# Integrating metabolomics and proteomics to identify novel drug targets for heart failure and atrial fibrillation

**DOI:** 10.1101/2023.10.19.23297247

**Authors:** Marion van Vugt, Chris Finan, Sandesh Chopade, Rui Providencia, Connie R. Bezzina, Folkert W. Asselbergs, Jessica van Setten, A. Floriaan Schmidt

## Abstract

**Background:** Altered metabolism plays a role in the pathophysiology of cardiac diseases, such as atrial fibrillation (AF) and heart failure (HF). We aimed to identify novel plasma metabolites and proteins associating with cardiac disease.

**Methods:** Mendelian randomisation (MR) was used to assess the association of 174 metabolites measured in up to 86,507 participants with AF, HF, dilated cardiomyopathy (DCM), and non-ischemic cardiomyopathy (NICM). Subsequently, we sourced data on 1,567 plasma proteins and performed *cis* MR to identify proteins affecting the identified metabolites as well as the cardiac diseases. Proteins were prioritised on cardiac expression and druggability, and mapped to biological pathways.

**Results:** We identified 35 metabolites associating with cardiac disease. AF was affected by seventeen metabolites, HF by nineteen, DCM by four, and NCIM by taurine. HF was particularly enriched for phosphatidylcholines (p=0.029) and DCM for acylcarnitines (p=0.001). Metabolite involvement with AF was more uniform, spanning for example phosphatidylcholines, amino acids, and acylcarnitines. We identified 38 druggable proteins expressed in cardiac tissue, with a directionally concordant effect on metabolites and cardiac disease. We recapitulated known associations, for example between the drug target of digoxin (AT1B2), taurine and NICM risk. Additionally, we identified numerous novel findings, such as higher RET values associating with phosphatidylcholines and decreasing AF and HF, and RET is targeted by drugs such as regorafenib which has known cardiotoxic side-effects. Pathway analysis implicated involvement of GDF15 signalling through RET, and ghrelin regulation of energy homeostasis in cardiac pathogenesis.

**Conclusion:** This study identified 35 plasma metabolites involved with cardiac diseases and linked these to 38 druggable proteins, providing actionable leads for drug development.

## Introduction

Atrial fibrillation (AF) and heart failure (HF) are common types of cardiac disease, resulting in substantial mortality and morbidity, and placing a major burden on healthcare and society^1,2^. Despite advances in management and understanding of disease pathophysiology, AF and HF prevalence remain high^1,2^. HF is a particularly heterogeneous disease potentially caused by cardiomyopathies, such as dilated cardiomyopathy (DCM) and non-ischemic cardiomyopathy (NICM).

Aside from N-terminal prohormone of brain natriuretic peptide (NT-proBNP) used in HF, troponin in ischemic cardiac disease, and D-dimers in pulmonary embolism, there are no plasma measurements to inform management of cardiac disease. Analyses of diseased hearts and cardiomyocytes have shown clear changes in cardiac metabolism, where diseased cardiomyocytes switch to ketone oxidation and glycolysis^3–5^. Small sample sized and predominantly cross-sectional studies of AF and HF patients have found elevated plasma values of acylcarnitines (ACs), which are markers of impaired fatty acid oxidation^6–12^.

Metabolic changes characterising cardiac diseases can inform drug development, which is especially important for HF and AF, where currently available drugs do not necessarily address underlying aetiology. For example, no novel AF drugs have been approved since the 2011 introduction of the anticoagulants rivaroxaban, apixaban, and edoxaban, which are indicated for stroke prevention in AF patient without addressing the underlying cause of AF.

Both metabolites and proteins, the main targets of most drugs^13^, are increasingly analysed by high-throughput assays measuring the plasma values of hundreds of analytes^14–16^. Genome-wide association studies (GWAS) have now identified genetic determinants of protein and metabolite values, which allow for integrative analyses identifying potential associations between the protein values, metabolite values, and cardiac disease in humans. Specifically, through two-sample Mendelian randomisation (MR), one can anticipate the effect an exposure (e.g. a metabolite) will have on the onset of disease by sourcing genetic variants strongly associated with the exposure and determining whether these variants show a dose-response association with disease^17,18^. This approach has been extensively validated for cardiovascular disease^19–22^, including a recent study on plasma proteins affecting cardiac function and structure as well as cardiac disease^23^.

In the current study, we sourced data on 174 metabolites measured in up to 86,507 participants and performed genome-wide MR to identify metabolites associated with AF, HF, DCM, and NICM. Subsequently, we leveraged data on 1,567 proteins measured in up to 35,559 participants and used *cis* MR to identify proteins affecting both the identified metabolites and the considered cardiac outcomes. We triangulated and prioritised proteins with concordant effects on metabolite values and cardiac outcomes. Through the integration of these omics data we identified 38 drugged and druggable proteins expressed in cardiac tissue, which affected 35 metabolites and cardiac disease.

## Methods

### Data sources

Genetic associations with 174 metabolites measured in up to 86,507 participants were available from Lotta *et al.*^24^ (**Supplementary Note**). Plasma protein data were available from eight GWAS: deCODE (SomaLogic assay, n=35,559)^25^, SCALLOP (OLINK assay, n=30,931)^26^, Ahola-Olli *et al.* (BioRad assay, n=8,293)^27^, Framingham (Luminex assay, n=6,861)^28^, AGES-Reykjavik (SomaLogic assay, n=5,368)^29^, INTERVAL (SomaLogic assay, n=3,301)^30^, Gilly *et al.* (Olink assay, n=1,328)^31^, and Yang *et al.* (SomaLogic assay, n=636)^32^. Genetic associations with AF were sourced from Nielsen *et al.* (60,620 cases)^33^, with HF from Shah *et al.* (47,309 cases)^34^, with DCM from Garnier *et al.* (2,719 cases)^35^, and with NICM from Aragam *et al.* (2,038 cases)^36^.

### Mendelian randomisation analyses

Genome-wide MR, considering genetic variants from across the genome, was used to establish associations between metabolite values and cardiac outcomes (step 1 in **Figure 1A**, **Appendix Table S1-2**). Subsequently *cis* MR, sourcing genetic variants from a 200 kilobase pair window around the protein encoding gene^17^, was used to identify plasma protein effects on metabolite values and cardiac outcomes (step 2-3 in **Figure 1A**). Irrespective of the type of MR analysis, instruments were selected based on an exposure F-statistic of at least 24 and a minor allele frequency of at least 0.01. Additionally, variants were clumped to a linkage disequilibrium r-squared of 0.3, based on a random sample of 5,000 unrelated UK biobank participants of European ancestry.

**Figure 1.**
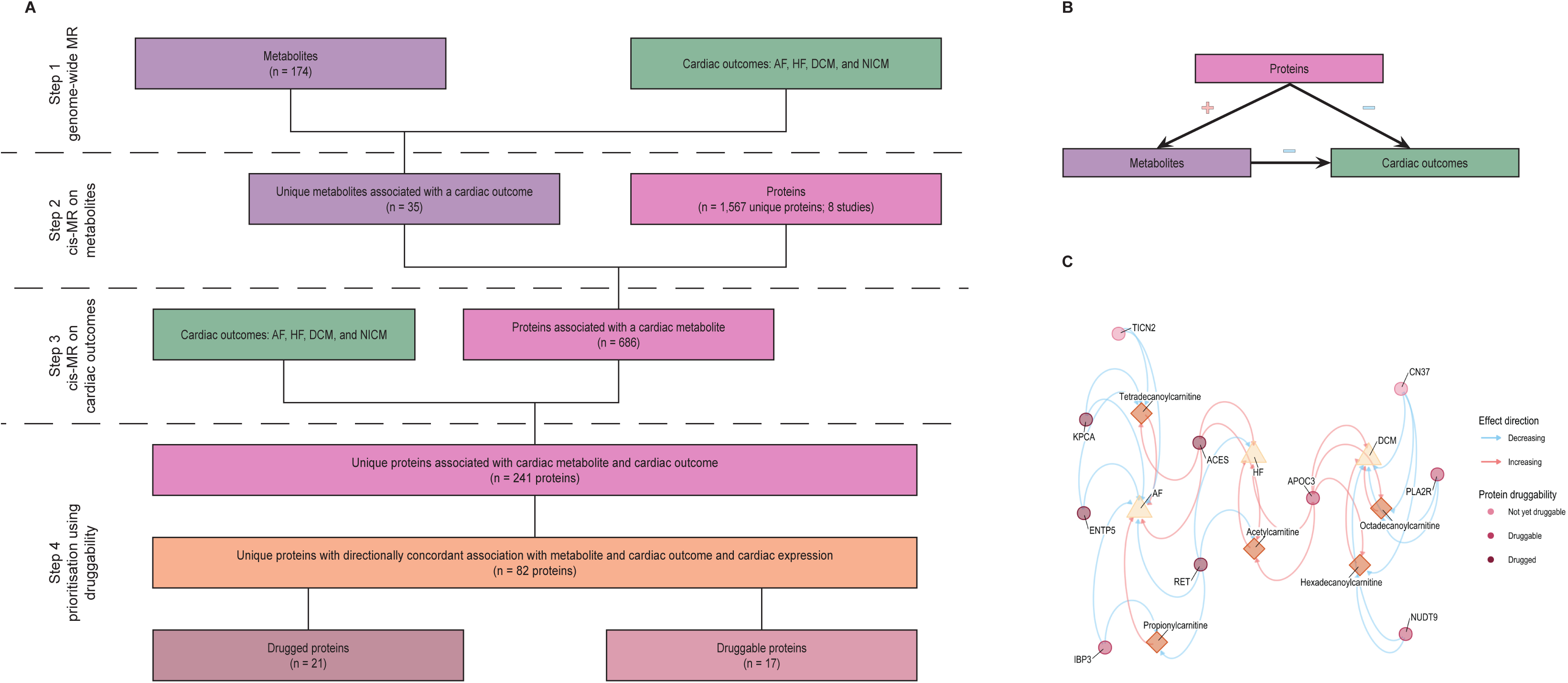
Study to identify plasma proteins and metabolites affecting cardiac disease. A) Flowchart of the analysis steps; B) A triangulation diagram, illustrating how a robust set of directionally concordant proteins was identified affecting plasma metabolite levels as well as cardiac disease. The red plus indicates a risk-increasing effect, while the blue minus indicates a risk-decreasing effect; C) Annotated network of prioritised metabolites, proteins, and outcomes for which the metabolites have at least 20% associated common proteins and belong to the metabolite class acylcarnitine. Prioritised directionally concordant proteins are represented by circles, metabolites by diamonds, outcomes by triangles. Circle colours represent protein druggability. Increasing effects are displayed by a red arrow, decreasing effects by a blue arrow. See **Supplementary Note** for more details. Abbreviations: AF = atrial fibrillation, DCM = dilated cardiomyopathy, HF = heart failure, MR = mendelian randomisation, NICM = non-ischemic cardiomyopathy.

Analyses were conducted using the generalised least squares implementation of the inverse-variance weighted (IVW) estimator, as well as with an Egger correction protecting against horizontal pleiotropy^37^. Influence of horizontal pleiotropy was minimised by excluding variants with a leverage statistic larger than three times the mean, and by excluding variants with an outlier (i.e. chi-square) statistic larger than 10.83^38^. To ensure we had sufficient data to accurately model the exposure effects, we discarded analyses with fewer than 6 variants. Additionally, a model selection strategy was used to select the most appropriate estimator (IVW or MR-Egger)^38,39^, where the MR-Egger is unbiased in the presence of directional horizontal pleiotropy. The model selection framework, originally developed by Rücker *et al.*^40^, uses the difference in heterogeneity between the IVW Q-statistic and the Egger Q-statistic to decide which method provides the best model to describe the available data. Furthermore, due to the two-sample nature of our analyses any potential weak-instrument bias would act toward a null effect, reducing power, rather than increasing type 1 errors^23^.

### Effect estimates, multiple testing, and nomenclature

MR results are presented as mean difference (MD) for continuous outcomes, or odds ratios (ORs) for binary outcomes, accompanied by 95% confidence intervals (CIs) and p-values. Metabolite effects on disease were filtered for a multiplicity corrected p-value threshold of 7×10^-5^ based on the 174 available metabolites and four outcomes, which is conservative, considering many metabolites are correlated. Similarly, protein MR effect estimates were evaluated against a multiplicity corrected p-value of 9×10^-7^ for the metabolites and 2×10^-5^ for the cardiac outcomes, based on the number of tested proteins (1,567 in the metabolite MR and 686 in the cardiac MR) and outcomes (36 in the metabolite MR and four in the cardiac MR). Proteins are referred to using their Uniprot label and presented in normal font to differentiate from gene names in italic font.

### Prioritisation and annotation of proteins

Proteins were prioritised based on the association with a plasma metabolite and a directionally concordant effect on cardiac outcome. For example, if an increase in a metabolite value decreased the risk of AF, a protein that increased the value of this metabolite should decrease AF risk to be deemed concordant. In this way, robust triangles of associations between the metabolites, proteins, and cardiac outcomes are created (**Figure 1B**). Given that this triangulation considers evidence from three separate MR analyses with distinct underlying assumptions on horizontal pleiotropy, this approach identifies a robust set of results, highly supported by the available data. For example, *cis* MR analyses assume the absence of pre-translational pleiotropy, which may be more robust compared to more distal assumptions on horizontal pleiotropy employed in genome-wide MR^17^.

The identified proteins were further annotated and prioritised on the presence of cardiac mRNA expression sourcing information from the human protein atlas (HPA)^41^. Subsequently, we identified genes which were overexpressed in cardiac tissue through comparisons against the average mRNA expression across the other 60 available tissues (**Supplementary Note**). We obtained information on druggability, indications and side-effects from ChEMBL v33 and the British National Formulary (BNF) identifying proteins as “druggable” if they are targeted by a developmental drug and “drugged” if they are targeted by an approved compound. Finally, the Reactome^42^ knowledgebase was queried to map all 1,567 proteins to biological pathways. Pathway involvement of the prioritised proteins was compared against the full set of 1,567 proteins using Wald tests.

### Replicating protein associations with metabolites and cardiac disease

The aforementioned *cis* MR analyses sourced data from eight proteomic GWAS, allowing for replication of the subset of identified proteins that were available in multiple studies. Of the 82 prioritised proteins (see results), 49 where available in multiple studies. For these 49 proteins the largest sample size GWAS was used for discovery and the remaining studies used for replication. Replication was sought by identifying associations with the same effect direction as the discovery association, and a p-value smaller than 0.05 (nominal replication) and 0.05/49=0.001 (conservative replication). Please see the **Supplementary Note** for additional details.

## Results

### Metabolite effects on cardiac outcomes

Genome-wide MR identified 35 metabolites associating with one or more cardiac disease (**Figure 2**, **Appendix Table S2-3**). Acylcarnitines (ACs) were enriched for DCM (p=0.001; **Appendix Table S4**), with higher values of hexadecanoylcarnitine (OR 1.51, 95%CI 1.28; 1.79), octadecanoylcarnitine (OR 1.47, 95%CI 1.25; 1.73), and octadecadienoylcarnitine (OR 1.38, 95%CI 1.22; 1.55) increasing DCM risk, while higher values of butyrylcarnitine decreased DCM risk (OR 0.87, 95%CI 0.82; 0.93). Phosphatidylcholines (PCs) were enriched for HF (p=0.029; **Appendix Table S4**) and all thirteen PCs associated with HF elicited a risk increasing effect. We observed more uniform effects for AF, which was affected by seventeen plasma metabolites. Finally, we observed that higher values of plasma taurine decreased NICM risk (OR 0.48, 95%CI 0.38; 0.60; **Figure 2**).

**Figure 2.**
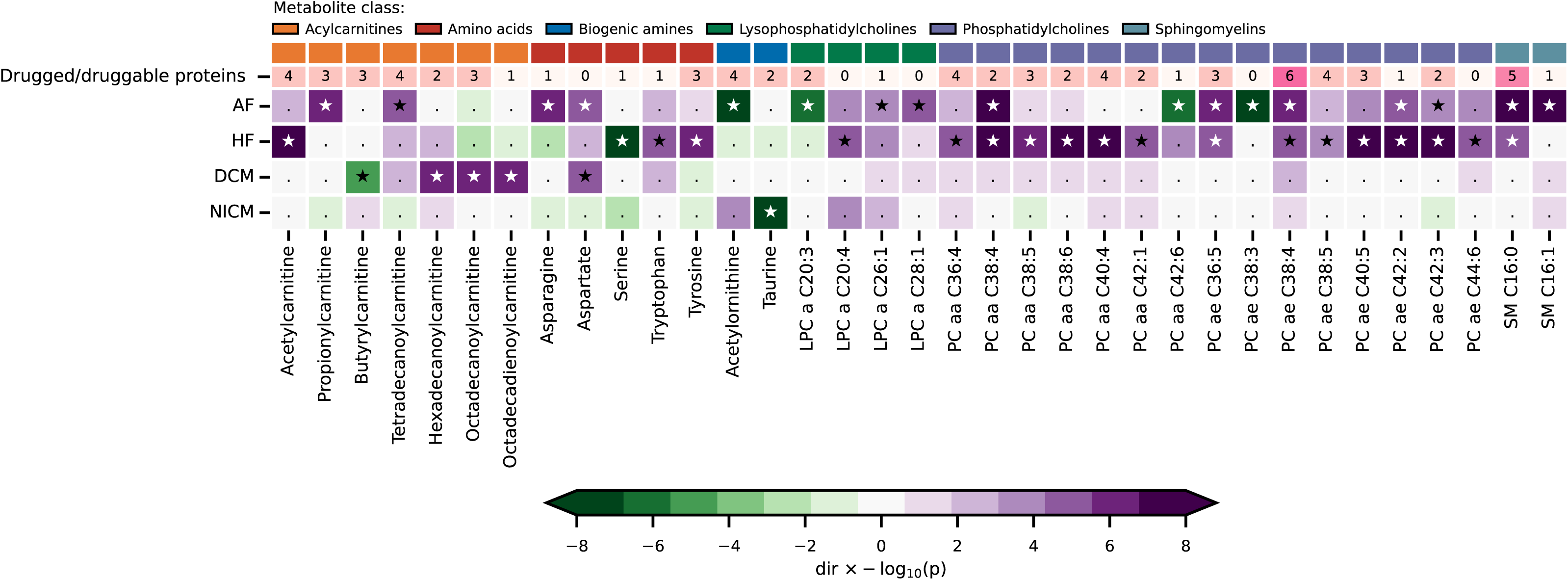
Plasma metabolites associating with at least one cardiac outcome. N.B. For visualisation purposes, the p-values were truncated to a -log(10) of 10. Multiplicity corrected significant associations are depicted by a star, non-significant associations by a dot. The top row indicates the number of druggable proteins reflecting information retrieved from BNF and ChEMBL. Genetic associations with the cardiac outcomes were obtained from Nielsen *et al.* (60,620 AF cases)^33^, Shah *et al.* (47,309 HF cases)^34^, Garnier *et al.* (2,719 DCM cases)^35^, and Aragam *et al.* (2,038 NICM cases)^36^. See the Methods section additional details, and Supplementary Table S2 for the underlying numerical data. Abbreviations: a = acyl residue, aa = diacyl residue, ae = acyl-alkyl residue, AF = atrial fibrillation, DCM = dilated cardiomyopathy, HF = heart failure, LPC = lysophosphatidylcholine, NICM = non-ischemic cardiomyopathy, PC = phosphatidylcholine, SM = sphingomyelin.

### Protein effects on metabolite values and cardiac outcomes

We evaluated 1,567 unique plasma proteins for associations with the 35 metabolites implicated in cardiac disease onset. This resulted in 686 proteins associated with one or more metabolite (step 2 in **Figure 1A**, **Appendix Figure S1**). Prioritising proteins on an association with cardiac outcomes identified 241 proteins (step 3 in **Figure 1A**, **Appendix Figure S2**), which were pruned down to 87 proteins by identifying the proteins with a directionally concordant effect on metabolites and cardiac outcomes (**Table 1-4**, **Appendix Table S5**). This triangulated subset of 87 proteins included 82 proteins that were expressed in cardiac tissue. Only CYTD, FETUB, PCSK9, PZP, and SPA11 were not expressed in cardiac tissue and six of the 82 prioritised proteins (ANX11, DNJA4, NAR3, PLXA1, RF1ML, and TIG1) were significantly overexpressed in the heart compared to other tissues (**Figure 3**, **Appendix Table S6**).

**Figure 3.**
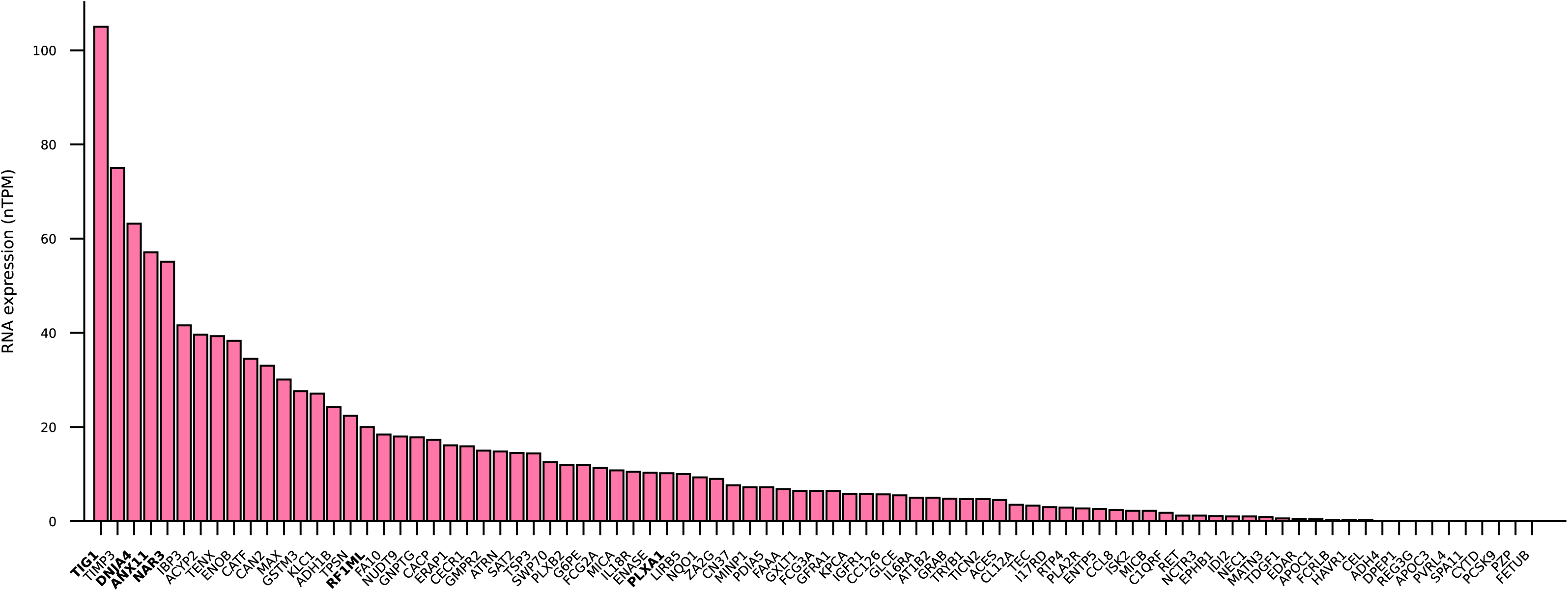
Cardiac mRNA expression of the subset of directionally concordant proteins affecting cardiac disease. N.B. Proteins are referred to using their Uniprot label and proteins in bold font are overexpressed in cardiac tissue relative to non-cardiac tissue. Data were sourced from the human protein atlas; see Methods section.

**Table 1.**
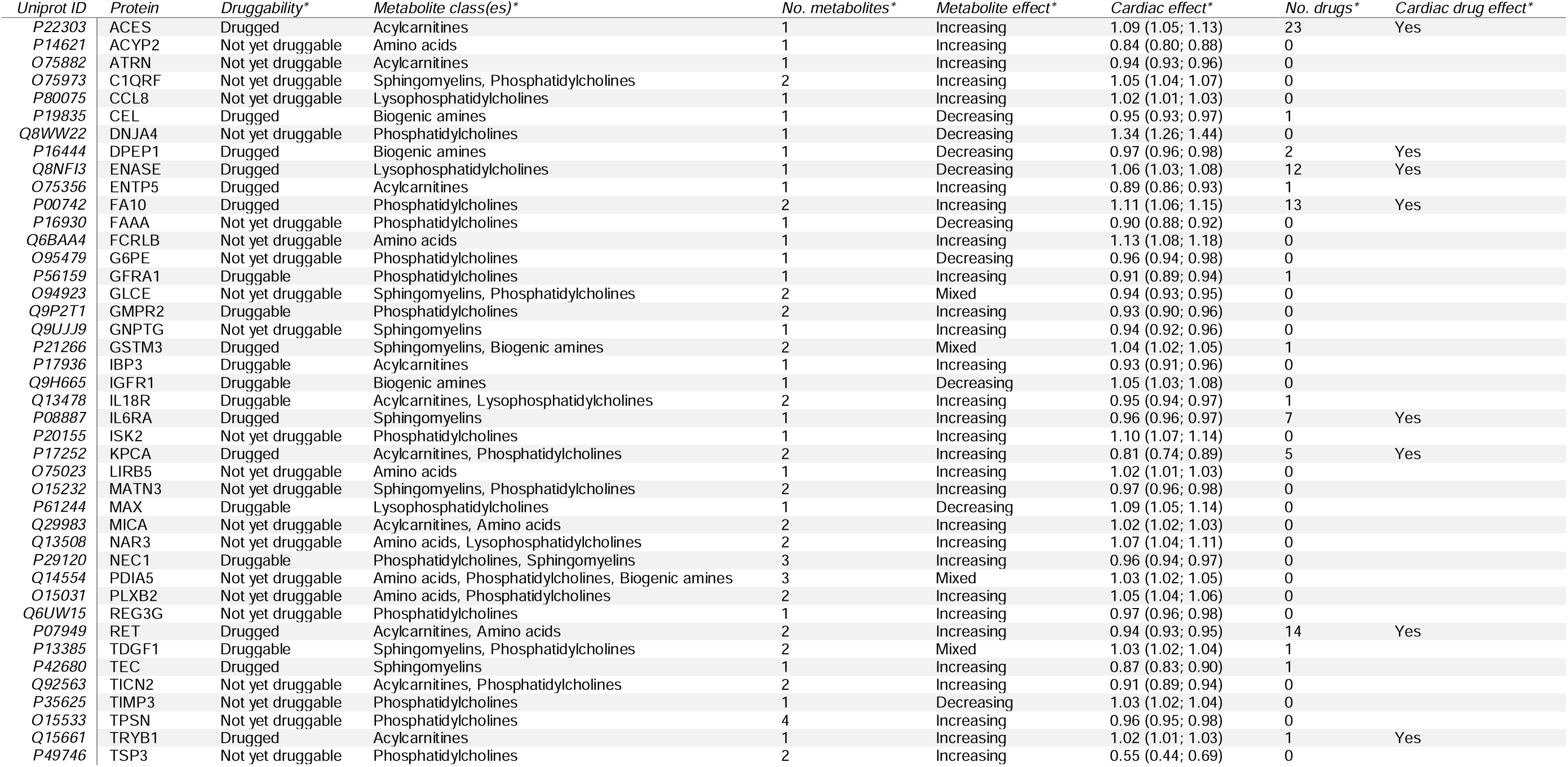
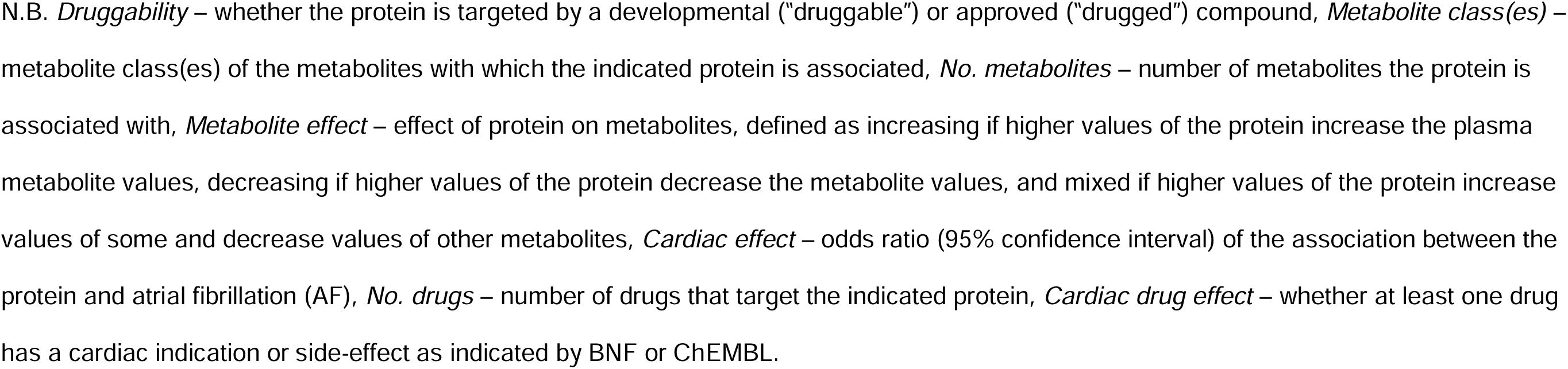
Overview and characteristics of prioritised proteins and metabolites associated with AF.

**Table 2.**
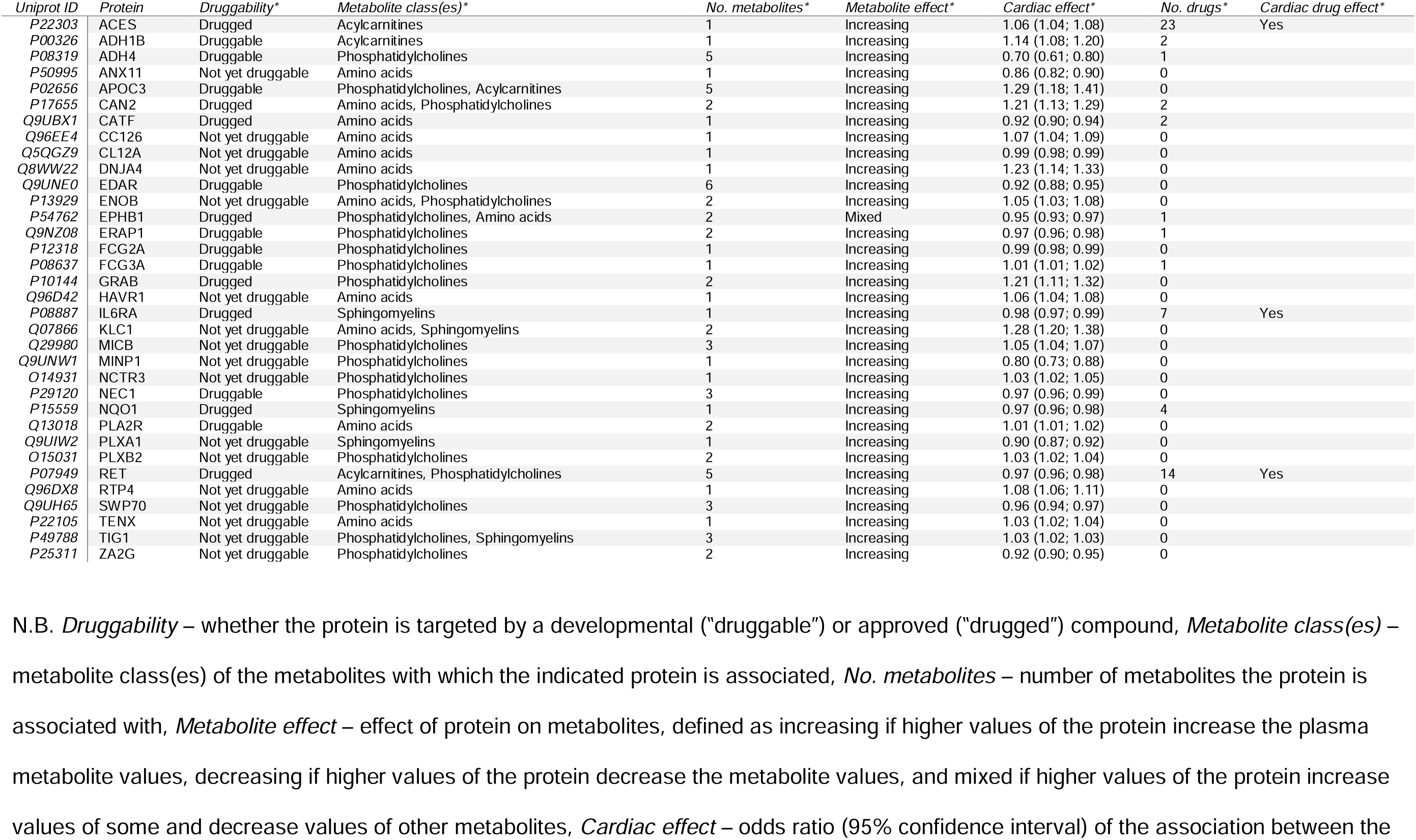
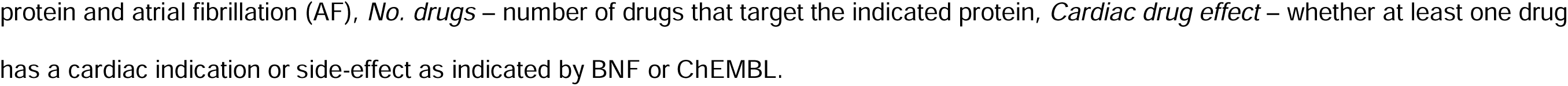
Overview and characteristics of prioritised proteins and metabolites associated with HF.

| Uniprot ID | Protein | Druggability* | Metabolite class(es)* | No. metabolites* | Metabolite effect* | Cardiac effect* | No. drugs* | Cardiac drug effect* |
| --- | --- | --- | --- | --- | --- | --- | --- | --- |
| P22303 | ACES | Drugged | Acylcarnitines | 1 | Increasing | 1.06 (1.04; 1.08) | 23 | Yes |
| P00326 | ADH1B | Druggable | Acylcarnitines | 1 | Increasing | 1.14 (1.08; 1.20) | 2 |  |
| P08319 | ADH4 | Druggable | Phosphatidylcholines | 5 | Increasing | 0.70 (0.61; 0.80) | 1 |  |
| P50995 | ANX11 | Not yet druggable | Amino acids | 1 | Increasing | 0.86 (0.82; 0.90) | 0 |  |
| P02656 | APOC3 | Druggable | Phosphatidylcholines, Acylcarnitines | 5 | Increasing | 1.29 (1.18; 1.41) | 0 |  |
| P17655 | CAN2 | Drugged | Amino acids, Phosphatidylcholines | 2 | Increasing | 1.21 (1.13; 1.29) | 2 |  |
| Q9UBX1 | CATF | Drugged | Amino acids | 1 | Increasing | 0.92 (0.90; 0.94) | 2 |  |
| Q96EE4 | CC126 | Not yet druggable | Amino acids | 1 | Increasing | 1.07 (1.04; 1.09) | 0 |  |
| Q5QGZ9 | CL12A | Not yet druggable | Amino acids | 1 | Increasing | 0.99 (0.98; 0.99) | 0 |  |
| Q8WW22 | DNJA4 | Not yet druggable | Amino acids | 1 | Increasing | 1.23 (1.14; 1.33) | 0 |  |
| Q9UNE0 | EDAR | Druggable | Phosphatidylcholines | 6 | Increasing | 0.92 (0.88; 0.95) | 0 |  |
| P13929 | ENOB | Not yet druggable | Amino acids, Phosphatidylcholines | 2 | Increasing | 1.05 (1.03; 1.08) | 0 |  |
| P54762 | EPHB1 | Drugged | Phosphatidylcholines, Amino acids | 2 | Mixed | 0.95 (0.93; 0.97) | 1 |  |
| Q9NZ08 | ERAP1 | Druggable | Phosphatidylcholines | 2 | Increasing | 0.97 (0.96; 0.98) | 1 |  |
| P12318 | FCG2A | Druggable | Phosphatidylcholines | 1 | Increasing | 0.99 (0.98; 0.99) | 0 |  |
| P08637 | FCG3A | Druggable | Phosphatidylcholines | 1 | Increasing | 1.01 (1.01; 1.02) | 1 |  |
| P10144 | GRAB | Drugged | Phosphatidylcholines | 2 | Increasing | 1.21 (1.11; 1.32) | 0 |  |
| Q96D42 | HAVR1 | Not yet druggable | Amino acids | 1 | Increasing | 1.06 (1.04; 1.08) | 0 |  |
| P08887 | IL6RA | Drugged | Sphingomyelins | 1 | Increasing | 0.98 (0.97; 0.99) | 7 | Yes |
| Q07866 | KLC1 | Not yet druggable | Amino acids, Sphingomyelins | 2 | Increasing | 1.28 (1.20; 1.38) | 0 |  |
| Q29980 | MICB | Not yet druggable | Phosphatidylcholines | 3 | Increasing | 1.05 (1.04; 1.07) | 0 |  |
| Q9UNW1 | MINP1 | Not yet druggable | Phosphatidylcholines | 1 | Increasing | 0.80 (0.73; 0.88) | 0 |  |
| O14931 | NCTR3 | Not yet druggable | Phosphatidylcholines | 1 | Increasing | 1.03 (1.02; 1.05) | 0 |  |
| P29120 | NEC1 | Druggable | Phosphatidylcholines | 3 | Increasing | 0.97 (0.96; 0.99) | 0 |  |
| P15559 | NQO1 | Drugged | Sphingomyelins | 1 | Increasing | 0.97 (0.96; 0.98) | 4 |  |
| Q13018 | PLA2R | Druggable | Amino acids | 2 | Increasing | 1.01 (1.01; 1.02) | 0 |  |
| Q9UIW2 | PLXA1 | Not yet druggable | Sphingomyelins | 1 | Increasing | 0.90 (0.87; 0.92) | 0 |  |
| O15031 | PLXB2 | Not yet druggable | Phosphatidylcholines | 2 | Increasing | 1.03 (1.02; 1.04) | 0 |  |
| P07949 | RET | Drugged | Acylcarnitines, Phosphatidylcholines | 5 | Increasing | 0.97 (0.96; 0.98) | 14 | Yes |
| Q96DX8 | RTP4 | Not yet druggable | Amino acids | 1 | Increasing | 1.08 (1.06; 1.11) | 0 |  |
| Q9UH65 | SWP70 | Not yet druggable | Phosphatidylcholines | 3 | Increasing | 0.96 (0.94; 0.97) | 0 |  |
| P22105 | TENX | Not yet druggable | Amino acids | 1 | Increasing | 1.03 (1.02; 1.04) | 0 |  |
| P49788 | TIG1 | Not yet druggable | Phosphatidylcholines, Sphingomyelins | 3 | Increasing | 1.03 (1.02; 1.03) | 0 |  |
| P25311 | ZA2G | Not yet druggable | Phosphatidylcholines | 2 | Increasing | 0.92 (0.90; 0.95) | 0 |  |

N.B. *Druggability* – whether the protein is targeted by a developmental (“druggable”) or approved (“drugged”) compound, *Metabolite class(es)* – metabolite class(es) of the metabolites with which the indicated protein is associated, *No. metabolites* – number of metabolites the protein is associated with, *Metabolite effect* – effect of protein on metabolites, defined as increasing if higher values of the protein increase the plasma metabolite values, decreasing if higher values of the protein decrease the metabolite values, and mixed if higher values of the protein increase values of some and decrease values of other metabolites, *Cardiac effect* – odds ratio (95% confidence interval) of the association between the
protein and atrial fibrillation (AF), *No. drugs* – number of drugs that target the indicated protein, *Cardiac drug effect* – whether at least one drug has a cardiac indication or side-effect as indicated by BNF or ChEMBL.

ChEMBL and BNF were consulted to identify seventeen proteins that were druggable (i.e., targeted by a developmental compound) and 21 that were drugged (i.e., targeted by an approved compound; **Figure 4, Appendix Table S7-8**). Eleven of the 21 drugged proteins were targeted by drugs with a cardiac indication, side-effect, or both. For example, higher values of AT1B2 (targeted by HF drug digoxin) increased NICM risk and decreased plasma taurine values. Furthermore, higher values of DPEP1 (associated with acetylornithine values) and KPCA (associated with tetradecanoylcarnitine and PC ae C38:4 values) decreased AF risk, while higher values of FA10 (associated with PC ae C42:2 and PC ae C38:4 values) increased AF risk (**Figure 4**). DPEP1 is inhibited by cilastatin, which is indicated for endocarditis and is implicated with tachycardia, KPCA is inhibited by midostaurin, which is known to elicit QT-interval prolongation, and FA10 is inhibited by drugs like apixaban, edoxaban, and rivaroxaban, which are commonly used to prevent stroke in AF. The remaining ten drugged proteins were targeted by compounds indicated for treatment of cancers (CAN2, EPHB1, ERAP1, GSTM3, NQO1, and PVRL4), mitochondrial and muscular disease (NQO1), and metabolic syndrome (CEL; **Appendix Table S8-9**).

**Figure 4.**
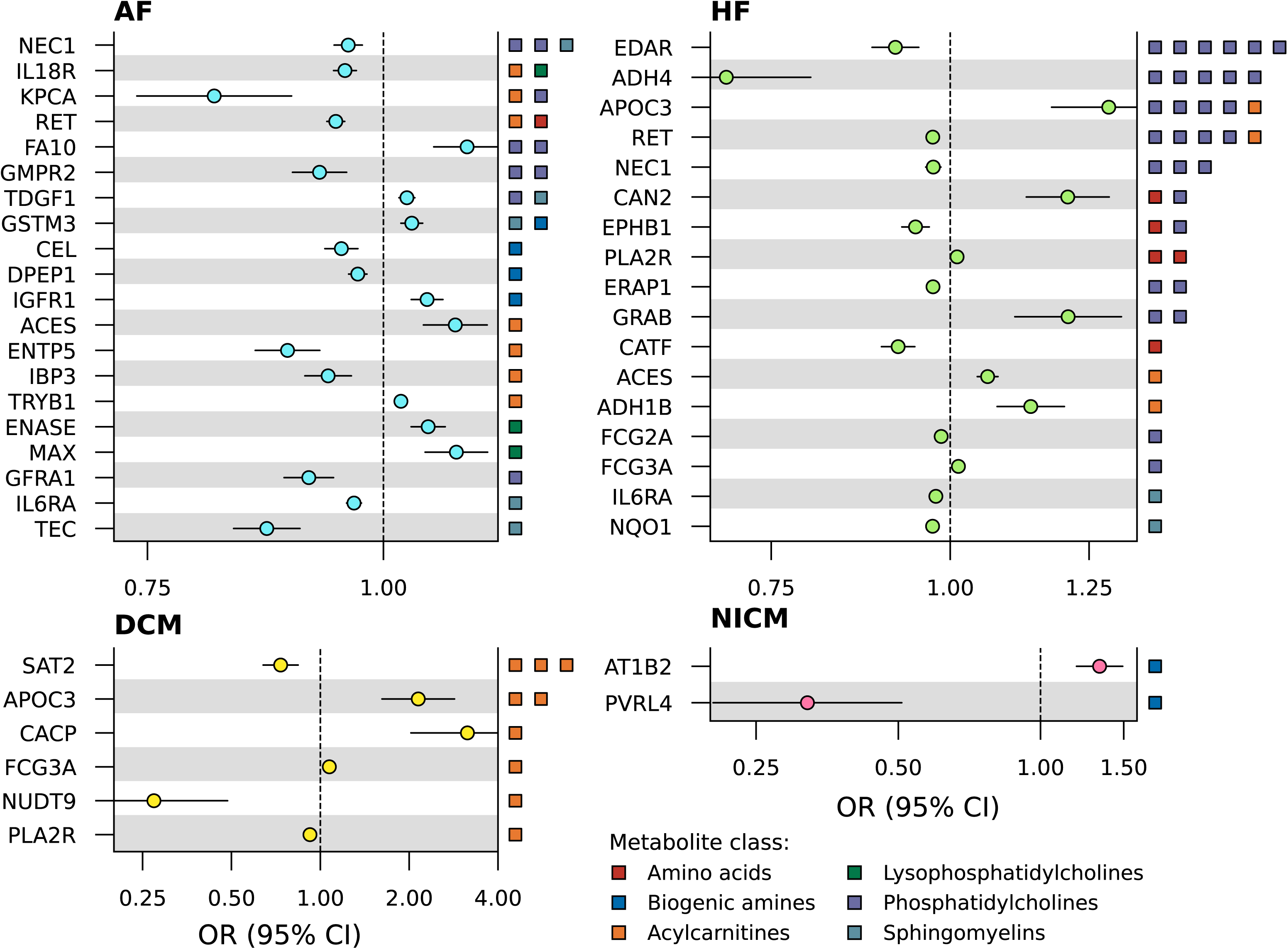
Forest plots of cardiac outcome effect of prioritised drugged or druggable proteins. N.B. Metabolite class of the metabolites affected by the protein are indicated on the right y-axis. Genetic associations with the cardiac outcomes were obtained from Nielsen *et al.* (60,620 AF cases)^33^, Shah *et al.* (47,309 HF cases)^34^, Garnier *et al.* (2,719 DCM cases)^35^, and Aragam *et al.* (2,038 NICM cases)^36^. See the Methods section for a more detailed description and **Appendix Table S7-8** for the full numerical results. Abbreviations: AF = atrial fibrillation, CI = confidence interval, DCM = dilated cardiomyopathy, HF = heart failure, NICM = non-ischemic cardiomyopathy, OR = odds ratio.

Irrespective of the indication or reported side-effect, drugged or druggable proteins associating with HF frequently affected phosphatidylcholines values. Higher values of ADH4 (targeted by nitrefazole), EDAR, EPHB1 (targeted by cancer drug vandetanib), ERAP1 (targeted by cancer drug tosedostat), and FCG2A decreased HF risk, while higher values of APOC3, CAN2 (targeted by amyloidosis drugs bortezomib and carfilzomib), and GRAB increased HF risk (**Figure 4**, **Appendix Table S7**). DCM was affected by six drugged or druggable proteins, all affecting one or more acylcarnitines. Higher values of SAT2 (targeted by trientine, which is in phase 2 clinical development for hypertrophic cardiomyopathy) and NUDT9 decreased DCM risk, while FCG3A (the target of imgatuzumab) and CACP (targeted by levocarnitine, which is being tested for AF and HF) increased DCM risk. Higher values of CEL (targeted by obesity medicine orlistat), ENTP5 (targeted by the psoriasis drug anthralin), GMPR2, GFRA1 (targeted by liatermin, a phase 1 compound for Parkinson’s disease), IBP3, and TEC (targeted by Pf-06651600, which is in clinical testing for arthritis, kidney and Crohn’s disease) decreased AF risk, while ENASE (targeted by AF drug edoxaban), GSTM3 (targeted by carmustine), IGFR1, TDGF1 (targeted by the antibody biib-015 used to treat cancer), and TRYB1 (targeted by arginine, which is being tested for HF) increased AF risk, affecting metabolites of various classes (**Figure 4**, **Appendix Table S7-8**). Four drugs target more than one prioritised protein: Cep-2563 targets KPCA and RET, edoxaban targets ENASE and FA10, nitrefazole targets ADH1B and ADH4, and vandetanib targets EPHB1 and RET (**Appendix Table S8**).

Ten pleiotropic proteins affected multiple cardiac outcomes (**Figure 5**). For example, higher values of ACES (inhibited by pyridostigmine bromide, which is in phase 2 testing for HF), DNJA4, and PLXB2 increased AF and HF risk, whereas higher values of IL6RA (targeted by tocilizumab), NEC1, and RET (targeted by regorafenib, which has cardiac side-effects) decreased AF and HF risk. Higher values of APOC3 and FCG3A increased HF and DCM risk. ENOB increased the risk of HF (OR 1.05, 95%CI 1.03; 1.08), DCM (OR 1.39, 95%CI 1.28; 1.50), and NICM (OR 1.24, 95%CI 1.13; 1.35; **Figure 5**, **Appendix Table S7**).

**Figure 5.**
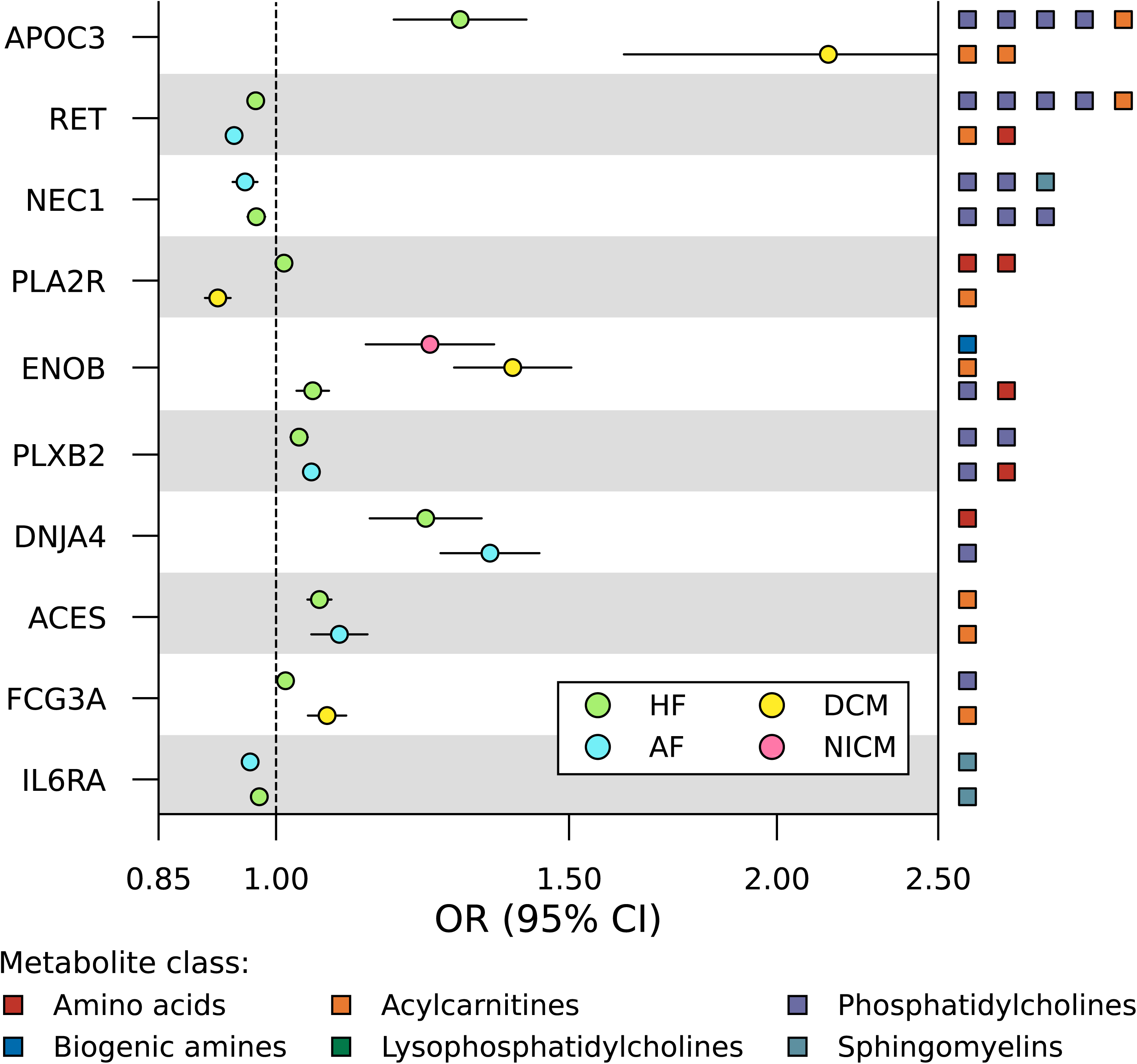
Forest plot of prioritised proteins associating with more than one cardiac outcome. N.B. . Metabolite class of the metabolites affected by the protein are indicated on the right y-axis. Genetic associations with the cardiac outcomes were obtained from Nielsen *et al.* (60,620 AF cases)^33^, Shah *et al.* (47,309 HF cases)^34^, Garnier *et al.* (2,719 DCM cases)^35^, and Aragam *et al.* (2,038 NICM cases)^36^. See the Methods section for a more detailed description and **Appendix Table S7** for the full numerical results. Abbreviations: AF = atrial fibrillation, CI = confidence interval, DCM = dilated cardiomyopathy, HF = heart failure, NICM = non-ischemic cardiomyopathy, OR = odds ratio.

### Identifying enriched biological pathways

We used the Reactome knowledgebase^42^ to identify enriched pathways among the prioritised proteins which identified three enriched pathways involving seven of the prioritised proteins. Specifically, GFRA1, KPCA, and RET were involved in the “RET signalling” pathway, TPSN and ERAP1 in “antigen presentation: folding, assembly and peptide loading of class I MHC”, and ACES and NEC1 in the “synthesis, secretion, and deacylation of ghrelin” pathway (**Appendix Table S10**).

### Replicating protein associations with cardiac disease

Replication data from independent plasma protein data were available for 49 out of 82 prioritised proteins. Applying a nominal replication p-value of 0.05, we were able to replicate the cardiac association of 38 proteins (77.6%; **Appendix Table S11**). The effects of 24 proteins were replicated more than once, with the IL6RA effects on AF and HF replicated in up to five studies. Applying a multiple testing adjusted p-value cut-off, resulted in 30 replicated proteins (61.2%), also including IL6RA. Please see the **Supplementary Note** for replication of the protein association with plasma metabolite values.

## Discussion

In the current study we integrated genetic associations with 174 metabolites and 1,567 proteins to identify determinants of cardiac disease relevant for de novo drug development and drug repurposing. Results were prioritised on concordant effect direction and cardiac tissue expression, identifying 82 triangulated proteins and 35 metabolites associating with AF, HF, DCM, and NICM. Plasma acylcarnitine (AC) values were strongly related with DCM, while phosphatidylcholine (PC) values were important determinants of HF. Metabolites affecting AF spanned six metabolite classes, potentially reflecting the heterogeneous aetiology of AF. We identified ten pleiotropic proteins affecting multiple cardiac traits: ACES, APOC3, DNJA4, ENOB, FCG3A, IL6RA, NEC1, PLA2R, PLXB2, and RET. The 38 drugged and druggable proteins included eleven proteins with a known cardiac indication, side-effect, or both: ACES (donepezil), AT1B2 (digoxin), CACP (levocarnitine), DPEP1 (cilastatin), ENASE (edoxaban), FA10 (apixaban), IL6RA (tocilizumab), KPCA (midostaurin), RET (regorafenib), SAT2 (trientine), and TRYB1 (arginine). Pathway enrichment identified proteins involved in GDF15-dependent RET signalling, ghrelin homeostasis, and MHC I antigen presentation.

ACs were strongly associated with DCM onset, with prioritised proteins affecting DCM also associating with long-chain (hexadecanoylcarnitine, octadecanoylcarnitine, octadecadienoylcarnitine) and short-chain (butyrylcarnitine) AC values (**Table 3**, **Appendix Table S7**). Higher AC values additionally increased AF (propionylcarnitine and tetradecanoylcarnitine), and HF risk (acetylcarnitine). AC values are indicators of fatty acid oxidation, the main energy substrate of the healthy heart^4^, with particularly long-chain ACs the results of fatty acid metabolism^43^, while short-chain ACs are more generally synthesised during glucose, AA and fatty acid metabolism^43^. Fourteen drugged and druggable proteins associated with AC plasma values (**Appendix Table S7**). We showed that ACES affects the plasma values of several ACs and was associated with a risk increasing effect on AF and HF. ACES is targeted by several drugs with cardiac side-effects and inhibited by pyridostigmine bromide, which is being tested for HF (**Appendix Table S8**), increases heart rate variability, and reduces ventricular arrhythmias in chronic HF patients^44^, supporting the identified associations. NEC1 affects several PCs, AF, and HF, is a druggable protein that processes hormones such as insulin and its encoding gene *PCSK1* associated with HF risk factors such as obesity^45^ and abnormal glucose homeostasis^46^. NEC1 and ACES are involved in the enriched ghrelin pathway (**Appendix Table S10**), which regulates energy homeostasis and plays a role in myocardial function^47–50^. Ghrelin was therefore proposed as a novel target for HF management^51^.

**Table 3.** Overview and characteristics of prioritised proteins and metabolites associated with DCM.

| *Uniprot ID* | *Protein* | *Druggability\** | *Metabolite class(es)\** | *No. metabolites\** | *Metabolite effect\** | *Cardiac effect\** | *No. drugs\** | *Cardiac drug effect\** |
| --- | --- | --- | --- | --- | --- | --- | --- | --- |
| P02654 | APOC1 | Not yet druggable | Acylcarnitines | 1 | Increasing | 1.49 (1.26; 1.77) | 0 |  |
| P02656 | APOC3 | Druggable | Acylcarnitines | 2 | Increasing | 2.15 (1.62; 2.85) | 0 |  |
| P43155 | CACP | Drugged | Acylcarnitines | 1 | Decreasing | 3.15 (2.03; 4.90) | 1 | Yes |
| Q9NZK5 | CECR1 | Not yet druggable | Acylcarnitines | 1 | Decreasing | 0.82 (0.76; 0.90) | 0 |  |
| P09543 | CN37 | Not yet druggable | Acylcarnitines | 2 | Increasing | 0.50 (0.43; 0.57) | 0 |  |
| P13929 | ENOB | Not yet druggable | Acylcarnitines | 1 | Decreasing | 1.39 (1.28; 1.50) | 0 |  |
| P08637 | FCG3A | Druggable | Acylcarnitines | 1 | Decreasing | 1.07 (1.05; 1.10) | 1 |  |
| Q4G148 | GXLT1 | Not yet druggable | Acylcarnitines | 1 | Increasing | 1.68 (1.34; 2.10) | 0 |  |
| Q8NFM7 | I17RD | Not yet druggable | Acylcarnitines | 1 | Increasing | 0.76 (0.68; 0.86) | 0 |  |
| Q9BXS1 | IDI2 | Not yet druggable | Acylcarnitines | 1 | Increasing | 1.94 (1.57; 2.41) | 0 |  |
| Q9BW91 | NUDT9 | Druggable | Acylcarnitines | 1 | Increasing | 0.27 (0.15; 0.49) | 0 |  |
| Q13018 | PLA2R | Druggable | Acylcarnitines | 1 | Increasing | 0.92 (0.91; 0.94) | 0 |  |
| Q9UGC7 | RF1ML | Not yet druggable | Acylcarnitines | 1 | Increasing | 2.35 (1.93; 2.86) | 0 |  |
| Q96F10 | SAT2 | Drugged | Acylcarnitines | 3 | Mixed | 0.73 (0.64; 0.84) | 1 | Yes |

N.B. *Druggability* – whether the protein is targeted by a developmental (“druggable”) or approved (“drugged”) compound, *Metabolite class(es)* – metabolite class(es) of the metabolites with which the indicated protein is associated, *No. metabolites* – number of metabolites the protein is associated with, *Metabolite effect* – effect of protein on metabolites, defined as increasing if higher values of the protein increase the plasma metabolite values, decreasing if higher values of the protein decrease the metabolite values, and mixed if higher values of the protein increase values of some and decrease values of other metabolites, *Cardiac effect* – odds ratio (95% confidence interval) of the association between the protein and atrial fibrillation (AF), *No. drugs* – number of drugs that target the indicated protein, *Cardiac drug effect* – whether at least one drug has a cardiac indication or side-effect as indicated by BNF or ChEMBL.

**Table 4.** Overview and characteristics of prioritised proteins and metabolites associated with NICM.

| *Uniprot ID* | *Protein* | *Druggability\** | *Metabolite class(es)\** | *No. metabolites\** | *Metabolite effect\** | *Cardiac effect\** | *No. drugs\** | *Cardiac drug effect\** |
| --- | --- | --- | --- | --- | --- | --- | --- | --- |
| *P13929* | ENOB | Not yet druggable | Biogenic amines | 1 | Decreasing | 1.24 (1.13; 1.35) | 0 |  |
| *P14415* | AT1B2 | Drugged | Biogenic amines | 1 | Decreasing | 1.33 (1.19; 1.49) | 6 | Yes |
| *Q96NY8* | PVRL4 | Drugged | Biogenic amines | 1 | Decreasing | 0.32 (0.20; 0.51) | 1 |  |

N.B. *Druggability* – whether the protein is targeted by a developmental (“druggable”) or approved (“drugged”) compound, *Metabolite class(es)* – metabolite class(es) of the metabolites with which the indicated protein is associated, *No. metabolites* – number of metabolites the protein is associated with, *Metabolite effect* – effect of protein on metabolites, defined as increasing if higher values of the protein increase the plasma metabolite values, decreasing if higher values of the protein decrease the metabolite values, and mixed if higher values of the protein increase values of some and decrease values of other metabolites, *Cardiac effect* – odds ratio (95% confidence interval) of the association between the protein and atrial fibrillation (AF), *No. drugs* – number of drugs that target the indicated protein, *Cardiac drug effect* – whether at least one drug has a cardiac indication or side-effect as indicated by BNF or ChEMBL.

We identified thirteen PCs that increased HF risk, of which five also increased AF risk. These results suggest that PCs play an important role in cardiac function, which is supported by the findings that PCs are implicated in aberrant calcium handling^52,53^. We identified sixteen drugged and druggable proteins affecting PC values as well as AF or HF, or both (**Appendix Table S7**). Higher ERAP1 values (targeted by tosedostat) were shown to decrease plasma PC values and HF risk, is targeted by cancer drug tosedostat, and plays a role in blood pressure regulation^54^. Together with the not yet druggable protein TPSN, ERAP1 is involved in the enriched MHC I antigen presentation pathway which is linked to CD8+ T-cell mediated cardiac remodelling leading to HF^55^. We additionally found that higher KPCA values decreased AC and PC values and protected against AF. KPCA is targeted by HF drug midostaurin, and is involved in the significantly enriched RET signalling pathway, together with GFRA1 and RET (**Appendix Table S10**). The RET receptor is activated by GFRA1 and is needed for GDF15 signalling^56^, where GDF15 concentrations are predictive of HF outcome^57,58^. GDF15 has also been shown to associate with stroke, bleeding, and mortality in AF patients^59^. Furthermore, our observation that increased RET values are protective against AF and HF may indicate that the adverse effects of RET-inhibiting cancer drug regorafenib, such as hypertension and myocardial infarction, are on-target (**Table 1-2**, **Figure 4-5**).

We wish to discuss the following potential limitations. Firstly, our analyses focussed on plasma metabolites and proteins, which may not necessarily reflect in cardiac tissue values^60^. Evidence suggests that plasma values may serve as biomarkers of intracellular energy metabolism^8,61^ and correlate with metabolite values in cardiac tissues, for example cardiac profiles of medium- and long-chain ACs have been found to reflect plasma concentrations^43^. This is further supported by our observation that 82 of the 87 triangulated plasma proteins were expressed in cardiac tissue. It is important to note that proteins without cardiac expression can affect cardiac function, thereby contributing to disease development. PCSK9, for example, was one of five proteins not expressed in cardiac tissue (**Figure 3**) but is anticipated to decrease HF risk through prevention of progressive CHD^20,62–64^. Additionally, animal studies suggest that PCSK9 inhibition may induce an LDL receptor-mediated improvement in cardiac lipid metabolism^65^, providing illustrations of plasma protein involvement with cardiac metabolism. A second potential limitation is our reliance on high throughput assays, which differ in terms of accuracy and coverage^66^ and measure relative protein or metabolite values rather than concentrations. This implies that the reported effect estimates and their magnitudes need not directly translate to anticipated clinical effects, hence small effects may nevertheless mark potentially clinically impactful targets. Rather we suggest a focus on effect direction, providing information on the type of perturbation (activating versus inhibiting), might be more informative. Third, while MR studies are susceptible to bias through horizontal pleiotropy, we mitigate this source of bias by removing potentially pleiotropic variants, employing a model selection framework, and triangulating results across analysis with distinct pleiotropy assumptions. Specifically, our triangulation approach minimized the potential for horizontal pleiotropy, by looking for agreement across three distinct MR analyses which also made distinct assumptions on horizontal pleiotropy. The robustness of this approach is highlighted by the rediscovery of multiple cardiac drug targets such as edoxaban, apixaban, and rivaroxaban for AF and digoxin for HF. The triangulation of AT1B2 with taurine and NICM is particularly illustrative, because taurine supplementation may be used to replace or reduce the dosage of digoxin prescription in HF^67^. Finally, we emphasise that we were able to replicate associations for up to 77.6% of the proteins available in multiple studies.

In conclusion, we identify 35 metabolites and 38 drugged or druggable proteins associating with AF, HF, DCM, and NICM. We were able to relate these findings to biological pathways relating to energy homeostasis through ghrelin and GDF15 signalling. Our findings provide insight into pathogenesis of cardiac disease and leads for cardiac drug development.

## Data availability

All scripts and aggregated data are available online at <URL available upon publication>.

## Non-standard abbreviations and acronyms

AA: amino acid
AC: acylcarnitine
AF: atrial fibrillation
BNF: British National Formulary
CI: confidence interval
DCM: dilated cardiomyopathy
EGFR: epidermal growth factor receptor
GWAS: genome-wide association studies
HF: heart failure
HPA: human protein atlas
IQR: interquartile range
IVW: inverse-variance weighted
LPC: lysophosphatidylcholine
MD: mean difference
MR: mendelian randomisation
NICM: non-ischemic cardiomyopathy
nTPM: normalised transcripts per million
NT-proBNP: N-terminal prohormone of brain natriuretic peptide
OR: odds ratio
PC: phosphatidylcholine
SM: sphingomyelin

## References

1. Elliott, A. D., Middeldorp, M. E., Van Gelder, I. C., Albert, C. M. & Sanders, P. Epidemiology and modifiable risk factors for atrial fibrillation. Nat. Rev. Cardiol. 1–14 (2023).

2. Ziaeian, B. & Fonarow, G. C. Epidemiology and aetiology of heart failure. Nat. Rev. Cardiol. 13, 368–378 (2016).

3. Lopaschuk, G. D., Karwi, Q. G., Tian, R., Wende, A. R. & Abel, E. D. Cardiac energy metabolism in heart failure. Circ. Res. 128, 1487–1513 (2021).

4. Dambrova, M. et al. Acylcarnitines: nomenclature, biomarkers, therapeutic potential, drug targets, and clinical trials. Pharmacol. Rev. 74, 506–551 (2022).

5. Wiersma, M. et al. Mitochondrial dysfunction underlies cardiomyocyte remodeling in experimental and clinical atrial fibrillation. Cells 8, 1202 (2019).

6. Ueland, T. et al. Disturbed carnitine regulation in chronic heart failure—increased plasma levels of palmitoyl-carnitine are associated with poor prognosis. Int. J. Cardiol. 167, 1892–1899 (2013).

7. Ahmad, T. et al. Prognostic implications of long-chain acylcarnitines in heart failure and reversibility with mechanical circulatory support. J. Am. Coll. Cardiol. 67, 291–299 (2016).

8. Hunter, W. G. et al. Metabolomic profiling identifies novel circulating biomarkers of mitochondrial dysfunction differentially elevated in heart failure with preserved versus reduced ejection fraction: evidence for shared metabolic impairments in clinical heart failure. J. Am. Heart Assoc. 5, e003190 (2016).

9. Ruiz, M. et al. Circulating acylcarnitine profile in human heart failure: a surrogate of fatty acid metabolic dysregulation in mitochondria and beyond. Am. J. Physiol.-Heart Circ. Physiol. 313, H768–H781 (2017).

10. Zordoky, B. N. et al. Metabolomic fingerprint of heart failure with preserved ejection fraction. PloS One 10, e0124844 (2015).

11. Ruiz-Canela, M., et al. Plasma acylcarnitines and risk of incident heart failure and atrial fibrillation: the Prevención con dieta mediterránea study. Rev. Esp. Cardiol. Engl. Ed. 75, 649–658 (2022).

12. Smith, E., Fernandez, C., Melander, O. & Ottosson, F. Altered acylcarnitine metabolism is associated with an increased risk of atrial fibrillation. J. Am. Heart Assoc. 9, e016737 (2020).

13. Finan, C. et al. The druggable genome and support for target identification and validation in drug development. Sci. Transl. Med. 9, eaag1166 (2017).

14. Soininen, P. et al. High-throughput serum NMR metabonomics for cost-effective holistic studies on systemic metabolism. Analyst 134, 1781–1785 (2009).

15. Würtz, P. et al. Quantitative serum nuclear magnetic resonance metabolomics in large-scale epidemiology: a primer on-omic technologies. Am. J. Epidemiol. 186, 1084–1096 (2017).

16. Gold, L. et al. Aptamer-based multiplexed proteomic technology for biomarker discovery. Nat. Preced. 1–1 (2010).

17. Schmidt, A. F. et al. Genetic drug target validation using Mendelian randomisation. Nat. Commun. 11, 3255 (2020).

18. Schmidt, A. F., Hingorani, A. D. & Finan, C. Human genomics and drug development. Cold Spring Harb. Perspect. Med. 12, (2022).

19. Consortium, I.-6 R. M. R. A. (IL6R M. & others. The interleukin-6 receptor as a target for prevention of coronary heart disease: a mendelian randomisation analysis. The Lancet 379, 1214–1224 (2012).

20. Schmidt, A. F. et al. Cholesteryl ester transfer protein (CETP) as a drug target for cardiovascular disease. Nat. Commun. 12, 5640 (2021).

21. Zheng, J. et al. Phenome-wide Mendelian randomization mapping the influence of the plasma proteome on complex diseases. Nat. Genet. 52, 1122–1131 (2020).

22. Cupido, A. J. et al. Joint genetic inhibition of PCSK9 and CETP and the association with coronary artery disease: a factorial Mendelian randomization study. JAMA Cardiol. 7, 955–964 (2022).

23. Schmidt, A. F. et al. Druggable proteins influencing cardiac structure and function: Implications for heart failure therapies and cancer cardiotoxicity. Sci. Adv. 9, eadd4984 (2023).

24. Lotta, L. A. et al. A cross-platform approach identifies genetic regulators of human metabolism and health. Nat. Genet. 53, 54–64 (2021).

25. Ferkingstad, E. et al. Large-scale integration of the plasma proteome with genetics and disease. Nat. Genet. 53, 1712–1721 (2021).

26. Folkersen, L. et al. Genomic and drug target evaluation of 90 cardiovascular proteins in 30,931 individuals. Nat. Metab. 2, 1135–1148 (2020).

27. Ahola-Olli, A. V. et al. Genome-wide association study identifies 27 loci influencing concentrations of circulating cytokines and growth factors. Am. J. Hum. Genet. 100, 40– 50 (2017).

28. Yao, C. et al. Genome-wide mapping of plasma protein QTLs identifies putatively causal genes and pathways for cardiovascular disease. Nat. Commun. 9, 3268 (2018).

29. Gudjonsson, A. et al. A genome-wide association study of serum proteins reveals shared loci with common diseases. Nat. Commun. 13, 480 (2022).

30. Sun, B. B. et al. Genomic atlas of the human plasma proteome. Nature 558, 73–79 (2018).

31. Gilly, A. et al. Whole-genome sequencing analysis of the cardiometabolic proteome. Nat. Commun. 11, 6336 (2020).

32. Yang, C. et al. Genomic atlas of the proteome from brain, CSF and plasma prioritizes proteins implicated in neurological disorders. Nat. Neurosci. 24, 1302–1312 (2021).

33. Nielsen, J. B. et al. Biobank-driven genomic discovery yields new insight into atrial fibrillation biology. Nat. Genet. 50, 1234–1239 (2018).

34. Shah, S. et al. Genome-wide association and Mendelian randomisation analysis provide insights into the pathogenesis of heart failure. Nat. Commun. 11, 1–12 (2020).

35. Garnier, S. et al. Genome-wide association analysis in dilated cardiomyopathy reveals two new players in systolic heart failure on chromosomes 3p25. 1 and 22q11. 23. Eur. Heart J. 42, 2000–2011 (2021).

36. Aragam, K. G. et al. Phenotypic refinement of heart failure in a national biobank facilitates genetic discovery. Circulation 139, 489–501 (2019).

37. Bowden, J., Davey Smith, G. & Burgess, S. Mendelian randomization with invalid instruments: effect estimation and bias detection through Egger regression. Int. J. Epidemiol. 44, 512–525 (2015).

38. Bowden, J. et al. A framework for the investigation of pleiotropy in two-sample summary data Mendelian randomization. Stat. Med. 36, 1783–1802 (2017).

39. Bowden, J. et al. Improving the visualization, interpretation and analysis of two-sample summary data Mendelian randomization via the Radial plot and Radial regression. Int. J. Epidemiol. 47, 1264–1278 (2018).

40. Rücker, G., Schwarzer, G., Carpenter, J. R., Binder, H. & Schumacher, M. Treatment-effect estimates adjusted for small-study effects via a limit meta-analysis. Biostatistics 12, 122–142 (2011).

41. Uhlen, M. et al. Towards a knowledge-based human protein atlas. Nat. Biotechnol. 28, 1248–1250 (2010).

42. Jassal, B. et al. The reactome pathway knowledgebase. Nucleic Acids Res. 48, D498–D503 (2020).

43. Makrecka-Kuka, M. et al. Plasma acylcarnitine concentrations reflect the acylcarnitine profile in cardiac tissues. Sci. Rep. 7, 17528 (2017).

44. Behling, A. et al. Cholinergic stimulation with pyridostigmine reduces ventricular arrhythmia and enhances heart rate variability in heart failure. Am. Heart J. 146, 494–500 (2003).

45. Benzinou, M. et al. Common nonsynonymous variants in PCSK1 confer risk of obesity. Nat. Genet. 40, 943–945 (2008).

46. Heni, M., et al. Association of obesity risk SNPs in PCSK1 with insulin sensitivity and proinsulin conversion. (2010).

47. Nagaya, N. et al. Effects of ghrelin administration on left ventricular function, exercise capacity, and muscle wasting in patients with chronic heart failure. Circulation 110, 3674–3679 (2004).

48. Beiras-Fernandez, A. et al. Altered myocardial expression of ghrelin and its receptor (GHSR-1a) in patients with severe heart failure. Peptides 31, 2222–2228 (2010).

49. Ma, T. et al. Ghrelin expression and significance in 92 patients with atrial fibrillation. Anatol. J. Cardiol. 18, 99 (2017).

50. Yuan, M.-J., Li, W. & Zhong, P. Research progress of ghrelin on cardiovascular disease. Biosci. Rep. 41, BSR20203387 (2021).

51. Nagaya, N. & Kangawa, K. Therapeutic potential of ghrelin in the treatment of heart failure. Drugs 66, 439–448 (2006).

52. Li, Y. et al. Enrichment of endoplasmic reticulum with cholesterol inhibits sarcoplasmic-endoplasmic reticulum calcium ATPase-2b activity in parallel with increased order of membrane lipids: implications for depletion of endoplasmic reticulum calcium stores and apoptosis in cholesterol-loaded macrophages. J. Biol. Chem. 279, 37030–37039 (2004).

53. Fu, S. et al. Aberrant lipid metabolism disrupts calcium homeostasis causing liver endoplasmic reticulum stress in obesity. Nature 473, 528–531 (2011).

54. Cifaldi, L., Romania, P., Lorenzi, S., Locatelli, F. & Fruci, D. Role of endoplasmic reticulum aminopeptidases in health and disease: from infection to cancer. Int. J. Mol. Sci. 13, 8338–8352 (2012).

55. Komai, K. et al. Single-cell analysis revealed the role of CD8+ effector T cells in preventing cardioprotective macrophage differentiation in the early phase of heart failure. Front. Immunol. 12, 763647 (2021).

56. Buchholz, K. et al. Expression of the Body-Weight Signaling Players: GDF15, GFRAL and RET and their clinical relevance in Gastric Cancer. J. Cancer 12, 4698 (2021).

57. Rochette, L., Zeller, M., Cottin, Y. & Vergely, C. Insights into mechanisms of GDF15 and receptor GFRAL: therapeutic targets. Trends Endocrinol. Metab. 31, 939–951 (2020).

58. Xu, J. et al. GDF15/MIC-1 functions as a protective and antihypertrophic factor released from the myocardium in association with SMAD protein activation. Circ. Res. 98, 342– 350 (2006).

59. Wallentin, L. et al. Growth differentiation factor 15, a marker of oxidative stress and inflammation, for risk assessment in patients with atrial fibrillation: insights from the Apixaban for Reduction in Stroke and Other Thromboembolic Events in Atrial Fibrillation (ARISTOTLE) trial. Circulation 130, 1847–1858 (2014).

60. Flam, E. et al. Integrated landscape of cardiac metabolism in end-stage human nonischemic dilated cardiomyopathy. *Nat*. Cardiovasc. Res. 1, 817–829 (2022).

61. Rinaldo, P., Cowan, T. M. & Matern, D. Acylcarnitine profile analysis. Genet. Med. 10, 151–156 (2008).

62. Cohen, J. C., Boerwinkle, E., Mosley Jr, T. H. & Hobbs, H. H. Sequence variations in PCSK9, low LDL, and protection against coronary heart disease. N. Engl. J. Med. 354, 1264–1272 (2006).

63. Sabatine, M. S. et al. Evolocumab and clinical outcomes in patients with cardiovascular disease. N. Engl. J. Med. 376, 1713–1722 (2017).

64. Schmidt, A. F. et al. PCSK9 monoclonal antibodies for the primary and secondary prevention of cardiovascular disease. Cochrane Database Syst. Rev. (2017).

65. Da Dalt, L. et al. PCSK9 deficiency rewires heart metabolism and drives heart failure with preserved ejection fraction. Eur. Heart J. 42, 3078–3090 (2021).

66. Katz, D. H. et al. Proteomic profiling platforms head to head: leveraging genetics and clinical traits to compare aptamer-and antibody-based methods. Sci. Adv. 8, eabm5164 (2022).

67. Schaffer, S. & Kim, H. W. Effects and mechanisms of taurine as a therapeutic agent. Biomol. Ther. 26, 225 (2018).

