## Supplemental Table S1 for "Integrating metabolomics and proteomics to identify novel drug targets for heart failure and atrial fibrillation"

| Table S1. Metabolite information | | | | | |
|---|---|---|---|---|---|
| **Metabolite** | **Short name*** | **Other names*** | **Metabolite class*** | **Sub-class*** | **Moiety*** |
| Carnitine | Carnitine | - | Acylcarnitines | - | c0 |
| Acetylcarnitine | Acetylcarnitine | - | Acylcarnitines | Short-chain | c2:0 |
| Propionylcarnitine | Propionylcarnitine | - | Acylcarnitines | Short-chain | c3:0 |
| Propenoylcarnitine | Propenoylcarnitine | - | Acylcarnitines | Unsaturated | c3:1 |
| Hydroxypropionylcarnitine | Hydroxypropionylcarnitine | - | Acylcarnitines | Hydroxyl-/dicarboxyl | c3-OH |
| Butyrylcarnitine | Butyrylcarnitine | - | Acylcarnitines | Short-chain | c4:0 |
| Butenoylcarnitine | Butenoylcarnitine | - | Acylcarnitines | Unsaturated | c4:1 |
| Hydroxybutyrylcarnitine | Hydroxybutyrylcarnitine | - | Acylcarnitines | Hydroxyl-/dicarboxyl | c4-OH |
| Valerylcarnitine | Valerylcarnitine | - | Acylcarnitines | Short-chain | c5:0 |
| Glutarylcarnitine | Glutarylcarnitine | - | Acylcarnitines | Hydroxyl-/dicarboxyl | c5-DC |
| Methylglutarylcarnitine | Methylglutarylcarnitine | - | Acylcarnitines | Hydroxyl-/dicarboxyl | c5-M-DC |
| Hydroxyvalerylcarnitine | Hydroxyvalerylcarnitine | - | Acylcarnitines | Hydroxyl-/dicarboxyl | c5-OH |
| Tiglylcarnitine | Tiglylcarnitine | - | Acylcarnitines | Branched | c5:1 M |
| Glutaconylcarnitine | Glutaconylcarnitine | Mesaconylcarnitine | Acylcarnitines | Hydroxyl-/dicarboxyl | c5:1-DC |
| Hexanoylcarnitine | Hexanoylcarnitine | Caproylcarnitine | Acylcarnitines | Medium-chain | c6:0 |
| Hexenoylcarnitine | Hexenoylcarnitine | - | Acylcarnitines | Unsaturated | c6:1 |
| Pimelylcarnitine | Pimelylcarnitine | - | Acylcarnitines | Hydroxyl-/dicarboxyl | c7-DC |
| Octanoylcarnitine | Octanoylcarnitine | - | Acylcarnitines | Medium-chain | c8:0 |
| Nonanoylcarnitine | Nonanoylcarnitine | - | Acylcarnitines | Medium-chain | c9:0 |
| Decanoylcarnitine | Decanoylcarnitine | - | Acylcarnitines | Medium-chain | c10:0 |
| Decenoylcarnitine | Decenoylcarnitine | - | Acylcarnitines | Unsaturated | c10:1 |
| Decadienoylcarnitine | Decadienoylcarnitine | - | Acylcarnitines | Unsaturated | c10:2 |
| Dodecanoylcarnitine | Dodecanoylcarnitine | Lauroylcarnitine | Acylcarnitines | Medium-chain | c12:0 |
| Dodecenoylcarnitine | Dodecenoylcarnitine | - | Acylcarnitines | Unsaturated | c12:1 |
| Tetradecanoylcarnitine | Tetradecanoylcarnitine | Myristoylcarnitine | Acylcarnitines | Long-chain | c14:0 |
| Tetradecenoylcarnitine | Tetradecenoylcarnitine | - | Acylcarnitines | Unsaturated | c14:1 |
| Hydroxytetradecenoylcarnitine | Hydroxytetradecenoylcarnitine | - | Acylcarnitines | Hydroxyl-/dicarboxyl | c14:1-OH |
| Tetradecadienoylcarnitine | Tetradecadienoylcarnitine | - | Acylcarnitines | Unsaturated | c14:2 |
| Hydroxytetradecadienoylcarnitine | Hydroxytetradecadienoylcarnitine | - | Acylcarnitines | Hydroxyl-/dicarboxyl | c14:2-OH |
| Hexadecanoylcarnitine | Hexadecanoylcarnitine | Palmitoylcarnitine | Acylcarnitines | Long-chain | c16:0 |
| Hydroxyhexadecanolycarnitine | Hydroxyhexadecanolycarnitine | - | Acylcarnitines | Hydroxyl-/dicarboxyl | c16-OH |

| Table S1. Metabolite information | | | | | |
|---|---|---|---|---|---|
| **Metabolite** | **Short name*** | **Other names*** | **Metabolite class*** | **Sub-class*** | **Moiety*** |
| Hexadecenoylcarnitine | Hexadecenoylcarnitine | - | Acylcarnitines | Unsaturated | c16:1 |
| Hydroxyhexadecenoylcarnitine | Hydroxyhexadecenoylcarnitine | - | Acylcarnitines | Hydroxyl-/dicarboxyl | c16:1-OH |
| Hexadecadienoylcarnitine | Hexadecadienoylcarnitine | - | Acylcarnitines | Unsaturated | c16:2 |
| Hydroxyhexadecadienoylcarnitine | Hydroxyhexadecadienoylcarnitine | - | Acylcarnitines | Hydroxyl-/dicarboxyl | c16:2-OH |
| Octadecanoylcarnitine | Octadecanoylcarnitine | Stearoylcarnitine | Acylcarnitines | Long-chain | c18:0 |
| Octadecenoylcarnitine | Octadecenoylcarnitine | - | Acylcarnitines | Unsaturated | c18:1 |
| Hydroxyoctadecenoylcarnitine | Hydroxyoctadecenoylcarnitine | - | Acylcarnitines | Hydroxyl-/dicarboxyl | c18:1-OH |
| Octadecadienoylcarnitine | Octadecadienoylcarnitine | - | Acylcarnitines | Unsaturated | c18:2 |
| Alanine | Alanine | ala | Amino acids | - | - |
| Arginine | Arginine | arg | Amino acids | - | - |
| Asparagine | Asparagine | asn | Amino acids | - | - |
| Aspartate | Aspartate | asp | Amino acids | - | - |
| Citrulline | Citrulline | - | Amino acids | - | - |
| Glutamate | Glutamate | glu | Amino acids | - | - |
| Glutamine | Glutamine | gln | Amino acids | - | - |
| Glycine | Glycine | gly | Amino acids | - | - |
| Histidine | Histidine | his | Amino acids | - | - |
| Isoleucine | Isoleucine | ile | Amino acids | - | - |
| Leucine | Leucine | leu | Amino acids | - | - |
| Lysine | Lysine | lys | Amino acids | - | - |
| Methionine | Methionine | met | Amino acids | - | - |
| Ornithine | Ornithine | orn | Amino acids | - | - |
| Phenylalanine | Phenylalanine | phe | Amino acids | - | - |
| Proline | Proline | pro | Amino acids | - | - |
| Serine | Serine | ser | Amino acids | - | - |
| Threonine | Threonine | thr | Amino acids | - | - |
| Tryptophan | Tryptophan | trp | Amino acids | - | - |
| Tyrosine | Tyrosine | tyr | Amino acids | - | - |
| Valine | Valine | val | Amino acids | - | - |
| Acetylornithine | Acetylornithine | - | Biogenic amines | - | - |

| Table S1. Metabolite information | | | | | |
|---|---|---|---|---|---|
| **Metabolite** | **Short name*** | **Other names*** | **Metabolite class*** | **Sub-class*** | **Moiety*** |
| alpha-Aminoadipic acid | alpha-Aminoadipic acid | - | Biogenic amines | - | - |
| cis-4-Hydroxyproline | cis-4-Hydroxyproline | - | Biogenic amines | - | - |
| Creatinine | Creatinine | - | Biogenic amines | - | - |
| Kynurenine | Kynurenine | - | Biogenic amines | - | - |
| Methionine-Sulfoxide | Methionine-Sulfoxide | - | Biogenic amines | - | - |
| Phenylethylamine | Phenylethylamine | - | Biogenic amines | - | - |
| Sarcosine | Sarcosine | - | Biogenic amines | - | - |
| Serotonin | Serotonin | - | Biogenic amines | - | - |
| Spermidine | Spermidine | - | Biogenic amines | - | - |
| Symmetric dimethylarginine | Symmetric dimethylarginine | - | Biogenic amines | - | - |
| Taurine | Taurine | - | Biogenic amines | - | - |
| trans-4-Hydroxyproline | trans-4-Hydroxyproline | - | Biogenic amines | - | - |
| Lysophosphatidylcholine with acyl residue C14:0 | LPC a C14:0 | - | Lysophosphatidylcholines | - | - |
| Lysophosphatidylcholine with acyl residue C16:0 | LPC a C16:0 | - | Lysophosphatidylcholines | - | - |
| Lysophosphatidylcholine with acyl residue C16:1 | LPC a C16:1 | - | Lysophosphatidylcholines | - | - |
| Lysophosphatidylcholine with acyl residue C17:0 | LPC a C17:0 | - | Lysophosphatidylcholines | - | - |
| Lysophosphatidylcholine with acyl residue C18:0 | LPC a C18:0 | - | Lysophosphatidylcholines | - | - |
| Lysophosphatidylcholine with acyl residue C18:1 | LPC a C18:1 | - | Lysophosphatidylcholines | - | - |
| Lysophosphatidylcholine with acyl residue C18:2 | LPC a C18:2 | - | Lysophosphatidylcholines | - | - |
| Lysophosphatidylcholine with acyl residue C20:3 | LPC a C20:3 | - | Lysophosphatidylcholines | - | - |
| Lysophosphatidylcholine with acyl residue C20:4 | LPC a C20:4 | - | Lysophosphatidylcholines | - | - |
| Lysophosphatidylcholine with acyl residue C24:0 | LPC a C24:0 | - | Lysophosphatidylcholines | - | - |
| Lysophosphatidylcholine with acyl residue C26:0 | LPC a C26:0 | - | Lysophosphatidylcholines | - | - |
| Lysophosphatidylcholine with acyl residue C26:1 | LPC a C26:1 | - | Lysophosphatidylcholines | - | - |
| Lysophosphatidylcholine with acyl residue C28:0 | LPC a C28:0 | - | Lysophosphatidylcholines | - | - |
| Lysophosphatidylcholine with acyl residue C28:1 | LPC a C28:1 | - | Lysophosphatidylcholines | - | - |
| Phosphatidylcholine with diacyl residue sum C24:0 | PC aa C24:0 | - | Phosphatidylcholines | - | - |
| Phosphatidylcholine with diacyl residue sum C26:0 | PC aa C26:0 | - | Phosphatidylcholines | - | - |
| Phosphatidylcholine with diacyl residue sum C28:1 | PC aa C28:1 | - | Phosphatidylcholines | - | - |
| Phosphatidylcholine with diacyl residue sum C30:0 | PC aa C30:0 | - | Phosphatidylcholines | - | - |

| Table S1. Metabolite information | | | | | |
|---|---|---|---|---|---|
| **Metabolite** | **Short name*** | **Other names*** | **Metabolite class*** | **Sub-class*** | **Moiety*** |
| Phosphatidylcholine with diacyl residue sum C32:0 | PC aa C32:0 | - | Phosphatidylcholines | - | - |
| Phosphatidylcholine with diacyl residue sum C32:1 | PC aa C32:1 | - | Phosphatidylcholines | - | - |
| Phosphatidylcholine with diacyl residue sum C32:2 | PC aa C32:2 | - | Phosphatidylcholines | - | - |
| Phosphatidylcholine with diacyl residue sum C32:3 | PC aa C32:3 | - | Phosphatidylcholines | - | - |
| Phosphatidylcholine with diacyl residue sum C34:1 | PC aa C34:1 | - | Phosphatidylcholines | - | - |
| Phosphatidylcholine with diacyl residue sum C34:2 | PC aa C34:2 | - | Phosphatidylcholines | - | - |
| Phosphatidylcholine with diacyl residue sum C34:3 | PC aa C34:3 | - | Phosphatidylcholines | - | - |
| Phosphatidylcholine with diacyl residue sum C34:4 | PC aa C34:4 | - | Phosphatidylcholines | - | - |
| Phosphatidylcholine with diacyl residue sum C36:0 | PC aa C36:0 | - | Phosphatidylcholines | - | - |
| Phosphatidylcholine with diacyl residue sum C36:1 | PC aa C36:1 | - | Phosphatidylcholines | - | - |
| Phosphatidylcholine with diacyl residue sum C36:2 | PC aa C36:2 | - | Phosphatidylcholines | - | - |
| Phosphatidylcholine with diacyl residue sum C36:3 | PC aa C36:3 | - | Phosphatidylcholines | - | - |
| Phosphatidylcholine with diacyl residue sum C36:4 | PC aa C36:4 | - | Phosphatidylcholines | - | - |
| Phosphatidylcholine with diacyl residue sum C36:5 | PC aa C36:5 | - | Phosphatidylcholines | - | - |
| Phosphatidylcholine with diacyl residue sum C36:6 | PC aa C36:6 | - | Phosphatidylcholines | - | - |
| Phosphatidylcholine with diacyl residue sum C38:0 | PC aa C38:0 | - | Phosphatidylcholines | - | - |
| Phosphatidylcholine with diacyl residue sum C38:1 | PC aa C38:1 | - | Phosphatidylcholines | - | - |
| Phosphatidylcholine with diacyl residue sum C38:3 | PC aa C38:3 | - | Phosphatidylcholines | - | - |
| Phosphatidylcholine with diacyl residue sum C38:4 | PC aa C38:4 | - | Phosphatidylcholines | - | - |
| Phosphatidylcholine with diacyl residue sum C38:5 | PC aa C38:5 | - | Phosphatidylcholines | - | - |
| Phosphatidylcholine with diacyl residue sum C38:6 | PC aa C38:6 | - | Phosphatidylcholines | - | - |
| Phosphatidylcholine with diacyl residue sum C40:1 | PC aa C40:1 | - | Phosphatidylcholines | - | - |
| Phosphatidylcholine with diacyl residue sum C40:2 | PC aa C40:2 | - | Phosphatidylcholines | - | - |
| Phosphatidylcholine with diacyl residue sum C40:3 | PC aa C40:3 | - | Phosphatidylcholines | - | - |
| Phosphatidylcholine with diacyl residue sum C40:4 | PC aa C40:4 | - | Phosphatidylcholines | - | - |
| Phosphatidylcholine with diacyl residue sum C40:5 | PC aa C40:5 | - | Phosphatidylcholines | - | - |
| Phosphatidylcholine with diacyl residue sum C40:6 | PC aa C40:6 | - | Phosphatidylcholines | - | - |
| Phosphatidylcholine with diacyl residue sum C42:0 | PC aa C42:0 | - | Phosphatidylcholines | - | - |
| Phosphatidylcholine with diacyl residue sum C42:1 | PC aa C42:1 | - | Phosphatidylcholines | - | - |
| Phosphatidylcholine with diacyl residue sum C42:1 | PC aa C42:1 | - | Phosphatidylcholines | - | - |

| Table S1. Metabolite information | | | | | |
|---|---|---|---|---|---|
| **Metabolite** | **Short name*** | **Other names*** | **Metabolite class*** | **Sub-class*** | **Moiety*** |
| Phosphatidylcholine with diacyl residue sum C42:4 | PC aa C42:4 | - | Phosphatidylcholines | - | - |
| Phosphatidylcholine with diacyl residue sum C42:5 | PC aa C42:5 | - | Phosphatidylcholines | - | - |
| Phosphatidylcholine with diacyl residue sum C42:6 | PC aa C42:6 | - | Phosphatidylcholines | - | - |
| Phosphatidylcholine with acyl-alkyl residue sum C30:0 | PC ae C30:0 | - | Phosphatidylcholines | - | - |
| Phosphatidylcholine with acyl-alkyl residue sum C30:1 | PC ae C30:1 | - | Phosphatidylcholines | - | - |
| Phosphatidylcholine with acyl-alkyl residue sum C30:2 | PC ae C30:2 | - | Phosphatidylcholines | - | - |
| Phosphatidylcholine with acyl-alkyl residue sum C32:1 | PC ae C32:1 | - | Phosphatidylcholines | - | - |
| Phosphatidylcholine with acyl-alkyl residue sum C32:2 | PC ae C32:2 | - | Phosphatidylcholines | - | - |
| Phosphatidylcholine with acyl-alkyl residue sum C34:0 | PC ae C34:0 | - | Phosphatidylcholines | - | - |
| Phosphatidylcholine with acyl-alkyl residue sum C34:1 | PC ae C34:1 | - | Phosphatidylcholines | - | - |
| Phosphatidylcholine with acyl-alkyl residue sum C34:2 | PC ae C34:2 | - | Phosphatidylcholines | - | - |
| Phosphatidylcholine with acyl-alkyl residue sum C34:3 | PC ae C34:3 | - | Phosphatidylcholines | - | - |
| Phosphatidylcholine with acyl-alkyl residue sum C36:0 | PC ae C36:0 | - | Phosphatidylcholines | - | - |
| Phosphatidylcholine with acyl-alkyl residue sum C36:1 | PC ae C36:1 | - | Phosphatidylcholines | - | - |
| Phosphatidylcholine with acyl-alkyl residue sum C36:2 | PC ae C36:2 | - | Phosphatidylcholines | - | - |
| Phosphatidylcholine with acyl-alkyl residue sum C36:3 | PC ae C36:3 | - | Phosphatidylcholines | - | - |
| Phosphatidylcholine with acyl-alkyl residue sum C36:4 | PC ae C36:4 | - | Phosphatidylcholines | - | - |
| Phosphatidylcholine with acyl-alkyl residue sum C36:5 | PC ae C36:5 | - | Phosphatidylcholines | - | - |
| Phosphatidylcholine with acyl-alkyl residue sum C38:0 | PC ae C38:0 | - | Phosphatidylcholines | - | - |
| Phosphatidylcholine with acyl-alkyl residue sum C38:1 | PC ae C38:1 | - | Phosphatidylcholines | - | - |
| Phosphatidylcholine with acyl-alkyl residue sum C38:2 | PC ae C38:2 | - | Phosphatidylcholines | - | - |
| Phosphatidylcholine with acyl-alkyl residue sum C38:3 | PC ae C38:3 | - | Phosphatidylcholines | - | - |
| Phosphatidylcholine with acyl-alkyl residue sum C38:4 | PC ae C38:4 | - | Phosphatidylcholines | - | - |
| Phosphatidylcholine with acyl-alkyl residue sum C38:5 | PC ae C38:5 | - | Phosphatidylcholines | - | - |
| Phosphatidylcholine with acyl-alkyl residue sum C38:6 | PC ae C38:6 | - | Phosphatidylcholines | - | - |
| Phosphatidylcholine with acyl-alkyl residue sum C40:1 | PC ae C40:1 | - | Phosphatidylcholines | - | - |
| Phosphatidylcholine with acyl-alkyl residue sum C40:2 | PC ae C40:2 | - | Phosphatidylcholines | - | - |
| Phosphatidylcholine with acyl-alkyl residue sum C40:3 | PC ae C40:3 | - | Phosphatidylcholines | - | - |
| Phosphatidylcholine with acyl-alkyl residue sum C40:4 | PC ae C40:4 | - | Phosphatidylcholines | - | - |
| Phosphatidylcholine with acyl-alkyl residue sum C40:5 | PC ae C40:5 | - | Phosphatidylcholines | - | - |

| **Table S1. Metabolite information** | | | | | |
|---|---|---|---|---|---|
| **Metabolite** | **Short name*** | **Other names*** | **Metabolite class*** | **Sub-class*** | **Moiety*** |
| Phosphatidylcholine with acyl-alkyl residue sum C40:6 | PC ae C40:6 | - | Phosphatidylcholines | - | - |
| Phosphatidylcholine with acyl-alkyl residue sum C42:0 | PC ae C42:0 | - | Phosphatidylcholines | - | - |
| Phosphatidylcholine with acyl-alkyl residue sum C42:1 | PC ae C42:1 | - | Phosphatidylcholines | - | - |
| Phosphatidylcholine with acyl-alkyl residue sum C42:2 | PC ae C42:2 | - | Phosphatidylcholines | - | - |
| Phosphatidylcholine with acyl-alkyl residue sum C42:3 | PC ae C42:3 | - | Phosphatidylcholines | - | - |
| Phosphatidylcholine with acyl-alkyl residue sum C42:4 | PC ae C42:4 | - | Phosphatidylcholines | - | - |
| Phosphatidylcholine with acyl-alkyl residue sum C42:5 | PC ae C42:5 | - | Phosphatidylcholines | - | - |
| Phosphatidylcholine with acyl-alkyl residue sum C44:5 | PC ae C44:5 | - | Phosphatidylcholines | - | - |
| Phosphatidylcholine with acyl-alkyl residue sum C44:6 | PC ae C44:6 | - | Phosphatidylcholines | - | - |
| Sphingomyelin with acyl residue sum C16:0 | SM C16:0 | - | Sphingomyelins | - | - |
| Sphingomyelin with acyl residue sum C16:1 | SM C16:1 | - | Sphingomyelins | - | - |
| Sphingomyelin with acyl residue sum C18:0 | SM C18:0 | - | Sphingomyelins | - | - |
| Sphingomyelin with acyl residue sum C18:1 | SM C18:1 | - | Sphingomyelins | - | - |
| Sphingomyelin with acyl residue sum C20:2 | SM C20:2 | - | Sphingomyelins | - | - |
| Sphingomyelin with acyl residue sum C24:0 | SM C24:0 | - | Sphingomyelins | - | - |
| Sphingomyelin with acyl residue sum C24:1 | SM C24:1 | - | Sphingomyelins | - | - |
| Hydroxysphingomyelin with acyl residue sum C14:1 | HydroxySM C14:1 | - | Sphingomyelins | - | - |
| Hydroxysphingomyelin with acyl residue sum C16:1 | HydroxySM C16:1 | - | Sphingomyelins | - | - |
| Hydroxysphingomyelin with acyl residue sum C22:1 | HydroxySM C22:1 | - | Sphingomyelins | - | - |
| Hydroxysphingomyelin with acyl residue sum C22:2 | HydroxySM C22:2 | - | Sphingomyelins | - | - |
| Hexose | Hexose | - | Hexoses | - | - |

* Columns: Short name - abbreviations used for the metabolites, Other names - synonyms or abbreviations used in literature for the metabolites,
Metabolite class - class metabolite belongs to, Sub-class - acylcarnitine sub-class, Moiety - specific group of atoms within the acylcarnitine that distinguishes it
Abbreviations: a = acyl residue, aa = diacyl residue, ae = acyl-alkyl residue, LPC = lysophosphatidylcholine, PC = phosphatidylcholine, SM = sphingomyelin
