## Supplemental Table S2 for "Integrating metabolomics and proteomics to identify novel drug targets for heart failure and atrial fibrillation"

**Table S2. Full MR results testing for the effect of metabolites on cardiac outcomes**

| Metabolite* | Metabolite class* | Cardiac outcome | OR (95% CI) | p-value | No. variants* | Model* | Heterogeneity p-value | Q-statistic |
|---|---|---|---|---|---|---|---|---|
| PC ae C44:6 | Phosphatidylcholines | DCM | 1.37 (1.05; 1.80) | $2.2\times10^{-2}$ | 13 | IVW | 0.355 | 13,201 |
| PC ae C44:6 | Phosphatidylcholines | HF | 1.12 (1.06; 1.18) | $3.5\times10^{-5}$ | 12 | IVW | 0.025 | 21,923 |
| PC ae C44:6 | Phosphatidylcholines | AF | 1.08 (1.04; 1.13) | $3.3\times10^{-4}$ | 13 | IVW | <0.001 | 35,884 |
| PC ae C44:6 | Phosphatidylcholines | NICM | 0.66 (0.16; 2.68) | $5.6\times10^{-1}$ | 13 | MR Egger | 0.526 | 10,047 |
| Serotonin | Biogenic amines | DCM | 0.93 (0.48; 1.80) | $8.4\times10^{-1}$ | 7 | IVW | 0.081 | 11,258 |
| Serotonin | Biogenic amines | AF | 0.93 (0.84; 1.02) | $1.3\times10^{-1}$ | 6 | IVW | 0.275 | 6,338 |
| Serotonin | Biogenic amines | NICM | 1.03 (0.71; 1.48) | $8.8\times10^{-1}$ | 7 | IVW | 0.885 | 2,353 |
| PC aa C38:0 | Phosphatidylcholines | DCM | 6.04 (1.52; 23.98) | $1.1\times10^{-2}$ | 8 | MR Egger | 0.981 | 1,105 |
| PC aa C38:0 | Phosphatidylcholines | HF | 0.99 (0.90; 1.09) | $8.9\times10^{-1}$ | 8 | IVW | 0.005 | 20,121 |
| PC aa C38:0 | Phosphatidylcholines | AF | 1.03 (0.92; 1.16) | $6.1\times10^{-1}$ | 8 | IVW | 0.062 | 13,459 |
| PC aa C38:0 | Phosphatidylcholines | NICM | 1.25 (0.82; 1.89) | $3.0\times10^{-1}$ | 8 | IVW | 0.712 | 4,571 |
| Symmetric dimethylarginine | Biogenic amines | DCM | 1.11 (0.61; 2.04) | $7.3\times10^{-1}$ | 6 | IVW | 0.089 | 9,549 |
| Symmetric dimethylarginine | Biogenic amines | AF | 1.04 (0.97; 1.13) | $2.7\times10^{-1}$ | 6 | IVW | 0.468 | 4,59 |
| Symmetric dimethylarginine | Biogenic amines | NICM | 0.84 (0.60; 1.18) | $3.2\times10^{-1}$ | 6 | IVW | 0.882 | 1,752 |
| Tiglylcarnitine | Acylcarnitines | DCM | 1.08 (0.58; 2.04) | $8.0\times10^{-1}$ | 7 | IVW | 0.049 | 12,625 |
| Tiglylcarnitine | Acylcarnitines | AF | 0.82 (0.72; 0.94) | $5.3\times10^{-3}$ | 6 | IVW | 0.247 | 6,665 |
| HydroxySM C22:2 | Sphingomyelins | DCM | 0.87 (0.57; 1.32) | $5.1\times10^{-1}$ | 7 | IVW | 0.358 | 6,617 |
| HydroxySM C22:2 | Sphingomyelins | HF | 1.07 (0.98; 1.17) | $1.2\times10^{-1}$ | 7 | IVW | 0.273 | 7,544 |
| HydroxySM C22:2 | Sphingomyelins | AF | 1.08 (1.01; 1.15) | $3.3\times10^{-2}$ | 7 | IVW | 0.004 | 19,121 |
| HydroxySM C22:2 | Sphingomyelins | NICM | 1.21 (0.86; 1.70) | $2.8\times10^{-1}$ | 7 | IVW | 0.585 | 4,681 |
| Lysine | Amino acids | DCM | 1.29 (0.85; 1.97) | $2.3\times10^{-1}$ | 60 | MR Egger | 0.011 | 85,372 |
| Lysine | Amino acids | HF | 1.01 (0.98; 1.05) | $4.4\times10^{-1}$ | 61 | IVW | 0.005 | 92,292 |
| Lysine | Amino acids | AF | 0.99 (0.96; 1.02) | $4.0\times10^{-1}$ | 60 | IVW | <0.001 | 111,719 |
| Lysine | Amino acids | NICM | 1.64 (1.07; 2.50) | $2.3\times10^{-2}$ | 60 | MR Egger | 0.073 | 74,305 |
| Tetradecanoylcarnitine | Acylcarnitines | DCM | 1.40 (1.09; 1.80) | $9.1\times10^{-3}$ | 33 | IVW | 0.551 | 30,343 |
| Tetradecanoylcarnitine | Acylcarnitines | HF | 1.08 (1.02; 1.15) | $1.1\times10^{-2}$ | 30 | IVW | 0.542 | 27,556 |
| Tetradecanoylcarnitine | Acylcarnitines | AF | 1.12 (1.06; 1.18) | $1.8\times10^{-5}$ | 30 | IVW | 0.012 | 48,95 |
| Tetradecanoylcarnitine | Acylcarnitines | NICM | 0.84 (0.65; 1.09) | $1.9\times10^{-1}$ | 27 | IVW | 0.440 | 26,423 |
| Nonanoylcarnitine | Acylcarnitines | DCM | 0.82 (0.70; 0.97) | $2.1\times10^{-2}$ | 23 | IVW | 0.257 | 25,883 |
| Nonanoylcarnitine | Acylcarnitines | HF | 1.01 (0.97; 1.05) | $5.8\times10^{-1}$ | 22 | IVW | 0.166 | 27,135 |
| Nonanoylcarnitine | Acylcarnitines | AF | 0.99 (0.97; 1.02) | $7.0\times10^{-1}$ | 22 | IVW | 0.342 | 23,034 |

| Table S2. Full MR results testing for the effect of metabolites on cardiac outcomes | | | | | | | | |
|---|---|---|---|---|---|---|---|---|
| **Metabolite*** | **Metabolite class*** | **Cardiac outcome** | **OR (95% CI)** | **p-value** | **No. variants*** | **Model*** | **Heterogeneity p-value** | **Q-statistic** |
| Nonanoylcarnitine | Acylcarnitines | NICM | 0.87 (0.75; 1.02) | $8.0\times10^{-2}$ | 22 | IVW | 0.151 | 27,613 |
| Tetradecenoylcarnitine | Acylcarnitines | DCM | 0.92 (0.68; 1.25) | $6.0\times10^{-1}$ | 29 | IVW | 0.471 | 27,872 |
| Tetradecenoylcarnitine | Acylcarnitines | HF | 1.00 (0.94; 1.07) | $9.1\times10^{-1}$ | 29 | IVW | 0.410 | 29,048 |
| Tetradecenoylcarnitine | Acylcarnitines | AF | 0.97 (0.92; 1.02) | $1.9\times10^{-1}$ | 30 | IVW | 0.042 | 43,368 |
| Tetradecenoylcarnitine | Acylcarnitines | NICM | 0.88 (0.68; 1.14) | $3.3\times10^{-1}$ | 28 | IVW | 0.021 | 43,839 |
| PC ae C38:1 | Phosphatidylcholines | DCM | 1.45 (0.99; 2.12) | $5.5\times10^{-2}$ | 10 | IVW | 0.957 | 3,175 |
| PC ae C38:1 | Phosphatidylcholines | HF | 1.05 (0.97; 1.14) | $2.2\times10^{-1}$ | 10 | IVW | 0.841 | 4,917 |
| PC ae C38:1 | Phosphatidylcholines | AF | 1.01 (0.94; 1.09) | $7.7\times10^{-1}$ | 10 | IVW | 0.246 | 11,444 |
| PC ae C38:1 | Phosphatidylcholines | NICM | 0.93 (0.68; 1.27) | $6.4\times10^{-1}$ | 10 | IVW | 0.696 | 6,434 |
| Proline | Amino acids | DCM | 1.01 (0.85; 1.20) | $9.2\times10^{-1}$ | 49 | MR Egger | <0.001 | 81,126 |
| Proline | Amino acids | HF | 1.03 (0.99; 1.07) | $9.2\times10^{-2}$ | 52 | MR Egger | 0.062 | 66,239 |
| Proline | Amino acids | AF | 1.01 (0.98; 1.04) | $5.1\times10^{-1}$ | 75 | MR Egger | 0.010 | 103,953 |
| Proline | Amino acids | NICM | 1.06 (0.97; 1.15) | $2.3\times10^{-1}$ | 58 | IVW | 0.009 | 85,571 |
| Hexadecanoylcarnitine | Acylcarnitines | DCM | 1.51 (1.28; 1.79) | $8.4\times10^{-7}$ | 63 | IVW | 0.276 | 68,15 |
| Hexadecanoylcarnitine | Acylcarnitines | HF | 1.06 (1.02; 1.09) | $1.7\times10^{-3}$ | 61 | IVW | 0.006 | 90,695 |
| Hexadecanoylcarnitine | Acylcarnitines | AF | 0.99 (0.96; 1.02) | $6.3\times10^{-1}$ | 60 | IVW | 0.002 | 94,711 |
| Hexadecanoylcarnitine | Acylcarnitines | NICM | 1.15 (0.99; 1.34) | $5.9\times10^{-2}$ | 55 | IVW | <0.001 | 103,338 |
| PC ae C42:3 | Phosphatidylcholines | DCM | 1.03 (0.93; 1.14) | $5.6\times10^{-1}$ | 44 | IVW | 0.521 | 41,859 |
| PC ae C42:3 | Phosphatidylcholines | HF | 1.06 (1.04; 1.08) | $1.3\times10^{-9}$ | 43 | IVW | 0.021 | 62,538 |
| PC ae C42:3 | Phosphatidylcholines | AF | 1.04 (1.02; 1.06) | $5.6\times10^{-5}$ | 46 | IVW | 0.062 | 60,406 |
| PC ae C42:3 | Phosphatidylcholines | NICM | 0.92 (0.84; 1.01) | $7.0\times10^{-2}$ | 45 | IVW | 0.182 | 52,34 |
| PC aa C32:1 | Phosphatidylcholines | DCM | 0.98 (0.66; 1.45) | $9.1\times10^{-1}$ | 10 | IVW | 0.511 | 8,23 |
| PC aa C32:1 | Phosphatidylcholines | HF | 0.95 (0.86; 1.04) | $2.6\times10^{-1}$ | 10 | IVW | 0.212 | 12,028 |
| PC aa C32:1 | Phosphatidylcholines | AF | 1.05 (0.96; 1.14) | $2.7\times10^{-1}$ | 10 | IVW | 0.160 | 13,059 |
| PC aa C32:1 | Phosphatidylcholines | NICM | 0.76 (0.53; 1.09) | $1.4\times10^{-1}$ | 10 | IVW | 0.332 | 10,23 |
| PC ae C34:1 | Phosphatidylcholines | DCM | 0.78 (0.53; 1.14) | $2.0\times10^{-1}$ | 10 | IVW | 0.644 | 6,93 |
| PC ae C34:1 | Phosphatidylcholines | HF | 0.93 (0.85; 1.03) | $1.7\times10^{-1}$ | 10 | IVW | 0.133 | 13,708 |
| PC ae C34:1 | Phosphatidylcholines | AF | 1.02 (0.96; 1.09) | $5.2\times10^{-1}$ | 10 | IVW | 0.441 | 8,963 |
| PC ae C34:1 | Phosphatidylcholines | NICM | 1.17 (0.84; 1.63) | $3.6\times10^{-1}$ | 9 | IVW | 0.822 | 4,375 |
| PC ae C44:5 | Phosphatidylcholines | DCM | 0.90 (0.70; 1.15) | $3.9\times10^{-1}$ | 18 | IVW | 0.985 | 6,851 |
| PC ae C44:5 | Phosphatidylcholines | HF | 1.07 (1.01; 1.12) | $1.5\times10^{-2}$ | 16 | IVW | <0.001 | 36,711 |

| Table S2. Full MR results testing for the effect of metabolites on cardiac outcomes | | | | | | | | |
|---|---|---|---|---|---|---|---|---|
| **Metabolite*** | **Metabolite class*** | **Cardiac outcome** | **OR (95% CI)** | **p-value** | **No. variants*** | **Model*** | **Heterogeneity p-value** | **Q-statistic** |
| PC ae C44:5 | Phosphatidylcholines | AF | 1.00 (0.96; 1.05) | $8.4\times10^{-1}$ | 18 | IVW | 0.030 | 29,515 |
| PC ae C44:5 | Phosphatidylcholines | NICM | 0.95 (0.74; 1.22) | $6.8\times10^{-1}$ | 17 | IVW | 0.143 | 22,018 |
| Serine | Amino acids | DCM | 1.00 (0.88; 1.13) | $9.7\times10^{-1}$ | 79 | IVW | 0.015 | 107,609 |
| Serine | Amino acids | HF | 0.90 (0.88; 0.93) | $2.6\times10^{-13}$ | 77 | IVW | 0.324 | 81,059 |
| Serine | Amino acids | AF | 1.01 (0.94; 1.08) | $8.0\times10^{-1}$ | 78 | MR Egger | <0.001 | 122,308 |
| Serine | Amino acids | NICM | 0.84 (0.74; 0.94) | $3.7\times10^{-3}$ | 77 | IVW | 0.154 | 88,566 |
| PC aa C36:0 | Phosphatidylcholines | DCM | 0.99 (0.58; 1.70) | $9.6\times10^{-1}$ | 6 | IVW | 0.515 | 4,241 |
| PC aa C36:0 | Phosphatidylcholines | HF | 0.94 (0.83; 1.06) | $2.9\times10^{-1}$ | 6 | IVW | 0.002 | 19,431 |
| PC aa C36:0 | Phosphatidylcholines | AF | 0.98 (0.89; 1.08) | $6.9\times10^{-1}$ | 6 | IVW | 0.510 | 4,277 |
| LPC a C26:0 | Lysophosphatidylcholines | DCM | 0.50 (0.31; 0.80) | $4.2\times10^{-3}$ | 6 | IVW | 0.129 | 8,535 |
| LPC a C26:0 | Lysophosphatidylcholines | HF | 0.95 (0.86; 1.05) | $3.3\times10^{-1}$ | 6 | IVW | 0.940 | 1,249 |
| LPC a C26:0 | Lysophosphatidylcholines | AF | 0.96 (0.89; 1.03) | $2.8\times10^{-1}$ | 9 | IVW | 0.118 | 12,835 |
| LPC a C26:0 | Lysophosphatidylcholines | NICM | 1.07 (0.64; 1.81) | $7.9\times10^{-1}$ | 7 | IVW | 0.112 | 10,326 |
| Ornithine | Amino acids | DCM | 1.12 (0.86; 1.45) | $4.0\times10^{-1}$ | 38 | IVW | 0.044 | 52,887 |
| Ornithine | Amino acids | HF | 0.98 (0.92; 1.05) | $6.0\times10^{-1}$ | 33 | IVW | 0.034 | 48,119 |
| Ornithine | Amino acids | AF | 0.96 (0.91; 1.01) | $9.1\times10^{-2}$ | 38 | IVW | 0.008 | 60,935 |
| Ornithine | Amino acids | NICM | 1.31 (1.04; 1.66) | $2.0\times10^{-2}$ | 34 | IVW | 0.558 | 31,182 |
| Tryptophan | Amino acids | DCM | 1.78 (1.19; 2.64) | $4.7\times10^{-3}$ | 14 | IVW | 0.086 | 20,386 |
| Tryptophan | Amino acids | HF | 1.15 (1.08; 1.23) | $2.8\times10^{-5}$ | 12 | IVW | 0.832 | 6,574 |
| Tryptophan | Amino acids | AF | 1.14 (1.05; 1.24) | $1.7\times10^{-3}$ | 13 | IVW | 0.125 | 17,718 |
| Tryptophan | Amino acids | NICM | 1.01 (0.78; 1.31) | $9.4\times10^{-1}$ | 14 | IVW | 0.502 | 12,313 |
| LPC a C20:3 | Lysophosphatidylcholines | DCM | 1.09 (0.91; 1.31) | $3.7\times10^{-1}$ | 32 | IVW | 0.077 | 42,792 |
| LPC a C20:3 | Lysophosphatidylcholines | HF | 0.98 (0.95; 1.01) | $2.2\times10^{-1}$ | 33 | IVW | 0.318 | 35,238 |
| LPC a C20:3 | Lysophosphatidylcholines | AF | 0.93 (0.91; 0.96) | $2.3\times10^{-7}$ | 42 | IVW | 0.094 | 53,299 |
| LPC a C20:3 | Lysophosphatidylcholines | NICM | 1.06 (0.95; 1.18) | $2.9\times10^{-1}$ | 37 | IVW | 0.027 | 54,017 |
| Butyrylcarnitine | Acylcarnitines | DCM | 0.87 (0.82; 0.93) | $4.9\times10^{-5}$ | 242 | MR Egger | <0.001 | 423,354 |
| Butyrylcarnitine | Acylcarnitines | HF | 1.00 (0.99; 1.01) | $5.1\times10^{-1}$ | 211 | IVW | <0.001 | 397,085 |
| Butyrylcarnitine | Acylcarnitines | AF | 1.00 (0.99; 1.02) | $7.8\times10^{-1}$ | 230 | MR Egger | <0.001 | 440,308 |
| Butyrylcarnitine | Acylcarnitines | NICM | 1.04 (1.00; 1.08) | $2.8\times10^{-2}$ | 226 | IVW | <0.001 | 375,737 |
| PC aa C42:0 | Phosphatidylcholines | DCM | 1.05 (0.56; 1.95) | $8.8\times10^{-1}$ | 8 | IVW | 0.079 | 12,747 |
| PC aa C42:0 | Phosphatidylcholines | HF | 1.03 (0.94; 1.13) | $5.1\times10^{-1}$ | 7 | IVW | 0.006 | 18,099 |

| Table S2. Full MR results testing for the effect of metabolites on cardiac outcomes | | | | | | | | |
|---|---|---|---|---|---|---|---|---|
| **Metabolite*** | **Metabolite class*** | **Cardiac outcome** | **OR (95% CI)** | **p-value** | **No. variants*** | **Model*** | **Heterogeneity p-value** | **Q-statistic** |
| PC aa C42:0 | Phosphatidylcholines | AF | 1.02 (0.95; 1.11) | $5.5\times10^{-1}$ | 9 | IVW | 0.244 | 10,308 |
| PC aa C42:0 | Phosphatidylcholines | NICM | 0.80 (0.55; 1.17) | $2.5\times10^{-1}$ | 9 | IVW | 0.202 | 11,001 |
| SM C18:1 | Sphingomyelins | DCM | 1.31 (0.99; 1.75) | $6.1\times10^{-2}$ | 21 | IVW | 0.163 | 26,077 |
| SM C18:1 | Sphingomyelins | HF | 0.96 (0.91; 1.01) | $9.6\times10^{-2}$ | 22 | IVW | 0.024 | 35,677 |
| SM C18:1 | Sphingomyelins | AF | 1.02 (0.98; 1.06) | $3.7\times10^{-1}$ | 23 | IVW | 0.011 | 39,972 |
| SM C18:1 | Sphingomyelins | NICM | 1.02 (0.83; 1.25) | $8.4\times10^{-1}$ | 23 | IVW | 0.917 | 13,551 |
| PC ae C36:3 | Phosphatidylcholines | DCM | 1.24 (0.87; 1.76) | $2.4\times10^{-1}$ | 28 | MR Egger | 0.546 | 24,53 |
| PC ae C36:3 | Phosphatidylcholines | HF | 0.94 (0.87; 1.01) | $8.4\times10^{-2}$ | 28 | MR Egger | 0.009 | 46,175 |
| PC ae C36:3 | Phosphatidylcholines | AF | 0.91 (0.86; 0.96) | $1.2\times10^{-3}$ | 30 | MR Egger | 0.003 | 52,888 |
| PC ae C36:3 | Phosphatidylcholines | NICM | 0.83 (0.60; 1.14) | $2.5\times10^{-1}$ | 29 | MR Egger | 0.376 | 28,687 |
| LPC a C14:0 | Lysophosphatidylcholines | DCM | 0.74 (0.46; 1.18) | $2.0\times10^{-1}$ | 7 | IVW | 0.166 | 9,13 |
| LPC a C14:0 | Lysophosphatidylcholines | HF | 1.02 (0.94; 1.11) | $5.8\times10^{-1}$ | 6 | IVW | 0.509 | 4,288 |
| LPC a C14:0 | Lysophosphatidylcholines | AF | 1.01 (0.95; 1.09) | $7.1\times10^{-1}$ | 7 | IVW | 0.622 | 4,403 |
| LPC a C14:0 | Lysophosphatidylcholines | NICM | 0.84 (0.59; 1.21) | $3.5\times10^{-1}$ | 6 | IVW | 0.387 | 5,24 |
| Acetylcarnitine | Acylcarnitines | DCM | 1.09 (0.91; 1.31) | $3.5\times10^{-1}$ | 57 | IVW | 0.106 | 69,488 |
| Acetylcarnitine | Acylcarnitines | HF | 1.10 (1.06; 1.14) | $1.5\times10^{-7}$ | 52 | IVW | 0.382 | 53,411 |
| Acetylcarnitine | Acylcarnitines | AF | 1.06 (1.02; 1.10) | $1.4\times10^{-3}$ | 55 | IVW | 0.088 | 68,532 |
| Acetylcarnitine | Acylcarnitines | NICM | 0.99 (0.86; 1.14) | $9.1\times10^{-1}$ | 53 | IVW | 0.004 | 82,915 |
| SM C16:0 | Sphingomyelins | DCM | 0.97 (0.82; 1.15) | $7.5\times10^{-1}$ | 78 | IVW | 0.020 | 104,7 |
| SM C16:0 | Sphingomyelins | HF | 1.10 (1.06; 1.14) | $3.3\times10^{-6}$ | 67 | IVW | <0.001 | 120,696 |
| SM C16:0 | Sphingomyelins | AF | 1.09 (1.05; 1.12) | $1.4\times10^{-7}$ | 78 | IVW | <0.001 | 146,558 |
| SM C16:0 | Sphingomyelins | NICM | 0.93 (0.81; 1.07) | $3.2\times10^{-1}$ | 76 | IVW | 0.552 | 72,744 |
| Threonine | Amino acids | DCM | 1.08 (0.87; 1.34) | $4.6\times10^{-1}$ | 50 | IVW | <0.001 | 86,029 |
| Threonine | Amino acids | HF | 0.93 (0.89; 0.97) | $7.5\times10^{-4}$ | 49 | IVW | 0.032 | 67,757 |
| Threonine | Amino acids | AF | 0.96 (0.92; 1.00) | $2.9\times10^{-2}$ | 52 | IVW | 0.010 | 77,576 |
| Threonine | Amino acids | NICM | 0.75 (0.63; 0.91) | $2.4\times10^{-3}$ | 53 | IVW | 0.561 | 49,792 |
| PC aa C34:2 | Phosphatidylcholines | DCM | 1.11 (0.91; 1.36) | $3.1\times10^{-1}$ | 30 | IVW | 0.013 | 48,621 |
| PC aa C34:2 | Phosphatidylcholines | HF | 0.78 (0.67; 0.90) | $5.7\times10^{-4}$ | 31 | MR Egger | 0.005 | 52,587 |
| PC aa C34:2 | Phosphatidylcholines | AF | 0.95 (0.92; 0.99) | $1.4\times10^{-2}$ | 29 | IVW | 0.406 | 29,136 |
| PC aa C34:2 | Phosphatidylcholines | NICM | 1.00 (0.84; 1.19) | $1.0\times10^{0}$ | 30 | IVW | 0.835 | 21,634 |
| PC aa C32:0 | Phosphatidylcholines | DCM | 1.12 (0.80; 1.57) | $4.9\times10^{-1}$ | 20 | IVW | 0.134 | 25,854 |

| Table S2. Full MR results testing for the effect of metabolites on cardiac outcomes | | | | | | | | |
|---|---|---|---|---|---|---|---|---|
| **Metabolite*** | **Metabolite class*** | **Cardiac outcome** | **OR (95% CI)** | **p-value** | **No. variants*** | **Model*** | **Heterogeneity p-value** | **Q-statistic** |
| PC aa C32:0 | Phosphatidylcholines | HF | 1.06 (1.00; 1.14) | $6.5\times10^{-2}$ | 18 | IVW | 0.285 | 19,791 |
| PC aa C32:0 | Phosphatidylcholines | AF | 1.09 (1.03; 1.16) | $2.6\times10^{-3}$ | 20 | IVW | 0.104 | 27,03 |
| PC aa C32:0 | Phosphatidylcholines | NICM | 1.04 (0.84; 1.29) | $6.9\times10^{-1}$ | 20 | IVW | 0.729 | 14,897 |
| PC ae C38:2 | Phosphatidylcholines | DCM | 3.20 (1.55; 6.63) | $1.7\times10^{-3}$ | 25 | MR Egger | 0.044 | 35,717 |
| PC ae C38:2 | Phosphatidylcholines | HF | 0.98 (0.94; 1.02) | $3.5\times10^{-1}$ | 24 | IVW | 0.007 | 42,875 |
| PC ae C38:2 | Phosphatidylcholines | AF | 0.93 (0.90; 0.97) | $1.3\times10^{-4}$ | 24 | IVW | 0.002 | 47,791 |
| PC ae C38:2 | Phosphatidylcholines | NICM | 0.96 (0.82; 1.13) | $6.4\times10^{-1}$ | 25 | IVW | 0.898 | 15,721 |
| Octadecadienoylcarnitine | Acylcarnitines | DCM | 1.38 (1.22; 1.55) | $2.2\times10^{-7}$ | 63 | IVW | 0.013 | 89,317 |
| Octadecadienoylcarnitine | Acylcarnitines | HF | 0.93 (0.86; 1.01) | $8.1\times10^{-2}$ | 61 | MR Egger | 0.027 | 81,682 |
| Octadecadienoylcarnitine | Acylcarnitines | AF | 0.97 (0.91; 1.04) | $4.3\times10^{-1}$ | 62 | MR Egger | <0.001 | 102,967 |
| Octadecadienoylcarnitine | Acylcarnitines | NICM | 1.05 (0.76; 1.45) | $7.8\times10^{-1}$ | 57 | MR Egger | 0.005 | 85,885 |
| Hexanoylcarnitine | Acylcarnitines | DCM | 1.08 (0.96; 1.22) | $2.0\times10^{-1}$ | 31 | IVW | 0.662 | 26,252 |
| Hexanoylcarnitine | Acylcarnitines | HF | 0.94 (0.89; 0.99) | $2.0\times10^{-2}$ | 29 | MR Egger | <0.001 | 68,771 |
| Hexanoylcarnitine | Acylcarnitines | AF | 1.01 (0.99; 1.03) | $3.8\times10^{-1}$ | 32 | IVW | 0.253 | 35,802 |
| Hexanoylcarnitine | Acylcarnitines | NICM | 0.79 (0.64; 0.98) | $3.3\times10^{-2}$ | 28 | MR Egger | 0.648 | 22,741 |
| PC aa C42:6 | Phosphatidylcholines | DCM | 1.13 (0.84; 1.53) | $4.1\times10^{-1}$ | 13 | IVW | 0.326 | 13,615 |
| PC aa C42:6 | Phosphatidylcholines | HF | 1.10 (1.04; 1.17) | $6.3\times10^{-4}$ | 12 | IVW | 0.021 | 22,464 |
| PC aa C42:6 | Phosphatidylcholines | AF | 0.76 (0.68; 0.85) | $2.7\times10^{-6}$ | 14 | MR Egger | 0.472 | 11,676 |
| PC aa C42:6 | Phosphatidylcholines | NICM | 1.03 (0.81; 1.31) | $8.2\times10^{-1}$ | 12 | IVW | 0.776 | 7,279 |
| Methionine-Sulfoxide | Biogenic amines | DCM | 0.94 (0.61; 1.45) | $7.9\times10^{-1}$ | 13 | IVW | 0.406 | 12,51 |
| Methionine-Sulfoxide | Biogenic amines | HF | 1.02 (0.91; 1.15) | $7.1\times10^{-1}$ | 12 | IVW | 0.122 | 16,539 |
| Methionine-Sulfoxide | Biogenic amines | AF | 1.08 (1.00; 1.17) | $6.5\times10^{-2}$ | 13 | IVW | 0.013 | 25,492 |
| Methionine-Sulfoxide | Biogenic amines | NICM | 0.94 (0.67; 1.34) | $7.5\times10^{-1}$ | 12 | IVW | 0.047 | 19,9 |
| LPC a C26:1 | Lysophosphatidylcholines | DCM | 1.11 (0.97; 1.26) | $1.3\times10^{-1}$ | 26 | IVW | 0.480 | 24,688 |
| LPC a C26:1 | Lysophosphatidylcholines | HF | 1.06 (1.03; 1.09) | $1.2\times10^{-4}$ | 25 | IVW | 0.258 | 28,052 |
| LPC a C26:1 | Lysophosphatidylcholines | AF | 1.06 (1.03; 1.09) | $6.8\times10^{-5}$ | 25 | IVW | 0.134 | 31,714 |
| LPC a C26:1 | Lysophosphatidylcholines | NICM | 1.56 (1.12; 2.19) | $8.7\times10^{-3}$ | 25 | MR Egger | 0.002 | 47,531 |
| LPC a C18:0 | Lysophosphatidylcholines | DCM | 1.32 (0.93; 1.85) | $1.2\times10^{-1}$ | 17 | IVW | 0.428 | 16,362 |
| LPC a C18:0 | Lysophosphatidylcholines | HF | 1.08 (1.01; 1.15) | $2.7\times10^{-2}$ | 17 | IVW | 0.538 | 14,819 |
| LPC a C18:0 | Lysophosphatidylcholines | AF | 0.99 (0.92; 1.05) | $6.8\times10^{-1}$ | 17 | IVW | 0.181 | 20,948 |
| LPC a C18:0 | Lysophosphatidylcholines | NICM | 1.12 (0.84; 1.50) | $4.4\times10^{-1}$ | 18 | IVW | 0.321 | 19,141 |

| Table S2. Full MR results testing for the effect of metabolites on cardiac outcomes | | | | | | | | |
|---|---|---|---|---|---|---|---|---|
| **Metabolite*** | **Metabolite class*** | **Cardiac outcome** | **OR (95% CI)** | **p-value** | **No. variants*** | **Model*** | **Heterogeneity p-value** | **Q-statistic** |
| SM C24:1 | Sphingomyelins | DCM | 0.90 (0.59; 1.37) | $6.3\times10^{-1}$ | 9 | IVW | 0.626 | 6,186 |
| SM C24:1 | Sphingomyelins | HF | 1.05 (0.93; 1.18) | $4.6\times10^{-1}$ | 7 | IVW | 0.172 | 9,029 |
| SM C24:1 | Sphingomyelins | AF | 1.03 (0.94; 1.13) | $4.9\times10^{-1}$ | 9 | IVW | 0.189 | 11,232 |
| SM C24:1 | Sphingomyelins | NICM | 1.12 (0.79; 1.58) | $5.3\times10^{-1}$ | 9 | IVW | 0.744 | 5,13 |
| Aspartate | Amino acids | DCM | 1.61 (1.30; 2.00) | $1.3\times10^{-5}$ | 36 | IVW | 0.002 | 64,476 |
| Aspartate | Amino acids | HF | 1.07 (1.02; 1.11) | $3.2\times10^{-3}$ | 35 | IVW | 0.185 | 41,191 |
| Aspartate | Amino acids | AF | 1.29 (1.15; 1.43) | $5.3\times10^{-6}$ | 39 | MR Egger | 0.002 | 66,146 |
| Aspartate | Amino acids | NICM | 0.83 (0.70; 0.97) | $1.9\times10^{-2}$ | 37 | IVW | 0.161 | 44,317 |
| PC aa C32:2 | Phosphatidylcholines | DCM | 0.95 (0.70; 1.28) | $7.4\times10^{-1}$ | 17 | IVW | 0.063 | 25,435 |
| PC aa C32:2 | Phosphatidylcholines | HF | 0.99 (0.94; 1.05) | $8.1\times10^{-1}$ | 16 | IVW | 0.102 | 22,208 |
| PC aa C32:2 | Phosphatidylcholines | AF | 1.00 (0.95; 1.05) | $9.3\times10^{-1}$ | 16 | IVW | 0.175 | 19,92 |
| PC aa C32:2 | Phosphatidylcholines | NICM | 0.96 (0.79; 1.18) | $7.1\times10^{-1}$ | 17 | IVW | 0.015 | 30,681 |
| LPC a C24:0 | Lysophosphatidylcholines | AF | 1.03 (0.97; 1.10) | $3.6\times10^{-1}$ | 6 | IVW | 0.856 | 1,953 |
| Dodecanoylcarnitine | Acylcarnitines | DCM | 1.43 (1.11; 1.85) | $5.4\times10^{-3}$ | 31 | IVW | 0.228 | 35,421 |
| Dodecanoylcarnitine | Acylcarnitines | HF | 0.93 (0.89; 0.98) | $8.2\times10^{-3}$ | 32 | IVW | 0.014 | 50,754 |
| Dodecanoylcarnitine | Acylcarnitines | AF | 0.94 (0.90; 0.99) | $2.0\times10^{-2}$ | 31 | IVW | 0.004 | 54,531 |
| Dodecanoylcarnitine | Acylcarnitines | NICM | 0.86 (0.69; 1.08) | $1.9\times10^{-1}$ | 31 | IVW | 0.370 | 31,953 |
| PC ae C36:4 | Phosphatidylcholines | DCM | 1.05 (0.85; 1.28) | $6.7\times10^{-1}$ | 31 | IVW | 0.040 | 44,877 |
| PC ae C36:4 | Phosphatidylcholines | HF | 1.02 (0.99; 1.06) | $2.1\times10^{-1}$ | 31 | IVW | <0.001 | 64,226 |
| PC ae C36:4 | Phosphatidylcholines | AF | 1.00 (0.97; 1.03) | $9.4\times10^{-1}$ | 30 | IVW | 0.888 | 20,158 |
| PC ae C36:4 | Phosphatidylcholines | NICM | 1.20 (0.98; 1.46) | $7.9\times10^{-2}$ | 31 | IVW | 0.195 | 36,41 |
| PC aa C36:6 | Phosphatidylcholines | DCM | 1.00 (0.76; 1.32) | $9.9\times10^{-1}$ | 18 | IVW | 0.367 | 18,349 |
| PC aa C36:6 | Phosphatidylcholines | HF | 0.81 (0.69; 0.96) | $1.7\times10^{-2}$ | 18 | MR Egger | 0.029 | 28,336 |
| PC aa C36:6 | Phosphatidylcholines | AF | 1.00 (0.96; 1.05) | $8.2\times10^{-1}$ | 19 | IVW | 0.684 | 14,682 |
| PC aa C36:6 | Phosphatidylcholines | NICM | 1.06 (0.85; 1.32) | $6.2\times10^{-1}$ | 18 | IVW | 0.773 | 12,431 |
| PC aa C38:3 | Phosphatidylcholines | DCM | 1.91 (1.04; 3.49) | $3.7\times10^{-2}$ | 28 | MR Egger | 0.062 | 37,868 |
| PC aa C38:3 | Phosphatidylcholines | HF | 0.99 (0.95; 1.03) | $6.4\times10^{-1}$ | 32 | IVW | 0.006 | 54,505 |
| PC aa C38:3 | Phosphatidylcholines | AF | 0.96 (0.93; 0.99) | $1.0\times10^{-2}$ | 36 | IVW | 0.003 | 62,19 |
| PC aa C38:3 | Phosphatidylcholines | NICM | 0.99 (0.84; 1.18) | $9.4\times10^{-1}$ | 32 | IVW | 0.125 | 40,179 |
| LPC a C28:1 | Lysophosphatidylcholines | DCM | 1.32 (0.86; 2.04) | $2.1\times10^{-1}$ | 8 | IVW | 0.122 | 11,404 |
| LPC a C28:1 | Lysophosphatidylcholines | HF | 1.44 (1.03; 2.00) | $3.1\times10^{-2}$ | 7 | MR Egger | 0.514 | 4,248 |

| Table S2. Full MR results testing for the effect of metabolites on cardiac outcomes | | | | | | | | |
|---|---|---|---|---|---|---|---|---|
| Metabolite* | Metabolite class* | Cardiac outcome | OR (95% CI) | p-value | No. variants* | Model* | Heterogeneity p-value | Q-statistic |
| LPC a C28:1 | Lysophosphatidylcholines | AF | 1.13 (1.07; 1.20) | $1.5\times10^{-5}$ | 8 | IVW | 0.507 | 6,287 |
| LPC a C28:1 | Lysophosphatidylcholines | NICM | 0.90 (0.67; 1.19) | $4.5\times10^{-1}$ | 8 | IVW | 0.503 | 6,321 |
| PC ae C38:4 | Phosphatidylcholines | DCM | 1.19 (1.05; 1.34) | $5.3\times10^{-3}$ | 42 | IVW | 0.005 | 68,102 |
| PC ae C38:4 | Phosphatidylcholines | HF | 1.19 (1.10; 1.29) | $1.4\times10^{-5}$ | 42 | MR Egger | 0.084 | 52,863 |
| PC ae C38:4 | Phosphatidylcholines | AF | 1.06 (1.04; 1.08) | $2.4\times10^{-7}$ | 41 | IVW | 0.021 | 60,295 |
| PC ae C38:4 | Phosphatidylcholines | NICM | 1.44 (1.03; 2.01) | $3.2\times10^{-2}$ | 43 | MR Egger | 0.104 | 52,733 |
| Arginine | Amino acids | DCM | 0.82 (0.68; 0.97) | $2.4\times10^{-2}$ | 59 | IVW | 0.040 | 78,15 |
| Arginine | Amino acids | HF | 1.01 (0.97; 1.05) | $5.7\times10^{-1}$ | 59 | IVW | 0.520 | 56,79 |
| Arginine | Amino acids | AF | 1.04 (1.01; 1.08) | $1.3\times10^{-2}$ | 62 | IVW | 0.306 | 66,089 |
| Arginine | Amino acids | NICM | 0.89 (0.75; 1.07) | $2.1\times10^{-1}$ | 59 | IVW | 0.213 | 66,267 |
| Carnitine | Acylcarnitines | DCM | 1.09 (0.98; 1.22) | $9.7\times10^{-2}$ | 142 | IVW | <0.001 | 213,333 |
| Carnitine | Acylcarnitines | HF | 1.03 (1.01; 1.05) | $1.3\times10^{-2}$ | 130 | IVW | 0.009 | 169,85 |
| Carnitine | Acylcarnitines | AF | 1.03 (1.01; 1.05) | $4.6\times10^{-3}$ | 145 | IVW | <0.001 | 207,781 |
| Carnitine | Acylcarnitines | NICM | 0.89 (0.82; 0.97) | $7.1\times10^{-3}$ | 137 | IVW | <0.001 | 293,455 |
| SM C18:0 | Sphingomyelins | DCM | 1.27 (1.01; 1.58) | $3.7\times10^{-2}$ | 65 | IVW | 0.051 | 83,506 |
| SM C18:0 | Sphingomyelins | HF | 1.07 (0.97; 1.19) | $1.7\times10^{-1}$ | 59 | MR Egger | <0.001 | 120,738 |
| SM C18:0 | Sphingomyelins | AF | 1.06 (1.02; 1.09) | $2.2\times10^{-3}$ | 63 | IVW | <0.001 | 106,889 |
| SM C18:0 | Sphingomyelins | NICM | 0.98 (0.84; 1.14) | $7.8\times10^{-1}$ | 66 | IVW | 0.181 | 75,228 |
| PC ae C38:3 | Phosphatidylcholines | DCM | 1.14 (0.87; 1.51) | $3.5\times10^{-1}$ | 15 | IVW | 0.332 | 15,697 |
| PC ae C38:3 | Phosphatidylcholines | HF | 0.97 (0.90; 1.03) | $3.1\times10^{-1}$ | 15 | IVW | 0.065 | 22,738 |
| PC ae C38:3 | Phosphatidylcholines | AF | 0.86 (0.83; 0.90) | $4.7\times10^{-11}$ | 15 | IVW | 0.022 | 26,53 |
| PC ae C38:3 | Phosphatidylcholines | NICM | 0.89 (0.72; 1.10) | $2.8\times10^{-1}$ | 15 | IVW | 0.009 | 29,309 |
| Acetylornithine | Biogenic amines | DCM | 1.01 (0.94; 1.09) | $7.0\times10^{-1}$ | 89 | IVW | <0.001 | 134,908 |
| Acetylornithine | Biogenic amines | HF | 0.99 (0.98; 1.00) | $1.3\times10^{-1}$ | 101 | IVW | <0.001 | 237,084 |
| Acetylornithine | Biogenic amines | AF | 0.97 (0.96; 0.98) | $9.9\times10^{-8}$ | 134 | IVW | <0.001 | 212,197 |
| Acetylornithine | Biogenic amines | NICM | 1.11 (1.05; 1.18) | $5.2\times10^{-4}$ | 118 | IVW | 0.109 | 136,145 |
| PC ae C32:2 | Phosphatidylcholines | DCM | 1.13 (0.86; 1.48) | $4.0\times10^{-1}$ | 15 | IVW | 0.883 | 8,124 |
| PC ae C32:2 | Phosphatidylcholines | HF | 0.99 (0.92; 1.06) | $7.5\times10^{-1}$ | 15 | IVW | 0.082 | 21,854 |
| PC ae C32:2 | Phosphatidylcholines | AF | 1.04 (0.99; 1.09) | $1.5\times10^{-1}$ | 15 | IVW | 0.023 | 26,367 |
| PC ae C32:2 | Phosphatidylcholines | NICM | 0.94 (0.73; 1.20) | $5.9\times10^{-1}$ | 15 | IVW | 0.386 | 14,892 |
| Propionylcarnitine | Acylcarnitines | DCM | 1.05 (0.81; 1.37) | $7.2\times10^{-1}$ | 100 | MR Egger | 0.005 | 137,546 |

| Table S2. Full MR results testing for the effect of metabolites on cardiac outcomes | | | | | | | | |
|---|---|---|---|---|---|---|---|---|
| **Metabolite*** | **Metabolite class*** | **Cardiac outcome** | **OR (95% CI)** | **p-value** | **No. variants*** | **Model*** | **Heterogeneity p-value** | **Q-statistic** |
| Propionylcarnitine | Acylcarnitines | HF | 0.99 (0.94; 1.05) | $7.6\times10^{-1}$ | 93 | MR Egger | <0.001 | 161,37 |
| Propionylcarnitine | Acylcarnitines | AF | 1.06 (1.03; 1.08) | $6.1\times10^{-7}$ | 99 | IVW | <0.001 | 176,849 |
| Propionylcarnitine | Acylcarnitines | NICM | 0.80 (0.65; 0.99) | $4.3\times10^{-2}$ | 91 | MR Egger | 0.002 | 131,429 |
| PC aa C40:3 | Phosphatidylcholines | DCM | 1.18 (0.77; 1.80) | $4.4\times10^{-1}$ | 7 | IVW | 0.876 | 2,434 |
| PC aa C40:3 | Phosphatidylcholines | HF | 0.89 (0.72; 1.10) | $2.8\times10^{-1}$ | 7 | MR Egger | 0.936 | 1,29 |
| PC aa C40:3 | Phosphatidylcholines | AF | 1.04 (0.98; 1.12) | $2.0\times10^{-1}$ | 8 | IVW | 0.361 | 7,682 |
| PC aa C40:3 | Phosphatidylcholines | NICM | 1.18 (0.84; 1.66) | $3.4\times10^{-1}$ | 8 | IVW | 0.363 | 7,665 |
| LPC a C18:1 | Lysophosphatidylcholines | DCM | 1.20 (0.83; 1.72) | $3.3\times10^{-1}$ | 7 | IVW | 0.005 | 18,656 |
| LPC a C18:1 | Lysophosphatidylcholines | HF | 1.06 (0.99; 1.14) | $1.2\times10^{-1}$ | 8 | IVW | 0.729 | 4,428 |
| LPC a C18:1 | Lysophosphatidylcholines | AF | 0.93 (0.88; 0.99) | $2.6\times10^{-2}$ | 8 | IVW | 0.757 | 4,195 |
| LPC a C18:1 | Lysophosphatidylcholines | NICM | 1.20 (0.90; 1.60) | $2.0\times10^{-1}$ | 8 | IVW | 0.661 | 4,991 |
| PC ae C40:3 | Phosphatidylcholines | DCM | 1.34 (0.96; 1.88) | $8.9\times10^{-2}$ | 12 | IVW | 0.864 | 6,131 |
| PC ae C40:3 | Phosphatidylcholines | HF | 1.00 (0.94; 1.06) | $9.3\times10^{-1}$ | 14 | IVW | 0.032 | 23,881 |
| PC ae C40:3 | Phosphatidylcholines | AF | 0.95 (0.90; 1.00) | $6.0\times10^{-2}$ | 14 | IVW | 0.763 | 9,127 |
| PC ae C40:3 | Phosphatidylcholines | NICM | 1.12 (0.90; 1.39) | $3.2\times10^{-1}$ | 13 | IVW | 0.443 | 12,034 |
| PC aa C40:2 | Phosphatidylcholines | HF | 1.12 (1.03; 1.22) | $6.7\times10^{-3}$ | 6 | IVW | 0.708 | 2,946 |
| PC aa C40:2 | Phosphatidylcholines | AF | 1.05 (0.95; 1.15) | $3.6\times10^{-1}$ | 6 | IVW | 0.093 | 9,423 |
| PC aa C40:2 | Phosphatidylcholines | NICM | 1.41 (0.99; 2.01) | $6.0\times10^{-2}$ | 6 | IVW | 0.995 | 0,428 |
| LPC a C17:0 | Lysophosphatidylcholines | DCM | 23.97 (2.14; 269.02) | $1.0\times10^{-2}$ | 6 | MR Egger | 0.944 | 0,758 |
| LPC a C17:0 | Lysophosphatidylcholines | HF | 1.07 (0.98; 1.16) | $1.4\times10^{-1}$ | 6 | IVW | 0.419 | 4,977 |
| LPC a C17:0 | Lysophosphatidylcholines | AF | 0.94 (0.87; 1.01) | $6.9\times10^{-2}$ | 6 | IVW | 0.490 | 4,425 |
| LPC a C17:0 | Lysophosphatidylcholines | NICM | 0.89 (0.62; 1.28) | $5.3\times10^{-1}$ | 6 | IVW | 0.034 | 12,071 |
| PC aa C38:6 | Phosphatidylcholines | DCM | 1.30 (0.99; 1.70) | $5.9\times10^{-2}$ | 23 | IVW | 0.058 | 33,26 |
| PC aa C38:6 | Phosphatidylcholines | HF | 1.14 (1.09; 1.20) | $7.0\times10^{-9}$ | 22 | IVW | <0.001 | 46,167 |
| PC aa C38:6 | Phosphatidylcholines | AF | 1.05 (1.01; 1.09) | $2.4\times10^{-2}$ | 22 | IVW | 0.045 | 33,138 |
| PC aa C38:6 | Phosphatidylcholines | NICM | 0.98 (0.81; 1.18) | $8.4\times10^{-1}$ | 23 | IVW | 0.581 | 20,035 |
| PC aa C40:4 | Phosphatidylcholines | DCM | 1.14 (0.95; 1.37) | $1.5\times10^{-1}$ | 23 | IVW | 0.475 | 21,747 |
| PC aa C40:4 | Phosphatidylcholines | HF | 1.12 (1.08; 1.15) | $1.1\times10^{-10}$ | 24 | IVW | 0.568 | 21,209 |
| PC aa C40:4 | Phosphatidylcholines | AF | 0.96 (0.87; 1.05) | $3.4\times10^{-1}$ | 24 | MR Egger | 0.050 | 33,891 |
| PC aa C40:4 | Phosphatidylcholines | NICM | 1.53 (1.06; 2.20) | $2.3\times10^{-2}$ | 25 | MR Egger | 0.507 | 22,218 |
| trans-4-Hydroxyproline | Biogenic amines | DCM | 0.63 (0.36; 1.12) | $1.1\times10^{-1}$ | 7 | IVW | 0.022 | 14,78 |

| Table S2. Full MR results testing for the effect of metabolites on cardiac outcomes | | | | | | | | |
|---|---|---|---|---|---|---|---|---|
| **Metabolite*** | **Metabolite class*** | **Cardiac outcome** | **OR (95% CI)** | **p-value** | **No. variants*** | **Model*** | **Heterogeneity p-value** | **Q-statistic** |
| trans-4-Hydroxyproline | Biogenic amines | HF | 1.03 (0.91; 1.18) | $6.1\times10^{-1}$ | 6 | IVW | 0.781 | 2,467 |
| trans-4-Hydroxyproline | Biogenic amines | AF | 0.98 (0.88; 1.09) | $7.3\times10^{-1}$ | 7 | IVW | 0.336 | 6,843 |
| trans-4-Hydroxyproline | Biogenic amines | NICM | 0.77 (0.48; 1.24) | $2.9\times10^{-1}$ | 7 | IVW | 0.48 | 5,512 |
| PC ae C40:2 | Phosphatidylcholines | DCM | 1.96 (1.15; 3.35) | $1.4\times10^{-2}$ | 7 | IVW | 0.184 | 8,813 |
| PC ae C40:2 | Phosphatidylcholines | HF | 0.95 (0.87; 1.04) | $2.7\times10^{-1}$ | 7 | IVW | 0.779 | 3,236 |
| PC ae C40:2 | Phosphatidylcholines | AF | 0.97 (0.89; 1.06) | $4.6\times10^{-1}$ | 7 | IVW | 0.302 | 7,206 |
| PC ae C40:2 | Phosphatidylcholines | NICM | 0.91 (0.63; 1.32) | $6.1\times10^{-1}$ | 7 | IVW | 0.646 | 4,228 |
| PC aa C40:5 | Phosphatidylcholines | DCM | 0.96 (0.81; 1.13) | $6.0\times10^{-1}$ | 34 | IVW | 0.405 | 34,297 |
| PC aa C40:5 | Phosphatidylcholines | HF | 1.06 (1.02; 1.10) | $2.1\times10^{-3}$ | 35 | IVW | 0.152 | 42,438 |
| PC aa C40:5 | Phosphatidylcholines | AF | 1.03 (1.01; 1.06) | $1.4\times10^{-2}$ | 35 | IVW | 0.025 | 51,999 |
| PC aa C40:5 | Phosphatidylcholines | NICM | 0.97 (0.85; 1.11) | $6.6\times10^{-1}$ | 34 | IVW | 0.482 | 32,694 |
| PC aa C42:1 | Phosphatidylcholines | DCM | 1.20 (0.89; 1.63) | $2.3\times10^{-1}$ | 14 | IVW | 0.397 | 13,677 |
| PC aa C42:1 | Phosphatidylcholines | HF | 1.15 (1.07; 1.23) | $4.7\times10^{-5}$ | 15 | IVW | 0.120 | 20,326 |
| PC aa C42:1 | Phosphatidylcholines | AF | 0.91 (0.77; 1.09) | $3.1\times10^{-1}$ | 15 | MR Egger | 0.042 | 22,963 |
| PC aa C42:1 | Phosphatidylcholines | NICM | 1.15 (0.92; 1.44) | $2.1\times10^{-1}$ | 16 | IVW | 0.900 | 8,543 |
| SM C20:2 | Sphingomyelins | DCM | 1.11 (0.71; 1.75) | $6.4\times10^{-1}$ | 11 | IVW | 0.123 | 15,259 |
| SM C20:2 | Sphingomyelins | HF | 0.91 (0.84; 0.97) | $7.1\times10^{-3}$ | 11 | IVW | 0.933 | 4,295 |
| SM C20:2 | Sphingomyelins | AF | 0.90 (0.85; 0.96) | $1.2\times10^{-3}$ | 11 | IVW | 0.035 | 19,479 |
| SM C20:2 | Sphingomyelins | NICM | 1.06 (0.78; 1.44) | $7.3\times10^{-1}$ | 10 | IVW | 0.536 | 7,978 |
| HydroxySM C16:1 | Sphingomyelins | DCM | 0.30 (0.09; 1.04) | $5.8\times10^{-2}$ | 12 | MR Egger | 0.066 | 17,406 |
| HydroxySM C16:1 | Sphingomyelins | HF | 0.99 (0.92; 1.07) | $8.3\times10^{-1}$ | 11 | IVW | 0.846 | 5,618 |
| HydroxySM C16:1 | Sphingomyelins | AF | 0.97 (0.89; 1.06) | $5.3\times10^{-1}$ | 10 | IVW | 0.126 | 13,909 |
| HydroxySM C16:1 | Sphingomyelins | NICM | 0.96 (0.71; 1.30) | $7.8\times10^{-1}$ | 12 | IVW | 0.850 | 6,336 |
| PC aa C36:5 | Phosphatidylcholines | DCM | 1.10 (0.90; 1.34) | $3.3\times10^{-1}$ | 24 | IVW | 0.107 | 31,661 |
| PC aa C36:5 | Phosphatidylcholines | HF | 1.05 (1.02; 1.09) | $3.6\times10^{-3}$ | 24 | IVW | 0.301 | 25,999 |
| PC aa C36:5 | Phosphatidylcholines | AF | 1.05 (1.01; 1.09) | $5.8\times10^{-3}$ | 24 | IVW | 0.065 | 34,005 |
| PC aa C36:5 | Phosphatidylcholines | NICM | 0.90 (0.76; 1.07) | $2.3\times10^{-1}$ | 24 | IVW | 0.060 | 34,357 |
| PC ae C38:6 | Phosphatidylcholines | DCM | 1.45 (0.74; 2.87) | $2.8\times10^{-1}$ | 22 | MR Egger | 0.651 | 17,032 |
| PC ae C38:6 | Phosphatidylcholines | HF | 1.04 (1.00; 1.08) | $7.3\times10^{-2}$ | 21 | IVW | 0.530 | 18,875 |
| PC ae C38:6 | Phosphatidylcholines | AF | 1.06 (1.03; 1.10) | $5.1\times10^{-4}$ | 20 | IVW | 0.007 | 37,463 |
| PC ae C38:6 | Phosphatidylcholines | NICM | 1.36 (0.63; 2.94) | $4.3\times10^{-1}$ | 23 | MR Egger | 0.764 | 16,109 |

| Table S2. Full MR results testing for the effect of metabolites on cardiac outcomes | | | | | | | | |
|---|---|---|---|---|---|---|---|---|
| **Metabolite*** | **Metabolite class*** | **Cardiac outcome** | **OR (95% CI)** | **p-value** | **No. variants*** | **Model*** | **Heterogeneity p-value** | **Q-statistic** |
| PC ae C32:1 | Phosphatidylcholines | DCM | 0.97 (0.80; 1.16) | $7.2\times10^{-1}$ | 26 | IVW | 0.589 | 22,803 |
| PC ae C32:1 | Phosphatidylcholines | HF | 0.95 (0.82; 1.11) | $5.5\times10^{-1}$ | 26 | MR Egger | 0.120 | 32,269 |
| PC ae C32:1 | Phosphatidylcholines | AF | 0.99 (0.96; 1.02) | $5.4\times10^{-1}$ | 27 | IVW | 0.760 | 20,654 |
| PC ae C32:1 | Phosphatidylcholines | NICM | 1.09 (0.93; 1.27) | $3.1\times10^{-1}$ | 27 | IVW | 0.388 | 27,407 |
| Decenoylcarnitine | Acylcarnitines | DCM | 1.17 (1.00; 1.36) | $5.5\times10^{-2}$ | 30 | IVW | 0.791 | 22,69 |
| Decenoylcarnitine | Acylcarnitines | HF | 0.98 (0.93; 1.02) | $3.2\times10^{-1}$ | 26 | IVW | 0.086 | 35,103 |
| Decenoylcarnitine | Acylcarnitines | AF | 1.00 (0.97; 1.03) | $7.7\times10^{-1}$ | 30 | IVW | 0.008 | 50,362 |
| Decenoylcarnitine | Acylcarnitines | NICM | 0.94 (0.81; 1.09) | $4.0\times10^{-1}$ | 27 | IVW | 0.728 | 21,269 |
| Valine | Amino acids | DCM | 1.30 (0.55; 3.10) | $5.5\times10^{-1}$ | 68 | MR Egger | 0.002 | 103,062 |
| Valine | Amino acids | HF | 0.94 (0.89; 1.00) | $3.9\times10^{-2}$ | 71 | IVW | 0.008 | 101,637 |
| Valine | Amino acids | AF | 1.05 (1.00; 1.11) | $3.5\times10^{-2}$ | 69 | IVW | <0.001 | 128,414 |
| Valine | Amino acids | NICM | 0.87 (0.70; 1.09) | $2.2\times10^{-1}$ | 73 | IVW | 0.022 | 98,08 |
| Hexose | Hexoses | DCM | 0.98 (0.66; 1.46) | $9.3\times10^{-1}$ | 11 | IVW | 0.988 | 2,677 |
| Hexose | Hexoses | HF | 1.07 (0.98; 1.17) | $1.2\times10^{-1}$ | 11 | IVW | 0.806 | 6,111 |
| Hexose | Hexoses | AF | 1.03 (0.96; 1.11) | $4.2\times10^{-1}$ | 11 | IVW | 0.302 | 11,75 |
| Hexose | Hexoses | NICM | 0.84 (0.57; 1.25) | $3.9\times10^{-1}$ | 9 | IVW | 0.863 | 3,93 |
| HydroxySM C22:1 | Sphingomyelins | DCM | 0.78 (0.52; 1.17) | $2.3\times10^{-1}$ | 8 | IVW | 0.89 | 2,942 |
| HydroxySM C22:1 | Sphingomyelins | HF | 1.13 (1.03; 1.25) | $1.1\times10^{-2}$ | 7 | IVW | 0.039 | 13,265 |
| HydroxySM C22:1 | Sphingomyelins | AF | 1.01 (0.93; 1.09) | $8.7\times10^{-1}$ | 8 | IVW | 0.964 | 1,92 |
| HydroxySM C22:1 | Sphingomyelins | NICM | 0.90 (0.61; 1.34) | $6.1\times10^{-1}$ | 7 | IVW | 0.971 | 1,315 |
| PC ae C34:2 | Phosphatidylcholines | DCM | 0.92 (0.71; 1.18) | $5.0\times10^{-1}$ | 21 | IVW | 0.144 | 26,691 |
| PC ae C34:2 | Phosphatidylcholines | HF | 0.95 (0.91; 0.99) | $2.6\times10^{-2}$ | 23 | IVW | 0.011 | 39,989 |
| PC ae C34:2 | Phosphatidylcholines | AF | 0.93 (0.89; 0.96) | $1.1\times10^{-4}$ | 22 | IVW | <0.001 | 51,881 |
| PC ae C34:2 | Phosphatidylcholines | NICM | 1.08 (0.90; 1.29) | $4.2\times10^{-1}$ | 23 | IVW | 0.513 | 21,124 |
| PC ae C40:5 | Phosphatidylcholines | DCM | 1.05 (0.94; 1.17) | $3.9\times10^{-1}$ | 53 | IVW | 0.093 | 65,946 |
| PC ae C40:5 | Phosphatidylcholines | HF | 1.07 (1.05; 1.09) | $2.7\times10^{-11}$ | 52 | IVW | 0.302 | 55,717 |
| PC ae C40:5 | Phosphatidylcholines | AF | 1.03 (1.01; 1.05) | $3.0\times10^{-4}$ | 53 | IVW | 0.127 | 63,757 |
| PC ae C40:5 | Phosphatidylcholines | NICM | 0.99 (0.90; 1.09) | $8.8\times10^{-1}$ | 53 | IVW | 0.074 | 67,415 |
| Octadecanoylcarnitine | Acylcarnitines | DCM | 1.47 (1.25; 1.73) | $2.3\times10^{-6}$ | 48 | IVW | 0.027 | 67,395 |
| Octadecanoylcarnitine | Acylcarnitines | HF | 0.81 (0.71; 0.92) | $1.0\times10^{-3}$ | 45 | MR Egger | 0.457 | 43,325 |
| Octadecanoylcarnitine | Acylcarnitines | AF | 0.84 (0.74; 0.97) | $1.5\times10^{-2}$ | 45 | MR Egger | 0.060 | 58,295 |

| Table S2. Full MR results testing for the effect of metabolites on cardiac outcomes | | | | | | | | |
|---|---|---|---|---|---|---|---|---|
| **Metabolite*** | **Metabolite class*** | **Cardiac outcome** | **OR (95% CI)** | **p-value** | **No. variants*** | **Model*** | **Heterogeneity p-value** | **Q-statistic** |
| Octadecanoylcarnitine | Acylcarnitines | NICM | 1.09 (0.94; 1.26) | $2.8 \times 10^{-1}$ | 42 | IVW | <0.001 | 76,092 |
| Kynurenine | Biogenic amines | DCM | 1.80 (1.14; 2.82) | $1.1 \times 10^{-2}$ | 57 | MR Egger | <0.001 | 99,216 |
| Kynurenine | Biogenic amines | HF | 1.01 (0.92; 1.10) | $8.9 \times 10^{-1}$ | 54 | MR Egger | <0.001 | 117,975 |
| Kynurenine | Biogenic amines | AF | 0.98 (0.95; 1.00) | $1.0 \times 10^{-1}$ | 58 | IVW | 0.031 | 78,492 |
| Kynurenine | Biogenic amines | NICM | 0.99 (0.86; 1.14) | $8.5 \times 10^{-1}$ | 54 | IVW | 0.050 | 70,966 |
| Isoleucine | Amino acids | DCM | 1.14 (0.68; 1.91) | $6.3 \times 10^{-1}$ | 31 | IVW | 0.378 | 31,779 |
| Isoleucine | Amino acids | HF | 0.96 (0.87; 1.07) | $4.5 \times 10^{-1}$ | 34 | IVW | 0.130 | 42,24 |
| Isoleucine | Amino acids | AF | 1.30 (1.02; 1.67) | $3.7 \times 10^{-2}$ | 34 | MR Egger | 0.035 | 47,95 |
| Isoleucine | Amino acids | NICM | 1.75 (1.08; 2.84) | $2.2 \times 10^{-2}$ | 33 | IVW | 0.071 | 44,442 |
| Pimelylcarnitine | Acylcarnitines | DCM | 1.49 (1.07; 2.07) | $1.8 \times 10^{-2}$ | 13 | IVW | 0.152 | 16,933 |
| Pimelylcarnitine | Acylcarnitines | HF | 0.97 (0.88; 1.07) | $5.4 \times 10^{-1}$ | 9 | IVW | 0.304 | 9,476 |
| Pimelylcarnitine | Acylcarnitines | AF | 1.43 (0.90; 2.25) | $1.3 \times 10^{-1}$ | 9 | MR Egger | 0.038 | 14,871 |
| Pimelylcarnitine | Acylcarnitines | NICM | 0.78 (0.54; 1.12) | $1.8 \times 10^{-1}$ | 9 | IVW | 0.972 | 2,258 |
| Decadienoylcarnitine | Acylcarnitines | DCM | 1.40 (0.95; 2.06) | $8.6 \times 10^{-2}$ | 6 | IVW | 0.421 | 4,956 |
| Decadienoylcarnitine | Acylcarnitines | HF | 1.16 (1.07; 1.26) | $1.7 \times 10^{-4}$ | 6 | IVW | 0.008 | 15,502 |
| Decadienoylcarnitine | Acylcarnitines | NICM | 1.33 (0.89; 1.99) | $1.6 \times 10^{-1}$ | 6 | IVW | 0.185 | 7,51 |
| Decanoylcarnitine | Acylcarnitines | DCM | 1.01 (0.89; 1.15) | $9.0 \times 10^{-1}$ | 74 | IVW | 0.471 | 73,216 |
| Decanoylcarnitine | Acylcarnitines | HF | 0.93 (0.87; 0.99) | $2.5 \times 10^{-2}$ | 67 | MR Egger | 0.005 | 98,24 |
| Decanoylcarnitine | Acylcarnitines | AF | 1.02 (0.99; 1.04) | $2.4 \times 10^{-1}$ | 67 | IVW | 0.003 | 102,702 |
| Decanoylcarnitine | Acylcarnitines | NICM | 1.00 (0.88; 1.14) | $9.9 \times 10^{-1}$ | 70 | IVW | 0.062 | 87,918 |
| PC ae C42:4 | Phosphatidylcholines | DCM | 1.15 (0.89; 1.48) | $2.9 \times 10^{-1}$ | 21 | IVW | 0.488 | 19,531 |
| PC ae C42:4 | Phosphatidylcholines | HF | 1.09 (0.91; 1.32) | $3.5 \times 10^{-1}$ | 21 | MR Egger | 0.296 | 21,767 |
| PC ae C42:4 | Phosphatidylcholines | AF | 0.99 (0.94; 1.03) | $5.3 \times 10^{-1}$ | 21 | IVW | 0.437 | 20,33 |
| PC ae C42:4 | Phosphatidylcholines | NICM | 0.90 (0.73; 1.11) | $3.2 \times 10^{-1}$ | 21 | IVW | 0.722 | 15,918 |
| PC ae C42:5 | Phosphatidylcholines | DCM | 0.97 (0.72; 1.32) | $8.6 \times 10^{-1}$ | 13 | IVW | 0.781 | 8,05 |
| PC ae C42:5 | Phosphatidylcholines | HF | 1.07 (1.01; 1.12) | $2.0 \times 10^{-2}$ | 14 | IVW | 0.877 | 7,466 |
| PC ae C42:5 | Phosphatidylcholines | AF | 1.09 (1.04; 1.14) | $1.5 \times 10^{-4}$ | 13 | IVW | 0.021 | 23,886 |
| PC ae C42:5 | Phosphatidylcholines | NICM | 0.94 (0.75; 1.18) | $6.0 \times 10^{-1}$ | 14 | IVW | 0.747 | 9,342 |
| SM C24:0 | Sphingomyelins | DCM | 2.89 (1.34; 6.21) | $6.7 \times 10^{-3}$ | 8 | MR Egger | 0.025 | 14,404 |
| SM C24:0 | Sphingomyelins | HF | 0.90 (0.77; 1.05) | $1.9 \times 10^{-1}$ | 7 | MR Egger | 0.037 | 11,853 |
| SM C24:0 | Sphingomyelins | AF | 0.98 (0.90; 1.06) | $5.6 \times 10^{-1}$ | 8 | IVW | 0.23 | 9,322 |

| Table S2. Full MR results testing for the effect of metabolites on cardiac outcomes | | | | | | | | |
|---|---|---|---|---|---|---|---|---|
| **Metabolite*** | **Metabolite class*** | **Cardiac outcome** | **OR (95% CI)** | **p-value** | **No. variants*** | **Model*** | **Heterogeneity p-value** | **Q-statistic** |
| SM C24:0 | Sphingomyelins | NICM | 0.91 (0.66; 1.24) | $5.4\times10^{-1}$ | 8 | IVW | 0.553 | 5,886 |
| HydroxySM C14:1 | Sphingomyelins | DCM | 1.03 (0.86; 1.23) | $7.5\times10^{-1}$ | 26 | IVW | <0.001 | 56,629 |
| HydroxySM C14:1 | Sphingomyelins | HF | 1.04 (1.00; 1.08) | $4.2\times10^{-2}$ | 26 | IVW | 0.225 | 29,981 |
| HydroxySM C14:1 | Sphingomyelins | AF | 1.10 (1.03; 1.17) | $4.0\times10^{-3}$ | 27 | MR Egger | 0.006 | 45,968 |
| HydroxySM C14:1 | Sphingomyelins | NICM | 1.13 (0.97; 1.31) | $1.2\times10^{-1}$ | 26 | IVW | 0.578 | 22,991 |
| Creatinine | Biogenic amines | DCM | 0.75 (0.58; 0.97) | $2.9\times10^{-2}$ | 115 | IVW | <0.001 | 190,451 |
| Creatinine | Biogenic amines | HF | 1.04 (0.84; 1.29) | $7.0\times10^{-1}$ | 117 | MR Egger | <0.001 | 176,291 |
| Creatinine | Biogenic amines | AF | 0.73 (0.61; 0.87) | $3.3\times10^{-4}$ | 123 | MR Egger | <0.001 | 205,326 |
| Creatinine | Biogenic amines | NICM | 0.53 (0.21; 1.34) | $1.8\times10^{-1}$ | 124 | MR Egger | 0.074 | 145,239 |
| Histidine | Amino acids | DCM | 0.93 (0.44; 1.96) | $8.5\times10^{-1}$ | 78 | MR Egger | <0.001 | 122,839 |
| Histidine | Amino acids | HF | 1.07 (1.03; 1.12) | $1.6\times10^{-3}$ | 79 | IVW | <0.001 | 138,256 |
| Histidine | Amino acids | AF | 1.06 (1.02; 1.10) | $2.4\times10^{-3}$ | 85 | IVW | <0.001 | 146,421 |
| Histidine | Amino acids | NICM | 0.99 (0.81; 1.21) | $8.9\times10^{-1}$ | 82 | IVW | 0.098 | 97,81 |
| PC aa C34:4 | Phosphatidylcholines | DCM | 1.09 (0.89; 1.34) | $3.8\times10^{-1}$ | 27 | IVW | 0.163 | 32,976 |
| PC aa C34:4 | Phosphatidylcholines | HF | 1.25 (1.11; 1.40) | $1.5\times10^{-4}$ | 29 | MR Egger | 0.038 | 41,327 |
| PC aa C34:4 | Phosphatidylcholines | AF | 1.04 (1.01; 1.08) | $1.4\times10^{-2}$ | 28 | IVW | 0.208 | 32,667 |
| PC aa C34:4 | Phosphatidylcholines | NICM | 1.33 (0.83; 2.14) | $2.4\times10^{-1}$ | 29 | MR Egger | 0.045 | 40,576 |
| PC ae C38:0 | Phosphatidylcholines | DCM | 0.99 (0.71; 1.37) | $9.3\times10^{-1}$ | 12 | IVW | 0.714 | 7,991 |
| PC ae C38:0 | Phosphatidylcholines | HF | 1.13 (1.05; 1.21) | $7.2\times10^{-4}$ | 11 | IVW | 0.023 | 20,742 |
| PC ae C38:0 | Phosphatidylcholines | AF | 0.99 (0.93; 1.04) | $6.3\times10^{-1}$ | 13 | IVW | 0.440 | 12,072 |
| PC ae C38:0 | Phosphatidylcholines | NICM | 1.11 (0.85; 1.46) | $4.4\times10^{-1}$ | 13 | IVW | 0.974 | 4,454 |
| Octadecenoylcarnitine | Acylcarnitines | DCM | 1.09 (0.87; 1.38) | $4.6\times10^{-1}$ | 25 | IVW | 0.753 | 18,989 |
| Octadecenoylcarnitine | Acylcarnitines | HF | 1.00 (0.96; 1.06) | $8.8\times10^{-1}$ | 23 | IVW | 0.428 | 22,544 |
| Octadecenoylcarnitine | Acylcarnitines | AF | 1.00 (0.95; 1.05) | $8.8\times10^{-1}$ | 25 | IVW | 0.056 | 35,949 |
| Octadecenoylcarnitine | Acylcarnitines | NICM | 0.81 (0.62; 1.06) | $1.3\times10^{-1}$ | 22 | IVW | 0.087 | 30,252 |
| LPC a C18:2 | Lysophosphatidylcholines | DCM | 1.28 (0.83; 2.00) | $2.7\times10^{-1}$ | 9 | IVW | 0.142 | 12,217 |
| LPC a C18:2 | Lysophosphatidylcholines | HF | 0.99 (0.92; 1.06) | $7.8\times10^{-1}$ | 9 | IVW | 0.013 | 19,459 |
| LPC a C18:2 | Lysophosphatidylcholines | AF | 0.92 (0.87; 0.99) | $1.7\times10^{-2}$ | 9 | IVW | 0.319 | 9,284 |
| LPC a C18:2 | Lysophosphatidylcholines | NICM | 1.07 (0.73; 1.56) | $7.3\times10^{-1}$ | 9 | IVW | 0.132 | 12,452 |
| LPC a C16:1 | Lysophosphatidylcholines | DCM | 1.17 (0.88; 1.56) | $2.9\times10^{-1}$ | 17 | IVW | 0.239 | 19,6 |
| LPC a C16:1 | Lysophosphatidylcholines | HF | 0.93 (0.79; 1.09) | $3.8\times10^{-1}$ | 18 | MR Egger | 0.261 | 19,151 |

| Table S2. Full MR results testing for the effect of metabolites on cardiac outcomes | | | | | | | | |
|---|---|---|---|---|---|---|---|---|
| **Metabolite*** | **Metabolite class*** | **Cardiac outcome** | **OR (95% CI)** | **p-value** | **No. variants*** | **Model*** | **Heterogeneity p-value** | **Q-statistic** |
| LPC a C16:1 | Lysophosphatidylcholines | AF | 1.00 (0.88; 1.14) | $9.8\times10^{-1}$ | 18 | MR Egger | 0.804 | 11,095 |
| LPC a C16:1 | Lysophosphatidylcholines | NICM | 1.30 (0.70; 2.42) | $4.1\times10^{-1}$ | 18 | MR Egger | 0.345 | 17,652 |
| PC ae C40:6 | Phosphatidylcholines | DCM | 1.07 (0.84; 1.36) | $5.9\times10^{-1}$ | 18 | IVW | 0.903 | 10,018 |
| PC ae C40:6 | Phosphatidylcholines | HF | 1.07 (1.02; 1.13) | $1.0\times10^{-2}$ | 17 | IVW | 0.344 | 17,665 |
| PC ae C40:6 | Phosphatidylcholines | AF | 1.03 (0.98; 1.07) | $2.9\times10^{-1}$ | 18 | IVW | 0.204 | 21,517 |
| PC ae C40:6 | Phosphatidylcholines | NICM | 0.95 (0.80; 1.13) | $5.8\times10^{-1}$ | 17 | IVW | 0.884 | 9,664 |
| PC ae C34:3 | Phosphatidylcholines | DCM | 1.20 (0.83; 1.75) | $3.3\times10^{-1}$ | 13 | IVW | 0.266 | 14,567 |
| PC ae C34:3 | Phosphatidylcholines | HF | 0.70 (0.52; 0.94) | $1.7\times10^{-2}$ | 14 | MR Egger | 0.149 | 17,01 |
| PC ae C34:3 | Phosphatidylcholines | AF | 0.95 (0.88; 1.01) | $1.0\times10^{-1}$ | 13 | IVW | 0.161 | 16,717 |
| PC ae C34:3 | Phosphatidylcholines | NICM | 1.19 (0.89; 1.58) | $2.4\times10^{-1}$ | 13 | IVW | 0.915 | 6,021 |
| Glutamine | Amino acids | DCM | 0.97 (0.60; 1.57) | $9.1\times10^{-1}$ | 126 | MR Egger | <0.001 | 217,679 |
| Glutamine | Amino acids | HF | 1.06 (1.02; 1.10) | $4.8\times10^{-3}$ | 129 | IVW | <0.001 | 194,102 |
| Glutamine | Amino acids | AF | 1.00 (0.97; 1.04) | $9.8\times10^{-1}$ | 126 | IVW | <0.001 | 227,818 |
| Glutamine | Amino acids | NICM | 1.06 (0.90; 1.24) | $4.8\times10^{-1}$ | 129 | IVW | 0.016 | 164,472 |
| Dodecenoylcarnitine | Acylcarnitines | DCM | 1.42 (1.05; 1.91) | $2.1\times10^{-2}$ | 13 | IVW | 0.884 | 6,576 |
| Dodecenoylcarnitine | Acylcarnitines | HF | 0.96 (0.91; 1.01) | $9.9\times10^{-2}$ | 14 | IVW | 0.025 | 24,786 |
| Dodecenoylcarnitine | Acylcarnitines | AF | 0.93 (0.88; 0.98) | $9.9\times10^{-3}$ | 12 | IVW | 0.002 | 29,464 |
| Dodecenoylcarnitine | Acylcarnitines | NICM | 0.82 (0.66; 1.03) | $9.4\times10^{-2}$ | 13 | IVW | 0.316 | 13,769 |
| PC ae C36:5 | Phosphatidylcholines | DCM | 0.96 (0.86; 1.07) | $4.2\times10^{-1}$ | 48 | IVW | 0.303 | 51,481 |
| PC ae C36:5 | Phosphatidylcholines | HF | 1.05 (1.03; 1.07) | $9.5\times10^{-6}$ | 50 | IVW | 0.254 | 55,135 |
| PC ae C36:5 | Phosphatidylcholines | AF | 1.04 (1.02; 1.06) | $1.4\times10^{-6}$ | 50 | IVW | 0.003 | 80,946 |
| PC ae C36:5 | Phosphatidylcholines | NICM | 1.00 (0.92; 1.10) | $9.4\times10^{-1}$ | 48 | IVW | 0.177 | 55,826 |
| PC aa C36:3 | Phosphatidylcholines | DCM | 1.07 (0.87; 1.31) | $5.3\times10^{-1}$ | 31 | IVW | 0.059 | 42,937 |
| PC aa C36:3 | Phosphatidylcholines | HF | 0.90 (0.82; 0.98) | $1.9\times10^{-2}$ | 30 | MR Egger | 0.222 | 33,389 |
| PC aa C36:3 | Phosphatidylcholines | AF | 0.90 (0.82; 0.98) | $1.2\times10^{-2}$ | 31 | MR Egger | 0.073 | 40,737 |
| PC aa C36:3 | Phosphatidylcholines | NICM | 1.01 (0.87; 1.18) | $8.9\times10^{-1}$ | 31 | IVW | 0.942 | 18,908 |
| PC ae C42:2 | Phosphatidylcholines | DCM | 1.04 (0.91; 1.19) | $5.9\times10^{-1}$ | 36 | IVW | 0.536 | 33,602 |
| PC ae C42:2 | Phosphatidylcholines | HF | 1.09 (1.06; 1.12) | $2.9\times10^{-8}$ | 34 | IVW | 0.284 | 37,131 |
| PC ae C42:2 | Phosphatidylcholines | AF | 1.06 (1.03; 1.08) | $3.1\times10^{-6}$ | 33 | IVW | 0.045 | 46,725 |
| PC ae C42:2 | Phosphatidylcholines | NICM | 1.00 (0.89; 1.12) | $1.0\times10^{0}$ | 34 | IVW | 0.557 | 31,19 |
| PC aa C40:6 | Phosphatidylcholines | DCM | 2.55 (1.09; 5.97) | $3.1\times10^{-2}$ | 31 | MR Egger | 0.036 | 44,054 |

| Table S2. Full MR results testing for the effect of metabolites on cardiac outcomes | | | | | | | | |
|---|---|---|---|---|---|---|---|---|
| **Metabolite*** | **Metabolite class*** | **Cardiac outcome** | **OR (95% CI)** | **p-value** | **No. variants*** | **Model*** | **Heterogeneity p-value** | **Q-statistic** |
| PC aa C40:6 | Phosphatidylcholines | HF | 1.07 (1.02; 1.12) | $5.6\times10^{-3}$ | 27 | IVW | 0.009 | 45,916 |
| PC aa C40:6 | Phosphatidylcholines | AF | 1.02 (0.98; 1.06) | $3.9\times10^{-1}$ | 31 | IVW | 0.832 | 22,578 |
| PC aa C40:6 | Phosphatidylcholines | NICM | 0.95 (0.80; 1.15) | $6.2\times10^{-1}$ | 32 | IVW | 0.471 | 30,909 |
| Alanine | Amino acids | DCM | 1.62 (0.71; 3.73) | $2.6\times10^{-1}$ | 119 | MR Egger | 0.002 | 166,188 |
| Alanine | Amino acids | HF | 0.99 (0.94; 1.04) | $6.0\times10^{-1}$ | 112 | IVW | 0.004 | 155,242 |
| Alanine | Amino acids | AF | 1.00 (0.96; 1.05) | $9.0\times10^{-1}$ | 121 | IVW | <0.001 | 208,563 |
| Alanine | Amino acids | NICM | 1.01 (0.82; 1.25) | $9.0\times10^{-1}$ | 115 | IVW | 0.033 | 143,195 |
| Citrulline | Amino acids | DCM | 0.29 (0.13; 0.66) | $3.3\times10^{-3}$ | 55 | MR Egger | 0.231 | 60,223 |
| Citrulline | Amino acids | HF | 1.03 (0.90; 1.19) | $6.6\times10^{-1}$ | 50 | MR Egger | 0.002 | 80,394 |
| Citrulline | Amino acids | AF | 0.99 (0.95; 1.02) | $4.9\times10^{-1}$ | 54 | IVW | 0.046 | 71,52 |
| Citrulline | Amino acids | NICM | 0.84 (0.69; 1.02) | $7.6\times10^{-2}$ | 51 | IVW | 0.153 | 60,198 |
| Asparagine | Amino acids | DCM | 0.99 (0.86; 1.14) | $8.8\times10^{-1}$ | 68 | IVW | 0.015 | 94,584 |
| Asparagine | Amino acids | HF | 0.96 (0.93; 0.99) | $7.2\times10^{-3}$ | 60 | IVW | <0.001 | 118,699 |
| Asparagine | Amino acids | AF | 1.06 (1.04; 1.09) | $4.3\times10^{-7}$ | 68 | IVW | <0.001 | 117,31 |
| Asparagine | Amino acids | NICM | 0.89 (0.78; 1.01) | $6.9\times10^{-2}$ | 61 | IVW | 0.011 | 87,958 |
| PC aa C38:5 | Phosphatidylcholines | DCM | 1.10 (0.98; 1.23) | $9.9\times10^{-2}$ | 46 | IVW | 0.182 | 53,445 |
| PC aa C38:5 | Phosphatidylcholines | HF | 1.06 (1.03; 1.08) | $1.8\times10^{-6}$ | 46 | IVW | 0.058 | 60,821 |
| PC aa C38:5 | Phosphatidylcholines | AF | 1.03 (0.99; 1.08) | $1.2\times10^{-1}$ | 44 | MR Egger | 0.034 | 60,205 |
| PC aa C38:5 | Phosphatidylcholines | NICM | 0.91 (0.84; 0.99) | $3.0\times10^{-2}$ | 47 | IVW | 0.017 | 68,491 |
| PC aa C38:4 | Phosphatidylcholines | DCM | 1.05 (0.97; 1.14) | $2.0\times10^{-1}$ | 64 | IVW | 0.272 | 69,365 |
| PC aa C38:4 | Phosphatidylcholines | HF | 1.08 (1.06; 1.11) | $1.2\times10^{-9}$ | 68 | MR Egger | 0.042 | 87,156 |
| PC aa C38:4 | Phosphatidylcholines | AF | 1.05 (1.03; 1.06) | $2.5\times10^{-14}$ | 66 | IVW | 0.003 | 101,268 |
| PC aa C38:4 | Phosphatidylcholines | NICM | 0.98 (0.92; 1.04) | $4.8\times10^{-1}$ | 65 | IVW | 0.163 | 75,014 |
| PC ae C40:1 | Phosphatidylcholines | DCM | 0.99 (0.69; 1.41) | $9.5\times10^{-1}$ | 13 | IVW | 0.104 | 18,418 |
| PC ae C40:1 | Phosphatidylcholines | HF | 1.10 (1.04; 1.16) | $1.5\times10^{-3}$ | 13 | IVW | 0.202 | 15,76 |
| PC ae C40:1 | Phosphatidylcholines | AF | 1.04 (0.99; 1.09) | $1.4\times10^{-1}$ | 13 | IVW | 0.851 | 7,106 |
| PC ae C40:1 | Phosphatidylcholines | NICM | 1.02 (0.82; 1.28) | $8.4\times10^{-1}$ | 14 | IVW | 0.323 | 14,749 |
| LPC a C20:4 | Lysophosphatidylcholines | DCM | 1.01 (0.93; 1.10) | $8.2\times10^{-1}$ | 66 | IVW | 0.256 | 72,05 |
| LPC a C20:4 | Lysophosphatidylcholines | HF | 1.03 (1.02; 1.05) | $4.6\times10^{-5}$ | 69 | IVW | 0.027 | 92,174 |
| LPC a C20:4 | Lysophosphatidylcholines | AF | 1.03 (1.01; 1.04) | $4.5\times10^{-4}$ | 67 | IVW | 0.004 | 100,267 |
| LPC a C20:4 | Lysophosphatidylcholines | NICM | 1.20 (1.09; 1.32) | $2.7\times10^{-4}$ | 73 | MR Egger | 0.010 | 101,51 |

| Table S2. Full MR results testing for the effect of metabolites on cardiac outcomes | | | | | | | | |
|---|---|---|---|---|---|---|---|---|
| **Metabolite*** | **Metabolite class*** | **Cardiac outcome** | **OR (95% CI)** | **p-value** | **No. variants*** | **Model*** | **Heterogeneity p-value** | **Q-statistic** |
| Spermidine | Biogenic amines | DCM | 1.04 (0.80; 1.34) | $7.9\times10^{-1}$ | 23 | IVW | 0.271 | 25,571 |
| Spermidine | Biogenic amines | HF | 0.99 (0.95; 1.04) | $7.1\times10^{-1}$ | 23 | IVW | 0.730 | 17,584 |
| Spermidine | Biogenic amines | AF | 0.98 (0.93; 1.02) | $3.3\times10^{-1}$ | 26 | IVW | 0.215 | 30,26 |
| Spermidine | Biogenic amines | NICM | 0.89 (0.71; 1.10) | $2.8\times10^{-1}$ | 22 | IVW | 0.131 | 28,33 |
| PC aa C24:0 | Phosphatidylcholines | DCM | 1.17 (0.99; 1.40) | $7.0\times10^{-2}$ | 22 | IVW | 0.156 | 27,481 |
| PC aa C24:0 | Phosphatidylcholines | HF | 1.05 (1.02; 1.08) | $1.8\times10^{-3}$ | 21 | IVW | 0.787 | 14,817 |
| PC aa C24:0 | Phosphatidylcholines | AF | 1.04 (1.02; 1.07) | $7.5\times10^{-4}$ | 21 | IVW | 0.019 | 35,287 |
| PC aa C24:0 | Phosphatidylcholines | NICM | 1.24 (0.82; 1.88) | $3.0\times10^{-1}$ | 22 | MR Egger | 0.605 | 17,737 |
| PC aa C28:1 | Phosphatidylcholines | DCM | 0.96 (0.78; 1.18) | $7.0\times10^{-1}$ | 19 | IVW | <0.001 | 51,986 |
| PC aa C28:1 | Phosphatidylcholines | HF | 1.07 (1.03; 1.12) | $5.3\times10^{-4}$ | 24 | IVW | 0.136 | 30,491 |
| PC aa C28:1 | Phosphatidylcholines | AF | 1.05 (1.01; 1.09) | $2.0\times10^{-2}$ | 24 | IVW | 0.250 | 27,151 |
| PC aa C28:1 | Phosphatidylcholines | NICM | 1.01 (0.89; 1.16) | $8.2\times10^{-1}$ | 25 | IVW | 0.013 | 42,067 |
| Phenylalanine | Amino acids | DCM | 1.09 (0.81; 1.45) | $5.8\times10^{-1}$ | 69 | IVW | 0.003 | 104,751 |
| Phenylalanine | Amino acids | HF | 1.00 (0.94; 1.08) | $9.1\times10^{-1}$ | 63 | IVW | 0.105 | 76,279 |
| Phenylalanine | Amino acids | AF | 1.03 (0.98; 1.09) | $2.7\times10^{-1}$ | 69 | IVW | 0.002 | 106,923 |
| Phenylalanine | Amino acids | NICM | 0.86 (0.66; 1.13) | $2.9\times10^{-1}$ | 67 | IVW | 0.317 | 70,93 |
| Valerylcarnitine | Acylcarnitines | DCM | 1.05 (0.86; 1.28) | $6.3\times10^{-1}$ | 24 | IVW | 0.253 | 27,072 |
| Valerylcarnitine | Acylcarnitines | HF | 0.91 (0.76; 1.08) | $2.8\times10^{-1}$ | 21 | MR Egger | 0.101 | 27,171 |
| Valerylcarnitine | Acylcarnitines | AF | 1.03 (1.00; 1.06) | $8.2\times10^{-2}$ | 24 | IVW | 0.008 | 42,588 |
| Valerylcarnitine | Acylcarnitines | NICM | 0.82 (0.70; 0.96) | $1.5\times10^{-2}$ | 21 | IVW | 0.009 | 37,8 |
| Methionine | Amino acids | DCM | 0.52 (0.35; 0.77) | $1.3\times10^{-3}$ | 16 | IVW | 0.835 | 9,753 |
| Methionine | Amino acids | HF | 0.92 (0.84; 1.01) | $7.3\times10^{-2}$ | 14 | IVW | 0.005 | 30,02 |
| Methionine | Amino acids | AF | 0.97 (0.90; 1.04) | $3.5\times10^{-1}$ | 16 | IVW | 0.009 | 30,993 |
| Methionine | Amino acids | NICM | 0.36 (0.11; 1.20) | $9.7\times10^{-2}$ | 16 | MR Egger | 0.324 | 15,83 |
| PC aa C36:2 | Phosphatidylcholines | DCM | 1.22 (1.01; 1.47) | $3.6\times10^{-2}$ | 29 | IVW | 0.004 | 52,223 |
| PC aa C36:2 | Phosphatidylcholines | HF | 0.94 (0.79; 1.11) | $4.4\times10^{-1}$ | 27 | MR Egger | <0.001 | 51,371 |
| PC aa C36:2 | Phosphatidylcholines | AF | 1.01 (0.87; 1.17) | $8.9\times10^{-1}$ | 30 | MR Egger | 0.085 | 38,731 |
| PC aa C36:2 | Phosphatidylcholines | NICM | 1.05 (0.90; 1.24) | $5.3\times10^{-1}$ | 29 | IVW | 0.413 | 28,982 |
| Glycine | Amino acids | DCM | 1.02 (0.93; 1.12) | $6.9\times10^{-1}$ | 266 | IVW | <0.001 | 398,052 |
| Glycine | Amino acids | HF | 0.95 (0.92; 0.98) | $6.2\times10^{-4}$ | 248 | MR Egger | <0.001 | 417,885 |
| Glycine | Amino acids | AF | 0.98 (0.96; 1.00) | $1.1\times10^{-1}$ | 277 | MR Egger | <0.001 | 392,232 |

| Table S2. Full MR results testing for the effect of metabolites on cardiac outcomes | | | | | | | | |
|---|---|---|---|---|---|---|---|---|
| **Metabolite*** | **Metabolite class*** | **Cardiac outcome** | **OR (95% CI)** | **p-value** | **No. variants*** | **Model*** | **Heterogeneity p-value** | **Q-statistic** |
| Glycine | Amino acids | NICM | 1.12 (1.05; 1.19) | $7.0\times10^{-4}$ | 269 | IVW | <0.001 | 427,559 |
| PC aa C42:5 | Phosphatidylcholines | DCM | 0.84 (0.56; 1.27) | $4.2\times10^{-1}$ | 6 | IVW | 0.463 | 4,624 |
| PC aa C42:5 | Phosphatidylcholines | HF | 0.99 (0.92; 1.07) | $8.6\times10^{-1}$ | 7 | IVW | <0.001 | 27,193 |
| PC aa C42:5 | Phosphatidylcholines | AF | 0.86 (0.71; 1.05) | $1.4\times10^{-1}$ | 7 | MR Egger | 0.980 | 0,757 |
| PC aa C42:5 | Phosphatidylcholines | NICM | 1.15 (0.83; 1.60) | $4.0\times10^{-1}$ | 7 | IVW | 0.714 | 3,724 |
| PC ae C40:4 | Phosphatidylcholines | DCM | 1.02 (0.79; 1.33) | $8.6\times10^{-1}$ | 18 | IVW | 0.116 | 24,12 |
| PC ae C40:4 | Phosphatidylcholines | HF | 1.00 (0.96; 1.05) | $9.2\times10^{-1}$ | 18 | IVW | 0.653 | 14,195 |
| PC ae C40:4 | Phosphatidylcholines | AF | 0.93 (0.88; 0.97) | $2.2\times10^{-3}$ | 17 | IVW | 0.097 | 23,681 |
| PC ae C40:4 | Phosphatidylcholines | NICM | 1.02 (0.85; 1.22) | $8.3\times10^{-1}$ | 18 | IVW | 0.424 | 17,447 |
| Tyrosine | Amino acids | DCM | 0.86 (0.71; 1.04) | $1.2\times10^{-1}$ | 116 | IVW | 0.015 | 150,464 |
| Tyrosine | Amino acids | HF | 1.09 (1.05; 1.13) | $2.3\times10^{-6}$ | 119 | IVW | 0.014 | 154,34 |
| Tyrosine | Amino acids | AF | 1.04 (1.01; 1.08) | $1.8\times10^{-2}$ | 115 | IVW | <0.001 | 215,243 |
| Tyrosine | Amino acids | NICM | 0.66 (0.45; 0.96) | $3.1\times10^{-2}$ | 120 | MR Egger | 0.098 | 138,242 |
| PC ae C42:1 | Phosphatidylcholines | DCM | 1.07 (0.84; 1.36) | $6.1\times10^{-1}$ | 24 | IVW | 0.904 | 14,727 |
| PC ae C42:1 | Phosphatidylcholines | HF | 0.92 (0.83; 1.03) | $1.5\times10^{-1}$ | 27 | MR Egger | 0.557 | 23,345 |
| PC ae C42:1 | Phosphatidylcholines | AF | 0.96 (0.93; 0.99) | $9.0\times10^{-3}$ | 30 | IVW | 0.015 | 47,961 |
| PC ae C42:1 | Phosphatidylcholines | NICM | 1.04 (0.87; 1.23) | $6.8\times10^{-1}$ | 27 | IVW | 0.629 | 23,061 |
| Sarcosine | Biogenic amines | DCM | 1.28 (0.97; 1.70) | $8.5\times10^{-2}$ | 24 | IVW | 0.823 | 16,721 |
| Sarcosine | Biogenic amines | HF | 1.10 (1.03; 1.18) | $6.6\times10^{-3}$ | 23 | IVW | 0.160 | 28,501 |
| Sarcosine | Biogenic amines | AF | 1.02 (0.96; 1.07) | $6.0\times10^{-1}$ | 24 | IVW | 0.499 | 22,359 |
| Sarcosine | Biogenic amines | NICM | 0.75 (0.60; 0.94) | $1.4\times10^{-2}$ | 23 | IVW | 0.532 | 20,816 |
| Taurine | Biogenic amines | DCM | 0.95 (0.65; 1.38) | $8.0\times10^{-1}$ | 15 | IVW | 0.998 | 3,5 |
| Taurine | Biogenic amines | HF | 0.95 (0.90; 1.01) | $9.2\times10^{-2}$ | 19 | IVW | 0.130 | 24,828 |
| Taurine | Biogenic amines | AF | 0.99 (0.94; 1.04) | $6.9\times10^{-1}$ | 22 | IVW | 0.016 | 37,241 |
| Taurine | Biogenic amines | NICM | 0.48 (0.38; 0.60) | $4.7\times10^{-10}$ | 19 | IVW | 0.441 | 18,221 |
| Leucine | Amino acids | DCM | 0.89 (0.53; 1.50) | $6.6\times10^{-1}$ | 38 | IVW | 0.111 | 47,726 |
| Leucine | Amino acids | HF | 1.00 (0.91; 1.09) | $9.9\times10^{-1}$ | 38 | IVW | <0.001 | 75,897 |
| Leucine | Amino acids | AF | 1.10 (1.02; 1.19) | $1.7\times10^{-2}$ | 39 | IVW | 0.042 | 54,283 |
| Leucine | Amino acids | NICM | 1.51 (1.02; 2.22) | $3.8\times10^{-2}$ | 37 | IVW | 0.004 | 62,554 |
| PC ae C36:2 | Phosphatidylcholines | DCM | 1.21 (0.95; 1.55) | $1.3\times10^{-1}$ | 20 | IVW | 0.076 | 28,425 |
| PC ae C36:2 | Phosphatidylcholines | HF | 0.94 (0.90; 0.98) | $4.7\times10^{-3}$ | 20 | IVW | <0.001 | 49,239 |

| Table S2. Full MR results testing for the effect of metabolites on cardiac outcomes | | | | | | | | |
|---|---|---|---|---|---|---|---|---|
| **Metabolite*** | **Metabolite class*** | **Cardiac outcome** | **OR (95% CI)** | **p-value** | **No. variants*** | **Model*** | **Heterogeneity p-value** | **Q-statistic** |
| PC ae C36:2 | Phosphatidylcholines | AF | 0.95 (0.91; 0.98) | $5.7×10^{-3}$ | 21 | IVW | 0.128 | 27,276 |
| PC ae C36:2 | Phosphatidylcholines | NICM | 1.11 (0.94; 1.31) | $2.4×10^{-1}$ | 21 | IVW | 0.627 | 17,391 |
| SM C16:1 | Sphingomyelins | DCM | 1.36 (1.04; 1.79) | $2.7×10^{-2}$ | 17 | IVW | 0.043 | 26,88 |
| SM C16:1 | Sphingomyelins | HF | 1.03 (0.98; 1.08) | $2.7×10^{-1}$ | 16 | IVW | 0.024 | 27,691 |
| SM C16:1 | Sphingomyelins | AF | 1.12 (1.08; 1.17) | $3.2×10^{-8}$ | 17 | IVW | 0.002 | 36,816 |
| SM C16:1 | Sphingomyelins | NICM | 1.18 (0.96; 1.45) | $1.1×10^{-1}$ | 18 | IVW | 0.801 | 11,979 |
| PC ae C38:5 | Phosphatidylcholines | DCM | 0.98 (0.84; 1.15) | $8.2×10^{-1}$ | 41 | IVW | 0.073 | 53,68 |
| PC ae C38:5 | Phosphatidylcholines | HF | 1.05 (1.03; 1.08) | $5.7×10^{-5}$ | 42 | IVW | 0.019 | 61,896 |
| PC ae C38:5 | Phosphatidylcholines | AF | 1.04 (1.02; 1.07) | $9.6×10^{-4}$ | 41 | IVW | 0.202 | 47,204 |
| PC ae C38:5 | Phosphatidylcholines | NICM | 1.06 (0.95; 1.18) | $3.0×10^{-1}$ | 42 | IVW | 0.548 | 39,256 |
| PC aa C34:3 | Phosphatidylcholines | DCM | 0.86 (0.65; 1.12) | $2.5×10^{-1}$ | 19 | IVW | 0.586 | 16,093 |
| PC aa C34:3 | Phosphatidylcholines | HF | 0.98 (0.93; 1.03) | $4.7×10^{-1}$ | 19 | IVW | <0.001 | 50,074 |
| PC aa C34:3 | Phosphatidylcholines | AF | 0.96 (0.91; 1.01) | $1.3×10^{-1}$ | 18 | IVW | 0.330 | 18,978 |
| PC aa C34:3 | Phosphatidylcholines | NICM | 0.90 (0.72; 1.13) | $3.7×10^{-1}$ | 19 | IVW | 0.992 | 6,759 |
| Tetradecadienoylcarnitine | Acylcarnitines | DCM | 1.22 (0.81; 1.86) | $3.4×10^{-1}$ | 8 | IVW | 0.641 | 5,159 |
| Tetradecadienoylcarnitine | Acylcarnitines | HF | 1.07 (0.97; 1.17) | $1.9×10^{-1}$ | 8 | IVW | 0.292 | 8,482 |
| Tetradecadienoylcarnitine | Acylcarnitines | AF | 0.91 (0.83; 0.99) | $3.1×10^{-2}$ | 7 | IVW | 0.017 | 15,388 |
| Tetradecadienoylcarnitine | Acylcarnitines | NICM | 1.16 (0.72; 1.89) | $5.4×10^{-1}$ | 8 | IVW | 0.065 | 13,294 |
| LPC a C16:0 | Lysophosphatidylcholines | DCM | 1.71 (1.18; 2.48) | $4.9×10^{-3}$ | 16 | IVW | 0.120 | 21,568 |
| LPC a C16:0 | Lysophosphatidylcholines | HF | 1.09 (1.02; 1.16) | $1.4×10^{-2}$ | 17 | IVW | 0.365 | 17,327 |
| LPC a C16:0 | Lysophosphatidylcholines | AF | 0.97 (0.92; 1.03) | $3.7×10^{-1}$ | 17 | IVW | 0.199 | 20,486 |
| LPC a C16:0 | Lysophosphatidylcholines | NICM | 0.76 (0.53; 1.07) | $1.1×10^{-1}$ | 16 | IVW | 0.090 | 22,725 |
| PC aa C30:0 | Phosphatidylcholines | DCM | 0.97 (0.60; 1.56) | $8.9×10^{-1}$ | 8 | IVW | 0.742 | 4,32 |
| PC aa C30:0 | Phosphatidylcholines | HF | 1.10 (0.98; 1.23) | $1.1×10^{-1}$ | 7 | IVW | 0.298 | 7,26 |
| PC aa C30:0 | Phosphatidylcholines | AF | 1.03 (0.95; 1.12) | $4.7×10^{-1}$ | 8 | IVW | 0.009 | 18,637 |
| PC aa C30:0 | Phosphatidylcholines | NICM | 1.18 (0.78; 1.77) | $4.3×10^{-1}$ | 8 | IVW | 0.803 | 3,798 |
| PC aa C36:1 | Phosphatidylcholines | DCM | 0.96 (0.61; 1.52) | $8.8×10^{-1}$ | 8 | IVW | 0.773 | 4,061 |
| PC aa C36:1 | Phosphatidylcholines | HF | 0.96 (0.85; 1.08) | $5.2×10^{-1}$ | 8 | IVW | 0.111 | 11,701 |
| PC aa C36:1 | Phosphatidylcholines | AF | 1.03 (0.95; 1.11) | $5.2×10^{-1}$ | 8 | IVW | 0.011 | 18,249 |
| PC aa C36:1 | Phosphatidylcholines | NICM | 1.02 (0.69; 1.51) | $9.1×10^{-1}$ | 8 | IVW | 0.811 | 3,724 |
| Octanoylcarnitine | Acylcarnitines | DCM | 0.98 (0.84; 1.14) | $8.0×10^{-1}$ | 95 | MR Egger | 0.560 | 90,297 |

| Table S2. Full MR results testing for the effect of metabolites on cardiac outcomes | | | | | | | | |
|---|---|---|---|---|---|---|---|---|
| **Metabolite*** | **Metabolite class*** | **Cardiac outcome** | **OR (95% CI)** | **p-value** | **No. variants*** | **Model*** | **Heterogeneity p-value** | **Q-statistic** |
| Octanoylcarnitine | Acylcarnitines | HF | 0.97 (0.94; 1.01) | $1.0\times10^{-1}$ | 88 | MR Egger | <0.001 | 141,624 |
| Octanoylcarnitine | Acylcarnitines | AF | 0.98 (0.95; 1.01) | $1.2\times10^{-1}$ | 94 | MR Egger | <0.001 | 151,554 |
| Octanoylcarnitine | Acylcarnitines | NICM | 0.89 (0.81; 0.97) | $7.9\times10^{-3}$ | 91 | IVW | <0.001 | 137,82 |
| PC ae C34:0 | Phosphatidylcholines | HF | 1.26 (0.92; 1.72) | $1.5\times10^{-1}$ | 6 | MR Egger | 0.204 | 5,936 |
| PC ae C34:0 | Phosphatidylcholines | AF | 1.08 (0.99; 1.18) | $9.2\times10^{-2}$ | 6 | IVW | 0.008 | 15,773 |
| PC ae C34:0 | Phosphatidylcholines | NICM | 1.31 (0.86; 1.99) | $2.1\times10^{-1}$ | 6 | IVW | 0.758 | 2,623 |
| PC aa C36:4 | Phosphatidylcholines | DCM | 1.09 (1.01; 1.18) | $3.7\times10^{-2}$ | 59 | IVW | 0.018 | 82,809 |
| PC aa C36:4 | Phosphatidylcholines | HF | 1.07 (1.04; 1.10) | $1.8\times10^{-5}$ | 61 | MR Egger | 0.089 | 74,099 |
| PC aa C36:4 | Phosphatidylcholines | AF | 1.04 (1.01; 1.07) | $2.5\times10^{-3}$ | 61 | MR Egger | 0.088 | 74,168 |
| PC aa C36:4 | Phosphatidylcholines | NICM | 1.06 (0.98; 1.16) | $1.7\times10^{-1}$ | 60 | IVW | 0.059 | 76,861 |
| Methylglutarylcarnitine | Acylcarnitines | DCM | 0.92 (0.55; 1.55) | $7.6\times10^{-1}$ | 58 | MR Egger | 0.160 | 66,433 |
| Methylglutarylcarnitine | Acylcarnitines | HF | 1.01 (0.98; 1.03) | $5.8\times10^{-1}$ | 62 | IVW | 0.012 | 88,611 |
| Methylglutarylcarnitine | Acylcarnitines | AF | 1.02 (0.94; 1.10) | $7.0\times10^{-1}$ | 62 | MR Egger | <0.001 | 128,924 |
| Methylglutarylcarnitine | Acylcarnitines | NICM | 1.13 (1.02; 1.26) | $2.5\times10^{-2}$ | 62 | IVW | 0.227 | 68,947 |

* Columns: Metabolite - metabolite name, full names are listed in Table S1, Metabolite class - class metabolite belongs to, No. variants - number of variants used in the MR, Model - model selected by the model selection framework (as described in the Methods section)

Abbreviations: a = acyl residue, aa = diacyl residue, ae = acyl-alkyl residue, AF = atrial fibrillation, DCM = dilated cardiomyopathy, HF = heart failure, IVW = inverse-variance weighted, LPC = lysophosphatidylcholine, NICM = non-ischemic cardiomyopathy, OR = odds ratio, PC = phosphatidylcholine, SM = sphingomyelin
