## Supplemental Table S3 for "Integrating metabolomics and proteomics to identify novel drug targets for heart failure and atrial fibrillation"

| Table S3. Number of metabolites per class associated with cardiac outcomes | | | | |
|---|---|---|---|---|
| | **AF** | **HF** | **DCM** | **NICM** |
| Amino acids | 2 | 3 | 1 | 0 |
| Biogenic amines | 1 | 0 | 0 | 1 |
| Acylcarnitines | 2 | 1 | 4 | 0 |
| Lysophosphatidylcholines | 3 | 1 | 0 | 0 |
| Phosphatidylcholines | 7 | 13 | 0 | 0 |
| Sphingomyelins | 2 | 1 | 0 | 0 |
| Hexoses | 0 | 0 | 0 | 0 |

Abbreviations: AF = atrial fibrillation, DCM = dilated cardiomyopathy,
HF = heart failure, NICM = non-ischemic cardiomyopathy
