## Supplemental Table S4 for "Integrating metabolomics and proteomics to identify novel drug targets for heart failure and atrial fibrillation"

**Table S4. Enrichment of significant metabolite classes per cardiac outcome**

| Indicated metabolite class | Number of metabolites of other classes associated with other outcomes | Number of metabolites of indicated class associated with other outcomes | Number of metabolites of other classes associated with indicated outcome | Number of metabolites of indicated class associated with indicated outcome | OR | p-value | Indicated cardiac outcome |
|---|---|---|---|---|---|---|---|
| Amino acids | 21 | 4 | 15 | 2 | 0,7 | 1 | AF |
| Biogenic amines | 24 | 1 | 16 | 1 | 1,5 | 1 | AF |
| Acylcarnitines | 20 | 5 | 15 | 2 | 0,533 | 0,681 | AF |
| Lysophosphatidylcholines | 24 | 1 | 14 | 3 | 5,143 | 0,286 | AF |
| Phosphatidylcholines | 12 | 13 | 10 | 7 | 0,646 | 0,543 | AF |
| Sphingomyelins | 24 | 1 | 15 | 2 | 3,2 | 0,556 | AF |
| Hexoses | 25 | 0 | 17 | 0 | - | - | AF |
| Amino acids | 20 | 3 | 16 | 3 | 1,25 | 1 | HF |
| Biogenic amines | 21 | 2 | 19 | - | - | - | HF |
| Acylcarnitines | 17 | 6 | 18 | 1 | 0,157 | 0,105 | HF |
| Lysophosphatidylcholines | 20 | 3 | 18 | 1 | 0,37 | 0,613 | HF |
| Phosphatidylcholines | 16 | 7 | 6 | 13 | 4,952 | 0,029 | HF |
| Sphingomyelins | 21 | 2 | 18 | 1 | 0,583 | 1 | HF |
| Hexoses | 23 | 0 | 19 | 0 | - | - | HF |
| Amino acids | 32 | 5 | 4 | 1 | 1,6 | 0,557 | DCM |
| Biogenic amines | 35 | 2 | 5 | - | - | - | DCM |
| Acylcarnitines | 34 | 3 | 1 | 4 | 45,333 | 0,001 | DCM |
| Lysophosphatidylcholines | 33 | 4 | 5 | - | - | - | DCM |
| Phosphatidylcholines | 17 | 20 | 5 | - | - | - | DCM |
| Sphingomyelins | 34 | 3 | 5 | - | - | - | DCM |
| Hexoses | 37 | 0 | 5 | 0 | - | - | DCM |
| Amino acids | 35 | 6 | 1 | - | - | - | NICM |
| Biogenic amines | 40 | 1 | - | 1 | - | - | NICM |
| Acylcarnitines | 34 | 7 | 1 | - | - | - | NICM |
| Lysophosphatidylcholines | 37 | 4 | 1 | - | - | - | NICM |
| Phosphatidylcholines | 21 | 20 | 1 | - | - | - | NICM |
| Sphingomyelins | 38 | 3 | 1 | - | - | - | NICM |
| Hexoses | 41 | 0 | 1 | 0 | - | - | NICM |

Abbreviations: AF = atrial fibrillation, DCM = dilated cardiomyopathy, HF = heart failure, NICM = non-ischemic cardiomyopathy, OR = odds ratio
