## Supplemental Table S5 for "Integrating metabolomics and proteomics to identify novel drug targets for heart failure and atrial fibrillation"

| Table S5. Number of overlapping prioritised proteins per outcome | | | | |
|---|---|---|---|---|
| **Outcome** | **AF** | **HF** | **DCM** | **NICM** |
| AF | 42 | 6 | 0 | 0 |
| HF | 6 | 34 | 4 | 1 |
| DCM | 0 | 4 | 14 | 1 |
| NICM | 0 | 1 | 1 | 3 |

Abbreviations: AF = atrial fibrillation,
DCM = dilated cardiomyopathy, HF = heart failure,
NICM = non-ischemic cardiomyopathy
