## Supplemental Table S6 for "Integrating metabolomics and proteomics to identify novel drug targets for heart failure and atrial fibrillation"

| Table S6. HPA RNA over-expression in heart | | | |
|---|---|---|---|
| Protein name | Cardiac RNA expression (nTPM) | Tau | p-value |
| TIG1 | 105 | 0,871 | $1.90\times10^{-4}$ |
| NAR3 | 55,1 | 0,954 | $5.02\times10^{-4}$ |
| DNJA4 | 63,2 | 0,802 | $8.56\times10^{-4}$ |
| RF1ML | 20 | 0,607 | 0,017 |
| ANX11 | 57,1 | 0,665 | 0,036 |
| PLXA1 | 10,2 | 0,803 | 0,045 |
| TENX | 39,3 | 0,815 | 0,113 |
| ACYP2 | 39,6 | 0,883 | 0,137 |
| RTP4 | 2,9 | 0,718 | 0,364 |
| GMPR2 | 15 | 0,568 | 0,402 |
| MINP1 | 7,2 | 0,638 | 0,416 |
| GNPTG | 17,8 | 0,627 | 0,417 |
| CATF | 34,5 | 0,67 | 0,423 |
| FA10 | 18,4 | 0,959 | 0,443 |
| MICA | 10,8 | 0,714 | 0,495 |
| TSP3 | 14,4 | 0,723 | 0,496 |
| LIRB5 | 10 | 0,868 | 0,5 |
| TIMP3 | 75 | 0,918 | 0,542 |
| PLXB2 | 12 | 0,67 | 0,55 |
| TICN2 | 4,7 | 0,795 | 0,55 |
| IBP3 | 41,6 | 0,957 | 0,596 |
| MATN3 | 0,9 | 0,869 | 0,6 |
| KPCA | 5,8 | 0,72 | 0,601 |
| GSTM3 | 27,6 | 0,869 | 0,603 |
| EDAR | 0,5 | 0,858 | 0,614 |
| PVRL4 | 0,1 | 0,913 | 0,642 |
| FCRLB | 0,2 | 0,913 | 0,656 |
| EPHB1 | 1,1 | 0,941 | 0,658 |
| I17RD | 3 | 0,764 | 0,659 |
| IL18R | 10,5 | 0,834 | 0,661 |
| GLCE | 5,5 | 0,864 | 0,663 |
| CN37 | 7,6 | 0,9 | 0,669 |
| GXLT1 | 6,4 | 0,696 | 0,671 |
| C1QRF | 1,8 | 0,898 | 0,687 |
| AT1B2 | 5 | 0,914 | 0,691 |
| TRYB1 | 4,7 | 0,757 | 0,696 |
| ENTP5 | 2,6 | 0,911 | 0,703 |
| NEC1 | 1 | 0,925 | 0,715 |
| CC126 | 5,7 | 0,891 | 0,718 |
| IL6RA | 5 | 0,873 | 0,719 |
| PLA2R | 2,7 | 0,9 | 0,732 |
| TDGF1 | 0,6 | 0,925 | 0,738 |
| NQO1 | 9,3 | 0,895 | 0,742 |
| PCSK9 | 0 | 0,972 | 0,747 |
| RET | 1,2 | 0,954 | 0,758 |
| CACP | 17,3 | 0,886 | 0,766 |

| Table S6. HPA RNA over-expression in heart | | | |
|---|---|---|---|
| Protein name | Cardiac RNA expression (nTPM) | Tau | p-value |
| IGFR1 | 5,8 | 0,927 | 0,772 |
| CECR1 | 15,9 | 0,811 | 0,779 |
| DPEP1 | 0,1 | 0,974 | 0,796 |
| PDIA5 | 7,2 | 0,87 | 0,804 |
| MAX | 30,1 | 0,633 | 0,804 |
| TEC | 3,3 | 0,82 | 0,811 |
| HAVR1 | 0,2 | 0,982 | 0,824 |
| NCTR3 | 1,2 | 0,935 | 0,826 |
| CCL8 | 2,4 | 0,853 | 0,83 |
| FCG3A | 6,4 | 0,959 | 0,84 |
| ENOB | 38,3 | 0,984 | 0,842 |
| SWP70 | 12,5 | 0,677 | 0,846 |
| CAN2 | 33 | 0,525 | 0,853 |
| MICB | 2,2 | 0,905 | 0,855 |
| ISK2 | 2,2 | 0,987 | 0,861 |
| FCG2A | 11,3 | 0,929 | 0,861 |
| ACES | 4,5 | 0,944 | 0,862 |
| REG3G | 0,1 | 0,994 | 0,865 |
| APOC1 | 0,4 | 0,994 | 0,866 |
| FETUB | 0 | 0,995 | 0,868 |
| PZP | 0 | 0,996 | 0,872 |
| GRAB | 4,8 | 0,979 | 0,874 |
| APOC3 | 0,1 | 0,996 | 0,876 |
| ADH4 | 0,1 | 0,998 | 0,89 |
| SPA11 | 0 | 0,999 | 0,895 |
| CYTD | 0 | 1 | 0,897 |
| CEL | 0,2 | 1 | 0,897 |
| KLC1 | 27,1 | 0,807 | 0,899 |
| CL12A | 3,5 | 0,914 | 0,903 |
| ENASE | 10,3 | 0,794 | 0,904 |
| SAT2 | 14,5 | 0,795 | 0,908 |
| GFRA1 | 6,4 | 0,747 | 0,908 |
| ERAP1 | 16,1 | 0,704 | 0,91 |
| FAAA | 6,8 | 0,939 | 0,911 |
| NUDT9 | 18 | 0,715 | 0,912 |
| IDI2 | 1 | 0,999 | 0,936 |
| ATRN | 14,8 | 0,698 | 0,943 |
| TPSN | 22,4 | 0,619 | 0,944 |
| ZA2G | 9 | 0,972 | 0,944 |
| G6PE | 11,9 | 0,908 | 0,97 |
| ADH1B | 24,2 | 0,963 | 0,995 |
