## Supplemental Table S7 for "Integrating metabolomics and proteomics to identify novel drug targets for heart failure and atrial fibrillation"

**Table S7. Final results of all MRs per prioritised protein**

| Protein information | | | | | Effect directions | | | Replications | | | | |
|---|---|---|---|---|---|---|---|---|---|---|---|---|
| Ensembl ID (GRCh38) | Uniprot ID | Protein name | Gene name | Druggability* | Metabolite-Outcome effect* | Protein-Metabolite effect* | Protein-Outcome effect* | Nominal metabolite replicates* | Adjusted metabolite replicates* | Nominal cardiac outcome replicates* | Adjusted cardiac outcome replicates* | Metabolite |
| ENSG00000087085 | P22303 | ACES | ACHE | Drugged | Risk increasing | Increasing | Risk increasing | - | - | - | - | Acetylcarnitine |
| ENSG00000087085 | P22303 | ACES | ACHE | Drugged | Risk increasing | Increasing | Risk increasing | - | - | - | - | Tetradecanoylcarnitine |
| ENSG00000170634 | P14621 | ACYP2 | ACYP2 | Not yet druggable | Risk increasing | Decreasing | Risk decreasing | 0/2 | 0/2 | 0/1 | 0/1 | Aspartate |
| ENSG00000196616 | P00326 | ADH1B | ADH1B | Druggable | Risk increasing | Increasing | Risk increasing | - | - | - | - | Acetylcarnitine |
| ENSG00000198099 | P08319 | ADH4 | ADH4 | Druggable | Risk increasing | Decreasing | Risk decreasing | 0/1 | 0/1 | 0/1 | 0/1 | PC aa C40:4 |
| ENSG00000198099 | P08319 | ADH4 | ADH4 | Druggable | Risk increasing | Decreasing | Risk decreasing | 0/1 | 0/1 | 0/1 | 0/1 | PC aa C42:1 |
| ENSG00000198099 | P08319 | ADH4 | ADH4 | Druggable | Risk increasing | Decreasing | Risk decreasing | 0/1 | 0/1 | 0/1 | 0/1 | PC ae C38:5 |
| ENSG00000198099 | P08319 | ADH4 | ADH4 | Druggable | Risk increasing | Decreasing | Risk decreasing | 1/1 | 0/1 | 0/1 | 0/1 | PC ae C40:5 |
| ENSG00000198099 | P08319 | ADH4 | ADH4 | Druggable | Risk increasing | Decreasing | Risk decreasing | 1/1 | 0/1 | 0/1 | 0/1 | PC aa C36:4 |
| ENSG00000122359 | P50995 | ANX11 | ANXA11 | Not yet druggable | Risk increasing | Decreasing | Risk decreasing | - | - | - | - | Tyrosine |
| ENSG00000130208 | P02654 | APOC1 | APOC1 | Not yet druggable | Risk increasing | Increasing | Risk increasing | - | - | - | - | Octadecanoylcarnitine |
| ENSG00000110245 | P02656 | APOC3 | APOC3 | Druggable | Risk increasing | Increasing | Risk increasing | 1/1 | 1/1 | 1/1 | 0/1 | PC aa C38:4 |
| ENSG00000110245 | P02656 | APOC3 | APOC3 | Druggable | Risk increasing | Increasing | Risk increasing | 1/1 | 1/1 | 1/1 | 0/1 | PC aa C38:5 |
| ENSG00000110245 | P02656 | APOC3 | APOC3 | Druggable | Risk increasing | Increasing | Risk increasing | 1/1 | 0/1 | 1/1 | 0/1 | Acetylcarnitine |
| ENSG00000110245 | P02656 | APOC3 | APOC3 | Druggable | Risk increasing | Increasing | Risk increasing | 1/1 | 1/1 | 1/1 | 0/1 | PC ae C40:5 |
| ENSG00000110245 | P02656 | APOC3 | APOC3 | Druggable | Risk increasing | Increasing | Risk increasing | 1/1 | 1/1 | 0/1 | 0/1 | Hexadecanoylcarnitine |
| ENSG00000110245 | P02656 | APOC3 | APOC3 | Druggable | Risk increasing | Increasing | Risk increasing | 1/1 | 1/1 | 1/1 | 0/1 | PC aa C38:6 |
| ENSG00000110245 | P02656 | APOC3 | APOC3 | Druggable | Risk increasing | Increasing | Risk increasing | 1/1 | 1/1 | 0/1 | 0/1 | Octadecanoylcarnitine |
| ENSG00000129244 | P14415 | AT1B2 | ATP1B2 | Drugged | Risk decreasing | Decreasing | Risk increasing | 1/2 | 1/2 | 2/2 | 2/2 | Taurine |
| ENSG00000088812 | O75882 | ATRN | ATRN | Not yet druggable | Risk increasing | Decreasing | Risk decreasing | - | - | - | - | Tetradecanoylcarnitine |
| ENSG00000131094 | O75973 | C1QRF | C1QL1 | Not yet druggable | Risk increasing | Increasing | Risk increasing | 1/1 | 1/1 | 1/1 | 1/1 | PC aa C38:4 |
| ENSG00000131094 | O75973 | C1QRF | C1QL1 | Not yet druggable | Risk increasing | Increasing | Risk increasing | 1/1 | 0/1 | 1/1 | 1/1 | SM C16:0 |
| ENSG00000095321 | P43155 | CACP | CRAT | Drugged | Risk decreasing | Decreasing | Risk increasing | - | - | - | - | Butyrylcarnitine |
| ENSG00000162909 | P17655 | CAN2 | CAPN2 | Drugged | Risk increasing | Increasing | Risk increasing | 0/1 | 0/1 | 1/1 | 0/1 | Tyrosine |
| ENSG00000162909 | P17655 | CAN2 | CAPN2 | Drugged | Risk increasing | Increasing | Risk increasing | 0/1 | 0/1 | 1/1 | 0/1 | PC ae C38:5 |
| ENSG00000174080 | Q9UBX1 | CATF | CTSF | Drugged | Risk increasing | Decreasing | Risk decreasing | 1/1 | 0/1 | 1/1 | 1/1 | Tyrosine |
| ENSG00000169193 | Q96EE4 | CC126 | CCDC126 | Not yet druggable | Risk increasing | Increasing | Risk increasing | 1/2 | 1/2 | 2/2 | 1/2 | Tyrosine |
| ENSG00000108700 | P80075 | CCL8 | CCL8 | Not yet druggable | Risk increasing | Increasing | Risk increasing | 0/1 | 0/1 | 1/1 | 0/1 | LPC a C28:1 |
| ENSG00000093072 | Q9NZK5 | CECR1 | CECR1 | Not yet druggable | Risk decreasing | Increasing | Risk decreasing | 2/2 | 1/2 | 2/2 | 1/2 | Butyrylcarnitine |
| ENSG00000170835 | P19835 | CEL | CEL | Drugged | Risk decreasing | Increasing | Risk decreasing | 0/2 | 0/2 | 0/2 | 0/2 | Acetylornithine |
| ENSG00000172322 | Q5QGZ9 | CL12A | CLEC12A | Not yet druggable | Risk increasing | Decreasing | Risk decreasing | 2/2 | 2/2 | 2/2 | 2/2 | Tryptophan |
| ENSG00000173786 | P09543 | CN37 | CNP | Not yet druggable | Risk increasing | Decreasing | Risk decreasing | - | - | - | - | Hexadecanoylcarnitine |
| ENSG00000173786 | P09543 | CN37 | CNP | Not yet druggable | Risk increasing | Decreasing | Risk decreasing | - | - | - | - | Octadecanoylcarnitine |
| ENSG00000140403 | Q8WW22 | DNJA4 | DNAJA4 | Not yet druggable | Risk increasing | Increasing | Risk increasing | - | - | - | - | Tryptophan |
| ENSG00000140403 | Q8WW22 | DNJA4 | DNAJA4 | Not yet druggable | Risk decreasing | Decreasing | Risk increasing | - | - | - | - | PC aa C42:6 |
| ENSG00000015413 | P16444 | DPEP1 | DPEP1 | Drugged | Risk decreasing | Increasing | Risk decreasing | - | - | - | - | Acetylornithine |
| ENSG00000135960 | Q9UNE0 | EDAR | EDAR | Druggable | Risk increasing | Decreasing | Risk decreasing | 1/2 | 0/2 | 0/2 | 0/2 | PC ae C42:3 |
| ENSG00000135960 | Q9UNE0 | EDAR | EDAR | Druggable | Risk increasing | Decreasing | Risk decreasing | 2/2 | 1/2 | 0/2 | 0/2 | PC ae C38:4 |
| ENSG00000135960 | Q9UNE0 | EDAR | EDAR | Druggable | Risk increasing | Decreasing | Risk decreasing | 2/2 | 0/2 | 0/2 | 0/2 | PC ae C40:5 |
| ENSG00000135960 | Q9UNE0 | EDAR | EDAR | Druggable | Risk increasing | Decreasing | Risk decreasing | 2/2 | 1/2 | 0/2 | 0/2 | PC aa C38:5 |
| ENSG00000135960 | Q9UNE0 | EDAR | EDAR | Druggable | Risk increasing | Decreasing | Risk decreasing | 1/2 | 0/2 | 0/2 | 0/2 | PC ae C38:5 |
| ENSG00000135960 | Q9UNE0 | EDAR | EDAR | Druggable | Risk increasing | Decreasing | Risk decreasing | 2/2 | 0/2 | 0/2 | 0/2 | PC aa C36:4 |
| ENSG00000167280 | Q8NFI3 | ENASE | ENGASE | Drugged | Risk decreasing | Decreasing | Risk increasing | - | - | - | - | LPC a C20:3 |
| ENSG00000108515 | P13929 | ENOB | ENO3 | Not yet druggable | Risk decreasing | Decreasing | Risk increasing | - | - | - | - | Taurine |
| ENSG00000108515 | P13929 | ENOB | ENO3 | Not yet druggable | Risk decreasing | Decreasing | Risk increasing | - | - | - | - | Butyrylcarnitine |
| ENSG00000108515 | P13929 | ENOB | ENO3 | Not yet druggable | Risk increasing | Increasing | Risk increasing | - | - | - | - | PC ae C40:5 |
| ENSG00000108515 | P13929 | ENOB | ENO3 | Not yet druggable | Risk increasing | Increasing | Risk increasing | - | - | - | - | Tyrosine |
| ENSG00000187097 | O75356 | ENTP5 | ENTPD5 | Drugged | Risk increasing | Decreasing | Risk decreasing | 3/3 | 0/3 | 2/3 | 1/3 | Tetradecanoylcarnitine |
| ENSG00000154928 | P54762 | EPHB1 | EPHB1 | Drugged | Risk decreasing | Increasing | Risk decreasing | - | - | - | - | Serine |
| ENSG00000154928 | P54762 | EPHB1 | EPHB1 | Drugged | Risk increasing | Decreasing | Risk decreasing | - | - | - | - | PC ae C42:3 |
| ENSG00000164307 | Q9NZ08 | ERAP1 | ERAP1 | Druggable | Risk increasing | Decreasing | Risk decreasing | 2/3 | 2/3 | 2/3 | 2/3 | PC aa C38:5 |
| ENSG00000164307 | Q9NZ08 | ERAP1 | ERAP1 | Druggable | Risk increasing | Decreasing | Risk decreasing | 2/3 | 2/3 | 2/3 | 2/3 | PC aa C38:6 |
| ENSG00000126218 | P00742 | FA10 | F10 | Drugged | Risk increasing | Increasing | Risk increasing | 2/2 | 0/2 | 2/2 | 1/2 | PC ae C42:2 |
| ENSG00000126218 | P00742 | FA10 | F10 | Drugged | Risk increasing | Increasing | Risk increasing | 2/2 | 1/2 | 2/2 | 1/2 | PC ae C38:4 |
| ENSG00000103876 | P16930 | FAAA | FAH | Not yet druggable | Risk decreasing | Increasing | Risk decreasing | 0/2 | 0/2 | 2/2 | 1/2 | PC ae C38:3 |
| ENSG00000143226 | P12318 | FCG2A | FCGR2A | Druggable | Risk increasing | Decreasing | Risk decreasing | 0/2 | 0/2 | 2/2 | 2/2 | PC aa C40:4 |
| ENSG00000203747 | P08637 | FCG3A | FCGR3A | Druggable | Risk increasing | Increasing | Risk increasing | - | - | - | - | PC aa C40:4 |
| ENSG00000203747 | P08637 | FCG3A | FCGR3A | Druggable | Risk decreasing | Decreasing | Risk increasing | - | - | - | - | Butyrylcarnitine |
| ENSG00000162746 | Q6BAA4 | FCRLB | FCRLB | Not yet druggable | Risk increasing | Increasing | Risk increasing | - | - | - | - | Aspartate |
| ENSG00000049239 | O95479 | G6PE | H6PD | Not yet druggable | Risk decreasing | Increasing | Risk decreasing | 2/2 | 2/2 | 0/2 | 0/2 | PC aa C42:6 |
| ENSG00000151892 | P56159 | GFRA1 | GFRA1 | Druggable | Risk increasing | Decreasing | Risk decreasing | 0/2 | 0/2 | 2/2 | 0/2 | PC ae C38:4 |
| ENSG00000138604 | O94923 | GLCE | GLCE | Not yet druggable | Risk increasing | Decreasing | Risk decreasing | 1/2 | 1/2 | 2/2 | 1/2 | SM C16:0 |
| ENSG00000138604 | O94923 | GLCE | GLCE | Not yet druggable | Risk decreasing | Increasing | Risk decreasing | 0/2 | 0/2 | 2/2 | 1/2 | PC aa C42:6 |
| ENSG00000100938 | Q9P2T1 | GMPR2 | GMPR2 | Druggable | Risk increasing | Decreasing | Risk decreasing | - | - | - | - | PC aa C38:4 |
| ENSG00000100938 | Q9P2T1 | GMPR2 | GMPR2 | Druggable | Risk increasing | Decreasing | Risk decreasing | - | - | - | - | PC ae C36:5 |
| ENSG00000090581 | Q9UJJ9 | GNPTG | GNPTG | Not yet druggable | Risk increasing | Decreasing | Risk decreasing | 2/2 | 1/2 | 1/2 | 0/2 | SM C16:0 |

Table S7. Final results of all MRs per prioritised pro

| Protein information | | | Metabolite effects on cardiac outcomes | | | | | Protein effects on metabolites | | | | | | Protein effects on cardiac outcomes | | | |
|---|---|---|---|---|---|---|---|---|---|---|---|---|---|---|---|---|---|
| Ensembl ID (GRCh38) | Uniprot ID | Protein name | Metabolite class | No. variants* | OR (95% CI) | p-value | Outcome | Gene name | No. variants* | MD (95% CI) | p-value | Metabolite | Gene name | No. variants* | OR (95% CI) | p-value | Outcome |
| ENSG00000087085 | P22303 | ACES | Acylcarnitines | 52 | 1.10 (1.06; 1.14) | $1.5\times10^{-7}$ | HF | ACHE | 27 | 0.06 (0.04; 0.07) | $6.2\times10^{-11}$ | Acetylcarnitine | ACHE | 25 | 1.06 (1.04; 1.08) | $1.4\times10^{-12}$ | HF |
| ENSG00000087085 | P22303 | ACES | Acylcarnitines | 30 | 1.12 (1.06; 1.18) | $1.8\times10^{-5}$ | AF | ACHE | 25 | 0.05 (0.03; 0.07) | $1.1\times10^{-7}$ | Tetradecanoylcarnitine | ACHE | 23 | 1.09 (1.05; 1.13) | $9.7\times10^{-6}$ | AF |
| ENSG00000170634 | P14621 | ACYP2 | Amino acids | 39 | 1.29 (1.15; 1.43) | $5.3\times10^{-6}$ | AF | ACYP2 | 53 | -0.10 (-0.13; -0.07) | $6.0\times10^{-12}$ | Aspartate | ACYP2 | 49 | 0.84 (0.80; 0.88) | $1.1\times10^{-10}$ | AF |
| ENSG00000196616 | P00326 | ADH1B | Acylcarnitines | 52 | 1.10 (1.06; 1.14) | $1.5\times10^{-7}$ | HF | ADH1B | 27 | 0.13 (0.09; 0.18) | $7.5\times10^{-9}$ | Acetylcarnitine | ADH1B | 23 | 1.14 (1.08; 1.20) | $2.8\times10^{-6}$ | HF |
| ENSG00000198099 | P08319 | ADH4 | Phosphatidylcholines | 24 | 1.12 (1.08; 1.15) | $1.1\times10^{-10}$ | HF | ADH4 | 17 | -0.25 (-0.33; -0.17) | $2.5\times10^{-9}$ | PC aa C40:4 | ADH4 | 15 | 0.70 (0.61; 0.80) | $2.0\times10^{-7}$ | HF |
| ENSG00000198099 | P08319 | ADH4 | Phosphatidylcholines | 15 | 1.15 (1.07; 1.23) | $4.7\times10^{-5}$ | HF | ADH4 | 16 | -0.30 (-0.38; -0.22) | $1.3\times10^{-14}$ | PC aa C42:1 | ADH4 | 15 | 0.70 (0.61; 0.80) | $2.0\times10^{-7}$ | HF |
| ENSG00000198099 | P08319 | ADH4 | Phosphatidylcholines | 42 | 1.05 (1.03; 1.08) | $5.7\times10^{-5}$ | HF | ADH4 | 17 | -0.61 (-0.84; -0.38) | $2.8\times10^{-7}$ | PC ae C38:5 | ADH4 | 15 | 0.70 (0.61; 0.80) | $2.0\times10^{-7}$ | HF |
| ENSG00000198099 | P08319 | ADH4 | Phosphatidylcholines | 52 | 1.07 (1.05; 1.09) | $2.7\times10^{-11}$ | HF | ADH4 | 16 | -0.73 (-0.99; -0.46) | $6.1\times10^{-8}$ | PC ae C40:5 | ADH4 | 15 | 0.70 (0.61; 0.80) | $2.0\times10^{-7}$ | HF |
| ENSG00000198099 | P08319 | ADH4 | Phosphatidylcholines | 61 | 1.07 (1.04; 1.10) | $1.8\times10^{-5}$ | HF | ADH4 | 16 | -0.76 (-1.02; -0.50) | $1.2\times10^{-8}$ | PC aa C36:4 | ADH4 | 15 | 0.70 (0.61; 0.80) | $2.0\times10^{-7}$ | HF |
| ENSG00000122359 | P50995 | ANX11 | Amino acids | 119 | 1.09 (1.05; 1.13) | $2.3\times10^{-6}$ | HF | ANXA11 | 31 | -0.10 (-0.12; -0.07) | $6.7\times10^{-15}$ | Tyrosine | ANXA11 | 29 | 0.86 (0.82; 0.90) | $3.8\times10^{-9}$ | HF |
| ENSG00000130208 | P02654 | APOC1 | Acylcarnitines | 48 | 1.47 (1.25; 1.73) | $2.3\times10^{-6}$ | DCM | APOC1 | 64 | 0.12 (0.08; 0.15) | $1.1\times10^{-10}$ | Octadecanoylcarnitine | APOC1 | 56 | 1.49 (1.26; 1.77) | $3.6\times10^{-6}$ | DCM |
| ENSG00000110245 | P02656 | APOC3 | Phosphatidylcholines | 68 | 1.08 (1.06; 1.11) | $1.2\times10^{-9}$ | HF | APOC3 | 24 | 0.39 (0.30; 0.48) | $1.0\times10^{-100}$ | PC aa C38:4 | APOC3 | 22 | 1.29 (1.18; 1.41) | $5.5\times10^{-8}$ | HF |
| ENSG00000110245 | P02656 | APOC3 | Phosphatidylcholines | 46 | 1.06 (1.03; 1.08) | $1.8\times10^{-6}$ | HF | APOC3 | 22 | 0.25 (0.16; 0.35) | $3.5\times10^{-7}$ | PC aa C38:5 | APOC3 | 22 | 1.29 (1.18; 1.41) | $5.5\times10^{-8}$ | HF |
| ENSG00000110245 | P02656 | APOC3 | Acylcarnitines | 52 | 1.10 (1.06; 1.14) | $1.5\times10^{-7}$ | HF | APOC3 | 24 | 0.19 (0.13; 0.25) | $4.7\times10^{-9}$ | Acetylcarnitine | APOC3 | 22 | 1.29 (1.18; 1.41) | $5.5\times10^{-8}$ | HF |
| ENSG00000110245 | P02656 | APOC3 | Phosphatidylcholines | 52 | 1.07 (1.05; 1.09) | $2.7\times10^{-11}$ | HF | APOC3 | 22 | 0.63 (0.39; 0.87) | $2.0\times10^{-7}$ | PC ae C40:5 | APOC3 | 22 | 1.29 (1.18; 1.41) | $5.5\times10^{-8}$ | HF |
| ENSG00000110245 | P02656 | APOC3 | Acylcarnitines | 63 | 1.51 (1.28; 1.79) | $8.4\times10^{-7}$ | DCM | APOC3 | 26 | 0.19 (0.12; 0.26) | $2.8\times10^{-7}$ | Hexadecanoylcarnitine | APOC3 | 24 | 2.15 (1.62; 2.85) | $1.2\times10^{-7}$ | DCM |
| ENSG00000110245 | P02656 | APOC3 | Phosphatidylcholines | 22 | 1.14 (1.09; 1.20) | $7.0\times10^{-9}$ | HF | APOC3 | 21 | 0.44 (0.33; 0.54) | $2.2\times10^{-16}$ | PC aa C38:6 | APOC3 | 22 | 1.29 (1.18; 1.41) | $5.5\times10^{-8}$ | HF |
| ENSG00000110245 | P02656 | APOC3 | Acylcarnitines | 48 | 1.47 (1.25; 1.73) | $2.3\times10^{-6}$ | DCM | APOC3 | 26 | 0.16 (0.10; 0.22) | $5.3\times10^{-7}$ | Octadecanoylcarnitine | APOC3 | 24 | 2.15 (1.62; 2.85) | $1.2\times10^{-7}$ | DCM |
| ENSG00000129244 | P14415 | AT1B2 | Biogenic amines | 19 | 0.48 (0.38; 0.60) | $4.7\times10^{-10}$ | NICM | ATP1B2 | 69 | -0.11 (-0.15; -0.07) | $3.3\times10^{-7}$ | Taurine | ATP1B2 | 57 | 1.33 (1.19; 1.49) | $5.4\times10^{-7}$ | NICM |
| ENSG00000088812 | O75882 | ATRN | Acylcarnitines | 30 | 1.12 (1.06; 1.18) | $1.8\times10^{-5}$ | AF | ATRN | 63 | -0.05 (-0.07; -0.03) | $3.9\times10^{-7}$ | Tetradecanoylcarnitine | ATRN | 64 | 0.94 (0.93; 0.96) | $5.6\times10^{-13}$ | AF |
| ENSG00000131094 | O75973 | C1QRF | Phosphatidylcholines | 66 | 1.05 (1.03; 1.06) | $2.5\times10^{-14}$ | AF | C1QL1 | 41 | 0.08 (0.05; 0.11) | $7.4\times10^{-8}$ | PC aa C38:4 | C1QL1 | 40 | 1.05 (1.04; 1.07) | $3.3\times10^{-9}$ | AF |
| ENSG00000131094 | O75973 | C1QRF | Sphingomyelins | 78 | 1.09 (1.05; 1.12) | $1.4\times10^{-7}$ | AF | C1QL1 | 45 | 0.07 (0.04; 0.09) | $6.3\times10^{-9}$ | SM C16:0 | C1QL1 | 40 | 1.05 (1.04; 1.07) | $3.3\times10^{-9}$ | AF |
| ENSG00000095321 | P43155 | CACP | Acylcarnitines | 242 | 0.87 (0.82; 0.93) | $4.9\times10^{-5}$ | DCM | CRAT | 10 | -0.31 (-0.42; -0.21) | $1.9\times10^{-9}$ | Butyrylcarnitine | CRAT | 10 | 3.15 (2.03; 4.90) | $3.4\times10^{-7}$ | DCM |
| ENSG00000162909 | P17655 | CAN2 | Amino acids | 119 | 1.09 (1.05; 1.13) | $2.3\times10^{-6}$ | HF | CAPN2 | 17 | 0.11 (0.07; 0.14) | $7.8\times10^{-10}$ | Tyrosine | CAPN2 | 15 | 1.21 (1.13; 1.29) | $2.7\times10^{-8}$ | HF |
| ENSG00000162909 | P17655 | CAN2 | Phosphatidylcholines | 42 | 1.05 (1.03; 1.08) | $5.7\times10^{-5}$ | HF | CAPN2 | 16 | 0.23 (0.14; 0.32) | $5.0\times10^{-7}$ | PC ae C38:5 | CAPN2 | 15 | 1.21 (1.13; 1.29) | $2.7\times10^{-8}$ | HF |
| ENSG00000174080 | Q9UBX1 | CATF | Amino acids | 119 | 1.09 (1.05; 1.13) | $2.3\times10^{-6}$ | HF | CTSF | 20 | -0.14 (-0.18; -0.10) | $2.6\times10^{-12}$ | Tyrosine | CTSF | 18 | 0.92 (0.90; 0.94) | $8.7\times10^{-10}$ | HF |
| ENSG00000169193 | Q96EE4 | CC126 | Amino acids | 119 | 1.09 (1.05; 1.13) | $2.3\times10^{-6}$ | HF | CCDC126 | 53 | 0.04 (0.03; 0.05) | $2.6\times10^{-8}$ | Tyrosine | CCDC126 | 46 | 1.07 (1.04; 1.09) | $2.2\times10^{-9}$ | HF |
| ENSG00000108700 | P80075 | CCL8 | Lysophosphatidylcholines | 8 | 1.13 (1.07; 1.20) | $1.5\times10^{-5}$ | AF | CCL8 | 112 | 0.04 (0.02; 0.06) | $7.2\times10^{-7}$ | LPC a C28:1 | CCL8 | 111 | 1.02 (1.01; 1.03) | $2.5\times10^{-7}$ | AF |
| ENSG00000093072 | Q9NZK5 | CECR1 | Acylcarnitines | 242 | 0.87 (0.82; 0.93) | $4.9\times10^{-5}$ | DCM | CECR1 | 116 | 0.05 (0.03; 0.06) | $9.5\times10^{-9}$ | Butyrylcarnitine | CECR1 | 76 | 0.82 (0.76; 0.90) | $8.3\times10^{-6}$ | DCM |
| ENSG00000170835 | P19835 | CEL | Biogenic amines | 134 | 0.97 (0.96; 0.98) | $9.9\times10^{-8}$ | AF | CEL | 63 | 0.07 (0.05; 0.10) | $8.7\times10^{-10}$ | Acetylornithine | CEL | 63 | 0.95 (0.93; 0.97) | $6.1\times10^{-7}$ | AF |
| ENSG00000172322 | Q5QGZ9 | CL12A | Amino acids | 12 | 1.15 (1.08; 1.23) | $2.8\times10^{-5}$ | HF | CLEC12A | 133 | -0.02 (-0.03; -0.01) | $3.7\times10^{-10}$ | Tryptophan | CLEC12A | 119 | 0.99 (0.98; 0.99) | $2.0\times10^{-6}$ | HF |
| ENSG00000173786 | P09543 | CN37 | Acylcarnitines | 63 | 1.51 (1.28; 1.79) | $8.4\times10^{-7}$ | DCM | CNP | 11 | -0.17 (-0.20; -0.14) | $1.0\times10^{-100}$ | Hexadecanoylcarnitine | CNP | 11 | 0.50 (0.43; 0.57) | $1.0\times10^{-100}$ | DCM |
| ENSG00000173786 | P09543 | CN37 | Acylcarnitines | 48 | 1.47 (1.25; 1.73) | $2.3\times10^{-6}$ | DCM | CNP | 11 | -0.12 (-0.15; -0.08) | $1.5\times10^{-12}$ | Octadecanoylcarnitine | CNP | 11 | 0.50 (0.43; 0.57) | $1.0\times10^{-100}$ | DCM |
| ENSG00000140403 | Q8WW22 | DNJA4 | Amino acids | 12 | 1.15 (1.08; 1.23) | $2.8\times10^{-5}$ | HF | DNAJA4 | 9 | 0.24 (0.15; 0.34) | $3.1\times10^{-7}$ | Tryptophan | DNAJA4 | 9 | 1.23 (1.14; 1.33) | $1.6\times10^{-7}$ | HF |
| ENSG00000140403 | Q8WW22 | DNJA4 | Phosphatidylcholines | 14 | 0.76 (0.68; 0.85) | $2.7\times10^{-6}$ | AF | DNAJA4 | 10 | -0.46 (-0.60; -0.32) | $5.9\times10^{-11}$ | PC aa C42:6 | DNAJA4 | 10 | 1.34 (1.26; 1.44) | $1.0\times10^{-100}$ | AF |
| ENSG00000015413 | P16444 | DPEP1 | Biogenic amines | 134 | 0.97 (0.96; 0.98) | $9.9\times10^{-8}$ | AF | DPEP1 | 70 | 0.07 (0.06; 0.08) | $1.0\times10^{-100}$ | Acetylornithine | DPEP1 | 78 | 0.97 (0.96; 0.98) | $4.0\times10^{-8}$ | AF |
| ENSG00000135960 | Q9UNE0 | EDAR | Phosphatidylcholines | 43 | 1.06 (1.04; 1.08) | $1.3\times10^{-9}$ | HF | EDAR | 50 | -0.13 (-0.18; -0.08) | $6.9\times10^{-8}$ | PC ae C42:3 | EDAR | 43 | 0.92 (0.88; 0.95) | $4.1\times10^{-6}$ | HF |
| ENSG00000135960 | Q9UNE0 | EDAR | Phosphatidylcholines | 42 | 1.19 (1.10; 1.29) | $1.4\times10^{-5}$ | HF | EDAR | 48 | -0.17 (-0.22; -0.13) | $3.3\times10^{-12}$ | PC ae C38:4 | EDAR | 43 | 0.92 (0.88; 0.95) | $4.1\times10^{-6}$ | HF |
| ENSG00000135960 | Q9UNE0 | EDAR | Phosphatidylcholines | 52 | 1.07 (1.05; 1.09) | $2.7\times10^{-11}$ | HF | EDAR | 48 | -0.22 (-0.30; -0.14) | $3.2\times10^{-8}$ | PC ae C40:5 | EDAR | 43 | 0.92 (0.88; 0.95) | $4.1\times10^{-6}$ | HF |
| ENSG00000135960 | Q9UNE0 | EDAR | Phosphatidylcholines | 46 | 1.06 (1.03; 1.08) | $1.8\times10^{-6}$ | HF | EDAR | 48 | -0.21 (-0.29; -0.13) | $1.3\times10^{-7}$ | PC aa C38:5 | EDAR | 43 | 0.92 (0.88; 0.95) | $4.1\times10^{-6}$ | HF |
| ENSG00000135960 | Q9UNE0 | EDAR | Phosphatidylcholines | 42 | 1.05 (1.03; 1.08) | $5.7\times10^{-5}$ | HF | EDAR | 46 | -0.25 (-0.33; -0.17) | $2.0\times10^{-9}$ | PC ae C38:5 | EDAR | 43 | 0.92 (0.88; 0.95) | $4.1\times10^{-6}$ | HF |
| ENSG00000135960 | Q9UNE0 | EDAR | Phosphatidylcholines | 61 | 1.07 (1.04; 1.10) | $1.8\times10^{-5}$ | HF | EDAR | 45 | -0.17 (-0.23; -0.12) | $1.0\times10^{-9}$ | PC aa C36:4 | EDAR | 43 | 0.92 (0.88; 0.95) | $4.1\times10^{-6}$ | HF |
| ENSG00000167280 | Q8NFI3 | ENASE | Lysophosphatidylcholines | 42 | 0.93 (0.91; 0.96) | $2.3\times10^{-7}$ | AF | ENGASE | 118 | -0.06 (-0.08; -0.04) | $2.9\times10^{-7}$ | LPC a C20:3 | ENGASE | 118 | 1.06 (1.03; 1.08) | $2.6\times10^{-7}$ | AF |
| ENSG00000108515 | P13929 | ENOB | Biogenic amines | 19 | 0.48 (0.38; 0.60) | $4.7\times10^{-10}$ | NICM | ENO3 | 62 | -0.10 (-0.12; -0.07) | $1.0\times10^{-100}$ | Taurine | ENO3 | 61 | 1.24 (1.13; 1.35) | $2.6\times10^{-6}$ | NICM |
| ENSG00000108515 | P13929 | ENOB | Acylcarnitines | 242 | 0.87 (0.82; 0.93) | $4.9\times10^{-5}$ | DCM | ENO3 | 66 | -0.06 (-0.08; -0.04) | $2.5\times10^{-11}$ | Butyrylcarnitine | ENO3 | 59 | 1.39 (1.28; 1.50) | $2.4\times10^{-15}$ | DCM |
| ENSG00000108515 | P13929 | ENOB | Phosphatidylcholines | 52 | 1.07 (1.05; 1.09) | $2.7\times10^{-11}$ | HF | ENO3 | 65 | 0.16 (0.10; 0.22) | $3.9\times10^{-7}$ | PC ae C40:5 | ENO3 | 58 | 1.05 (1.03; 1.08) | $9.4\times10^{-6}$ | HF |
| ENSG00000108515 | P13929 | ENOB | Amino acids | 119 | 1.09 (1.05; 1.13) | $2.3\times10^{-6}$ | HF | ENO3 | 65 | 0.02 (0.01; 0.03) | $8.0\times10^{-7}$ | Tyrosine | ENO3 | 58 | 1.05 (1.03; 1.08) | $9.4\times10^{-6}$ | HF |
| ENSG00000187097 | O75356 | ENTP5 | Acylcarnitines | 30 | 1.12 (1.06; 1.18) | $1.8\times10^{-5}$ | AF | ENTPD5 | 34 | -0.13 (-0.16; -0.09) | $2.1\times10^{-10}$ | Tetradecanoylcarnitine | ENTPD5 | 29 | 0.89 (0.86; 0.93) | $5.8\times10^{-9}$ | AF |
| ENSG00000154928 | P54762 | EPHB1 | Amino acids | 77 | 0.90 (0.88; 0.93) | $2.6\times10^{-13}$ | HF | EPHB1 | 98 | 0.14 (0.10; 0.18) | $8.1\times10^{-11}$ | Serine | EPHB1 | 94 | 0.95 (0.93; 0.97) | $7.0\times10^{-7}$ | HF |
| ENSG00000154928 | P54762 | EPHB1 | Phosphatidylcholines | 43 | 1.06 (1.04; 1.08) | $1.3\times10^{-9}$ | HF | EPHB1 | 104 | -0.09 (-0.12; -0.06) | $4.3\times10^{-9}$ | PC ae C42:3 | EPHB1 | 94 | 0.95 (0.93; 0.97) | $7.0\times10^{-7}$ | HF |
| ENSG00000164307 | Q9NZ08 | ERAP1 | Phosphatidylcholines | 46 | 1.06 (1.03; 1.08) | $1.8\times10^{-6}$ | HF | ERAP1 | 79 | -0.03 (-0.04; -0.02) | $7.2\times10^{-7}$ | PC aa C38:5 | ERAP1 | 70 | 0.97 (0.96; 0.98) | $3.5\times10^{-11}$ | HF |
| ENSG00000164307 | Q9NZ08 | ERAP1 | Phosphatidylcholines | 22 | 1.14 (1.09; 1.20) | $7.0\times10^{-9}$ | HF | ERAP1 | 78 | -0.04 (-0.05; -0.02) | $4.9\times10^{-8}$ | PC aa C38:6 | ERAP1 | 70 | 0.97 (0.96; 0.98) | $3.5\times10^{-11}$ | HF |
| ENSG00000126218 | P00742 | FA10 | Phosphatidylcholines | 33 | 1.06 (1.03; 1.08) | $3.1\times10^{-6}$ | AF | F10 | 39 | 0.19 (0.11; 0.26) | $8.6\times10^{-7}$ | PC ae C42:2 | F10 | 37 | 1.11 (1.06; 1.15) | $8.6\times10^{-7}$ | AF |
| ENSG00000126218 | P00742 | FA10 | Phosphatidylcholines | 41 | 1.06 (1.04; 1.08) | $2.4\times10^{-7}$ | AF | F10 | 38 | 0.20 (0.13; 0.28) | $4.2\times10^{-7}$ | PC ae C38:4 | F10 | 37 | 1.11 (1.06; 1.15) | $8.6\times10^{-7}$ | AF |
| ENSG00000103876 | P16930 | FAAA | Phosphatidylcholines | 15 | 0.86 (0.83; 0.90) | $4.7\times10^{-11}$ | AF | FAH | 58 | 0.11 (0.07; 0.15) | $1.5\times10^{-7}$ | PC ae C38:3 | FAH | 55 | 0.90 (0.88; 0.92) | $1.0\times10^{-100}$ | AF |
| ENSG00000143226 | P12318 | FCG2A | Phosphatidylcholines | 24 | 1.12 (1.08; 1.15) | $1.1\times10^{-10}$ | HF | FCGR2A | 93 | -0.02 (-0.03; -0.02) | $8.3\times10^{-9}$ | PC aa C40:4 | FCGR2A | 57 | 0.99 (0.98; 0.99) | $3.5\times10^{-7}$ | HF |
| ENSG00000203747 | P08637 | FCG3A | Phosphatidylcholines | 24 | 1.12 (1.08; 1.15) | $1.1\times10^{-10}$ | HF | FCGR3A | 102 | 0.04 (0.02; 0.05) | $1.5\times10^{-7}$ | PC aa C40:4 | FCGR3A | 74 | 1.01 (1.01; 1.02) | $5.7\times10^{-7}$ | HF |
| ENSG00000203747 | P08637 | FCG3A | Acylcarnitines | 242 | 0.87 (0.82; 0.93) | $4.9\times10^{-5}$ | DCM | FCGR3A | 117 | -0.02 (-0.03; -0.02) | $3.7\times10^{-14}$ | Butyrylcarnitine | FCGR3A | 82 | 1.07 (1.05; 1.10) | $1.8\times10^{-7}$ | DCM |
| ENSG00000162746 | Q6BAA4 | FCRLB | Amino acids | 39 | 1.29 (1.15; 1.43) | $5.3\times10^{-6}$ | AF | FCRLB | 48 | 0.07 (0.04; 0.09) | $2.5\times10^{-7}$ | Aspartate | FCRLB | 48 | 1.13 (1.08; 1.18) | $2.9\times10^{-7}$ | AF |
| ENSG00000049239 | O95479 | G6PE | Phosphatidylcholines | 14 | 0.76 (0.68; 0.85) | $2.7\times10^{-6}$ | AF | H6PD | 111 | 0.09 (0.07; 0.11) | $2.2\times10^{-16}$ | PC aa C42:6 | H6PD | 110 | 0.96 (0.94; 0.98) | $7.8\times10^{-7}$ | AF |
| ENSG00000151892 | P56159 | GFRA1 | Phosphatidylcholines | 41 | 1.06 (1.04; 1.08) | $2.4\times10^{-7}$ | AF | GFRA1 | 28 | -0.16 (-0.21; -0.10) | $1.2\times10^{-7}$ | PC ae C38:4 | GFRA1 | 26 | 0.91 (0.89; 0.94) | $3.4\times10^{-9}$ | AF |
| ENSG00000138604 | O94923 | GLCE | Sphingomyelins | 78 | 1.09 (1.05; 1.12) | $1.4\times10^{-7}$ | AF | GLCE | 53 | -0.05 (-0.07; -0.03) | $4.2\times10^{-7}$ | SM C16:0 | GLCE | 60 | 0.94 (0.93; 0.95) | $1.0\times10^{-100}$ | AF |
| ENSG00000138604 | O94923 | GLCE | Phosphatidylcholines | 14 | 0.76 (0.68; 0.85) | $2.7\times10^{-6}$ | AF | GLCE | 58 | 0.07 (0.05; 0.09) | $5.2\times10^{-9}$ | PC aa C42:6 | GLCE | 60 | 0.94 (0.93; 0.95) | $1.0\times10^{-100}$ | AF |
| ENSG00000100938 | Q9P2T1 | GMPR2 | Phosphatidylcholines | 66 | 1.05 (1.03; 1.06) | $2.5\times10^{-14}$ | AF | GMPR2 | 26 | -0.19 (-0.25; -0.12) | $1.5\times10^{-8}$ | PC aa C38:4 | GMPR2 | 29 | 0.93 (0.90; 0.96) | $3.8\times10^{-6}$ | AF |
| ENSG00000100938 | Q9P2T1 | GMPR2 | Phosphatidylcholines | 50 | 1.04 (1.02; 1.06) | $1.4\times10^{-6}$ | AF | GMPR2 | 27 | -0.19 (-0.25; -0.12) | $5.4\times10^{-9}$ | PC ae C36:5 | GMPR2 | 29 | 0.93 (0.90; 0.96) | $3.8\times10^{-6}$ | AF |
| ENSG00000090581 | Q9UJJ9 | GNPTG | Sphingomyelins | 78 | 1.09 (1.05; 1.12) | $1.4\times10^{-7}$ | AF | GNPTG | 68 | -0.08 (-0.10; -0.05) | $8.8\times10^{-10}$ | SM C16:0 | GNPTG | 70 | 0.94 (0.92; 0.96) | $4.3\times10^{-9}$ | AF |

| Table S7. Final results of all MRs per prioritised protein | | | | | | | | | | | | |
|---|---|---|---|---|---|---|---|---|---|---|---|---|
| Protein information | | | | | Effect directions | | | Replications | | | | |
| Ensembl ID (GRCh38) | Uniprot ID | Protein name | Gene name | Druggability* | Metabolite-Outcome effect* | Protein-Metabolite effect* | Protein-Outcome effect* | Nominal metabolite replicates* | Adjusted metabolite replicates* | Nominal cardiac outcome replicates* | Adjusted cardiac outcome replicates* | Metabolite |
| ENSG00000100453 | P10144 | GRAB | GZMB | Drugged | Risk increasing | Increasing | Risk increasing | 0/1 | 0/1 | 0/1 | 0/1 | PC ae C38:4 |
| ENSG00000100453 | P10144 | GRAB | GZMB | Drugged | Risk increasing | Increasing | Risk increasing | 0/1 | 0/1 | 0/1 | 0/1 | PC ae C36:5 |
| ENSG00000134202 | P21266 | GSTM3 | GSTM3 | Drugged | Risk increasing | Increasing | Risk increasing | 1/1 | 1/1 | 0/1 | 0/1 | SM C16:0 |
| ENSG00000134202 | P21266 | GSTM3 | GSTM3 | Drugged | Risk decreasing | Decreasing | Risk increasing | 1/1 | 0/1 | 0/1 | 0/1 | Acetylornithine |
| ENSG00000151233 | Q4G148 | GXLT1 | GXYLT1 | Not yet druggable | Risk increasing | Increasing | Risk increasing | 2/2 | 1/2 | 2/2 | 1/2 | Octadecadienoylcarnitine |
| ENSG00000113249 | Q96D42 | HAVR1 | HAVCR1 | Not yet druggable | Risk increasing | Increasing | Risk increasing | 3/4 | 0/4 | 2/4 | 1/4 | Tryptophan |
| ENSG00000144730 | Q8NFM7 | I17RD | IL17RD | Not yet druggable | Risk increasing | Decreasing | Risk decreasing | 2/2 | 1/2 | 2/2 | 1/2 | Octadecanoylcarnitine |
| ENSG00000146674 | P17936 | IBP3 | IGFBP3 | Druggable | Risk increasing | Decreasing | Risk decreasing | - | - | - | - | Propionylcarnitine |
| ENSG00000148377 | Q9BXS1 | IDI2 | IDI2 | Not yet druggable | Risk increasing | Increasing | Risk increasing | - | - | - | - | Hexadecanoylcarnitine |
| ENSG00000126246 | Q9H665 | IGFR1 | IGFLR1 | Druggable | Risk decreasing | Decreasing | Risk increasing | 1/2 | 0/2 | 1/2 | 1/2 | Acetylornithine |
| ENSG00000115604 | Q13478 | IL18R | IL18R1 | Druggable | Risk increasing | Decreasing | Risk decreasing | 1/2 | 1/2 | 2/2 | 2/2 | Propionylcarnitine |
| ENSG00000115604 | Q13478 | IL18R | IL18R1 | Druggable | Risk increasing | Decreasing | Risk decreasing | 2/2 | 1/2 | 2/2 | 2/2 | LPC a C26:1 |
| ENSG00000160712 | P08887 | IL6RA | IL6R | Drugged | Risk increasing | Decreasing | Risk decreasing | 4/5 | 2/5 | 5/5 | 5/5 | SM C16:0 |
| ENSG00000160712 | P08887 | IL6RA | IL6R | Drugged | Risk increasing | Decreasing | Risk decreasing | 4/5 | 2/5 | 4/5 | 1/5 | SM C16:0 |
| ENSG00000128040 | P20155 | ISK2 | SPINK2 | Not yet druggable | Risk increasing | Increasing | Risk increasing | 2/2 | 1/2 | 1/2 | 1/2 | PC ae C42:2 |
| ENSG00000126214 | Q07866 | KLC1 | KLC1 | Not yet druggable | Risk increasing | Increasing | Risk increasing | - | - | - | - | Tyrosine |
| ENSG00000126214 | Q07866 | KLC1 | KLC1 | Not yet druggable | Risk increasing | Increasing | Risk increasing | - | - | - | - | SM C16:0 |
| ENSG00000154229 | P17252 | KPCA | PRKCA | Drugged | Risk increasing | Decreasing | Risk decreasing | - | - | - | - | Tetradecanoylcarnitine |
| ENSG00000154229 | P17252 | KPCA | PRKCA | Drugged | Risk increasing | Decreasing | Risk decreasing | - | - | - | - | PC ae C38:4 |
| ENSG00000105609 | O75023 | LIRB5 | LILRB5 | Not yet druggable | Risk increasing | Increasing | Risk increasing | 2/2 | 1/2 | 1/2 | 0/2 | Asparagine |
| ENSG00000132031 | O15232 | MATN3 | MATN3 | Not yet druggable | Risk increasing | Decreasing | Risk decreasing | - | - | - | - | SM C16:0 |
| ENSG00000132031 | O15232 | MATN3 | MATN3 | Not yet druggable | Risk increasing | Decreasing | Risk decreasing | - | - | - | - | PC ae C42:2 |
| ENSG00000125952 | P61244 | MAX | MAX | Druggable | Risk decreasing | Decreasing | Risk increasing | - | - | - | - | LPC a C20:3 |
| ENSG00000204520 | Q29983 | MICA | MICA | Not yet druggable | Risk increasing | Increasing | Risk increasing | - | - | - | - | Asparagine |
| ENSG00000204520 | Q29983 | MICA | MICA | Not yet druggable | Risk increasing | Increasing | Risk increasing | - | - | - | - | Propionylcarnitine |
| ENSG00000204516 | Q29980 | MICB | MICB | Not yet druggable | Risk increasing | Increasing | Risk increasing | - | - | - | - | PC ae C40:5 |
| ENSG00000204516 | Q29980 | MICB | MICB | Not yet druggable | Risk increasing | Increasing | Risk increasing | - | - | - | - | PC ae C38:4 |
| ENSG00000204516 | Q29980 | MICB | MICB | Not yet druggable | Risk increasing | Increasing | Risk increasing | - | - | - | - | PC aa C38:4 |
| ENSG00000107789 | Q9UNW1 | MINP1 | MINPP1 | Not yet druggable | Risk increasing | Decreasing | Risk decreasing | 2/2 | 0/2 | 0/2 | 0/2 | PC aa C38:6 |
| ENSG00000156219 | Q13508 | NAR3 | ART3 | Not yet druggable | Risk increasing | Increasing | Risk increasing | 0/2 | 0/2 | 0/2 | 0/2 | Aspartate |
| ENSG00000156219 | Q13508 | NAR3 | ART3 | Not yet druggable | Risk increasing | Increasing | Risk increasing | 1/2 | 0/2 | 0/2 | 0/2 | LPC a C28:1 |
| ENSG00000204475 | O14931 | NCTR3 | NCR3 | Not yet druggable | Risk increasing | Increasing | Risk increasing | - | - | - | - | PC aa C40:4 |
| ENSG00000175426 | P29120 | NEC1 | PCSK1 | Druggable | Risk increasing | Decreasing | Risk decreasing | 1/2 | 0/2 | 2/2 | 2/2 | SM C16:1 |
| ENSG00000175426 | P29120 | NEC1 | PCSK1 | Druggable | Risk increasing | Decreasing | Risk decreasing | 0/2 | 0/2 | 2/2 | 0/2 | PC aa C36:4 |
| ENSG00000175426 | P29120 | NEC1 | PCSK1 | Druggable | Risk increasing | Decreasing | Risk decreasing | 0/2 | 0/2 | 2/2 | 2/2 | PC ae C38:4 |
| ENSG00000175426 | P29120 | NEC1 | PCSK1 | Druggable | Risk increasing | Decreasing | Risk decreasing | 1/2 | 0/2 | 2/2 | 2/2 | PC ae C36:5 |
| ENSG00000175426 | P29120 | NEC1 | PCSK1 | Druggable | Risk increasing | Decreasing | Risk decreasing | 1/2 | 0/2 | 2/2 | 0/2 | PC ae C36:5 |
| ENSG00000175426 | P29120 | NEC1 | PCSK1 | Druggable | Risk increasing | Decreasing | Risk decreasing | 0/2 | 0/2 | 2/2 | 0/2 | PC ae C38:4 |
| ENSG00000181019 | P15559 | NQO1 | NQO1 | Drugged | Risk increasing | Decreasing | Risk decreasing | 1/2 | 1/2 | 2/2 | 1/2 | SM C16:0 |
| ENSG00000170502 | Q9BW91 | NUDT9 | NUDT9 | Druggable | Risk increasing | Decreasing | Risk decreasing | 2/2 | 1/2 | 0/2 | 0/2 | Hexadecanoylcarnitine |
| ENSG00000065485 | Q14554 | PDIA5 | PDIA5 | Not yet druggable | Risk increasing | Increasing | Risk increasing | 0/2 | 0/2 | 2/2 | 1/2 | Aspartate |
| ENSG00000065485 | Q14554 | PDIA5 | PDIA5 | Not yet druggable | Risk decreasing | Decreasing | Risk increasing | 1/2 | 1/2 | 2/2 | 1/2 | Acetylornithine |
| ENSG00000065485 | Q14554 | PDIA5 | PDIA5 | Not yet druggable | Risk decreasing | Decreasing | Risk increasing | 2/2 | 2/2 | 2/2 | 1/2 | PC ae C38:3 |
| ENSG00000153246 | Q13018 | PLA2R | PLA2R1 | Druggable | Risk increasing | Decreasing | Risk decreasing | 1/2 | 1/2 | 2/2 | 1/2 | Octadecanoylcarnitine |
| ENSG00000153246 | Q13018 | PLA2R | PLA2R1 | Druggable | Risk increasing | Increasing | Risk increasing | 1/2 | 0/2 | 0/2 | 0/2 | Tryptophan |
| ENSG00000153246 | Q13018 | PLA2R | PLA2R1 | Druggable | Risk increasing | Increasing | Risk increasing | 2/2 | 2/2 | 0/2 | 0/2 | Tyrosine |
| ENSG00000114554 | Q9UIW2 | PLXA1 | PLXNA1 | Not yet druggable | Risk increasing | Decreasing | Risk decreasing | 1/1 | 1/1 | 1/1 | 0/1 | SM C16:0 |
| ENSG00000196576 | O15031 | PLXB2 | PLXNB2 | Not yet druggable | Risk increasing | Increasing | Risk increasing | 1/2 | 1/2 | 2/2 | 2/2 | PC ae C42:2 |
| ENSG00000196576 | O15031 | PLXB2 | PLXNB2 | Not yet druggable | Risk increasing | Increasing | Risk increasing | 1/2 | 1/2 | 2/2 | 1/2 | PC ae C42:2 |
| ENSG00000196576 | O15031 | PLXB2 | PLXNB2 | Not yet druggable | Risk increasing | Increasing | Risk increasing | 2/2 | 0/2 | 2/2 | 1/2 | PC aa C42:1 |
| ENSG00000196576 | O15031 | PLXB2 | PLXNB2 | Not yet druggable | Risk increasing | Increasing | Risk increasing | 1/2 | 1/2 | 2/2 | 2/2 | Aspartate |
| ENSG00000143217 | Q96NY8 | PVRL4 | PVRL4 | Drugged | Risk decreasing | Increasing | Risk decreasing | 1/1 | 0/1 | 1/1 | 1/1 | Taurine |
| ENSG00000143954 | Q6UW15 | REG3G | REG3G | Not yet druggable | Risk increasing | Decreasing | Risk decreasing | 0/1 | 0/1 | 1/1 | 1/1 | PC aa C38:4 |
| ENSG00000165731 | P07949 | RET | RET | Drugged | Risk increasing | Decreasing | Risk decreasing | 1/2 | 0/2 | 2/2 | 1/2 | Propionylcarnitine |
| ENSG00000165731 | P07949 | RET | RET | Drugged | Risk increasing | Decreasing | Risk decreasing | 1/2 | 0/2 | 2/2 | 0/2 | Acetylcarnitine |
| ENSG00000165731 | P07949 | RET | RET | Drugged | Risk increasing | Decreasing | Risk decreasing | 2/2 | 2/2 | 2/2 | 0/2 | PC aa C40:4 |
| ENSG00000165731 | P07949 | RET | RET | Drugged | Risk increasing | Decreasing | Risk decreasing | 2/2 | 2/2 | 2/2 | 0/2 | PC aa C36:4 |
| ENSG00000165731 | P07949 | RET | RET | Drugged | Risk increasing | Decreasing | Risk decreasing | 1/2 | 0/2 | 2/2 | 0/2 | PC ae C38:5 |
| ENSG00000165731 | P07949 | RET | RET | Drugged | Risk increasing | Decreasing | Risk decreasing | 0/2 | 0/2 | 2/2 | 1/2 | Asparagine |
| ENSG00000165731 | P07949 | RET | RET | Drugged | Risk increasing | Decreasing | Risk decreasing | 1/2 | 0/2 | 2/2 | 0/2 | PC aa C42:1 |
| ENSG00000112031 | Q9UGC7 | RF1ML | MTRF1L | Not yet druggable | Risk increasing | Increasing | Risk increasing | - | - | - | - | Octadecanoylcarnitine |
| ENSG00000136514 | Q96DX8 | RTP4 | RTP4 | Not yet druggable | Risk increasing | Increasing | Risk increasing | - | - | - | - | Tryptophan |
| ENSG00000141504 | Q96F10 | SAT2 | SAT2 | Drugged | Risk decreasing | Increasing | Risk decreasing | - | - | - | - | Butyrylcarnitine |
| ENSG00000141504 | Q96F10 | SAT2 | SAT2 | Drugged | Risk increasing | Decreasing | Risk decreasing | - | - | - | - | Octadecadienoylcarnitine |
| ENSG00000141504 | Q96F10 | SAT2 | SAT2 | Drugged | Risk increasing | Decreasing | Risk decreasing | - | - | - | - | Octadecanoylcarnitine |
| ENSG00000133789 | Q9UH65 | SWP70 | SWAP70 | Not yet druggable | Risk increasing | Decreasing | Risk decreasing | 1/2 | 1/2 | 1/2 | 0/2 | PC aa C38:6 |

Table S7. Final results of all MRs per prioritised prc

| Protein informatio | | | Metabolite effects on cardiac outcomes | | | | | Protein effects on metabolites | | | | | | Protein effects on cardiac outcomes | | | |
|---|---|---|---|---|---|---|---|---|---|---|---|---|---|---|---|---|---|
| Ensembl ID (GRCh38) | Uniprot ID | Protein name | Metabolite class | No. variants* | OR (95% CI) | p-value | Outcome | Gene name | No. variants* | MD (95% CI) | p-value | Metabolite | Gene name | No. variants* | OR (95% CI) | p-value | Outcome |
| ENSG00000100453 | P10144 | GRAB | Phosphatidylcholines | 42 | 1.19 (1.10; 1.29) | $1.4\times10^{-5}$ | HF | GZMB | 12 | 0.71 (0.46; 0.96) | $3.4\times10^{-8}$ | PC ae C38:4 | GZMB | 12 | 1.21 (1.11; 1.32) | $1.5\times10^{-5}$ | HF |
| ENSG00000100453 | P10144 | GRAB | Phosphatidylcholines | 50 | 1.05 (1.03; 1.07) | $9.5\times10^{-6}$ | HF | GZMB | 12 | 0.68 (0.43; 0.93) | $1.0\times10^{-7}$ | PC ae C36:5 | GZMB | 12 | 1.21 (1.11; 1.32) | $1.5\times10^{-5}$ | HF |
| ENSG00000134202 | P21266 | GSTM3 | Sphingomyelins | 78 | 1.09 (1.05; 1.12) | $1.4\times10^{-7}$ | AF | GSTM3 | 105 | 0.03 (0.02; 0.04) | $3.9\times10^{-7}$ | SM C16:0 | GSTM3 | 109 | 1.04 (1.02; 1.05) | $3.1\times10^{-7}$ | AF |
| ENSG00000134202 | P21266 | GSTM3 | Biogenic amines | 134 | 0.97 (0.96; 0.98) | $9.9\times10^{-8}$ | AF | GSTM3 | 109 | -0.03 (-0.04; -0.02) | $3.1\times10^{-8}$ | Acetylornithine | GSTM3 | 109 | 1.04 (1.02; 1.05) | $3.1\times10^{-7}$ | AF |
| ENSG00000151233 | Q4G148 | GXLT1 | Acylcarnitines | 63 | 1.38 (1.22; 1.55) | $2.2\times10^{-7}$ | DCM | GXYLT1 | 41 | 0.12 (0.09; 0.15) | $4.0\times10^{-15}$ | Octadecadienoylcarnitine | GXYLT1 | 35 | 1.68 (1.34; 2.10) | $6.0\times10^{-6}$ | DCM |
| ENSG00000113249 | Q96D42 | HAVR1 | Amino acids | 12 | 1.15 (1.08; 1.23) | $2.8\times10^{-5}$ | HF | HAVCR1 | 62 | 0.04 (0.02; 0.06) | $6.8\times10^{-7}$ | Tryptophan | HAVCR1 | 52 | 1.06 (1.04; 1.08) | $7.6\times10^{-8}$ | HF |
| ENSG00000144730 | Q8NFM7 | I17RD | Acylcarnitines | 48 | 1.47 (1.25; 1.73) | $2.3\times10^{-6}$ | DCM | IL17RD | 60 | -0.05 (-0.07; -0.03) | $7.2\times10^{-9}$ | Octadecanoylcarnitine | IL17RD | 64 | 0.76 (0.68; 0.86) | $1.0\times10^{-5}$ | DCM |
| ENSG00000146674 | P17936 | IBP3 | Acylcarnitines | 99 | 1.06 (1.03; 1.08) | $6.1\times10^{-7}$ | AF | IGFBP3 | 47 | -0.09 (-0.12; -0.06) | $1.3\times10^{-8}$ | Propionylcarnitine | IGFBP3 | 48 | 0.93 (0.91; 0.96) | $3.2\times10^{-6}$ | AF |
| ENSG00000148377 | Q9BXS1 | IDI2 | Acylcarnitines | 63 | 1.51 (1.28; 1.79) | $8.4\times10^{-7}$ | DCM | IDI2 | 30 | 0.10 (0.06; 0.14) | $1.0\times10^{-7}$ | Hexadecanoylcarnitine | IDI2 | 28 | 1.94 (1.57; 2.41) | $1.7\times10^{-9}$ | DCM |
| ENSG00000126246 | Q9H665 | IGFR1 | Biogenic amines | 134 | 0.97 (0.96; 0.98) | $9.9\times10^{-8}$ | AF | IGFLR1 | 61 | -0.04 (-0.06; -0.03) | $5.8\times10^{-7}$ | Acetylornithine | IGFLR1 | 63 | 1.05 (1.03; 1.08) | $6.5\times10^{-8}$ | AF |
| ENSG00000115604 | Q13478 | IL18R | Acylcarnitines | 99 | 1.06 (1.03; 1.08) | $6.1\times10^{-7}$ | AF | IL18R1 | 48 | -0.04 (-0.05; -0.03) | $1.3\times10^{-9}$ | Propionylcarnitine | IL18R1 | 43 | 0.95 (0.94; 0.97) | $3.8\times10^{-11}$ | AF |
| ENSG00000115604 | Q13478 | IL18R | Lysophosphatidylcholines | 25 | 1.06 (1.03; 1.09) | $6.8\times10^{-5}$ | AF | IL18R1 | 57 | -0.05 (-0.07; -0.03) | $2.1\times10^{-9}$ | LPC a C26:1 | IL18R1 | 43 | 0.95 (0.94; 0.97) | $3.8\times10^{-11}$ | AF |
| ENSG00000160712 | P08887 | IL6RA | Sphingomyelins | 78 | 1.09 (1.05; 1.12) | $1.4\times10^{-7}$ | AF | IL6R | 71 | -0.03 (-0.04; -0.02) | $2.2\times10^{-11}$ | SM C16:0 | IL6R | 66 | 0.96 (0.96; 0.97) | $9.1\times10^{-15}$ | AF |
| ENSG00000160712 | P08887 | IL6RA | Sphingomyelins | 67 | 1.10 (1.06; 1.14) | $3.3\times10^{-6}$ | HF | IL6R | 71 | -0.03 (-0.04; -0.02) | $2.2\times10^{-11}$ | SM C16:0 | IL6R | 58 | 0.98 (0.97; 0.99) | $6.0\times10^{-7}$ | HF |
| ENSG00000128040 | P20155 | ISK2 | Phosphatidylcholines | 33 | 1.06 (1.03; 1.08) | $3.1\times10^{-6}$ | AF | SPINK2 | 58 | 0.13 (0.09; 0.18) | $6.5\times10^{-10}$ | PC ae C42:2 | SPINK2 | 63 | 1.10 (1.07; 1.14) | $7.8\times10^{-9}$ | AF |
| ENSG00000126214 | Q07866 | KLC1 | Amino acids | 119 | 1.09 (1.05; 1.13) | $2.3\times10^{-6}$ | HF | KLC1 | 12 | 0.18 (0.12; 0.23) | $3.7\times10^{-10}$ | Tyrosine | KLC1 | 12 | 1.28 (1.20; 1.38) | $1.5\times10^{-12}$ | HF |
| ENSG00000126214 | Q07866 | KLC1 | Sphingomyelins | 67 | 1.10 (1.06; 1.14) | $3.3\times10^{-6}$ | HF | KLC1 | 12 | 0.26 (0.17; 0.36) | $5.1\times10^{-8}$ | SM C16:0 | KLC1 | 12 | 1.28 (1.20; 1.38) | $1.5\times10^{-12}$ | HF |
| ENSG00000154229 | P17252 | KPCA | Acylcarnitines | 30 | 1.12 (1.06; 1.18) | $1.8\times10^{-5}$ | AF | PRKCA | 13 | -0.50 (-0.62; -0.38) | $2.2\times10^{-16}$ | Tetradecanoylcarnitine | PRKCA | 14 | 0.81 (0.74; 0.89) | $1.8\times10^{-5}$ | AF |
| ENSG00000154229 | P17252 | KPCA | Phosphatidylcholines | 41 | 1.06 (1.04; 1.08) | $2.4\times10^{-7}$ | AF | PRKCA | 13 | -0.61 (-0.84; -0.37) | $4.6\times10^{-7}$ | PC ae C38:4 | PRKCA | 14 | 0.81 (0.74; 0.89) | $1.8\times10^{-5}$ | AF |
| ENSG00000105609 | O75023 | LIRB5 | Amino acids | 68 | 1.06 (1.04; 1.09) | $4.3\times10^{-7}$ | AF | LILRB5 | 173 | 0.02 (0.01; 0.03) | $4.9\times10^{-8}$ | Asparagine | LILRB5 | 178 | 1.02 (1.01; 1.03) | $2.3\times10^{-6}$ | AF |
| ENSG00000132031 | O15232 | MATN3 | Sphingomyelins | 78 | 1.09 (1.05; 1.12) | $1.4\times10^{-7}$ | AF | MATN3 | 93 | -0.03 (-0.04; -0.02) | $5.6\times10^{-12}$ | SM C16:0 | MATN3 | 97 | 0.97 (0.96; 0.98) | $8.6\times10^{-10}$ | AF |
| ENSG00000132031 | O15232 | MATN3 | Phosphatidylcholines | 33 | 1.06 (1.03; 1.08) | $3.1\times10^{-6}$ | AF | MATN3 | 85 | -0.06 (-0.08; -0.04) | $2.4\times10^{-7}$ | PC ae C42:2 | MATN3 | 97 | 0.97 (0.96; 0.98) | $8.6\times10^{-10}$ | AF |
| ENSG00000125952 | P61244 | MAX | Lysophosphatidylcholines | 42 | 0.93 (0.91; 0.96) | $2.3\times10^{-7}$ | AF | MAX | 40 | -0.26 (-0.33; -0.20) | $5.4\times10^{-14}$ | LPC a C20:3 | MAX | 36 | 1.09 (1.05; 1.14) | $4.7\times10^{-6}$ | AF |
| ENSG00000204520 | Q29983 | MICA | Amino acids | 68 | 1.06 (1.04; 1.09) | $4.3\times10^{-7}$ | AF | MICA | 49 | 0.03 (0.02; 0.04) | $9.4\times10^{-14}$ | Asparagine | MICA | 53 | 1.02 (1.02; 1.03) | $1.0\times10^{-100}$ | AF |
| ENSG00000204520 | Q29983 | MICA | Acylcarnitines | 99 | 1.06 (1.03; 1.08) | $6.1\times10^{-7}$ | AF | MICA | 52 | 0.02 (0.01; 0.03) | $1.6\times10^{-7}$ | Propionylcarnitine | MICA | 53 | 1.02 (1.02; 1.03) | $1.0\times10^{-100}$ | AF |
| ENSG00000204516 | Q29980 | MICB | Phosphatidylcholines | 52 | 1.07 (1.05; 1.09) | $2.7\times10^{-11}$ | HF | MICB | 55 | 0.03 (0.02; 0.04) | $1.0\times10^{-9}$ | PC ae C40:5 | MICB | 39 | 1.05 (1.04; 1.07) | $1.0\times10^{-100}$ | HF |
| ENSG00000204516 | Q29980 | MICB | Phosphatidylcholines | 42 | 1.19 (1.10; 1.29) | $1.4\times10^{-5}$ | HF | MICB | 53 | 0.03 (0.02; 0.03) | $4.6\times10^{-7}$ | PC ae C38:4 | MICB | 39 | 1.05 (1.04; 1.07) | $1.0\times10^{-100}$ | HF |
| ENSG00000204516 | Q29980 | MICB | Phosphatidylcholines | 68 | 1.08 (1.06; 1.11) | $1.2\times10^{-9}$ | HF | MICB | 57 | 0.04 (0.02; 0.06) | $4.0\times10^{-7}$ | PC aa C38:4 | MICB | 39 | 1.05 (1.04; 1.07) | $1.0\times10^{-100}$ | HF |
| ENSG00000107789 | Q9UNW1 | MINP1 | Phosphatidylcholines | 22 | 1.14 (1.09; 1.20) | $7.0\times10^{-9}$ | HF | MINPP1 | 16 | -0.43 (-0.58; -0.29) | $4.0\times10^{-9}$ | PC aa C38:6 | MINPP1 | 15 | 0.80 (0.73; 0.88) | $5.6\times10^{-6}$ | HF |
| ENSG00000156219 | Q13508 | NAR3 | Amino acids | 39 | 1.29 (1.15; 1.43) | $5.3\times10^{-6}$ | AF | ART3 | 60 | 0.06 (0.04; 0.08) | $2.3\times10^{-10}$ | Aspartate | ART3 | 60 | 1.07 (1.04; 1.11) | $1.6\times10^{-5}$ | AF |
| ENSG00000156219 | Q13508 | NAR3 | Lysophosphatidylcholines | 8 | 1.13 (1.07; 1.20) | $1.5\times10^{-5}$ | AF | ART3 | 63 | 0.08 (0.05; 0.11) | $9.5\times10^{-9}$ | LPC a C28:1 | ART3 | 60 | 1.07 (1.04; 1.11) | $1.6\times10^{-5}$ | AF |
| ENSG00000204475 | O14931 | NCTR3 | Phosphatidylcholines | 24 | 1.12 (1.08; 1.15) | $1.1\times10^{-10}$ | HF | NCR3 | 41 | 0.07 (0.04; 0.09) | $5.0\times10^{-7}$ | PC aa C40:4 | NCR3 | 37 | 1.03 (1.02; 1.05) | $7.1\times10^{-6}$ | HF |
| ENSG00000175426 | P29120 | NEC1 | Sphingomyelins | 17 | 1.12 (1.08; 1.17) | $3.2\times10^{-8}$ | AF | PCSK1 | 91 | -0.05 (-0.07; -0.03) | $1.5\times10^{-7}$ | SM C16:1 | PCSK1 | 93 | 0.96 (0.94; 0.97) | $1.0\times10^{-6}$ | AF |
| ENSG00000175426 | P29120 | NEC1 | Phosphatidylcholines | 61 | 1.07 (1.04; 1.10) | $1.8\times10^{-5}$ | HF | PCSK1 | 88 | -0.07 (-0.09; -0.05) | $2.8\times10^{-10}$ | PC aa C36:4 | PCSK1 | 76 | 0.97 (0.96; 0.99) | $1.1\times10^{-5}$ | HF |
| ENSG00000175426 | P29120 | NEC1 | Phosphatidylcholines | 41 | 1.06 (1.04; 1.08) | $2.4\times10^{-7}$ | AF | PCSK1 | 92 | -0.06 (-0.08; -0.04) | $6.4\times10^{-9}$ | PC ae C38:4 | PCSK1 | 93 | 0.96 (0.94; 0.97) | $1.0\times10^{-6}$ | AF |
| ENSG00000175426 | P29120 | NEC1 | Phosphatidylcholines | 50 | 1.04 (1.02; 1.06) | $1.4\times10^{-6}$ | AF | PCSK1 | 91 | -0.07 (-0.09; -0.05) | $9.1\times10^{-11}$ | PC ae C36:5 | PCSK1 | 93 | 0.96 (0.94; 0.97) | $1.0\times10^{-6}$ | AF |
| ENSG00000175426 | P29120 | NEC1 | Phosphatidylcholines | 50 | 1.05 (1.03; 1.07) | $9.5\times10^{-6}$ | HF | PCSK1 | 91 | -0.07 (-0.09; -0.05) | $9.1\times10^{-11}$ | PC ae C36:5 | PCSK1 | 76 | 0.97 (0.96; 0.99) | $1.1\times10^{-5}$ | HF |
| ENSG00000175426 | P29120 | NEC1 | Phosphatidylcholines | 42 | 1.19 (1.10; 1.29) | $1.4\times10^{-5}$ | HF | PCSK1 | 92 | -0.06 (-0.08; -0.04) | $6.4\times10^{-9}$ | PC ae C38:4 | PCSK1 | 76 | 0.97 (0.96; 0.99) | $1.1\times10^{-5}$ | HF |
| ENSG00000181019 | P15559 | NQO1 | Sphingomyelins | 67 | 1.10 (1.06; 1.14) | $3.3\times10^{-6}$ | HF | NQO1 | 48 | -0.03 (-0.04; -0.02) | $1.3\times10^{-7}$ | SM C16:0 | NQO1 | 30 | 0.97 (0.96; 0.98) | $1.1\times10^{-9}$ | HF |
| ENSG00000170502 | Q9BW91 | NUDT9 | Acylcarnitines | 63 | 1.51 (1.28; 1.79) | $8.4\times10^{-7}$ | DCM | NUDT9 | 24 | -0.14 (-0.19; -0.09) | $4.9\times10^{-7}$ | Hexadecanoylcarnitine | NUDT9 | 23 | 0.27 (0.15; 0.49) | $9.6\times10^{-6}$ | DCM |
| ENSG00000065485 | Q14554 | PDIA5 | Amino acids | 39 | 1.29 (1.15; 1.43) | $5.3\times10^{-6}$ | AF | PDIA5 | 125 | 0.04 (0.03; 0.06) | $5.2\times10^{-10}$ | Aspartate | PDIA5 | 121 | 1.03 (1.02; 1.05) | $1.5\times10^{-6}$ | AF |
| ENSG00000065485 | Q14554 | PDIA5 | Biogenic amines | 134 | 0.97 (0.96; 0.98) | $9.9\times10^{-8}$ | AF | PDIA5 | 125 | -0.06 (-0.09; -0.04) | $2.7\times10^{-7}$ | Acetylornithine | PDIA5 | 121 | 1.03 (1.02; 1.05) | $1.5\times10^{-6}$ | AF |
| ENSG00000065485 | Q14554 | PDIA5 | Phosphatidylcholines | 15 | 0.86 (0.83; 0.90) | $4.7\times10^{-11}$ | AF | PDIA5 | 127 | -0.06 (-0.08; -0.05) | $2.3\times10^{-11}$ | PC ae C38:3 | PDIA5 | 121 | 1.03 (1.02; 1.05) | $1.5\times10^{-6}$ | AF |
| ENSG00000153246 | Q13018 | PLA2R | Acylcarnitines | 48 | 1.47 (1.25; 1.73) | $2.3\times10^{-6}$ | DCM | PLA2R1 | 87 | -0.02 (-0.02; -0.01) | $4.9\times10^{-12}$ | Octadecanoylcarnitine | PLA2R1 | 79 | 0.92 (0.91; 0.94) | $1.0\times10^{-100}$ | DCM |
| ENSG00000153246 | Q13018 | PLA2R | Amino acids | 12 | 1.15 (1.08; 1.23) | $2.8\times10^{-5}$ | HF | PLA2R1 | 90 | 0.01 (0.01; 0.02) | $2.5\times10^{-9}$ | Tryptophan | PLA2R1 | 79 | 1.01 (1.01; 1.02) | $8.6\times10^{-6}$ | HF |
| ENSG00000153246 | Q13018 | PLA2R | Amino acids | 119 | 1.09 (1.05; 1.13) | $2.3\times10^{-6}$ | HF | PLA2R1 | 87 | 0.02 (0.02; 0.02) | $1.0\times10^{-100}$ | Tyrosine | PLA2R1 | 79 | 1.01 (1.01; 1.02) | $8.6\times10^{-6}$ | HF |
| ENSG00000114554 | Q9UIW2 | PLXA1 | Sphingomyelins | 67 | 1.10 (1.06; 1.14) | $3.3\times10^{-6}$ | HF | PLXNA1 | 37 | -0.09 (-0.12; -0.05) | $8.2\times10^{-8}$ | SM C16:0 | PLXNA1 | 33 | 0.90 (0.87; 0.92) | $1.0\times10^{-100}$ | HF |
| ENSG00000196576 | O15031 | PLXB2 | Phosphatidylcholines | 33 | 1.06 (1.03; 1.08) | $3.1\times10^{-6}$ | AF | PLXNB2 | 94 | 0.09 (0.05; 0.12) | $8.3\times10^{-8}$ | PC ae C42:2 | PLXNB2 | 99 | 1.05 (1.04; 1.06) | $1.0\times10^{-100}$ | AF |
| ENSG00000196576 | O15031 | PLXB2 | Phosphatidylcholines | 34 | 1.09 (1.06; 1.12) | $2.9\times10^{-8}$ | HF | PLXNB2 | 94 | 0.09 (0.05; 0.12) | $8.3\times10^{-8}$ | PC ae C42:2 | PLXNB2 | 87 | 1.03 (1.02; 1.04) | $1.6\times10^{-7}$ | HF |
| ENSG00000196576 | O15031 | PLXB2 | Phosphatidylcholines | 15 | 1.15 (1.07; 1.23) | $4.7\times10^{-5}$ | HF | PLXNB2 | 105 | 0.12 (0.09; 0.14) | $1.0\times10^{-100}$ | PC aa C42:1 | PLXNB2 | 87 | 1.03 (1.02; 1.04) | $1.6\times10^{-7}$ | HF |
| ENSG00000196576 | O15031 | PLXB2 | Amino acids | 39 | 1.29 (1.15; 1.43) | $5.3\times10^{-6}$ | AF | PLXNB2 | 100 | 0.03 (0.02; 0.04) | $2.5\times10^{-8}$ | Aspartate | PLXNB2 | 99 | 1.05 (1.04; 1.06) | $1.0\times10^{-100}$ | AF |
| ENSG00000143217 | Q96NY8 | PVRL4 | Biogenic amines | 19 | 0.48 (0.38; 0.60) | $4.7\times10^{-10}$ | NICM | PVRL4 | 10 | 0.43 (0.30; 0.56) | $7.7\times10^{-11}$ | Taurine | PVRL4 | 10 | 0.32 (0.20; 0.51) | $1.3\times10^{-6}$ | NICM |
| ENSG00000143954 | Q6UW15 | REG3G | Phosphatidylcholines | 66 | 1.05 (1.03; 1.06) | $2.5\times10^{-14}$ | AF | REG3G | 87 | -0.06 (-0.08; -0.04) | $1.8\times10^{-7}$ | PC aa C38:4 | REG3G | 82 | 0.97 (0.96; 0.98) | $3.2\times10^{-6}$ | AF |
| ENSG00000165731 | P07949 | RET | Acylcarnitines | 99 | 1.06 (1.03; 1.08) | $6.1\times10^{-7}$ | AF | RET | 64 | -0.04 (-0.05; -0.03) | $1.0\times10^{-100}$ | Propionylcarnitine | RET | 63 | 0.94 (0.93; 0.95) | $1.0\times10^{-100}$ | AF |
| ENSG00000165731 | P07949 | RET | Acylcarnitines | 52 | 1.10 (1.06; 1.14) | $1.5\times10^{-7}$ | HF | RET | 68 | -0.03 (-0.04; -0.02) | $7.0\times10^{-10}$ | Acetylcarnitine | RET | 65 | 0.97 (0.96; 0.98) | $2.2\times10^{-10}$ | HF |
| ENSG00000165731 | P07949 | RET | Phosphatidylcholines | 24 | 1.12 (1.08; 1.15) | $1.1\times10^{-10}$ | HF | RET | 66 | -0.08 (-0.09; -0.06) | $1.0\times10^{-100}$ | PC aa C40:4 | RET | 65 | 0.97 (0.96; 0.98) | $2.2\times10^{-10}$ | HF |
| ENSG00000165731 | P07949 | RET | Phosphatidylcholines | 61 | 1.07 (1.04; 1.10) | $1.8\times10^{-5}$ | HF | RET | 66 | -0.09 (-0.13; -0.06) | $2.9\times10^{-7}$ | PC aa C36:4 | RET | 65 | 0.97 (0.96; 0.98) | $2.2\times10^{-10}$ | HF |
| ENSG00000165731 | P07949 | RET | Phosphatidylcholines | 42 | 1.05 (1.03; 1.08) | $5.7\times10^{-5}$ | HF | RET | 65 | -0.04 (-0.06; -0.03) | $3.7\times10^{-7}$ | PC ae C38:5 | RET | 65 | 0.97 (0.96; 0.98) | $2.2\times10^{-10}$ | HF |
| ENSG00000165731 | P07949 | RET | Amino acids | 68 | 1.06 (1.04; 1.09) | $4.3\times10^{-7}$ | AF | RET | 61 | -0.04 (-0.05; -0.02) | $7.5\times10^{-10}$ | Asparagine | RET | 63 | 0.94 (0.93; 0.95) | $1.0\times10^{-100}$ | AF |
| ENSG00000165731 | P07949 | RET | Phosphatidylcholines | 15 | 1.15 (1.07; 1.23) | $4.7\times10^{-5}$ | HF | RET | 61 | -0.11 (-0.15; -0.06) | $4.6\times10^{-7}$ | PC aa C42:1 | RET | 65 | 0.97 (0.96; 0.98) | $2.2\times10^{-10}$ | HF |
| ENSG00000112031 | Q9UGC7 | RF1ML | Acylcarnitines | 48 | 1.47 (1.25; 1.73) | $2.3\times10^{-6}$ | DCM | MTRF1L | 36 | 0.27 (0.17; 0.36) | $1.2\times10^{-7}$ | Octadecanoylcarnitine | MTRF1L | 34 | 2.35 (1.93; 2.86) | $1.0\times10^{-100}$ | DCM |
| ENSG00000136514 | Q96DX8 | RTP4 | Amino acids | 12 | 1.15 (1.08; 1.23) | $2.8\times10^{-5}$ | HF | RTP4 | 71 | 0.06 (0.04; 0.08) | $5.7\times10^{-7}$ | Tryptophan | RTP4 | 71 | 1.08 (1.06; 1.11) | $6.4\times10^{-13}$ | HF |
| ENSG00000141504 | Q96F10 | SAT2 | Acylcarnitines | 242 | 0.87 (0.82; 0.93) | $4.9\times10^{-5}$ | DCM | SAT2 | 48 | 0.08 (0.05; 0.11) | $6.3\times10^{-7}$ | Butyrylcarnitine | SAT2 | 45 | 0.73 (0.64; 0.84) | $8.7\times10^{-6}$ | DCM |
| ENSG00000141504 | Q96F10 | SAT2 | Acylcarnitines | 63 | 1.38 (1.22; 1.55) | $2.2\times10^{-7}$ | DCM | SAT2 | 47 | -0.08 (-0.11; -0.05) | $2.3\times10^{-7}$ | Octadecadienoylcarnitine | SAT2 | 45 | 0.73 (0.64; 0.84) | $8.7\times10^{-6}$ | DCM |
| ENSG00000141504 | Q96F10 | SAT2 | Acylcarnitines | 48 | 1.47 (1.25; 1.73) | $2.3\times10^{-6}$ | DCM | SAT2 | 46 | -0.09 (-0.12; -0.06) | $4.6\times10^{-8}$ | Octadecanoylcarnitine | SAT2 | 45 | 0.73 (0.64; 0.84) | $8.7\times10^{-6}$ | DCM |
| ENSG00000133789 | Q9UH65 | SWP70 | Phosphatidylcholines | 22 | 1.14 (1.09; 1.20) | $7.0\times10^{-9}$ | HF | SWAP70 | 79 | -0.07 (-0.09; -0.05) | $1.5\times10^{-9}$ | PC aa C38:6 | SWAP70 | 65 | 0.96 (0.94; 0.97) | $4.0\times10^{-9}$ | HF |

**Table S7. Final results of all MRs per prioritised protein**

| Protein information | | | | | Effect directions | | | Replications | | | | |
|---|---|---|---|---|---|---|---|---|---|---|---|---|
| **Ensembl ID (GRCh38)** | **Uniprot ID** | **Protein name** | **Gene name** | **Druggability*** | **Metabolite-Outcome effect*** | **Protein-Metabolite effect*** | **Protein-Outcome effect*** | **Nominal metabolite replicates*** | **Adjusted metabolite replicates*** | **Nominal cardiac outcome replicates*** | **Adjusted cardiac outcome replicates*** | **Metabolite** |
| ENSG00000133789 | Q9UH65 | SWP70 | SWAP70 | Not yet druggable | Risk increasing | Decreasing | Risk decreasing | 1/2 | 1/2 | 1/2 | 0/2 | PC ae C42:2 |
| ENSG00000133789 | Q9UH65 | SWP70 | SWAP70 | Not yet druggable | Risk increasing | Decreasing | Risk decreasing | 1/2 | 0/2 | 1/2 | 0/2 | PC aa C38:5 |
| ENSG00000241186 | P13385 | TDGF1 | TDGF1 | Druggable | Risk decreasing | Decreasing | Risk increasing | 2/3 | 2/3 | 1/3 | 0/3 | PC aa C42:6 |
| ENSG00000241186 | P13385 | TDGF1 | TDGF1 | Druggable | Risk increasing | Increasing | Risk increasing | 0/3 | 0/3 | 1/3 | 0/3 | SM C16:0 |
| ENSG00000135605 | P42680 | TEC | TEC | Drugged | Risk increasing | Decreasing | Risk decreasing | - | - | - | - | SM C16:0 |
| ENSG00000168477 | P22105 | TENX | TNXB | Not yet druggable | Risk increasing | Increasing | Risk increasing | - | - | - | - | Tyrosine |
| ENSG00000107742 | Q92563 | TICN2 | SPOCK2 | Not yet druggable | Risk increasing | Decreasing | Risk decreasing | 1/2 | 1/2 | 0/2 | 0/2 | PC ae C42:3 |
| ENSG00000107742 | Q92563 | TICN2 | SPOCK2 | Not yet druggable | Risk increasing | Decreasing | Risk decreasing | 1/2 | 0/2 | 0/2 | 0/2 | Tetradecanoylcarnitine |
| ENSG00000118849 | P49788 | TIG1 | RARRES1 | Not yet druggable | Risk increasing | Increasing | Risk increasing | 0/2 | 0/2 | 1/2 | 0/2 | PC ae C38:4 |
| ENSG00000118849 | P49788 | TIG1 | RARRES1 | Not yet druggable | Risk increasing | Increasing | Risk increasing | 1/2 | 1/2 | 1/2 | 0/2 | SM C16:0 |
| ENSG00000118849 | P49788 | TIG1 | RARRES1 | Not yet druggable | Risk increasing | Increasing | Risk increasing | 0/2 | 0/2 | 1/2 | 0/2 | PC ae C38:5 |
| ENSG00000100234 | P35625 | TIMP3 | TIMP3 | Not yet druggable | Risk decreasing | Decreasing | Risk increasing | 1/1 | 1/1 | 0/1 | 0/1 | PC aa C42:6 |
| ENSG00000231925 | O15533 | TPSN | TAPBP | Not yet druggable | Risk increasing | Decreasing | Risk decreasing | - | - | - | - | PC aa C38:4 |
| ENSG00000231925 | O15533 | TPSN | TAPBP | Not yet druggable | Risk increasing | Decreasing | Risk decreasing | - | - | - | - | PC ae C42:3 |
| ENSG00000231925 | O15533 | TPSN | TAPBP | Not yet druggable | Risk increasing | Decreasing | Risk decreasing | - | - | - | - | PC ae C36:5 |
| ENSG00000231925 | O15533 | TPSN | TAPBP | Not yet druggable | Risk increasing | Decreasing | Risk decreasing | - | - | - | - | PC ae C38:4 |
| ENSG00000172236 | Q15661 | TRYB1 | TPSAB1 | Drugged | Risk increasing | Increasing | Risk increasing | - | - | - | - | Tetradecanoylcarnitine |
| ENSG00000169231 | P49746 | TSP3 | THBS3 | Not yet druggable | Risk increasing | Decreasing | Risk decreasing | - | - | - | - | PC aa C38:4 |
| ENSG00000169231 | P49746 | TSP3 | THBS3 | Not yet druggable | Risk increasing | Decreasing | Risk decreasing | - | - | - | - | PC ae C38:4 |
| ENSG00000160862 | P25311 | ZA2G | AZGP1 | Not yet druggable | Risk increasing | Decreasing | Risk decreasing | - | - | - | - | PC aa C40:4 |
| ENSG00000160862 | P25311 | ZA2G | AZGP1 | Not yet druggable | Risk increasing | Decreasing | Risk decreasing | - | - | - | - | PC aa C38:6 |

* Columns: Druggability - druggability per protein, where druggable is defined as a protein targeted by a compound that is being tested, and drugged is defined as a protein targeted by an approved compound (see Methods section), Metabolite-Outcome effect - whether higher metabolite values increase or decrease the risk of the specified outcome, Protei
Nominal metabolite replicates - number of times association of indicated protein and metabolite was replicated in other studies with a p-value < 0.05 / number of times association was performed (see Methods section), Adjusted metabolite replicates - number of times association of indicated protein and metabolite was replicated in other studies with an a
Nominal cardiac replicates - number of times association of indicated protein and cardiac outcome was replicated in other studies with a p-value < 0.05 / number of times association was performed (see Methods section), Adjusted cardiac replicates - number of times association of indicated protein and cardiac outcome was replicated in other studies with
Abbreviations: a = acyl residue, aa = diacyl residue, ae = acyl-alkyl residue, AF = atrial fibrillation, CI = confidence interval, DCM = dilated cardiomyopathy, HF = heart failure, LPC = lysophosphatidylcholine, MD = mean difference, NICM = non-ischemic cardiomyopathy, OR = odds ratio, PC = phosphatidylcholine, SM = sphingomyelin

**Table S7. Final results of all MRs per prioritised pro**

| Protein informatio | | | Metabolite effects on cardiac outcomes | | | | | Protein effects on metabolites | | | | | | Protein effects on cardiac outcomes | | | |
|---|---|---|---|---|---|---|---|---|---|---|---|---|---|---|---|---|---|
| Ensembl ID (GRCh38) | Uniprot ID | Protein name | Metabolite class | No. variants* | OR (95% CI) | p-value | Outcome | Gene name | No. variants* | MD (95% CI) | p-value | Metabolite | Gene name | No. variants* | OR (95% CI) | p-value | Outcome |
| ENSG00000133789 | Q9UH65 | SWP70 | Phosphatidylcholines | 34 | 1.09 (1.06; 1.12) | $2.9\times10^{-8}$ | HF | SWAP70 | 75 | -0.10 (-0.14; -0.06) | $4.2\times10^{-7}$ | PC ae C42:2 | SWAP70 | 65 | 0.96 (0.94; 0.97) | $4.0\times10^{-9}$ | HF |
| ENSG00000133789 | Q9UH65 | SWP70 | Phosphatidylcholines | 46 | 1.06 (1.03; 1.08) | $1.8\times10^{-6}$ | HF | SWAP70 | 77 | -0.06 (-0.08; -0.04) | $2.2\times10^{-7}$ | PC aa C38:5 | SWAP70 | 65 | 0.96 (0.94; 0.97) | $4.0\times10^{-9}$ | HF |
| ENSG00000241186 | P13385 | TDGF1 | Phosphatidylcholines | 14 | 0.76 (0.68; 0.85) | $2.7\times10^{-6}$ | AF | TDGF1 | 82 | -0.04 (-0.05; -0.03) | $5.7\times10^{-9}$ | PC aa C42:6 | TDGF1 | 87 | 1.03 (1.02; 1.04) | $6.3\times10^{-9}$ | AF |
| ENSG00000241186 | P13385 | TDGF1 | Sphingomyelins | 78 | 1.09 (1.05; 1.12) | $1.4\times10^{-7}$ | AF | TDGF1 | 87 | 0.03 (0.02; 0.04) | $1.2\times10^{-11}$ | SM C16:0 | TDGF1 | 87 | 1.03 (1.02; 1.04) | $6.3\times10^{-9}$ | AF |
| ENSG00000135605 | P42680 | TEC | Sphingomyelins | 78 | 1.09 (1.05; 1.12) | $1.4\times10^{-7}$ | AF | TEC | 23 | -0.21 (-0.26; -0.15) | $2.3\times10^{-12}$ | SM C16:0 | TEC | 25 | 0.87 (0.83; 0.90) | $5.6\times10^{-12}$ | AF |
| ENSG00000168477 | P22105 | TENX | Amino acids | 119 | 1.09 (1.05; 1.13) | $2.3\times10^{-6}$ | HF | TNXB | 31 | 0.03 (0.02; 0.05) | $5.4\times10^{-7}$ | Tyrosine | TNXB | 33 | 1.03 (1.02; 1.04) | $1.0\times10^{-8}$ | HF |
| ENSG00000107742 | Q92563 | TICN2 | Phosphatidylcholines | 46 | 1.04 (1.02; 1.06) | $5.6\times10^{-5}$ | AF | SPOCK2 | 32 | -0.20 (-0.27; -0.14) | $1.4\times10^{-9}$ | PC ae C42:3 | SPOCK2 | 34 | 0.91 (0.89; 0.94) | $1.2\times10^{-9}$ | AF |
| ENSG00000107742 | Q92563 | TICN2 | Acylcarnitines | 30 | 1.12 (1.06; 1.18) | $1.8\times10^{-5}$ | AF | SPOCK2 | 34 | -0.12 (-0.16; -0.07) | $1.6\times10^{-7}$ | Tetradecanoylcarnitine | SPOCK2 | 34 | 0.91 (0.89; 0.94) | $1.2\times10^{-9}$ | AF |
| ENSG00000118849 | P49788 | TIG1 | Phosphatidylcholines | 42 | 1.19 (1.10; 1.29) | $1.4\times10^{-5}$ | HF | RARRES1 | 56 | 0.03 (0.02; 0.04) | $7.4\times10^{-12}$ | PC ae C38:4 | RARRES1 | 49 | 1.03 (1.02; 1.03) | $1.0\times10^{-100}$ | HF |
| ENSG00000118849 | P49788 | TIG1 | Sphingomyelins | 67 | 1.10 (1.06; 1.14) | $3.3\times10^{-6}$ | HF | RARRES1 | 58 | 0.03 (0.02; 0.04) | $4.6\times10^{-14}$ | SM C16:0 | RARRES1 | 49 | 1.03 (1.02; 1.03) | $1.0\times10^{-100}$ | HF |
| ENSG00000118849 | P49788 | TIG1 | Phosphatidylcholines | 42 | 1.05 (1.03; 1.08) | $5.7\times10^{-5}$ | HF | RARRES1 | 54 | 0.04 (0.03; 0.05) | $1.0\times10^{-100}$ | PC ae C38:5 | RARRES1 | 49 | 1.03 (1.02; 1.03) | $1.0\times10^{-100}$ | HF |
| ENSG00000100234 | P35625 | TIMP3 | Phosphatidylcholines | 14 | 0.76 (0.68; 0.85) | $2.7\times10^{-6}$ | AF | TIMP3 | 88 | -0.06 (-0.08; -0.04) | $1.3\times10^{-7}$ | PC aa C42:6 | TIMP3 | 105 | 1.03 (1.02; 1.04) | $1.3\times10^{-11}$ | AF |
| ENSG00000231925 | O15533 | TPSN | Phosphatidylcholines | 66 | 1.05 (1.03; 1.06) | $2.5\times10^{-14}$ | AF | TAPBP | 28 | -0.06 (-0.09; -0.04) | $7.6\times10^{-8}$ | PC aa C38:4 | TAPBP | 28 | 0.96 (0.95; 0.98) | $1.4\times10^{-7}$ | AF |
| ENSG00000231925 | O15533 | TPSN | Phosphatidylcholines | 46 | 1.04 (1.02; 1.06) | $5.6\times10^{-5}$ | AF | TAPBP | 29 | -0.05 (-0.07; -0.04) | $4.3\times10^{-10}$ | PC ae C42:3 | TAPBP | 28 | 0.96 (0.95; 0.98) | $1.4\times10^{-7}$ | AF |
| ENSG00000231925 | O15533 | TPSN | Phosphatidylcholines | 50 | 1.04 (1.02; 1.06) | $1.4\times10^{-6}$ | AF | TAPBP | 29 | -0.05 (-0.06; -0.03) | $1.9\times10^{-8}$ | PC ae C36:5 | TAPBP | 28 | 0.96 (0.95; 0.98) | $1.4\times10^{-7}$ | AF |
| ENSG00000231925 | O15533 | TPSN | Phosphatidylcholines | 41 | 1.06 (1.04; 1.08) | $2.4\times10^{-7}$ | AF | TAPBP | 29 | -0.08 (-0.09; -0.06) | $1.0\times10^{-100}$ | PC ae C38:4 | TAPBP | 28 | 0.96 (0.95; 0.98) | $1.4\times10^{-7}$ | AF |
| ENSG00000172236 | Q15661 | TRYB1 | Acylcarnitines | 30 | 1.12 (1.06; 1.18) | $1.8\times10^{-5}$ | AF | TPSAB1 | 137 | 0.06 (0.04; 0.07) | $1.4\times10^{-13}$ | Tetradecanoylcarnitine | TPSAB1 | 161 | 1.02 (1.01; 1.03) | $1.4\times10^{-7}$ | AF |
| ENSG00000169231 | P49746 | TSP3 | Phosphatidylcholines | 66 | 1.05 (1.03; 1.06) | $2.5\times10^{-14}$ | AF | THBS3 | 13 | -0.42 (-0.58; -0.26) | $1.4\times10^{-7}$ | PC aa C38:4 | THBS3 | 10 | 0.55 (0.44; 0.69) | $3.7\times10^{-7}$ | AF |
| ENSG00000169231 | P49746 | TSP3 | Phosphatidylcholines | 41 | 1.06 (1.04; 1.08) | $2.4\times10^{-7}$ | AF | THBS3 | 13 | -0.42 (-0.58; -0.27) | $1.1\times10^{-7}$ | PC ae C38:4 | THBS3 | 10 | 0.55 (0.44; 0.69) | $3.7\times10^{-7}$ | AF |
| ENSG00000160862 | P25311 | ZA2G | Phosphatidylcholines | 24 | 1.12 (1.08; 1.15) | $1.1\times10^{-10}$ | HF | AZGP1 | 27 | -0.12 (-0.16; -0.08) | $1.3\times10^{-8}$ | PC aa C40:4 | AZGP1 | 26 | 0.92 (0.90; 0.95) | $2.4\times10^{-8}$ | HF |
| ENSG00000160862 | P25311 | ZA2G | Phosphatidylcholines | 22 | 1.14 (1.09; 1.20) | $7.0\times10^{-9}$ | HF | AZGP1 | 27 | -0.17 (-0.23; -0.12) | $7.9\times10^{-9}$ | PC aa C38:6 | AZGP1 | 26 | 0.92 (0.90; 0.95) | $2.4\times10^{-8}$ | HF |

* Columns: Druggability - druggability per protein, vn-Metabolite effect - whether higher protein values increase or decrease metabolite values, Protein-Outcome effect - whether higher protein values increase or decrease indicated outcome risk,
Nominal metabolite replicates - number of times asadjusted p-value / number of times association was performed (see Methods section),
Nominal cardiac replicates - number of times associan adjusted p-value / number of times association was performed (see Methods section), No. variants - number of variants used in MR-analysis
Abbreviations: a = acyl residue, aa = diacyl residue,
