## Supplemental Table S8 for "Integrating metabolomics and proteomics to identify novel drug targets for heart failure and atrial fibrillation"

**Table S8. Druggability results of the prioritised proteins**

| Target Uniprot ID* | Target ChEMBL ID | Target protein name | Druggability* | Target type | Drug ChEMBL ID | Drug name | Drug molecule type | Drug mechanism | Drug effect source | Drug effect type | Drug effect frequency | Max phase* | Drug effect | Drug effect class |
|---|---|---|---|---|---|---|---|---|---|---|---|---|---|---|
| P22303 | CHEMBL2095233 | ACES | Drugged | Selectivity group | CHEMBL636 | Rivastigmine | Small molecule | Inhibitor | BNF | Side-effect | Common or very common | 4 | Drowsiness | No |
| P22303 | CHEMBL220 | ACES | Drugged | Single protein | CHEMBL748 | Pralidoxime chloride | Small molecule | Activator | BNF | Side-effect | Not known | 4 | Hyperventilation | No |
| P22303 | CHEMBL220 | ACES | Drugged | Single protein | CHEMBL748 | Pralidoxime chloride | Small molecule | Activator | BNF | Side-effect | Not known | 4 | Headache | No |
| P22303 | CHEMBL220 | ACES | Drugged | Single protein | CHEMBL748 | Pralidoxime chloride | Small molecule | Activator | BNF | Side-effect | Not known | 4 | Drowsiness | No |
| P22303 | CHEMBL220 | ACES | Drugged | Single protein | CHEMBL748 | Pralidoxime chloride | Small molecule | Activator | BNF | Side-effect | Not known | 4 | Dizziness | No |
| P22303 | CHEMBL220 | ACES | Drugged | Single protein | CHEMBL748 | Pralidoxime chloride | Small molecule | Activator | BNF | Indication | All | 4 | Adjunct to atropine in the treatment of poisoning by organophosphorus insecticide or nerve agent | No |
| P22303 | CHEMBL220 | ACES | Drugged | Single protein | CHEMBL1678 | Donepezil hydrochloride | Small molecule | Inhibitor | BNF | Side-effect | Uncommon | 4 | Seizure | No |
| P22303 | CHEMBL220 | ACES | Drugged | Single protein | CHEMBL1678 | Donepezil hydrochloride | Small molecule | Inhibitor | BNF | Side-effect | Uncommon | 4 | Hypersalivation | No |
| P22303 | CHEMBL220 | ACES | Drugged | Single protein | CHEMBL1678 | Donepezil hydrochloride | Small molecule | Inhibitor | BNF | Side-effect | Uncommon | 4 | Gastrointestinal hemorrhage | No |
| P22303 | CHEMBL220 | ACES | Drugged | Single protein | CHEMBL1678 | Donepezil hydrochloride | Small molecule | Inhibitor | BNF | Side-effect | Uncommon | 4 | Bradycardia | Yes |
| P22303 | CHEMBL220 | ACES | Drugged | Single protein | CHEMBL1678 | Donepezil hydrochloride | Small molecule | Inhibitor | BNF | Side-effect | Rare or very rare | 4 | Rhabdomyolysis | No |
| P22303 | CHEMBL220 | ACES | Drugged | Single protein | CHEMBL1678 | Donepezil hydrochloride | Small molecule | Inhibitor | BNF | Side-effect | Rare or very rare | 4 | Neuroleptic malignant syndrome | No |
| P22303 | CHEMBL220 | ACES | Drugged | Single protein | CHEMBL1678 | Donepezil hydrochloride | Small molecule | Inhibitor | BNF | Side-effect | Rare or very rare | 4 | Hepatic disorder | No |
| P22303 | CHEMBL220 | ACES | Drugged | Single protein | CHEMBL748 | Pralidoxime chloride | Small molecule | Activator | BNF | Side-effect | Not known | 4 | Muscle weakness | No |
| P22303 | CHEMBL220 | ACES | Drugged | Single protein | CHEMBL1678 | Donepezil hydrochloride | Small molecule | Inhibitor | BNF | Side-effect | Rare or very rare | 4 | Extrapyramidal symptoms | No |
| P22303 | CHEMBL220 | ACES | Drugged | Single protein | CHEMBL1678 | Donepezil hydrochloride | Small molecule | Inhibitor | BNF | Side-effect | Common or very common | 4 | Vomiting | No |
| P22303 | CHEMBL220 | ACES | Drugged | Single protein | CHEMBL1678 | Donepezil hydrochloride | Small molecule | Inhibitor | BNF | Side-effect | Common or very common | 4 | Urinary incontinence | No |
| P22303 | CHEMBL220 | ACES | Drugged | Single protein | CHEMBL1678 | Donepezil hydrochloride | Small molecule | Inhibitor | BNF | Side-effect | Common or very common | 4 | Syncope | No |
| P22303 | CHEMBL220 | ACES | Drugged | Single protein | CHEMBL211471 | Neostigmine methylsulfate | Small molecule | Inhibitor | ChEMBL | Indication | All | 0,5 | Snoring | No |
| P22303 | CHEMBL220 | ACES | Drugged | Single protein | CHEMBL211471 | Neostigmine methylsulfate | Small molecule | Inhibitor | ChEMBL | Indication | All | 4 | Myasthenia gravis | No |
| P22303 | CHEMBL220 | ACES | Drugged | Single protein | CHEMBL1678 | Donepezil hydrochloride | Small molecule | Inhibitor | ChEMBL | Indication | All | 1 | Cognitive dysfunction | No |
| P22303 | CHEMBL220 | ACES | Drugged | Single protein | CHEMBL1678 | Donepezil hydrochloride | Small molecule | Inhibitor | ChEMBL | Indication | All | 2 | Autistic disorder | No |
| P22303 | CHEMBL220 | ACES | Drugged | Single protein | CHEMBL1678 | Donepezil hydrochloride | Small molecule | Inhibitor | ChEMBL | Indication | All | 2 | Migraine disorder | No |
| P22303 | CHEMBL220 | ACES | Drugged | Single protein | CHEMBL1678 | Donepezil hydrochloride | Small molecule | Inhibitor | ChEMBL | Indication | All | 2 | Neuralgia | No |
| P22303 | CHEMBL220 | ACES | Drugged | Single protein | CHEMBL1678 | Donepezil hydrochloride | Small molecule | Inhibitor | ChEMBL | Indication | All | 2 | Cancers | No |
| P22303 | CHEMBL220 | ACES | Drugged | Single protein | CHEMBL1678 | Donepezil hydrochloride | Small molecule | Inhibitor | ChEMBL | Indication | All | 3 | Delirium | No |
| P22303 | CHEMBL220 | ACES | Drugged | Single protein | CHEMBL1678 | Donepezil hydrochloride | Small molecule | Inhibitor | ChEMBL | Indication | All | 3 | Depressive disorder | No |
| P22303 | CHEMBL220 | ACES | Drugged | Single protein | CHEMBL1678 | Donepezil hydrochloride | Small molecule | Inhibitor | BNF | Side-effect | Rare or very rare | 4 | Cardiac conduction disorder | Yes |
| P22303 | CHEMBL220 | ACES | Drugged | Single protein | CHEMBL748 | Pralidoxime chloride | Small molecule | Activator | BNF | Side-effect | Not known | 4 | Nausea | No |
| P22303 | CHEMBL220 | ACES | Drugged | Single protein | CHEMBL748 | Pralidoxime chloride | Small molecule | Activator | BNF | Side-effect | Not known | 4 | Tachycardia | Yes |
| P22303 | CHEMBL220 | ACES | Drugged | Single protein | CHEMBL748 | Pralidoxime chloride | Small molecule | Activator | BNF | Side-effect | Not known | 4 | Vision disorder | No |
| P22303 | CHEMBL2095233 | ACES | Drugged | Selectivity group | CHEMBL636 | Rivastigmine | Small molecule | Inhibitor | ChEMBL | Indication | All | 3 | Cognitive dysfunction | No |
| P22303 | CHEMBL2095233 | ACES | Drugged | Selectivity group | CHEMBL636 | Rivastigmine | Small molecule | Inhibitor | ChEMBL | Indication | All | 3 | Neurobehavioral manifestations | No |
| P22303 | CHEMBL2095233 | ACES | Drugged | Selectivity group | CHEMBL636 | Rivastigmine | Small molecule | Inhibitor | ChEMBL | Indication | All | 3 | Dementia | No |
| P22303 | CHEMBL2095233 | ACES | Drugged | Selectivity group | CHEMBL636 | Rivastigmine | Small molecule | Inhibitor | ChEMBL | Indication | All | 3 | Supranuclear palsy, progressive | No |
| P22303 | CHEMBL2095233 | ACES | Drugged | Selectivity group | CHEMBL636 | Rivastigmine | Small molecule | Inhibitor | ChEMBL | Indication | All | 3 | Schizophrenia | No |
| P22303 | CHEMBL2095233 | ACES | Drugged | Selectivity group | CHEMBL636 | Rivastigmine | Small molecule | Inhibitor | ChEMBL | Indication | All | 3 | Depressive disorder | No |
| P22303 | CHEMBL2095233 | ACES | Drugged | Selectivity group | CHEMBL636 | Rivastigmine | Small molecule | Inhibitor | ChEMBL | Indication | All | 3 | Delirium | No |
| P22303 | CHEMBL2095233 | ACES | Drugged | Selectivity group | CHEMBL636 | Rivastigmine | Small molecule | Inhibitor | ChEMBL | Indication | All | 3 | Brain injuries | No |
| P22303 | CHEMBL2095233 | ACES | Drugged | Selectivity group | CHEMBL636 | Rivastigmine | Small molecule | Inhibitor | ChEMBL | Indication | All | 3 | Anxiety | No |
| P22303 | CHEMBL2095233 | ACES | Drugged | Selectivity group | CHEMBL636 | Rivastigmine | Small molecule | Inhibitor | ChEMBL | Indication | All | 1 | Cocaine-related disorder | No |
| P22303 | CHEMBL2095233 | ACES | Drugged | Selectivity group | CHEMBL636 | Rivastigmine | Small molecule | Inhibitor | ChEMBL | Indication | All | 1 | Amphetamine-related disorder | No |
| P22303 | CHEMBL2095233 | ACES | Drugged | Selectivity group | CHEMBL636 | Rivastigmine | Small molecule | Inhibitor | ChEMBL | Indication | All | 1 | Substance-related disorder | No |
| P22303 | CHEMBL2095233 | ACES | Drugged | Selectivity group | CHEMBL636 | Rivastigmine | Small molecule | Inhibitor | ChEMBL | Indication | All | 1 | Down syndrome | No |
| P22303 | CHEMBL2095233 | ACES | Drugged | Selectivity group | CHEMBL215645 | Rivastigmine tartrate | Small molecule | Inhibitor | ChEMBL | Indication | All | 4 | Parkinson disease | No |
| P22303 | CHEMBL2095233 | ACES | Drugged | Selectivity group | CHEMBL215645 | Rivastigmine tartrate | Small molecule | Inhibitor | ChEMBL | Indication | All | 4 | Dementia | No |
| P22303 | CHEMBL2095233 | ACES | Drugged | Selectivity group | CHEMBL215645 | Rivastigmine tartrate | Small molecule | Inhibitor | ChEMBL | Indication | All | 1 | Hypotension, orthostatic | No |
| P22303 | CHEMBL220 | ACES | Drugged | Single protein | CHEMBL812 | Pyridostigmine bromide | Small molecule | Inhibitor | BNF | Side-effect | Not known | 4 | Vomiting | No |
| P22303 | CHEMBL220 | ACES | Drugged | Single protein | CHEMBL812 | Pyridostigmine bromide | Small molecule | Inhibitor | BNF | Side-effect | Not known | 4 | Rash | No |
| P22303 | CHEMBL220 | ACES | Drugged | Single protein | CHEMBL812 | Pyridostigmine bromide | Small molecule | Inhibitor | BNF | Side-effect | Not known | 4 | Nausea | No |
| P22303 | CHEMBL220 | ACES | Drugged | Single protein | CHEMBL812 | Pyridostigmine bromide | Small molecule | Inhibitor | BNF | Side-effect | Not known | 4 | Muscle cramps | No |
| P22303 | CHEMBL220 | ACES | Drugged | Single protein | CHEMBL812 | Pyridostigmine bromide | Small molecule | Inhibitor | BNF | Side-effect | Not known | 4 | Hypersalivation | No |
| P22303 | CHEMBL220 | ACES | Drugged | Single protein | CHEMBL812 | Pyridostigmine bromide | Small molecule | Inhibitor | BNF | Side-effect | Not known | 4 | Excessive tearing | No |
| P22303 | CHEMBL220 | ACES | Drugged | Single protein | CHEMBL812 | Pyridostigmine bromide | Small molecule | Inhibitor | BNF | Side-effect | Not known | 4 | Diarrhoea | No |
| P22303 | CHEMBL220 | ACES | Drugged | Single protein | CHEMBL812 | Pyridostigmine bromide | Small molecule | Inhibitor | BNF | Side-effect | Not known | 4 | Gastrointestinal hypermotility | No |
| P22303 | CHEMBL220 | ACES | Drugged | Single protein | CHEMBL1678 | Donepezil hydrochloride | Small molecule | Inhibitor | BNF | Side-effect | Common or very common | 4 | Skin reactions | No |
| P22303 | CHEMBL220 | ACES | Drugged | Single protein | CHEMBL812 | Pyridostigmine bromide | Small molecule | Inhibitor | BNF | Side-effect | Not known | 4 | Abdominal cramps | No |
| P22303 | CHEMBL220 | ACES | Drugged | Single protein | CHEMBL812 | Pyridostigmine bromide | Small molecule | Inhibitor | BNF | Indication | All | 4 | Myasthenia gravis | No |
| P22303 | CHEMBL220 | ACES | Drugged | Single protein | CHEMBL1678 | Donepezil hydrochloride | Small molecule | Inhibitor | ChEMBL | Indication | All | 3 | Down syndrome | No |
| P22303 | CHEMBL2095233 | ACES | Drugged | Selectivity group | CHEMBL636 | Rivastigmine | Small molecule | Inhibitor | ChEMBL | Indication | All | 4 | Alzheimer disease | No |
| P22303 | CHEMBL220 | ACES | Drugged | Single protein | CHEMBL1678 | Donepezil hydrochloride | Small molecule | Inhibitor | ChEMBL | Indication | All | 3 | Dementia | No |
| P22303 | CHEMBL220 | ACES | Drugged | Single protein | CHEMBL1678 | Donepezil hydrochloride | Small molecule | Inhibitor | ChEMBL | Indication | All | 4 | Alzheimer disease | No |
| P22303 | CHEMBL220 | ACES | Drugged | Single protein | CHEMBL2107457 | Itopride | Small molecule | Inhibitor | ChEMBL | Indication | All | 1 | Gastroparesis | No |
| P22303 | CHEMBL220 | ACES | Drugged | Single protein | CHEMBL2107457 | Itopride | Small molecule | Inhibitor | ChEMBL | Indication | All | 2 | Constipation | No |
| P22303 | CHEMBL220 | ACES | Drugged | Single protein | CHEMBL2107457 | Itopride | Small molecule | Inhibitor | ChEMBL | Indication | All | 2 | Irritable bowel syndrome | No |
| P22303 | CHEMBL220 | ACES | Drugged | Single protein | CHEMBL2107457 | Itopride | Small molecule | Inhibitor | ChEMBL | Indication | All | 3 | Digestive system diseases | No |
| P22303 | CHEMBL220 | ACES | Drugged | Single protein | CHEMBL2107457 | Itopride | Small molecule | Inhibitor | ChEMBL | Indication | All | 3 | Dyspepsia | No |
| P22303 | CHEMBL220 | ACES | Drugged | Single protein | CHEMBL2107457 | Itopride | Small molecule | Inhibitor | ChEMBL | Indication | All | 3 | Peritonitis | No |
| P22303 | CHEMBL220 | ACES | Drugged | Single protein | CHEMBL1199307 | Distigmine | Small molecule | Inhibitor | ChEMBL | Indication | All | - | - | No |
| P22303 | CHEMBL220 | ACES | Drugged | Single protein | CHEMBL1255901 | Huperzine a | Small molecule | Inhibitor | ChEMBL | Indication | All | 1 | Substance-related disorder | No |
| P22303 | CHEMBL220 | ACES | Drugged | Single protein | CHEMBL1255901 | Huperzine a | Small molecule | Inhibitor | ChEMBL | Indication | All | 1 | Cocaine-related disorder | No |
| P22303 | CHEMBL220 | ACES | Drugged | Single protein | CHEMBL1255901 | Huperzine a | Small molecule | Inhibitor | ChEMBL | Indication | All | 2 | Alzheimer disease | No |
| P22303 | CHEMBL220 | ACES | Drugged | Single protein | CHEMBL1255901 | Huperzine a | Small molecule | Inhibitor | ChEMBL | Indication | All | 2 | Brain injuries | No |
| P22303 | CHEMBL220 | ACES | Drugged | Single protein | CHEMBL1678 | Donepezil hydrochloride | Small molecule | Inhibitor | BNF | Indication | All | 4 | Mild to moderate dementia in alzheimer's disease | No |
| P22303 | CHEMBL220 | ACES | Drugged | Single protein | CHEMBL338975 | Physostigmine salicylate | Small molecule | Inhibitor | ChEMBL | Indication | All | - | - | No |
| P22303 | CHEMBL220 | ACES | Drugged | Single protein | CHEMBL1678 | Donepezil hydrochloride | Small molecule | Inhibitor | BNF | Side-effect | Common or very common | 4 | Aggression | No |
| P22303 | CHEMBL220 | ACES | Drugged | Single protein | CHEMBL1678 | Donepezil hydrochloride | Small molecule | Inhibitor | BNF | Side-effect | Common or very common | 4 | Appetite decreased | No |
| P22303 | CHEMBL220 | ACES | Drugged | Single protein | CHEMBL1678 | Donepezil hydrochloride | Small molecule | Inhibitor | BNF | Side-effect | Common or very common | 4 | Common cold | No |
| P22303 | CHEMBL220 | ACES | Drugged | Single protein | CHEMBL1678 | Donepezil hydrochloride | Small molecule | Inhibitor | BNF | Side-effect | Common or very common | 4 | Diarrhoea | No |
| P22303 | CHEMBL220 | ACES | Drugged | Single protein | CHEMBL1678 | Donepezil hydrochloride | Small molecule | Inhibitor | BNF | Side-effect | Common or very common | 4 | Dizziness | No |
| P22303 | CHEMBL220 | ACES | Drugged | Single protein | CHEMBL1678 | Donepezil hydrochloride | Small molecule | Inhibitor | BNF | Side-effect | Common or very common | 4 | Fatigue | No |
| P22303 | CHEMBL220 | ACES | Drugged | Single protein | CHEMBL1678 | Donepezil hydrochloride | Small molecule | Inhibitor | BNF | Side-effect | Common or very common | 4 | Gastrointestinal disorder | No |
| P22303 | CHEMBL220 | ACES | Drugged | Single protein | CHEMBL1678 | Donepezil hydrochloride | Small molecule | Inhibitor | BNF | Side-effect | Common or very common | 4 | Hallucination | No |
| P22303 | CHEMBL220 | ACES | Drugged | Single protein | CHEMBL1678 | Donepezil hydrochloride | Small molecule | Inhibitor | BNF | Side-effect | Common or very common | 4 | Headache | No |
| P22303 | CHEMBL220 | ACES | Drugged | Single protein | CHEMBL1678 | Donepezil hydrochloride | Small molecule | Inhibitor | BNF | Side-effect | Common or very common | 4 | Injury | No |
| P22303 | CHEMBL220 | ACES | Drugged | Single protein | CHEMBL1678 | Donepezil hydrochloride | Small molecule | Inhibitor | BNF | Side-effect | Common or very common | 4 | Muscle cramps | No |
| P22303 | CHEMBL220 | ACES | Drugged | Single protein | CHEMBL1678 | Donepezil hydrochloride | Small molecule | Inhibitor | BNF | Side-effect | Common or very common | 4 | Nausea | No |

| Table S8. Druggability results of the prioritised proteins | | | | | | | | | | | | | | |
|---|---|---|---|---|---|---|---|---|---|---|---|---|---|---|
| Target Uniprot ID* | Target ChEMBL ID | Target protein name | Druggability* | Target type | Drug ChEMBL ID | Drug name | Drug molecule type | Drug mechanism | Drug effect source | Drug effect type | Drug effect frequency | Max phase* | Drug effect | Drug effect class |
| P22303 | CHEMBL220 | ACES | Drugged | Single protein | CHEMBL1678 | Donepezil hydrochloride | Small molecule | Inhibitor | BNF | Side-effect | Common or very common | 4 | Pain | No |
| P22303 | CHEMBL220 | ACES | Drugged | Single protein | CHEMBL1678 | Donepezil hydrochloride | Small molecule | Inhibitor | BNF | Side-effect | Common or very common | 4 | Agitation | No |
| P22303 | CHEMBL220 | ACES | Drugged | Single protein | CHEMBL2107308 | Acotiamide hydrochloride | Small molecule | Inhibitor | ChEMBL | Indication | All | 3 | Dyspepsia | No |
| P22303 | CHEMBL220 | ACES | Drugged | Single protein | CHEMBL278819 | Minaprine | Small molecule | Inhibitor | ChEMBL | Indication | All | 4 | Depressive disorder | No |
| P22303 | CHEMBL220 | ACES | Drugged | Single protein | CHEMBL1364551 | Minaprine hydrochloride | Small molecule | Inhibitor | ChEMBL | Indication | All | - | - | No |
| P22303 | CHEMBL220 | ACES | Drugged | Single protein | CHEMBL1678 | Donepezil hydrochloride | Small molecule | Inhibitor | ChEMBL | Indication | All | 4 | Dementia | No |
| P22303 | CHEMBL220 | ACES | Drugged | Single protein | CHEMBL1200933 | Hexafluorenium bromide | Small molecule | Inhibitor | ChEMBL | Indication | All | - | - | No |
| P22303 | CHEMBL220 | ACES | Drugged | Single protein | CHEMBL812 | Pyridostigmine bromide | Small molecule | Inhibitor | ChEMBL | Indication | All | 0,5 | Glycogen storage disease type ii | No |
| P22303 | CHEMBL220 | ACES | Drugged | Single protein | CHEMBL812 | Pyridostigmine bromide | Small molecule | Inhibitor | ChEMBL | Indication | All | 1 | Hypotension, orthostatic | No |
| P22303 | CHEMBL220 | ACES | Drugged | Single protein | CHEMBL812 | Pyridostigmine bromide | Small molecule | Inhibitor | ChEMBL | Indication | All | 2 | Heart failure | Yes |
| P22303 | CHEMBL220 | ACES | Drugged | Single protein | CHEMBL812 | Pyridostigmine bromide | Small molecule | Inhibitor | ChEMBL | Indication | All | 2 | Hypotension | No |
| P22303 | CHEMBL220 | ACES | Drugged | Single protein | CHEMBL812 | Pyridostigmine bromide | Small molecule | Inhibitor | ChEMBL | Indication | All | 2 | Parkinson disease | No |
| P22303 | CHEMBL220 | ACES | Drugged | Single protein | CHEMBL812 | Pyridostigmine bromide | Small molecule | Inhibitor | ChEMBL | Indication | All | 2 | Spinal muscular atrophies of childhood | No |
| P22303 | CHEMBL220 | ACES | Drugged | Single protein | CHEMBL812 | Pyridostigmine bromide | Small molecule | Inhibitor | ChEMBL | Indication | All | 2 | Severe acute respiratory syndrome | No |
| P22303 | CHEMBL220 | ACES | Drugged | Single protein | CHEMBL812 | Pyridostigmine bromide | Small molecule | Inhibitor | ChEMBL | Indication | All | 2 | Ileus | No |
| P22303 | CHEMBL220 | ACES | Drugged | Single protein | CHEMBL812 | Pyridostigmine bromide | Small molecule | Inhibitor | ChEMBL | Indication | All | 4 | Myasthenia gravis | No |
| P22303 | CHEMBL220 | ACES | Drugged | Single protein | CHEMBL1555 | Galantamine hydrobromide | Small molecule | Inhibitor | ChEMBL | Indication | All | 2 | Frontotemporal dementia | No |
| P22303 | CHEMBL220 | ACES | Drugged | Single protein | CHEMBL1555 | Galantamine hydrobromide | Small molecule | Inhibitor | ChEMBL | Indication | All | 3 | Mental disorder | No |
| P22303 | CHEMBL220 | ACES | Drugged | Single protein | CHEMBL1555 | Galantamine hydrobromide | Small molecule | Inhibitor | ChEMBL | Indication | All | 3 | Schizophrenia | No |
| P22303 | CHEMBL220 | ACES | Drugged | Single protein | CHEMBL1555 | Galantamine hydrobromide | Small molecule | Inhibitor | ChEMBL | Indication | All | 3 | Dementia | No |
| P22303 | CHEMBL220 | ACES | Drugged | Single protein | CHEMBL1555 | Galantamine hydrobromide | Small molecule | Inhibitor | ChEMBL | Indication | All | 3 | Cognitive dysfunction | No |
| P22303 | CHEMBL220 | ACES | Drugged | Single protein | CHEMBL1555 | Galantamine hydrobromide | Small molecule | Inhibitor | ChEMBL | Indication | All | 4 | Alzheimer disease | No |
| P22303 | CHEMBL220 | ACES | Drugged | Single protein | CHEMBL1555 | Galantamine hydrobromide | Small molecule | Inhibitor | ChEMBL | Indication | All | 4 | Dementia | No |
| P22303 | CHEMBL220 | ACES | Drugged | Single protein | CHEMBL1025 | Isoflurophate | Small molecule | Inhibitor | ChEMBL | Indication | All | 4 | Cancers | No |
| P22303 | CHEMBL220 | ACES | Drugged | Single protein | CHEMBL1128 | Edrophonium chloride | Small molecule | Inhibitor | ChEMBL | Indication | All | - | - | No |
| P22303 | CHEMBL220 | ACES | Drugged | Single protein | CHEMBL1200367 | Echothiophate iodide | Small molecule | Inhibitor | ChEMBL | Indication | All | - | - | No |
| P22303 | CHEMBL220 | ACES | Drugged | Single protein | CHEMBL1200514 | Demecarium bromide | Small molecule | Inhibitor | ChEMBL | Indication | All | - | - | No |
| P22303 | CHEMBL220 | ACES | Drugged | Single protein | CHEMBL1200541 | Ambenonium chloride | Small molecule | Inhibitor | ChEMBL | Indication | All | - | - | No |
| P22303 | CHEMBL220 | ACES | Drugged | Single protein | CHEMBL748 | Pralidoxime chloride | Small molecule | Activator | ChEMBL | Indication | All | 4 | Myasthenia gravis | No |
| P22303 | CHEMBL220 | ACES | Drugged | Single protein | CHEMBL748 | Pralidoxime chloride | Small molecule | Activator | ChEMBL | Indication | All | 4 | Poisoning | No |
| P22303 | CHEMBL220 | ACES | Drugged | Single protein | CHEMBL1201341 | Echothiophate | Small molecule | Inhibitor | ChEMBL | Indication | All | 4 | Cancers | No |
| P22303 | CHEMBL220 | ACES | Drugged | Single protein | CHEMBL2105345 | Propanidid | Small molecule | Inhibitor | ChEMBL | Indication | All | - | - | No |
| P22303 | CHEMBL220 | ACES | Drugged | Single protein | CHEMBL1678 | Donepezil hydrochloride | Small molecule | Inhibitor | ChEMBL | Indication | All | 3 | Rett syndrome | No |
| P22303 | CHEMBL2095233 | ACES | Drugged | Selectivity group | CHEMBL636 | Rivastigmine | Small molecule | Inhibitor | ChEMBL | Indication | All | 4 | Dementia | No |
| P22303 | CHEMBL220 | ACES | Drugged | Single protein | CHEMBL1678 | Donepezil hydrochloride | Small molecule | Inhibitor | BNF | Side-effect | Common or very common | 4 | Sleep disorder | No |
| P22303 | CHEMBL2095233 | ACES | Drugged | Selectivity group | CHEMBL1677 | Tacrine hydrochloride | Small molecule | Inhibitor | ChEMBL | Indication | All | - | - | No |
| P22303 | CHEMBL2095233 | ACES | Drugged | Selectivity group | CHEMBL636 | Rivastigmine | Small molecule | Inhibitor | BNF | Side-effect | Common or very common | 4 | Skin reactions | No |
| P22303 | CHEMBL2095233 | ACES | Drugged | Selectivity group | CHEMBL636 | Rivastigmine | Small molecule | Inhibitor | BNF | Side-effect | Uncommon | 4 | Hypotension | No |
| P22303 | CHEMBL2095233 | ACES | Drugged | Selectivity group | CHEMBL636 | Rivastigmine | Small molecule | Inhibitor | BNF | Side-effect | Common or very common | 4 | Sleep disorder | No |
| P22303 | CHEMBL2095233 | ACES | Drugged | Selectivity group | CHEMBL636 | Rivastigmine | Small molecule | Inhibitor | BNF | Side-effect | Uncommon | 4 | Gastric ulcer | No |
| P22303 | CHEMBL2095233 | ACES | Drugged | Selectivity group | CHEMBL636 | Rivastigmine | Small molecule | Inhibitor | BNF | Side-effect | Uncommon | 4 | Atrioventricular block | Yes |
| P22303 | CHEMBL2095233 | ACES | Drugged | Selectivity group | CHEMBL636 | Rivastigmine | Small molecule | Inhibitor | BNF | Side-effect | Uncommon | 4 | Aggression | No |
| P22303 | CHEMBL2095233 | ACES | Drugged | Selectivity group | CHEMBL636 | Rivastigmine | Small molecule | Inhibitor | BNF | Side-effect | Rare or very rare | 4 | Seizure | No |
| P22303 | CHEMBL2095233 | ACES | Drugged | Selectivity group | CHEMBL636 | Rivastigmine | Small molecule | Inhibitor | BNF | Side-effect | Rare or very rare | 4 | Gastrointestinal hemorrhage | No |
| P22303 | CHEMBL2095233 | ACES | Drugged | Selectivity group | CHEMBL636 | Rivastigmine | Small molecule | Inhibitor | BNF | Side-effect | Rare or very rare | 4 | Gastrointestinal disorder | No |
| P22303 | CHEMBL2095233 | ACES | Drugged | Selectivity group | CHEMBL636 | Rivastigmine | Small molecule | Inhibitor | BNF | Side-effect | Rare or very rare | 4 | Angina | Yes |
| P22303 | CHEMBL2095233 | ACES | Drugged | Selectivity group | CHEMBL636 | Rivastigmine | Small molecule | Inhibitor | BNF | Side-effect | Rare or very rare | 4 | Pancreatitis | No |
| P22303 | CHEMBL2095233 | ACES | Drugged | Selectivity group | CHEMBL636 | Rivastigmine | Small molecule | Inhibitor | BNF | Side-effect | Not known | 4 | Nightmare | No |
| P22303 | CHEMBL2095233 | ACES | Drugged | Selectivity group | CHEMBL636 | Rivastigmine | Small molecule | Inhibitor | BNF | Side-effect | Not known | 4 | Hallucination | No |
| P22303 | CHEMBL2095233 | ACES | Drugged | Selectivity group | CHEMBL636 | Rivastigmine | Small molecule | Inhibitor | BNF | Side-effect | Not known | 4 | Hepatitis | No |
| P22303 | CHEMBL2095233 | ACES | Drugged | Selectivity group | CHEMBL636 | Rivastigmine | Small molecule | Inhibitor | ChEMBL | Indication | All | 4 | Parkinson disease | No |
| P22303 | CHEMBL2095233 | ACES | Drugged | Selectivity group | CHEMBL636 | Rivastigmine | Small molecule | Inhibitor | BNF | Side-effect | Common or very common | 4 | Weight decreased | No |
| P22303 | CHEMBL2095233 | ACES | Drugged | Selectivity group | CHEMBL636 | Rivastigmine | Small molecule | Inhibitor | BNF | Side-effect | Common or very common | 4 | Vomiting | No |
| P22303 | CHEMBL2095233 | ACES | Drugged | Selectivity group | CHEMBL636 | Rivastigmine | Small molecule | Inhibitor | BNF | Side-effect | Common or very common | 4 | Urinary tract infection | No |
| P22303 | CHEMBL2095233 | ACES | Drugged | Selectivity group | CHEMBL636 | Rivastigmine | Small molecule | Inhibitor | BNF | Side-effect | Common or very common | 4 | Urinary incontinence | No |
| P22303 | CHEMBL2095233 | ACES | Drugged | Selectivity group | CHEMBL636 | Rivastigmine | Small molecule | Inhibitor | BNF | Side-effect | Common or very common | 4 | Parkinsonism | No |
| P22303 | CHEMBL2095233 | ACES | Drugged | Selectivity group | CHEMBL636 | Rivastigmine | Small molecule | Inhibitor | BNF | Side-effect | Common or very common | 4 | Tremor | No |
| P22303 | CHEMBL2095233 | ACES | Drugged | Selectivity group | CHEMBL636 | Rivastigmine | Small molecule | Inhibitor | BNF | Side-effect | Common or very common | 4 | Nausea | No |
| P22303 | CHEMBL2095233 | ACES | Drugged | Selectivity group | CHEMBL636 | Rivastigmine | Small molecule | Inhibitor | BNF | Side-effect | Common or very common | 4 | Malaise | No |
| P22303 | CHEMBL2095233 | ACES | Drugged | Selectivity group | CHEMBL636 | Rivastigmine | Small molecule | Inhibitor | BNF | Indication | All | 4 | Mild to moderate dementia in alzheimer's disease | No |
| P22303 | CHEMBL2095233 | ACES | Drugged | Selectivity group | CHEMBL636 | Rivastigmine | Small molecule | Inhibitor | BNF | Indication | All | 4 | Mild to moderate dementia in parkinson's disease | No |
| P22303 | CHEMBL2095233 | ACES | Drugged | Selectivity group | CHEMBL636 | Rivastigmine | Small molecule | Inhibitor | BNF | Side-effect | Common or very common | 4 | Anxiety | No |
| P22303 | CHEMBL2095233 | ACES | Drugged | Selectivity group | CHEMBL636 | Rivastigmine | Small molecule | Inhibitor | BNF | Side-effect | Common or very common | 4 | Appetite decreased | No |
| P22303 | CHEMBL2095233 | ACES | Drugged | Selectivity group | CHEMBL636 | Rivastigmine | Small molecule | Inhibitor | BNF | Side-effect | Common or very common | 4 | Arrhythmias | Yes |
| P22303 | CHEMBL2095233 | ACES | Drugged | Selectivity group | CHEMBL636 | Rivastigmine | Small molecule | Inhibitor | BNF | Side-effect | Common or very common | 4 | Asthenia | No |
| P22303 | CHEMBL2095233 | ACES | Drugged | Selectivity group | CHEMBL636 | Rivastigmine | Small molecule | Inhibitor | BNF | Side-effect | Common or very common | 4 | Confusion | No |
| P22303 | CHEMBL2095233 | ACES | Drugged | Selectivity group | CHEMBL636 | Rivastigmine | Small molecule | Inhibitor | BNF | Side-effect | Common or very common | 4 | Dehydration | No |
| P22303 | CHEMBL2095233 | ACES | Drugged | Selectivity group | CHEMBL636 | Rivastigmine | Small molecule | Inhibitor | BNF | Side-effect | Common or very common | 4 | Depression | No |
| P22303 | CHEMBL2095233 | ACES | Drugged | Selectivity group | CHEMBL636 | Rivastigmine | Small molecule | Inhibitor | BNF | Side-effect | Common or very common | 4 | Diarrhoea | No |
| P22303 | CHEMBL2095233 | ACES | Drugged | Selectivity group | CHEMBL636 | Rivastigmine | Small molecule | Inhibitor | BNF | Side-effect | Common or very common | 4 | Movement disorder | No |
| P22303 | CHEMBL2095233 | ACES | Drugged | Selectivity group | CHEMBL636 | Rivastigmine | Small molecule | Inhibitor | BNF | Side-effect | Common or very common | 4 | Dizziness | No |
| P22303 | CHEMBL2095233 | ACES | Drugged | Selectivity group | CHEMBL636 | Rivastigmine | Small molecule | Inhibitor | BNF | Side-effect | Common or very common | 4 | Fall | No |
| P22303 | CHEMBL2095233 | ACES | Drugged | Selectivity group | CHEMBL636 | Rivastigmine | Small molecule | Inhibitor | BNF | Side-effect | Common or very common | 4 | Gait abnormal | No |
| P22303 | CHEMBL2095233 | ACES | Drugged | Selectivity group | CHEMBL636 | Rivastigmine | Small molecule | Inhibitor | BNF | Side-effect | Common or very common | 4 | Gastrointestinal discomfort | No |
| P22303 | CHEMBL2095233 | ACES | Drugged | Selectivity group | CHEMBL636 | Rivastigmine | Small molecule | Inhibitor | BNF | Side-effect | Common or very common | 4 | Hallucination | No |
| P22303 | CHEMBL2095233 | ACES | Drugged | Selectivity group | CHEMBL636 | Rivastigmine | Small molecule | Inhibitor | BNF | Side-effect | Common or very common | 4 | Headache | No |
| P22303 | CHEMBL2095233 | ACES | Drugged | Selectivity group | CHEMBL636 | Rivastigmine | Small molecule | Inhibitor | BNF | Side-effect | Common or very common | 4 | Hyperhidrosis | No |
| P22303 | CHEMBL2095233 | ACES | Drugged | Selectivity group | CHEMBL636 | Rivastigmine | Small molecule | Inhibitor | BNF | Side-effect | Common or very common | 4 | Hypersalivation | No |
| P22303 | CHEMBL2095233 | ACES | Drugged | Selectivity group | CHEMBL636 | Rivastigmine | Small molecule | Inhibitor | BNF | Side-effect | Common or very common | 4 | Hypertension | Yes |
| P22303 | CHEMBL2095233 | ACES | Drugged | Selectivity group | CHEMBL636 | Rivastigmine | Small molecule | Inhibitor | BNF | Side-effect | Common or very common | 4 | Syncope | No |
| P14621 | - | ACYP2 | Not yet druggable | - | - | - | - | - | - | - | - | - | - | No |
| P08319 | CHEMBL2096668 | ADH4 | Druggable | Protein family | CHEMBL1909285 | Nitrefazole | Small molecule | Inhibitor | ChEMBL | Indication | All | - | - | No |
| P08319 | CHEMBL2990 | ADH4 | Druggable | Single protein | - | - | - | - | - | - | - | - | - | No |
| P50995 | - | ANX11 | Not yet druggable | - | - | - | - | - | - | - | - | - | - | No |
| P02654 | - | APOC1 | Not yet druggable | - | - | - | - | - | - | - | - | - | - | No |
| P02656 | CHEMBL4523160 | APOC3 | Druggable | Single protein | - | - | - | - | - | - | - | - | - | No |
| P14415 | CHEMBL2095186 | AT1B2 | Drugged | Protein complex group | CHEMBL1751 | Digoxin | Small molecule | Inhibitor | ChEMBL | Indication | All | 2 | Cancers | No |

**Table S8. Druggability results of the prioritised proteins**

| Target Uniprot ID* | Target ChEMBL ID | Target protein name | Druggability* | Target type | Drug ChEMBL ID | Drug name | Drug molecule type | Drug mechanism | Drug effect source | Drug effect type | Drug effect frequency | Max phase* | Drug effect | Drug effect class |
|---|---|---|---|---|---|---|---|---|---|---|---|---|---|---|
| P14415 | CHEMBL2095186 | AT1B2 | Drugged | Protein complex group | CHEMBL1751 | Digoxin | Small molecule | Inhibitor | BNF | Side-effect | Rare or very rare | 4 | Malaise | No |
| P14415 | CHEMBL2095186 | AT1B2 | Drugged | Protein complex group | CHEMBL1751 | Digoxin | Small molecule | Inhibitor | BNF | Side-effect | Rare or very rare | 4 | Psychosis | No |
| P14415 | CHEMBL2095186 | AT1B2 | Drugged | Protein complex group | CHEMBL1751 | Digoxin | Small molecule | Inhibitor | BNF | Side-effect | Rare or very rare | 4 | Thrombocytopenia | No |
| P14415 | CHEMBL2095186 | AT1B2 | Drugged | Protein complex group | CHEMBL1751 | Digoxin | Small molecule | Inhibitor | BNF | Side-effect | Uncommon | 4 | Depression | No |
| P14415 | CHEMBL2095186 | AT1B2 | Drugged | Protein complex group | CHEMBL1751 | Digoxin | Small molecule | Inhibitor | ChEMBL | Indication | All | 1 | Angiodema | No |
| P14415 | CHEMBL2095186 | AT1B2 | Drugged | Protein complex group | CHEMBL1751 | Digoxin | Small molecule | Inhibitor | ChEMBL | Indication | All | 1 | Substance-related disorder | No |
| P14415 | CHEMBL2095186 | AT1B2 | Drugged | Protein complex group | CHEMBL1751 | Digoxin | Small molecule | Inhibitor | ChEMBL | Indication | All | 1 | Hepatitis b, chronic | No |
| P14415 | CHEMBL2095186 | AT1B2 | Drugged | Protein complex group | CHEMBL1751 | Digoxin | Small molecule | Inhibitor | ChEMBL | Indication | All | 1 | Hiv infection | No |
| P14415 | CHEMBL2095186 | AT1B2 | Drugged | Protein complex group | CHEMBL1751 | Digoxin | Small molecule | Inhibitor | ChEMBL | Indication | All | 1 | Nausea | No |
| P14415 | CHEMBL2095186 | AT1B2 | Drugged | Protein complex group | CHEMBL1751 | Digoxin | Small molecule | Inhibitor | ChEMBL | Indication | All | 1 | Influenza, human | No |
| P14415 | CHEMBL2095186 | AT1B2 | Drugged | Protein complex group | CHEMBL1751 | Digoxin | Small molecule | Inhibitor | ChEMBL | Indication | All | 1 | Inflammation | No |
| P14415 | CHEMBL2095186 | AT1B2 | Drugged | Protein complex group | CHEMBL1751 | Digoxin | Small molecule | Inhibitor | ChEMBL | Indication | All | 1 | Infections | No |
| P14415 | CHEMBL2095186 | AT1B2 | Drugged | Protein complex group | CHEMBL1751 | Digoxin | Small molecule | Inhibitor | ChEMBL | Indication | All | 1 | Von hippel-lindau disease | No |
| P14415 | CHEMBL2095186 | AT1B2 | Drugged | Protein complex group | CHEMBL1751 | Digoxin | Small molecule | Inhibitor | ChEMBL | Indication | All | 1 | Hepatitis c | No |
| P14415 | CHEMBL2095186 | AT1B2 | Drugged | Protein complex group | CHEMBL1751 | Digoxin | Small molecule | Inhibitor | ChEMBL | Indication | All | 1 | Cancers | No |
| P14415 | CHEMBL2095186 | AT1B2 | Drugged | Protein complex group | CHEMBL1751 | Digoxin | Small molecule | Inhibitor | ChEMBL | Indication | All | 1 | Epilepsy | No |
| P14415 | CHEMBL2095186 | AT1B2 | Drugged | Protein complex group | CHEMBL1751 | Digoxin | Small molecule | Inhibitor | ChEMBL | Indication | All | 1 | Diabetes mellitus, type 2 | No |
| P14415 | CHEMBL2095186 | AT1B2 | Drugged | Protein complex group | CHEMBL1751 | Digoxin | Small molecule | Inhibitor | ChEMBL | Indication | All | 1 | Diabetes mellitus | No |
| P14415 | CHEMBL2095186 | AT1B2 | Drugged | Protein complex group | CHEMBL1751 | Digoxin | Small molecule | Inhibitor | ChEMBL | Indication | All | 1 | Depressive disorder | No |
| P14415 | CHEMBL2095186 | AT1B2 | Drugged | Protein complex group | CHEMBL1751 | Digoxin | Small molecule | Inhibitor | ChEMBL | Indication | All | 1 | Cytomegalovirus infection | No |
| P14415 | CHEMBL2095186 | AT1B2 | Drugged | Protein complex group | CHEMBL1751 | Digoxin | Small molecule | Inhibitor | ChEMBL | Indication | All | 1 | Hypertension | Yes |
| P14415 | CHEMBL2095186 | AT1B2 | Drugged | Protein complex group | CHEMBL254219 | Digitoxin | Small molecule | Inhibitor | ChEMBL | Indication | All | 4 | Cardiovascular diseases | Yes |
| P14415 | CHEMBL2095186 | AT1B2 | Drugged | Protein complex group | CHEMBL1614 | Deslanoside | Small molecule | Inhibitor | ChEMBL | Indication | All | 4 | Cardiovascular diseases | Yes |
| P14415 | CHEMBL2095186 | AT1B2 | Drugged | Protein complex group | CHEMBL1751 | Digoxin | Small molecule | Inhibitor | ChEMBL | Indication | All | 2 | Arthritis | No |
| P14415 | CHEMBL2095186 | AT1B2 | Drugged | Protein complex group | CHEMBL1751 | Digoxin | Small molecule | Inhibitor | BNF | Side-effect | Rare or very rare | 4 | Headache | No |
| P14415 | CHEMBL2095186 | AT1B2 | Drugged | Protein complex group | CHEMBL1751 | Digoxin | Small molecule | Inhibitor | BNF | Side-effect | Rare or very rare | 4 | Gynaecomastia | No |
| P14415 | CHEMBL2095186 | AT1B2 | Drugged | Protein complex group | CHEMBL254219 | Digitoxin | Small molecule | Inhibitor | ChEMBL | Indication | All | 2 | Cystic fibrosis | No |
| P14415 | CHEMBL2095186 | AT1B2 | Drugged | Protein complex group | CHEMBL1751 | Digoxin | Small molecule | Inhibitor | BNF | Side-effect | Rare or very rare | 4 | Confusion | No |
| P14415 | CHEMBL2095186 | AT1B2 | Drugged | Protein complex group | CHEMBL1751 | Digoxin | Small molecule | Inhibitor | ChEMBL | Indication | All | 2 | Keratosis, actinic | No |
| P14415 | CHEMBL2095186 | AT1B2 | Drugged | Protein complex group | CHEMBL1751 | Digoxin | Small molecule | Inhibitor | BNF | Side-effect | Rare or very rare | 4 | Gastrointestinal disorder | No |
| P14415 | CHEMBL2095186 | AT1B2 | Drugged | Protein complex group | CHEMBL1751 | Digoxin | Small molecule | Inhibitor | ChEMBL | Indication | All | 2 | Non-alcoholic fatty liver disease | No |
| P14415 | CHEMBL2095186 | AT1B2 | Drugged | Protein complex group | CHEMBL1751 | Digoxin | Small molecule | Inhibitor | ChEMBL | Indication | All | 3 | Tachycardia | Yes |
| P14415 | CHEMBL2095186 | AT1B2 | Drugged | Protein complex group | CHEMBL1751 | Digoxin | Small molecule | Inhibitor | ChEMBL | Indication | All | 4 | Atrial fibrillation | Yes |
| P14415 | CHEMBL2095186 | AT1B2 | Drugged | Protein complex group | CHEMBL1751 | Digoxin | Small molecule | Inhibitor | ChEMBL | Indication | All | 4 | Heart failure | Yes |
| P14415 | CHEMBL2095186 | AT1B2 | Drugged | Protein complex group | CHEMBL3545057 | Acetyldigitoxin | Small molecule | Inhibitor | ChEMBL | Indication | All | 4 | Cardiovascular diseases | Yes |
| P14415 | CHEMBL2095186 | AT1B2 | Drugged | Protein complex group | CHEMBL2093999 | Istaroxime | Small molecule | Inhibitor | ChEMBL | Indication | All | 2 | Heart failure | Yes |
| P14415 | CHEMBL2095186 | AT1B2 | Drugged | Protein complex group | CHEMBL2093999 | Istaroxime | Small molecule | Inhibitor | ChEMBL | Indication | All | 2 | Shock | No |
| P14415 | CHEMBL2095186 | AT1B2 | Drugged | Protein complex group | CHEMBL506569 | Lanatoside c | Small molecule | Inhibitor | ChEMBL | Indication | All | 4 | Cardiovascular diseases | Yes |
| P14415 | CHEMBL2095186 | AT1B2 | Drugged | Protein complex group | CHEMBL1751 | Digoxin | Small molecule | Inhibitor | BNF | Indication | All | 4 | Emergency loading dose, for atrial fibrillation or flutter | Yes |
| P14415 | CHEMBL2095186 | AT1B2 | Drugged | Protein complex group | CHEMBL1751 | Digoxin | Small molecule | Inhibitor | BNF | Indication | All | 4 | Heart failure | No |
| P14415 | CHEMBL2095186 | AT1B2 | Drugged | Protein complex group | CHEMBL1751 | Digoxin | Small molecule | Inhibitor | BNF | Indication | All | 4 | Maintenance, for atrial fibrillation or flutter | No |
| P14415 | CHEMBL2095186 | AT1B2 | Drugged | Protein complex group | CHEMBL1751 | Digoxin | Small molecule | Inhibitor | ChEMBL | Indication | All | 4 | Cardiovascular diseases | Yes |
| P14415 | CHEMBL2095186 | AT1B2 | Drugged | Protein complex group | CHEMBL1751 | Digoxin | Small molecule | Inhibitor | BNF | Side-effect | Common or very common | 4 | Arrhythmias | Yes |
| P14415 | CHEMBL2095186 | AT1B2 | Drugged | Protein complex group | CHEMBL1751 | Digoxin | Small molecule | Inhibitor | BNF | Side-effect | Rare or very rare | 4 | Appetite decreased | No |
| P14415 | CHEMBL2095186 | AT1B2 | Drugged | Protein complex group | CHEMBL1751 | Digoxin | Small molecule | Inhibitor | BNF | Indication | All | 4 | Rapid digitalisation, for atrial fibrillation or flutter | Yes |
| P14415 | CHEMBL2095186 | AT1B2 | Drugged | Protein complex group | CHEMBL1751 | Digoxin | Small molecule | Inhibitor | BNF | Side-effect | Rare or very rare | 4 | Asthenia | No |
| P14415 | CHEMBL2095186 | AT1B2 | Drugged | Protein complex group | CHEMBL1751 | Digoxin | Small molecule | Inhibitor | BNF | Side-effect | Common or very common | 4 | Vomiting | No |
| P14415 | CHEMBL2095186 | AT1B2 | Drugged | Protein complex group | CHEMBL1751 | Digoxin | Small molecule | Inhibitor | BNF | Side-effect | Common or very common | 4 | Skin reactions | No |
| P14415 | CHEMBL2095186 | AT1B2 | Drugged | Protein complex group | CHEMBL1751 | Digoxin | Small molecule | Inhibitor | BNF | Side-effect | Common or very common | 4 | Vision disorder | No |
| P14415 | CHEMBL2095186 | AT1B2 | Drugged | Protein complex group | CHEMBL1751 | Digoxin | Small molecule | Inhibitor | BNF | Side-effect | Common or very common | 4 | Eosinophilia | No |
| P14415 | CHEMBL2095186 | AT1B2 | Drugged | Protein complex group | CHEMBL1751 | Digoxin | Small molecule | Inhibitor | BNF | Side-effect | Common or very common | 4 | Dizziness | No |
| P14415 | CHEMBL2095186 | AT1B2 | Drugged | Protein complex group | CHEMBL1751 | Digoxin | Small molecule | Inhibitor | BNF | Side-effect | Common or very common | 4 | Diarrhoea | No |
| P14415 | CHEMBL2095186 | AT1B2 | Drugged | Protein complex group | CHEMBL1751 | Digoxin | Small molecule | Inhibitor | BNF | Side-effect | Common or very common | 4 | Cerebral impairment | No |
| P14415 | CHEMBL2095186 | AT1B2 | Drugged | Protein complex group | CHEMBL1751 | Digoxin | Small molecule | Inhibitor | BNF | Side-effect | Common or very common | 4 | Cardiac conduction disorder | Yes |
| P14415 | CHEMBL2095186 | AT1B2 | Drugged | Protein complex group | CHEMBL1751 | Digoxin | Small molecule | Inhibitor | BNF | Side-effect | Common or very common | 4 | Nausea | No |
| O75882 | - | ATRN | Not yet druggable | - | - | - | - | - | - | - | - | - | - | No |
| O75973 | - | C1QRF | Not yet druggable | - | - | - | - | - | - | - | - | - | - | No |
| P43155 | CHEMBL3184 | CACP | Drugged | Single protein | CHEMBL1149 | Levocarnitine | Small molecule | - | ChEMBL | Indication | All | 3 | Cytomegalovirus infection | No |
| P43155 | CHEMBL3184 | CACP | Drugged | Single protein | CHEMBL1149 | Levocarnitine | Small molecule | - | ChEMBL | Indication | All | 3 | Hyperthyroidism | No |
| P43155 | CHEMBL3184 | CACP | Drugged | Single protein | CHEMBL1149 | Levocarnitine | Small molecule | - | ChEMBL | Indication | All | 0,5 | Respiratory insufficiency | No |
| P43155 | CHEMBL3184 | CACP | Drugged | Single protein | CHEMBL1149 | Levocarnitine | Small molecule | - | ChEMBL | Indication | All | 2 | Heart failure | Yes |
| P43155 | CHEMBL3184 | CACP | Drugged | Single protein | CHEMBL1149 | Levocarnitine | Small molecule | - | ChEMBL | Indication | All | 3 | Sarcopenia | No |
| P43155 | CHEMBL3184 | CACP | Drugged | Single protein | CHEMBL1149 | Levocarnitine | Small molecule | - | ChEMBL | Indication | All | 4 | Muscle weakness | No |
| P43155 | CHEMBL3184 | CACP | Drugged | Single protein | CHEMBL1149 | Levocarnitine | Small molecule | - | ChEMBL | Indication | All | 2 | Cancers | No |
| P43155 | CHEMBL3184 | CACP | Drugged | Single protein | CHEMBL1149 | Levocarnitine | Small molecule | - | ChEMBL | Indication | All | 3 | Hiv infection | No |
| P43155 | CHEMBL3184 | CACP | Drugged | Single protein | CHEMBL1149 | Levocarnitine | Small molecule | - | ChEMBL | Indication | All | 4 | Brain diseases | No |
| P43155 | CHEMBL3184 | CACP | Drugged | Single protein | CHEMBL1149 | Levocarnitine | Small molecule | - | ChEMBL | Indication | All | 2 | Acquired immunodeficiency syndrome | No |
| P43155 | CHEMBL3184 | CACP | Drugged | Single protein | CHEMBL1149 | Levocarnitine | Small molecule | - | ChEMBL | Indication | All | 3 | Multiple sclerosis | No |
| P43155 | CHEMBL3184 | CACP | Drugged | Single protein | CHEMBL1149 | Levocarnitine | Small molecule | - | ChEMBL | Indication | All | 3 | Cancers | No |
| P43155 | CHEMBL3184 | CACP | Drugged | Single protein | CHEMBL1149 | Levocarnitine | Small molecule | - | ChEMBL | Indication | All | 2 | Spinal cord injuries | No |
| P43155 | CHEMBL3184 | CACP | Drugged | Single protein | CHEMBL1149 | Levocarnitine | Small molecule | - | ChEMBL | Indication | All | 3 | Lipid metabolism disorder | No |
| P43155 | CHEMBL3184 | CACP | Drugged | Single protein | CHEMBL1149 | Levocarnitine | Small molecule | - | ChEMBL | Indication | All | 2 | Anemia | No |
| P43155 | CHEMBL3184 | CACP | Drugged | Single protein | CHEMBL1149 | Levocarnitine | Small molecule | - | ChEMBL | Indication | All | 4 | Kidney failure, chronic | No |
| P43155 | CHEMBL3184 | CACP | Drugged | Single protein | CHEMBL1149 | Levocarnitine | Small molecule | - | ChEMBL | Indication | All | 2 | Muscle cramp | No |
| P43155 | CHEMBL3184 | CACP | Drugged | Single protein | CHEMBL1149 | Levocarnitine | Small molecule | - | ChEMBL | Indication | All | 2 | Diabetes mellitus, type 2 | No |
| P43155 | CHEMBL3184 | CACP | Drugged | Single protein | CHEMBL1149 | Levocarnitine | Small molecule | - | ChEMBL | Indication | All | 3 | Acute kidney injury | No |
| P43155 | CHEMBL3184 | CACP | Drugged | Single protein | CHEMBL1149 | Levocarnitine | Small molecule | - | ChEMBL | Indication | All | 3 | Atrial fibrillation | Yes |
| P43155 | CHEMBL3184 | CACP | Drugged | Single protein | CHEMBL1149 | Levocarnitine | Small molecule | - | ChEMBL | Indication | All | 2 | Polycystic ovary syndrome | No |
| P43155 | CHEMBL3184 | CACP | Drugged | Single protein | CHEMBL1149 | Levocarnitine | Small molecule | - | ChEMBL | Indication | All | 3 | Peripheral arterial disease | No |
| P43155 | CHEMBL3184 | CACP | Drugged | Single protein | CHEMBL1149 | Levocarnitine | Small molecule | - | ChEMBL | Indication | All | 2 | Renal insufficiency | No |
| P43155 | CHEMBL3184 | CACP | Drugged | Single protein | CHEMBL1149 | Levocarnitine | Small molecule | - | ChEMBL | Indication | All | 3 | Muscular atrophy, spinal | No |
| P43155 | CHEMBL3184 | CACP | Drugged | Single protein | CHEMBL1149 | Levocarnitine | Small molecule | - | ChEMBL | Indication | All | 1 | Shock, septic | No |
| P43155 | CHEMBL3184 | CACP | Drugged | Single protein | CHEMBL1149 | Levocarnitine | Small molecule | - | ChEMBL | Indication | All | 2 | Non-alcoholic fatty liver disease | No |
| P43155 | CHEMBL3184 | CACP | Drugged | Single protein | CHEMBL1149 | Levocarnitine | Small molecule | - | ChEMBL | Indication | All | 3 | Pyelonephritis | No |
| P17655 | CHEMBL2382 | CAN2 | Drugged | Single protein | CHEMBL325041 | Bortezomib | Small molecule | Inhibitor | ChEMBL | Indication | All | 3 | Plasmacytoma | No |
| P17655 | CHEMBL2382 | CAN2 | Drugged | Single protein | CHEMBL325041 | Bortezomib | Small molecule | Inhibitor | ChEMBL | Indication | All | 3 | Cancers | No |
| P17655 | CHEMBL2382 | CAN2 | Drugged | Single protein | CHEMBL325041 | Bortezomib | Small molecule | Inhibitor | ChEMBL | Indication | All | 2 | Cancers | No |

**Table S8. Druggability results of the prioritised proteins**

| Target Uniprot ID* | Target ChEMBL ID | Target protein name | Druggability* | Target type | Drug ChEMBL ID | Drug name | Drug molecule type |
|---|---|---|---|---|---|---|---|
| P17655 | CHEMBL2382 | CAN2 | Drugged | Single protein | CHEMBL325041 | Bortezomib | Small molecule |
| P17655 | CHEMBL2382 | CAN2 | Drugged | Single protein | CHEMBL325041 | Bortezomib | Small molecule |
| P17655 | CHEMBL2382 | CAN2 | Drugged | Single protein | CHEMBL325041 | Bortezomib | Small molecule |
| P17655 | CHEMBL2382 | CAN2 | Drugged | Single protein | CHEMBL325041 | Bortezomib | Small molecule |
| P17655 | CHEMBL2382 | CAN2 | Drugged | Single protein | CHEMBL325041 | Bortezomib | Small molecule |
| P17655 | CHEMBL2382 | CAN2 | Drugged | Single protein | CHEMBL325041 | Bortezomib | Small molecule |
| P17655 | CHEMBL2382 | CAN2 | Drugged | Single protein | CHEMBL451887 | Carfilzomib | Protein |
| P17655 | CHEMBL2382 | CAN2 | Drugged | Single protein | CHEMBL451887 | Carfilzomib | Protein |
| P17655 | CHEMBL2382 | CAN2 | Drugged | Single protein | CHEMBL451887 | Carfilzomib | Protein |
| P17655 | CHEMBL2382 | CAN2 | Drugged | Single protein | CHEMBL451887 | Carfilzomib | Protein |
| P17655 | CHEMBL2382 | CAN2 | Drugged | Single protein | CHEMBL451887 | Carfilzomib | Protein |
| P17655 | CHEMBL2382 | CAN2 | Drugged | Single protein | CHEMBL451887 | Carfilzomib | Protein |
| P17655 | CHEMBL2382 | CAN2 | Drugged | Single protein | CHEMBL451887 | Carfilzomib | Protein |
| P17655 | CHEMBL2382 | CAN2 | Drugged | Single protein | CHEMBL451887 | Carfilzomib | Protein |
| P17655 | CHEMBL2382 | CAN2 | Drugged | Single protein | CHEMBL325041 | Bortezomib | Small molecule |
| P17655 | CHEMBL2382 | CAN2 | Drugged | Single protein | CHEMBL451887 | Carfilzomib | Protein |
| P17655 | CHEMBL2382 | CAN2 | Drugged | Single protein | CHEMBL325041 | Bortezomib | Small molecule |
| P17655 | CHEMBL2382 | CAN2 | Drugged | Single protein | CHEMBL325041 | Bortezomib | Small molecule |
| P17655 | CHEMBL2382 | CAN2 | Drugged | Single protein | CHEMBL325041 | Bortezomib | Small molecule |
| P17655 | CHEMBL2382 | CAN2 | Drugged | Single protein | CHEMBL451887 | Carfilzomib | Protein |
| P17655 | CHEMBL2382 | CAN2 | Drugged | Single protein | CHEMBL325041 | Bortezomib | Small molecule |
| P17655 | CHEMBL2382 | CAN2 | Drugged | Single protein | CHEMBL325041 | Bortezomib | Small molecule |
| P17655 | CHEMBL2382 | CAN2 | Drugged | Single protein | CHEMBL325041 | Bortezomib | Small molecule |
| P17655 | CHEMBL2382 | CAN2 | Drugged | Single protein | CHEMBL325041 | Bortezomib | Small molecule |
| P17655 | CHEMBL2382 | CAN2 | Drugged | Single protein | CHEMBL325041 | Bortezomib | Small molecule |
| P17655 | CHEMBL2382 | CAN2 | Drugged | Single protein | CHEMBL325041 | Bortezomib | Small molecule |
| P17655 | CHEMBL2382 | CAN2 | Drugged | Single protein | CHEMBL325041 | Bortezomib | Small molecule |
| P17655 | CHEMBL2382 | CAN2 | Drugged | Single protein | CHEMBL325041 | Bortezomib | Small molecule |
| P17655 | CHEMBL2382 | CAN2 | Drugged | Single protein | CHEMBL325041 | Bortezomib | Small molecule |
| P17655 | CHEMBL2382 | CAN2 | Drugged | Single protein | CHEMBL325041 | Bortezomib | Small molecule |
| P17655 | CHEMBL2382 | CAN2 | Drugged | Single protein | CHEMBL325041 | Bortezomib | Small molecule |
| P17655 | CHEMBL2382 | CAN2 | Drugged | Single protein | CHEMBL325041 | Bortezomib | Small molecule |
| P17655 | CHEMBL2382 | CAN2 | Drugged | Single protein | CHEMBL325041 | Bortezomib | Small molecule |
| P17655 | CHEMBL2382 | CAN2 | Drugged | Single protein | CHEMBL325041 | Bortezomib | Small molecule |
| Q9UBX1 | CHEMBL2517 | CATF | Drugged | Single protein | CHEMBL231813 | Telaprevir | Protein |
| Q9UBX1 | CHEMBL2517 | CATF | Drugged | Single protein | CHEMBL218394 | Boceprevir | Small molecule |
| Q9UBX1 | CHEMBL2517 | CATF | Drugged | Single protein | CHEMBL231813 | Telaprevir | Protein |
| Q9UBX1 | CHEMBL2517 | CATF | Drugged | Single protein | CHEMBL231813 | Telaprevir | Protein |
| Q9UBX1 | CHEMBL2517 | CATF | Drugged | Single protein | CHEMBL231813 | Telaprevir | Protein |
| Q9UBX1 | CHEMBL2517 | CATF | Drugged | Single protein | CHEMBL231813 | Telaprevir | Protein |
| Q9UBX1 | CHEMBL2517 | CATF | Drugged | Single protein | CHEMBL218394 | Boceprevir | Small molecule |
| Q9UBX1 | CHEMBL2517 | CATF | Drugged | Single protein | CHEMBL218394 | Boceprevir | Small molecule |
| Q9UBX1 | CHEMBL2517 | CATF | Drugged | Single protein | CHEMBL231813 | Telaprevir | Protein |
| Q9UBX1 | CHEMBL2517 | CATF | Drugged | Single protein | CHEMBL218394 | Boceprevir | Small molecule |
| Q96EE4 | - | CC126 | Not yet druggable | - | - | - | - |
| P80075 | - | CCL8 | Not yet druggable | - | - | - | - |
| Q9NZK5 | - | CECR1 | Not yet druggable | - | - | - | - |
| P19835 | CHEMBL3219 | CEL | Drugged | Single protein | CHEMBL175247 | Orlistat | Small molecule |
| P19835 | CHEMBL3219 | CEL | Drugged | Single protein | CHEMBL175247 | Orlistat | Small molecule |
| P19835 | CHEMBL3219 | CEL | Drugged | Single protein | CHEMBL175247 | Orlistat | Small molecule |
| P19835 | CHEMBL3219 | CEL | Drugged | Single protein | CHEMBL175247 | Orlistat | Small molecule |
| P19835 | CHEMBL3219 | CEL | Drugged | Single protein | CHEMBL175247 | Orlistat | Small molecule |
| Q5QGZ9 | - | CL12A | Not yet druggable | - | - | - | - |
| P09543 | - | CN37 | Not yet druggable | - | - | - | - |
| Q8WW22 | - | DNJA4 | Not yet druggable | - | - | - | - |
| P16444 | CHEMBL1989 | DPEP1 | Drugged | Single protein | CHEMBL766 | Cilastatin | Small molecule |
| P16444 | CHEMBL1989 | DPEP1 | Drugged | Single protein | CHEMBL766 | Cilastatin | Small molecule |
| P16444 | CHEMBL1989 | DPEP1 | Drugged | Single protein | CHEMBL766 | Cilastatin | Small molecule |
| P16444 | CHEMBL1989 | DPEP1 | Drugged | Single protein | CHEMBL766 | Cilastatin | Small molecule |
| P16444 | CHEMBL1989 | DPEP1 | Drugged | Single protein | CHEMBL766 | Cilastatin | Small molecule |
| P16444 | CHEMBL1989 | DPEP1 | Drugged | Single protein | CHEMBL766 | Cilastatin | Small molecule |
| P16444 | CHEMBL1989 | DPEP1 | Drugged | Single protein | CHEMBL766 | Cilastatin | Small molecule |
| P16444 | CHEMBL1989 | DPEP1 | Drugged | Single protein | CHEMBL766 | Cilastatin | Small molecule |
| P16444 | CHEMBL1989 | DPEP1 | Drugged | Single protein | CHEMBL766 | Cilastatin | Small molecule |
| P16444 | CHEMBL1989 | DPEP1 | Drugged | Single protein | CHEMBL766 | Cilastatin | Small molecule |
| P16444 | CHEMBL1989 | DPEP1 | Drugged | Single protein | CHEMBL766 | Cilastatin | Small molecule |
| P16444 | CHEMBL1989 | DPEP1 | Drugged | Single protein | CHEMBL766 | Cilastatin | Small molecule |
| P16444 | CHEMBL1989 | DPEP1 | Drugged | Single protein | CHEMBL766 | Cilastatin | Small molecule |
| P16444 | CHEMBL1989 | DPEP1 | Drugged | Single protein | CHEMBL766 | Cilastatin | Small molecule |
| P16444 | CHEMBL1989 | DPEP1 | Drugged | Single protein | CHEMBL766 | Cilastatin | Small molecule |
| P16444 | CHEMBL1989 | DPEP1 | Drugged | Single protein | CHEMBL766 | Cilastatin | Small molecule |
| P16444 | CHEMBL1989 | DPEP1 | Drugged | Single protein | CHEMBL766 | Cilastatin | Small molecule |
| P16444 | CHEMBL1989 | DPEP1 | Drugged | Single protein | CHEMBL766 | Cilastatin | Small molecule |
| P16444 | CHEMBL1989 | DPEP1 | Drugged | Single protein | CHEMBL766 | Cilastatin | Small molecule |
| P16444 | CHEMBL1989 | DPEP1 | Drugged | Single protein | CHEMBL1201057 | Cilastatin sodium | Small molecule |
| P16444 | CHEMBL1989 | DPEP1 | Drugged | Single protein | CHEMBL1201057 | Cilastatin sodium | Small molecule |
| P16444 | CHEMBL1989 | DPEP1 | Drugged | Single protein | CHEMBL1201057 | Cilastatin sodium | Small molecule |
| P16444 | CHEMBL1989 | DPEP1 | Drugged | Single protein | CHEMBL1201057 | Cilastatin sodium | Small molecule |
| P16444 | CHEMBL1989 | DPEP1 | Drugged | Single protein | CHEMBL766 | Cilastatin | Small molecule |
| P16444 | CHEMBL1989 | DPEP1 | Drugged | Single protein | CHEMBL766 | Cilastatin | Small molecule |
| P16444 | CHEMBL1989 | DPEP1 | Drugged | Single protein | CHEMBL766 | Cilastatin | Small molecule |
| P16444 | CHEMBL1989 | DPEP1 | Drugged | Single protein | CHEMBL766 | Cilastatin | Small molecule |
| P16444 | CHEMBL1989 | DPEP1 | Drugged | Single protein | CHEMBL766 | Cilastatin | Small molecule |
| P16444 | CHEMBL1989 | DPEP1 | Drugged | Single protein | CHEMBL766 | Cilastatin | Small molecule |
| P16444 | CHEMBL1989 | DPEP1 | Drugged | Single protein | CHEMBL766 | Cilastatin | Small molecule |
| P16444 | CHEMBL1989 | DPEP1 | Drugged | Single protein | CHEMBL766 | Cilastatin | Small molecule |
| P16444 | CHEMBL1989 | DPEP1 | Drugged | Single protein | CHEMBL766 | Cilastatin | Small molecule |

| Target Uniprot ID* | Drug mechanism | Drug effect source | Drug effect type | Drug effect frequency | Max phase* | Drug effect | Drug effect class |
|---|---|---|---|---|---|---|---|
| P17655 | Inhibitor | ChEMBL | Indication | All | 1 | Cancers | No |
| P17655 | Inhibitor | ChEMBL | Indication | All | 3 | Amyloidosis, familial | No |
| P17655 | Inhibitor | ChEMBL | Indication | All | 2 | Lymphoproliferative disorder | No |
| P17655 | Inhibitor | ChEMBL | Indication | All | 3 | Renal insufficiency | No |
| P17655 | Inhibitor | ChEMBL | Indication | All | 2 | Immune system diseases | No |
| P17655 | Inhibitor | ChEMBL | Indication | All | 4 | Cancers | No |
| P17655 | Inhibitor | ChEMBL | Indication | All | 1 | Immunoglobulin light-chain amyloidosis | No |
| P17655 | Inhibitor | ChEMBL | Indication | All | 3 | Waldenstrom macroglobulinemia | No |
| P17655 | Inhibitor | ChEMBL | Indication | All | 1 | Hodgkin disease | No |
| P17655 | Inhibitor | ChEMBL | Indication | All | 1 | Sezary syndrome | No |
| P17655 | Inhibitor | ChEMBL | Indication | All | 1 | Plasmacytoma | No |
| P17655 | Inhibitor | ChEMBL | Indication | All | 1 | Amyloidosis | No |
| P17655 | Inhibitor | ChEMBL | Indication | All | 2 | Cancers | No |
| P17655 | Inhibitor | ChEMBL | Indication | All | 1 | Cancers | No |
| P17655 | Inhibitor | ChEMBL | Indication | All | 2 | Hodgkin disease | No |
| P17655 | Inhibitor | ChEMBL | Indication | All | 4 | Cancers | No |
| P17655 | Inhibitor | ChEMBL | Indication | All | 2 | Bronchiolitis obliterans | No |
| P17655 | Inhibitor | ChEMBL | Indication | All | 3 | Waldenstrom macroglobulinemia | No |
| P17655 | Inhibitor | ChEMBL | Indication | All | 1 | Immunoblastic lymphadenopathy | No |
| P17655 | Inhibitor | ChEMBL | Indication | All | 1 | Mycosis fungoides | No |
| P17655 | Inhibitor | ChEMBL | Indication | All | 1 | Sezary syndrome | No |
| P17655 | Inhibitor | ChEMBL | Indication | All | 3 | Immunoglobulin light-chain amyloidosis | No |
| P17655 | Inhibitor | ChEMBL | Indication | All | 2 | Graft vs host disease | No |
| P17655 | Inhibitor | ChEMBL | Indication | All | 2 | Red-cell aplasia, pure | No |
| P17655 | Inhibitor | ChEMBL | Indication | All | 2 | Neuromyelitis optica | No |
| P17655 | Inhibitor | ChEMBL | Indication | All | 3 | Amyloidosis | No |
| P17655 | Inhibitor | ChEMBL | Indication | All | 1 | Smoldering multiple myeloma | No |
| P17655 | Inhibitor | ChEMBL | Indication | All | 0,5 | Myeloproliferative disorder | No |
| P17655 | Inhibitor | ChEMBL | Indication | All | 3 | Purpura, thrombocytopenic, idiopathic | No |
| P17655 | Inhibitor | ChEMBL | Indication | All | 2 | Castleman disease | No |
| P17655 | Inhibitor | ChEMBL | Indication | All | 1 | Uremia | No |
| P17655 | Inhibitor | ChEMBL | Indication | All | 2 | Encephalitis, japanese | No |
| P17655 | Inhibitor | ChEMBL | Indication | All | 1 | Mycosis fungoides | No |
| P17655 | Inhibitor | ChEMBL | Indication | All | 1 | Anemia | No |
| Q9UBX1 | Inhibitor | ChEMBL | Indication | All | 3 | Infections | No |
| Q9UBX1 | Inhibitor | ChEMBL | Indication | All | 3 | Hepatitis c | No |
| Q9UBX1 | Inhibitor | ChEMBL | Indication | All | 4 | Virus diseases | No |
| Q9UBX1 | Inhibitor | ChEMBL | Indication | All | 1 | Liver diseases | No |
| Q9UBX1 | Inhibitor | ChEMBL | Indication | All | 3 | Hepatitis c | No |
| Q9UBX1 | Inhibitor | ChEMBL | Indication | All | 2 | Thrombocytopenia | No |
| Q9UBX1 | Inhibitor | ChEMBL | Indication | All | 4 | Virus diseases | No |
| Q9UBX1 | Inhibitor | ChEMBL | Indication | All | 1 | Thrombocytopenia | No |
| Q9UBX1 | Inhibitor | ChEMBL | Indication | All | 3 | Hiv infection | No |
| Q9UBX1 | Inhibitor | ChEMBL | Indication | All | 4 | Hepatitis c | No |
| Q96EE4 | - | - | - | - | - | - | No |
| P80075 | - | - | - | - | - | - | No |
| Q9NZK5 | - | - | - | - | - | - | No |
| P19835 | Inhibitor | ChEMBL | Indication | All | 3 | Infertility | No |
| P19835 | Inhibitor | ChEMBL | Indication | All | 1 | Metabolic syndrome | No |
| P19835 | Inhibitor | ChEMBL | Indication | All | 2 | Hyperlipoproteinemia type i | No |
| P19835 | Inhibitor | ChEMBL | Indication | All | 4 | Obesity | No |
| P19835 | Inhibitor | ChEMBL | Indication | All | 2 | Polycystic ovary syndrome | No |
| Q5QGZ9 | - | - | - | - | - | - | No |
| P09543 | - | - | - | - | - | - | No |
| Q8WW22 | - | - | - | - | - | - | No |
| P16444 | Inhibitor | BNF | Side-effect | Rare or very rare | 4 | Encephalopathy | No |
| P16444 | Inhibitor | BNF | Side-effect | Common or very common | 4 | Eosinophilia | No |
| P16444 | Inhibitor | BNF | Side-effect | Common or very common | 4 | Nausea | No |
| P16444 | Inhibitor | BNF | Side-effect | Common or very common | 4 | Skin reactions | No |
| P16444 | Inhibitor | BNF | Side-effect | Common or very common | 4 | Thrombophlebitis | No |
| P16444 | Inhibitor | BNF | Side-effect | Common or very common | 4 | Vomiting | No |
| P16444 | Inhibitor | BNF | Side-effect | Not known | 4 | Agitation | No |
| P16444 | Inhibitor | BNF | Side-effect | Rare or very rare | 4 | Flushing | No |
| P16444 | Inhibitor | BNF | Side-effect | Rare or very rare | 4 | Agranulocytosis | No |
| P16444 | Inhibitor | BNF | Side-effect | Rare or very rare | 4 | Angiodema | No |
| P16444 | Inhibitor | BNF | Side-effect | Rare or very rare | 4 | Antibiotic associated colitis | No |
| P16444 | Inhibitor | BNF | Side-effect | Rare or very rare | 4 | Chest discomfort | Yes |
| P16444 | Inhibitor | BNF | Side-effect | Rare or very rare | 4 | Colitis hemorrhagic | No |
| P16444 | Inhibitor | BNF | Side-effect | Common or very common | 4 | Diarrhoea | No |
| P16444 | Inhibitor | BNF | Side-effect | Rare or very rare | 4 | Dyspnoea | No |
| P16444 | Inhibitor | BNF | Side-effect | Rare or very rare | 4 | Anaphylactic reaction | No |
| P16444 | Inhibitor | BNF | Side-effect | Rare or very rare | 4 | Cyanosis | No |
| P16444 | Inhibitor | BNF | Indication | All | 4 | Empirical treatment of infection in febrile patients with neutropenia | No |
| P16444 | Inhibitor | BNF | Indication | All | 4 | Infection caused by | No |
| P16444 | Inhibitor | ChEMBL | Indication | All | 4 | Pneumonia | No |
| P16444 | Inhibitor | ChEMBL | Indication | All | 4 | Urinary tract infection | No |
| P16444 | Inhibitor | ChEMBL | Indication | All | 4 | Sepsis | No |
| P16444 | Inhibitor | ChEMBL | Indication | All | 4 | Intraabdominal infection | No |
| P16444 | Inhibitor | ChEMBL | Indication | All | 0,5 | Osteomyelitis | No |
| P16444 | Inhibitor | ChEMBL | Indication | All | 1 | Sepsis | No |
| P16444 | Inhibitor | ChEMBL | Indication | All | 2 | Infections | No |
| P16444 | Inhibitor | ChEMBL | Indication | All | 2 | Urinary tract infection | No |
| P16444 | Inhibitor | ChEMBL | Indication | All | 3 | Bacterial infection | No |
| P16444 | Inhibitor | ChEMBL | Indication | All | 3 | Pyelonephritis | No |
| P16444 | Inhibitor | ChEMBL | Indication | All | 3 | Pneumonia | No |
| P16444 | Inhibitor | BNF | Indication | All | 4 | Or other less sensitive organisms | No |
| P16444 | Inhibitor | BNF | Indication | All | 4 | Aerobic and anaerobic gram-positive and gram-negative infection | No |

**Table S8. Druggability results of the prioritised proteins**

| Target Uniprot ID* | Target ChEMBL ID | Target protein name | Druggability* | Target type | Drug ChEMBL ID | Drug name | Drug molecule type | Drug mechanism | Drug effect source | Drug effect type | Drug effect frequency | Max phase* | Drug effect | Drug effect class |
|---|---|---|---|---|---|---|---|---|---|---|---|---|---|---|
| P16444 | CHEMBL1989 | DPEP1 | Drugged | Single protein | CHEMBL766 | Cilastatin | Small molecule | Inhibitor | BNF | Side-effect | Rare or very rare | 4 | Focal tremor | No |
| P16444 | CHEMBL1989 | DPEP1 | Drugged | Single protein | CHEMBL766 | Cilastatin | Small molecule | Inhibitor | BNF | Indication | All | 4 | Hospital-acquired septicemia | No |
| P16444 | CHEMBL1989 | DPEP1 | Drugged | Single protein | CHEMBL766 | Cilastatin | Small molecule | Inhibitor | BNF | Indication | All | 4 | Life-threatening infection | No |
| P16444 | CHEMBL1989 | DPEP1 | Drugged | Single protein | CHEMBL766 | Cilastatin | Small molecule | Inhibitor | BNF | Side-effect | Rare or very rare | 4 | Gastrointestinal discomfort | No |
| P16444 | CHEMBL1989 | DPEP1 | Drugged | Single protein | CHEMBL766 | Cilastatin | Small molecule | Inhibitor | BNF | Side-effect | Rare or very rare | 4 | Hyperhidrosis | No |
| P16444 | CHEMBL1989 | DPEP1 | Drugged | Single protein | CHEMBL766 | Cilastatin | Small molecule | Inhibitor | BNF | Side-effect | Rare or very rare | 4 | Headache | No |
| P16444 | CHEMBL1989 | DPEP1 | Drugged | Single protein | CHEMBL766 | Cilastatin | Small molecule | Inhibitor | BNF | Side-effect | Rare or very rare | 4 | Urine discolouration | No |
| P16444 | CHEMBL1989 | DPEP1 | Drugged | Single protein | CHEMBL766 | Cilastatin | Small molecule | Inhibitor | BNF | Side-effect | Rare or very rare | 4 | Vertigo | No |
| P16444 | CHEMBL1989 | DPEP1 | Drugged | Single protein | CHEMBL766 | Cilastatin | Small molecule | Inhibitor | BNF | Side-effect | Uncommon | 4 | Bone marrow disorder | No |
| P16444 | CHEMBL1989 | DPEP1 | Drugged | Single protein | CHEMBL766 | Cilastatin | Small molecule | Inhibitor | BNF | Side-effect | Uncommon | 4 | Confusion | No |
| P16444 | CHEMBL1989 | DPEP1 | Drugged | Single protein | CHEMBL766 | Cilastatin | Small molecule | Inhibitor | BNF | Side-effect | Uncommon | 4 | Dizziness | No |
| P16444 | CHEMBL1989 | DPEP1 | Drugged | Single protein | CHEMBL766 | Cilastatin | Small molecule | Inhibitor | BNF | Side-effect | Uncommon | 4 | Drowsiness | No |
| P16444 | CHEMBL1989 | DPEP1 | Drugged | Single protein | CHEMBL766 | Cilastatin | Small molecule | Inhibitor | BNF | Side-effect | Uncommon | 4 | Hallucination | No |
| P16444 | CHEMBL1989 | DPEP1 | Drugged | Single protein | CHEMBL766 | Cilastatin | Small molecule | Inhibitor | BNF | Side-effect | Uncommon | 4 | Hypotension | No |
| P16444 | CHEMBL1989 | DPEP1 | Drugged | Single protein | CHEMBL766 | Cilastatin | Small molecule | Inhibitor | BNF | Side-effect | Uncommon | 4 | Leucopenia | No |
| P16444 | CHEMBL1989 | DPEP1 | Drugged | Single protein | CHEMBL766 | Cilastatin | Small molecule | Inhibitor | BNF | Side-effect | Uncommon | 4 | Movement disorder | No |
| P16444 | CHEMBL1989 | DPEP1 | Drugged | Single protein | CHEMBL766 | Cilastatin | Small molecule | Inhibitor | BNF | Side-effect | Uncommon | 4 | Psychiatric disorder | No |
| P16444 | CHEMBL1989 | DPEP1 | Drugged | Single protein | CHEMBL766 | Cilastatin | Small molecule | Inhibitor | BNF | Side-effect | Uncommon | 4 | Seizure | No |
| P16444 | CHEMBL1989 | DPEP1 | Drugged | Single protein | CHEMBL766 | Cilastatin | Small molecule | Inhibitor | BNF | Side-effect | Uncommon | 4 | Thrombocytopenia | No |
| P16444 | CHEMBL1989 | DPEP1 | Drugged | Single protein | CHEMBL766 | Cilastatin | Small molecule | Inhibitor | BNF | Side-effect | Uncommon | 4 | Thrombocytosis | No |
| P16444 | CHEMBL1989 | DPEP1 | Drugged | Single protein | CHEMBL766 | Cilastatin | Small molecule | Inhibitor | BNF | Indication | All | 4 | Aerobic gram-negative infection | No |
| P16444 | CHEMBL1989 | DPEP1 | Drugged | Single protein | CHEMBL766 | Cilastatin | Small molecule | Inhibitor | BNF | Side-effect | Rare or very rare | 4 | Tooth discolouration | No |
| P16444 | CHEMBL1989 | DPEP1 | Drugged | Single protein | CHEMBL766 | Cilastatin | Small molecule | Inhibitor | BNF | Side-effect | Rare or very rare | 4 | Anemia | No |
| P16444 | CHEMBL1989 | DPEP1 | Drugged | Single protein | CHEMBL766 | Cilastatin | Small molecule | Inhibitor | BNF | Side-effect | Rare or very rare | 4 | Tongue discolouration | No |
| P16444 | CHEMBL1989 | DPEP1 | Drugged | Single protein | CHEMBL766 | Cilastatin | Small molecule | Inhibitor | BNF | Side-effect | Rare or very rare | 4 | Taste altered | No |
| P16444 | CHEMBL1989 | DPEP1 | Drugged | Single protein | CHEMBL766 | Cilastatin | Small molecule | Inhibitor | BNF | Side-effect | Rare or very rare | 4 | Hearing loss | No |
| P16444 | CHEMBL1989 | DPEP1 | Drugged | Single protein | CHEMBL766 | Cilastatin | Small molecule | Inhibitor | BNF | Side-effect | Rare or very rare | 4 | Hepatic disorder | No |
| P16444 | CHEMBL1989 | DPEP1 | Drugged | Single protein | CHEMBL1201057 | Cilastatin sodium | Small molecule | Inhibitor | ChEMBL | Indication | All | 4 | Infections | No |
| P16444 | CHEMBL1989 | DPEP1 | Drugged | Single protein | CHEMBL766 | Cilastatin | Small molecule | Inhibitor | BNF | Side-effect | Rare or very rare | 4 | Hyperventilation | No |
| P16444 | CHEMBL1989 | DPEP1 | Drugged | Single protein | CHEMBL766 | Cilastatin | Small molecule | Inhibitor | BNF | Side-effect | Rare or very rare | 4 | Increased risk of infection | No |
| P16444 | CHEMBL1989 | DPEP1 | Drugged | Single protein | CHEMBL766 | Cilastatin | Small molecule | Inhibitor | BNF | Side-effect | Rare or very rare | 4 | Myasthenia gravis | No |
| P16444 | CHEMBL1989 | DPEP1 | Drugged | Single protein | CHEMBL766 | Cilastatin | Small molecule | Inhibitor | BNF | Side-effect | Rare or very rare | 4 | Oral disorder | No |
| P16444 | CHEMBL1989 | DPEP1 | Drugged | Single protein | CHEMBL766 | Cilastatin | Small molecule | Inhibitor | BNF | Side-effect | Rare or very rare | 4 | Palpitations | Yes |
| P16444 | CHEMBL1989 | DPEP1 | Drugged | Single protein | CHEMBL766 | Cilastatin | Small molecule | Inhibitor | BNF | Side-effect | Rare or very rare | 4 | Paraesthesia | No |
| P16444 | CHEMBL1989 | DPEP1 | Drugged | Single protein | CHEMBL766 | Cilastatin | Small molecule | Inhibitor | BNF | Side-effect | Rare or very rare | 4 | Polyarthralgia | No |
| P16444 | CHEMBL1989 | DPEP1 | Drugged | Single protein | CHEMBL766 | Cilastatin | Small molecule | Inhibitor | BNF | Side-effect | Rare or very rare | 4 | Polyuria | No |
| P16444 | CHEMBL1989 | DPEP1 | Drugged | Single protein | CHEMBL766 | Cilastatin | Small molecule | Inhibitor | BNF | Side-effect | Rare or very rare | 4 | Renal impairment | No |
| P16444 | CHEMBL1989 | DPEP1 | Drugged | Single protein | CHEMBL766 | Cilastatin | Small molecule | Inhibitor | BNF | Side-effect | Rare or very rare | 4 | Severe cutaneous adverse reactions scars | No |
| P16444 | CHEMBL1989 | DPEP1 | Drugged | Single protein | CHEMBL766 | Cilastatin | Small molecule | Inhibitor | BNF | Side-effect | Rare or very rare | 4 | Spinal pain | No |
| P16444 | CHEMBL1989 | DPEP1 | Drugged | Single protein | CHEMBL766 | Cilastatin | Small molecule | Inhibitor | BNF | Side-effect | Rare or very rare | 4 | Tachycardia | Yes |
| P16444 | CHEMBL1989 | DPEP1 | Drugged | Single protein | CHEMBL766 | Cilastatin | Small molecule | Inhibitor | BNF | Side-effect | Rare or very rare | 4 | Tinnitus | No |
| P16444 | CHEMBL1989 | DPEP1 | Drugged | Single protein | CHEMBL1201057 | Cilastatin sodium | Small molecule | Inhibitor | ChEMBL | Indication | All | 4 | Endocarditis | Yes |
| P16444 | CHEMBL1989 | DPEP1 | Drugged | Single protein | CHEMBL766 | Cilastatin | Small molecule | Inhibitor | ChEMBL | Indication | All | 2 | Cancers | No |
| Q9UNE0 | CHEMBL1250376 | EDAR | Druggable | Single protein | - | - | - | - | - | - | - | - | - | No |
| Q8NFI3 | CHEMBL5172 | ENASE | Drugged | Single protein | CHEMBL1269025 | Edoxaban | Small molecule | Inhibitor | ChEMBL | Indication | All | 4 | Embolism | No |
| Q8NFI3 | CHEMBL5172 | ENASE | Drugged | Single protein | CHEMBL1503 | Omeprazole | Small molecule | Inhibitor | ChEMBL | Indication | All | 3 | Hemorrhage | No |
| Q8NFI3 | CHEMBL5172 | ENASE | Drugged | Single protein | CHEMBL1503 | Omeprazole | Small molecule | Inhibitor | ChEMBL | Indication | All | 3 | Barrett esophagus | No |
| Q8NFI3 | CHEMBL5172 | ENASE | Drugged | Single protein | CHEMBL1503 | Omeprazole | Small molecule | Inhibitor | ChEMBL | Indication | All | 3 | Wounds and injuries | No |
| Q8NFI3 | CHEMBL5172 | ENASE | Drugged | Single protein | CHEMBL1503 | Omeprazole | Small molecule | Inhibitor | ChEMBL | Indication | All | 2 | Idiopathic pulmonary fibrosis | No |
| Q8NFI3 | CHEMBL5172 | ENASE | Drugged | Single protein | CHEMBL1201863 | Dexlansoprazole | Small molecule | Inhibitor | ChEMBL | Indication | All | 2 | Barrett esophagus | No |
| Q8NFI3 | CHEMBL5172 | ENASE | Drugged | Single protein | CHEMBL1201863 | Dexlansoprazole | Small molecule | Inhibitor | ChEMBL | Indication | All | 3 | Esophagitis, peptic | No |
| Q8NFI3 | CHEMBL5172 | ENASE | Drugged | Single protein | CHEMBL1201863 | Dexlansoprazole | Small molecule | Inhibitor | ChEMBL | Indication | All | 4 | Heartburn | No |
| Q8NFI3 | CHEMBL5172 | ENASE | Drugged | Single protein | CHEMBL1201863 | Dexlansoprazole | Small molecule | Inhibitor | ChEMBL | Indication | All | 4 | Esophagitis | No |
| Q8NFI3 | CHEMBL5172 | ENASE | Drugged | Single protein | CHEMBL1201863 | Dexlansoprazole | Small molecule | Inhibitor | ChEMBL | Indication | All | 4 | Gastroesophageal reflux | No |
| Q8NFI3 | CHEMBL5172 | ENASE | Drugged | Single protein | CHEMBL1502 | Pantoprazole | Small molecule | Inhibitor | ChEMBL | Indication | All | 1 | Lecithin cholesterol acyltransferase deficiency | No |
| Q8NFI3 | CHEMBL5172 | ENASE | Drugged | Single protein | CHEMBL1502 | Pantoprazole | Small molecule | Inhibitor | ChEMBL | Indication | All | 1 | Cancers | No |
| Q8NFI3 | CHEMBL5172 | ENASE | Drugged | Single protein | CHEMBL1502 | Pantoprazole | Small molecule | Inhibitor | ChEMBL | Indication | All | 4 | Gastroesophageal reflux | No |
| Q8NFI3 | CHEMBL5172 | ENASE | Drugged | Single protein | CHEMBL1502 | Pantoprazole | Small molecule | Inhibitor | ChEMBL | Indication | All | 4 | Zollinger-ellison syndrome | No |
| Q8NFI3 | CHEMBL5172 | ENASE | Drugged | Single protein | CHEMBL1502 | Pantoprazole | Small molecule | Inhibitor | ChEMBL | Indication | All | 4 | Esophagitis | No |
| Q8NFI3 | CHEMBL5172 | ENASE | Drugged | Single protein | CHEMBL1219 | Rabeprazole | Small molecule | Inhibitor | ChEMBL | Indication | All | 4 | Peptic ulcer | No |
| Q8NFI3 | CHEMBL5172 | ENASE | Drugged | Single protein | CHEMBL1219 | Rabeprazole | Small molecule | Inhibitor | ChEMBL | Indication | All | 2 | Esophagitis, peptic | No |
| Q8NFI3 | CHEMBL5172 | ENASE | Drugged | Single protein | CHEMBL1219 | Rabeprazole | Small molecule | Inhibitor | ChEMBL | Indication | All | 1 | Amyotrophic lateral sclerosis | No |
| Q8NFI3 | CHEMBL5172 | ENASE | Drugged | Single protein | CHEMBL1219 | Rabeprazole | Small molecule | Inhibitor | ChEMBL | Indication | All | 3 | Gastrointestinal hemorrhage | No |
| Q8NFI3 | CHEMBL5172 | ENASE | Drugged | Single protein | CHEMBL1219 | Rabeprazole | Small molecule | Inhibitor | ChEMBL | Indication | All | 3 | Cerebrovascular disorder | No |
| Q8NFI3 | CHEMBL5172 | ENASE | Drugged | Single protein | CHEMBL1219 | Rabeprazole | Small molecule | Inhibitor | ChEMBL | Indication | All | 1 | Cancers | No |
| Q8NFI3 | CHEMBL5172 | ENASE | Drugged | Single protein | CHEMBL1219 | Rabeprazole | Small molecule | Inhibitor | ChEMBL | Indication | All | 4 | Duodenal ulcer | No |
| Q8NFI3 | CHEMBL5172 | ENASE | Drugged | Single protein | CHEMBL1219 | Rabeprazole | Small molecule | Inhibitor | ChEMBL | Indication | All | 2 | Dyspepsia | No |
| Q8NFI3 | CHEMBL5172 | ENASE | Drugged | Single protein | CHEMBL1219 | Rabeprazole | Small molecule | Inhibitor | ChEMBL | Indication | All | 2 | Laryngopharyngeal reflux | No |
| Q8NFI3 | CHEMBL5172 | ENASE | Drugged | Single protein | CHEMBL1219 | Rabeprazole | Small molecule | Inhibitor | ChEMBL | Indication | All | 4 | Gastroesophageal reflux | No |
| Q8NFI3 | CHEMBL5172 | ENASE | Drugged | Single protein | CHEMBL1219 | Rabeprazole | Small molecule | Inhibitor | ChEMBL | Indication | All | 1 | Communicable diseases | No |
| Q8NFI3 | CHEMBL5172 | ENASE | Drugged | Single protein | CHEMBL1219 | Rabeprazole | Small molecule | Inhibitor | ChEMBL | Indication | All | 3 | Helicobacter infection | No |
| Q8NFI3 | CHEMBL5172 | ENASE | Drugged | Single protein | CHEMBL1219 | Rabeprazole | Small molecule | Inhibitor | ChEMBL | Indication | All | 4 | Zollinger-ellison syndrome | No |
| Q8NFI3 | CHEMBL5172 | ENASE | Drugged | Single protein | CHEMBL1503 | Omeprazole | Small molecule | Inhibitor | ChEMBL | Indication | All | 2 | Gastrinoma | No |
| Q8NFI3 | CHEMBL5172 | ENASE | Drugged | Single protein | CHEMBL480 | Lansoprazole | Small molecule | Inhibitor | ChEMBL | Indication | All | 2 | Premature birth | No |
| Q8NFI3 | CHEMBL5172 | ENASE | Drugged | Single protein | CHEMBL1503 | Omeprazole | Small molecule | Inhibitor | ChEMBL | Indication | All | 1 | Chronic pain | No |
| Q8NFI3 | CHEMBL5172 | ENASE | Drugged | Single protein | CHEMBL1503 | Omeprazole | Small molecule | Inhibitor | ChEMBL | Indication | All | 3 | Dyspepsia | No |
| Q8NFI3 | CHEMBL5172 | ENASE | Drugged | Single protein | CHEMBL1503 | Omeprazole | Small molecule | Inhibitor | ChEMBL | Indication | All | 4 | Stomach ulcer | No |
| Q8NFI3 | CHEMBL5172 | ENASE | Drugged | Single protein | CHEMBL1503 | Omeprazole | Small molecule | Inhibitor | ChEMBL | Indication | All | 3 | Pain | No |
| Q8NFI3 | CHEMBL5172 | ENASE | Drugged | Single protein | CHEMBL1503 | Omeprazole | Small molecule | Inhibitor | ChEMBL | Indication | All | 3 | Cysticercosis | No |
| Q8NFI3 | CHEMBL5172 | ENASE | Drugged | Single protein | CHEMBL1503 | Omeprazole | Small molecule | Inhibitor | ChEMBL | Indication | All | 4 | Gastroesophageal reflux | No |
| Q8NFI3 | CHEMBL5172 | ENASE | Drugged | Single protein | CHEMBL1503 | Omeprazole | Small molecule | Inhibitor | ChEMBL | Indication | All | 4 | Peptic ulcer | No |
| Q8NFI3 | CHEMBL5172 | ENASE | Drugged | Single protein | CHEMBL1503 | Omeprazole | Small molecule | Inhibitor | ChEMBL | Indication | All | 0,5 | Cancers | No |
| Q8NFI3 | CHEMBL5172 | ENASE | Drugged | Single protein | CHEMBL1503 | Omeprazole | Small molecule | Inhibitor | ChEMBL | Indication | All | 3 | Spondylitis, ankylosing | No |
| Q8NFI3 | CHEMBL5172 | ENASE | Drugged | Single protein | CHEMBL1503 | Omeprazole | Small molecule | Inhibitor | ChEMBL | Indication | All | 2 | Squamous intraepithelial lesions | No |
| Q8NFI3 | CHEMBL5172 | ENASE | Drugged | Single protein | CHEMBL1503 | Omeprazole | Small molecule | Inhibitor | ChEMBL | Indication | All | 2 | Cystic fibrosis | No |
| Q8NFI3 | CHEMBL5172 | ENASE | Drugged | Single protein | CHEMBL1503 | Omeprazole | Small molecule | Inhibitor | ChEMBL | Indication | All | 1 | Pulmonary disease, chronic obstructive | No |
| Q8NFI3 | CHEMBL5172 | ENASE | Drugged | Single protein | CHEMBL1503 | Omeprazole | Small molecule | Inhibitor | ChEMBL | Indication | All | 3 | Diabetes mellitus, type 1 | No |

**Table S8. Druggability results of the prioritised proteins**

| Target Uniprot ID* | Target ChEMBL ID | Target protein name | Druggability* | Target type | Drug ChEMBL ID | Drug name | Drug molecule type | Drug mechanism | Drug effect source | Drug effect type | Drug effect frequency | Max phase* | Drug effect | Drug effect class |
|---|---|---|---|---|---|---|---|---|---|---|---|---|---|---|
| Q8NFI3 | CHEMBL5172 | ENASE | Drugged | Single protein | CHEMBL1503 | Omeprazole | Small molecule | Inhibitor | ChEMBL | Indication | All | 1 | Diabetes mellitus, type 2 | No |
| Q8NFI3 | CHEMBL5172 | ENASE | Drugged | Single protein | CHEMBL1503 | Omeprazole | Small molecule | Inhibitor | ChEMBL | Indication | All | 4 | Heartburn | No |
| Q8NFI3 | CHEMBL5172 | ENASE | Drugged | Single protein | CHEMBL1503 | Omeprazole | Small molecule | Inhibitor | ChEMBL | Indication | All | 1 | Clostridium infection | No |
| Q8NFI3 | CHEMBL5172 | ENASE | Drugged | Single protein | CHEMBL1503 | Omeprazole | Small molecule | Inhibitor | ChEMBL | Indication | All | 3 | Exocrine pancreatic insufficiency | No |
| Q8NFI3 | CHEMBL5172 | ENASE | Drugged | Single protein | CHEMBL1503 | Omeprazole | Small molecule | Inhibitor | ChEMBL | Indication | All | 2 | Zollinger-ellison syndrome | No |
| Q8NFI3 | CHEMBL5172 | ENASE | Drugged | Single protein | CHEMBL1503 | Omeprazole | Small molecule | Inhibitor | ChEMBL | Indication | All | 1 | Cancers | No |
| Q8NFI3 | CHEMBL5172 | ENASE | Drugged | Single protein | CHEMBL1503 | Omeprazole | Small molecule | Inhibitor | ChEMBL | Indication | All | 1 | Alzheimer disease | No |
| Q8NFI3 | CHEMBL5172 | ENASE | Drugged | Single protein | CHEMBL1503 | Omeprazole | Small molecule | Inhibitor | ChEMBL | Indication | All | 1 | Colitis, ulcerative | No |
| Q8NFI3 | CHEMBL5172 | ENASE | Drugged | Single protein | CHEMBL1503 | Omeprazole | Small molecule | Inhibitor | ChEMBL | Indication | All | 1 | Child development disorder, pervasive | No |
| Q8NFI3 | CHEMBL5172 | ENASE | Drugged | Single protein | CHEMBL1503 | Omeprazole | Small molecule | Inhibitor | ChEMBL | Indication | All | 3 | Multiple sclerosis, relapsing-remitting | No |
| Q8NFI3 | CHEMBL5172 | ENASE | Drugged | Single protein | CHEMBL1503 | Omeprazole | Small molecule | Inhibitor | ChEMBL | Indication | All | 1 | Mitochondrial diseases | No |
| Q8NFI3 | CHEMBL5172 | ENASE | Drugged | Single protein | CHEMBL1503 | Omeprazole | Small molecule | Inhibitor | ChEMBL | Indication | All | 1 | Anemia | No |
| Q8NFI3 | CHEMBL5172 | ENASE | Drugged | Single protein | CHEMBL1503 | Omeprazole | Small molecule | Inhibitor | ChEMBL | Indication | All | 3 | Helicobacter infection | No |
| Q8NFI3 | CHEMBL5172 | ENASE | Drugged | Single protein | CHEMBL1503 | Omeprazole | Small molecule | Inhibitor | ChEMBL | Indication | All | 3 | Cough | No |
| Q8NFI3 | CHEMBL5172 | ENASE | Drugged | Single protein | CHEMBL1503 | Omeprazole | Small molecule | Inhibitor | ChEMBL | Indication | All | 4 | Esophagitis | No |
| Q8NFI3 | CHEMBL5172 | ENASE | Drugged | Single protein | CHEMBL1503 | Omeprazole | Small molecule | Inhibitor | ChEMBL | Indication | All | 1 | Erectile dysfunction | No |
| Q8NFI3 | CHEMBL5172 | ENASE | Drugged | Single protein | CHEMBL1503 | Omeprazole | Small molecule | Inhibitor | ChEMBL | Indication | All | 1 | Muscular atrophy, spinal | No |
| Q8NFI3 | CHEMBL5172 | ENASE | Drugged | Single protein | CHEMBL1503 | Omeprazole | Small molecule | Inhibitor | ChEMBL | Indication | All | 2 | Granuloma | No |
| Q8NFI3 | CHEMBL5172 | ENASE | Drugged | Single protein | CHEMBL480 | Lansoprazole | Small molecule | Inhibitor | ChEMBL | Indication | All | 1 | Cystic fibrosis | No |
| Q8NFI3 | CHEMBL5172 | ENASE | Drugged | Single protein | CHEMBL480 | Lansoprazole | Small molecule | Inhibitor | ChEMBL | Indication | All | 3 | Duodenal ulcer | No |
| Q8NFI3 | CHEMBL5172 | ENASE | Drugged | Single protein | CHEMBL480 | Lansoprazole | Small molecule | Inhibitor | ChEMBL | Indication | All | 3 | Gastritis | No |
| Q8NFI3 | CHEMBL5172 | ENASE | Drugged | Single protein | CHEMBL2141296 | Ixazomib | Small molecule | Inhibitor | ChEMBL | Indication | All | 2 | Waldenstrom macroglobulinemia | No |
| Q8NFI3 | CHEMBL5172 | ENASE | Drugged | Single protein | CHEMBL2141296 | Ixazomib | Small molecule | Inhibitor | ChEMBL | Indication | All | 1 | Hiv infection | No |
| Q8NFI3 | CHEMBL5172 | ENASE | Drugged | Single protein | CHEMBL2141296 | Ixazomib | Small molecule | Inhibitor | ChEMBL | Indication | All | 2 | Cancers | No |
| Q8NFI3 | CHEMBL5172 | ENASE | Drugged | Single protein | CHEMBL641 | Atomoxetine | Small molecule | Inhibitor | ChEMBL | Indication | All | 3 | Attention deficit and disruptive behavior disorder | No |
| Q8NFI3 | CHEMBL5172 | ENASE | Drugged | Single protein | CHEMBL641 | Atomoxetine | Small molecule | Inhibitor | ChEMBL | Indication | All | 4 | Attention deficit disorder with hyperactivity | No |
| Q8NFI3 | CHEMBL5172 | ENASE | Drugged | Single protein | CHEMBL641 | Atomoxetine | Small molecule | Inhibitor | ChEMBL | Indication | All | 2 | Alzheimer disease | No |
| Q8NFI3 | CHEMBL5172 | ENASE | Drugged | Single protein | CHEMBL1738 | Dexrazoxane | Small molecule | - | ChEMBL | Indication | All | 4 | Cardiomyopathies | Yes |
| Q8NFI3 | CHEMBL5172 | ENASE | Drugged | Single protein | CHEMBL1738 | Dexrazoxane | Small molecule | - | ChEMBL | Indication | All | 1 | Cancers | No |
| Q8NFI3 | CHEMBL5172 | ENASE | Drugged | Single protein | CHEMBL1503 | Omeprazole | Small molecule | Inhibitor | ChEMBL | Indication | All | 4 | Duodenal ulcer | No |
| Q8NFI3 | CHEMBL5172 | ENASE | Drugged | Single protein | CHEMBL1738 | Dexrazoxane | Small molecule | - | ChEMBL | Indication | All | 4 | Cancers | No |
| Q8NFI3 | CHEMBL5172 | ENASE | Drugged | Single protein | CHEMBL1738 | Dexrazoxane | Small molecule | - | ChEMBL | Indication | All | 3 | Cancers | No |
| Q8NFI3 | CHEMBL5172 | ENASE | Drugged | Single protein | CHEMBL1738 | Dexrazoxane | Small molecule | - | ChEMBL | Indication | All | 2 | Cancers | No |
| Q8NFI3 | CHEMBL5172 | ENASE | Drugged | Single protein | CHEMBL1269025 | Edoxaban | Small molecule | Inhibitor | ChEMBL | Indication | All | 4 | Stroke | No |
| Q8NFI3 | CHEMBL5172 | ENASE | Drugged | Single protein | CHEMBL2141296 | Ixazomib | Small molecule | Inhibitor | ChEMBL | Indication | All | 2 | Renal insufficiency | No |
| Q8NFI3 | CHEMBL5172 | ENASE | Drugged | Single protein | CHEMBL1269025 | Edoxaban | Small molecule | Inhibitor | ChEMBL | Indication | All | 3 | Severe acute respiratory syndrome | No |
| Q8NFI3 | CHEMBL5172 | ENASE | Drugged | Single protein | CHEMBL1269025 | Edoxaban | Small molecule | Inhibitor | ChEMBL | Indication | All | 2 | Coronary disease | Yes |
| Q8NFI3 | CHEMBL5172 | ENASE | Drugged | Single protein | CHEMBL1269025 | Edoxaban | Small molecule | Inhibitor | ChEMBL | Indication | All | 2 | Myocardial infarction | Yes |
| Q8NFI3 | CHEMBL5172 | ENASE | Drugged | Single protein | CHEMBL1269025 | Edoxaban | Small molecule | Inhibitor | ChEMBL | Indication | All | 3 | Venous thromboembolism | No |
| Q8NFI3 | CHEMBL5172 | ENASE | Drugged | Single protein | CHEMBL1269025 | Edoxaban | Small molecule | Inhibitor | ChEMBL | Indication | All | 4 | Thrombosis | No |
| Q8NFI3 | CHEMBL5172 | ENASE | Drugged | Single protein | CHEMBL1201863 | Dexlansoprazole | Small molecule | Inhibitor | ChEMBL | Indication | All | 4 | Duodenal ulcer | No |
| Q8NFI3 | CHEMBL5172 | ENASE | Drugged | Single protein | CHEMBL1201863 | Dexlansoprazole | Small molecule | Inhibitor | ChEMBL | Indication | All | 4 | Stomach ulcer | No |
| Q8NFI3 | CHEMBL5172 | ENASE | Drugged | Single protein | CHEMBL1201863 | Dexlansoprazole | Small molecule | Inhibitor | ChEMBL | Indication | All | 4 | Peptic ulcer | No |
| Q8NFI3 | CHEMBL5172 | ENASE | Drugged | Single protein | CHEMBL1269025 | Edoxaban | Small molecule | Inhibitor | ChEMBL | Indication | All | 4 | Atrial fibrillation | Yes |
| Q8NFI3 | CHEMBL5172 | ENASE | Drugged | Single protein | CHEMBL1269025 | Edoxaban | Small molecule | Inhibitor | ChEMBL | Indication | All | 2 | Hemorrhage | No |
| Q8NFI3 | CHEMBL5172 | ENASE | Drugged | Single protein | CHEMBL1269025 | Edoxaban | Small molecule | Inhibitor | ChEMBL | Indication | All | 3 | Aortic valve stenosis | Yes |
| Q8NFI3 | CHEMBL5172 | ENASE | Drugged | Single protein | CHEMBL1269025 | Edoxaban | Small molecule | Inhibitor | ChEMBL | Indication | All | 3 | Heart diseases | Yes |
| Q8NFI3 | CHEMBL5172 | ENASE | Drugged | Single protein | CHEMBL1269025 | Edoxaban | Small molecule | Inhibitor | ChEMBL | Indication | All | 2 | Peripheral arterial disease | No |
| Q8NFI3 | CHEMBL5172 | ENASE | Drugged | Single protein | CHEMBL1269025 | Edoxaban | Small molecule | Inhibitor | ChEMBL | Indication | All | 2 | Ischemic stroke | Yes |
| Q8NFI3 | CHEMBL5172 | ENASE | Drugged | Single protein | CHEMBL1269025 | Edoxaban | Small molecule | Inhibitor | ChEMBL | Indication | All | 2 | Blood coagulation disorder | No |
| Q8NFI3 | CHEMBL5172 | ENASE | Drugged | Single protein | CHEMBL2141296 | Ixazomib | Small molecule | Inhibitor | ChEMBL | Indication | All | 2 | Anemia | No |
| Q8NFI3 | CHEMBL5172 | ENASE | Drugged | Single protein | CHEMBL2141296 | Ixazomib | Small molecule | Inhibitor | ChEMBL | Indication | All | 1 | Cancers | No |
| Q8NFI3 | CHEMBL5172 | ENASE | Drugged | Single protein | CHEMBL2141296 | Ixazomib | Small molecule | Inhibitor | ChEMBL | Indication | All | 3 | Amyloidosis, familial | No |
| Q8NFI3 | CHEMBL5172 | ENASE | Drugged | Single protein | CHEMBL480 | Lansoprazole | Small molecule | Inhibitor | ChEMBL | Indication | All | 3 | Idiopathic pulmonary fibrosis | No |
| Q8NFI3 | CHEMBL5172 | ENASE | Drugged | Single protein | CHEMBL480 | Lansoprazole | Small molecule | Inhibitor | ChEMBL | Indication | All | 3 | Arthritis | No |
| Q8NFI3 | CHEMBL5172 | ENASE | Drugged | Single protein | CHEMBL480 | Lansoprazole | Small molecule | Inhibitor | ChEMBL | Indication | All | 3 | Esophagitis, peptic | No |
| Q8NFI3 | CHEMBL5172 | ENASE | Drugged | Single protein | CHEMBL480 | Lansoprazole | Small molecule | Inhibitor | ChEMBL | Indication | All | 3 | Helicobacter infection | No |
| Q8NFI3 | CHEMBL5172 | ENASE | Drugged | Single protein | CHEMBL480 | Lansoprazole | Small molecule | Inhibitor | ChEMBL | Indication | All | 4 | Peptic ulcer | No |
| Q8NFI3 | CHEMBL5172 | ENASE | Drugged | Single protein | CHEMBL480 | Lansoprazole | Small molecule | Inhibitor | ChEMBL | Indication | All | 1 | Laryngopharyngeal reflux | No |
| Q8NFI3 | CHEMBL5172 | ENASE | Drugged | Single protein | CHEMBL480 | Lansoprazole | Small molecule | Inhibitor | ChEMBL | Indication | All | 3 | Diabetes mellitus, type 1 | No |
| Q8NFI3 | CHEMBL5172 | ENASE | Drugged | Single protein | CHEMBL480 | Lansoprazole | Small molecule | Inhibitor | ChEMBL | Indication | All | 2 | Eosinophilic esophagitis | No |
| Q8NFI3 | CHEMBL5172 | ENASE | Drugged | Single protein | CHEMBL480 | Lansoprazole | Small molecule | Inhibitor | ChEMBL | Indication | All | 1 | Cancers | No |
| Q8NFI3 | CHEMBL5172 | ENASE | Drugged | Single protein | CHEMBL480 | Lansoprazole | Small molecule | Inhibitor | ChEMBL | Indication | All | 1 | Pain | No |
| Q8NFI3 | CHEMBL5172 | ENASE | Drugged | Single protein | CHEMBL480 | Lansoprazole | Small molecule | Inhibitor | ChEMBL | Indication | All | 4 | Gastroesophageal reflux | No |
| Q8NFI3 | CHEMBL5172 | ENASE | Drugged | Single protein | CHEMBL480 | Lansoprazole | Small molecule | Inhibitor | ChEMBL | Indication | All | 4 | Stomach ulcer | No |
| Q8NFI3 | CHEMBL5172 | ENASE | Drugged | Single protein | CHEMBL480 | Lansoprazole | Small molecule | Inhibitor | ChEMBL | Indication | All | 3 | Cancers | No |
| Q8NFI3 | CHEMBL5172 | ENASE | Drugged | Single protein | CHEMBL480 | Lansoprazole | Small molecule | Inhibitor | ChEMBL | Indication | All | 2 | Cancers | No |
| Q8NFI3 | CHEMBL5172 | ENASE | Drugged | Single protein | CHEMBL480 | Lansoprazole | Small molecule | Inhibitor | ChEMBL | Indication | All | 3 | Gout | No |
| Q8NFI3 | CHEMBL5172 | ENASE | Drugged | Single protein | CHEMBL480 | Lansoprazole | Small molecule | Inhibitor | ChEMBL | Indication | All | 2 | Pseudomyxoma peritonei | No |
| Q8NFI3 | CHEMBL5172 | ENASE | Drugged | Single protein | CHEMBL862 | Guanfacine | Small molecule | Agonist | ChEMBL | Indication | All | 4 | Attention deficit disorder with hyperactivity | No |
| Q8NFI3 | CHEMBL5172 | ENASE | Drugged | Single protein | CHEMBL49 | Buspirone | Small molecule | Partial agonist | ChEMBL | Indication | All | 2 | Sexual dysfunction, physiological | No |
| Q8NFI3 | CHEMBL5172 | ENASE | Drugged | Single protein | CHEMBL49 | Buspirone | Small molecule | Partial agonist | ChEMBL | Indication | All | 2 | Postoperative nausea and vomiting | No |
| Q8NFI3 | CHEMBL5172 | ENASE | Drugged | Single protein | CHEMBL49 | Buspirone | Small molecule | Partial agonist | ChEMBL | Indication | All | 2 | Spinal cord injuries | No |
| Q8NFI3 | CHEMBL5172 | ENASE | Drugged | Single protein | CHEMBL49 | Buspirone | Small molecule | Partial agonist | ChEMBL | Indication | All | 3 | Psychotic disorder | No |
| Q8NFI3 | CHEMBL5172 | ENASE | Drugged | Single protein | CHEMBL49 | Buspirone | Small molecule | Partial agonist | ChEMBL | Indication | All | 4 | Anxiety disorder | No |
| Q8NFI3 | CHEMBL5172 | ENASE | Drugged | Single protein | CHEMBL49 | Buspirone | Small molecule | Partial agonist | ChEMBL | Indication | All | 4 | Anxiety | No |
| Q8NFI3 | CHEMBL5172 | ENASE | Drugged | Single protein | CHEMBL49 | Buspirone | Small molecule | Partial agonist | ChEMBL | Indication | All | 3 | Schizophrenia | No |
| Q8NFI3 | CHEMBL5172 | ENASE | Drugged | Single protein | CHEMBL2141296 | Ixazomib | Small molecule | Inhibitor | ChEMBL | Indication | All | 3 | Plasmacytoma | No |
| Q8NFI3 | CHEMBL5172 | ENASE | Drugged | Single protein | CHEMBL2141296 | Ixazomib | Small molecule | Inhibitor | ChEMBL | Indication | All | 1 | Lupus nephritis | No |
| Q8NFI3 | CHEMBL5172 | ENASE | Drugged | Single protein | CHEMBL2141296 | Ixazomib | Small molecule | Inhibitor | ChEMBL | Indication | All | 2 | Purpura, thrombocytopenic, idiopathic | No |
| Q8NFI3 | CHEMBL5172 | ENASE | Drugged | Single protein | CHEMBL2141296 | Ixazomib | Small molecule | Inhibitor | ChEMBL | Indication | All | 4 | Cancers | No |
| Q8NFI3 | CHEMBL5172 | ENASE | Drugged | Single protein | CHEMBL2141296 | Ixazomib | Small molecule | Inhibitor | ChEMBL | Indication | All | 3 | Cancers | No |
| Q8NFI3 | CHEMBL5172 | ENASE | Drugged | Single protein | CHEMBL1269025 | Edoxaban | Small molecule | Inhibitor | ChEMBL | Indication | All | 4 | Venous thrombosis | No |
| Q8NFI3 | CHEMBL5172 | ENASE | Drugged | Single protein | CHEMBL1503 | Omeprazole | Small molecule | Inhibitor | ChEMBL | Indication | All | 1 | Laryngomalacia | No |
| Q8NFI3 | CHEMBL5172 | ENASE | Drugged | Single protein | CHEMBL1503 | Omeprazole | Small molecule | Inhibitor | ChEMBL | Indication | All | 1 | Central serous chorioretinopathy | No |
| Q8NFI3 | CHEMBL5172 | ENASE | Drugged | Single protein | CHEMBL1503 | Omeprazole | Small molecule | Inhibitor | ChEMBL | Indication | All | 2 | Ulcer | No |
| Q8NFI3 | CHEMBL5172 | ENASE | Drugged | Single protein | CHEMBL20 | Acetazolamide | Small molecule | Inhibitor | ChEMBL | Indication | All | 2 | Landau-kleffner syndrome | No |

**Table S8. Druggability results of the prioritised proteins**

| Target Uniprot ID* | Target ChEMBL ID | Target protein name | Druggability* | Target type | Drug ChEMBL ID | Drug name | Drug molecule type | Drug mechanism | Drug effect source | Drug effect type | Drug effect frequency | Max phase* | Drug effect | Drug effect class |
|---|---|---|---|---|---|---|---|---|---|---|---|---|---|---|
| Q8NFI3 | CHEMBL5172 | ENASE | Drugged | Single protein | CHEMBL20 | Acetazolamide | Small molecule | Inhibitor | ChEMBL | Indication | All | 2 | Helicobacter infection | No |
| Q8NFI3 | CHEMBL5172 | ENASE | Drugged | Single protein | CHEMBL20 | Acetazolamide | Small molecule | Inhibitor | ChEMBL | Indication | All | 2 | Macular edema | No |
| Q8NFI3 | CHEMBL5172 | ENASE | Drugged | Single protein | CHEMBL20 | Acetazolamide | Small molecule | Inhibitor | ChEMBL | Indication | All | 3 | Apnea | No |
| Q8NFI3 | CHEMBL5172 | ENASE | Drugged | Single protein | CHEMBL20 | Acetazolamide | Small molecule | Inhibitor | ChEMBL | Indication | All | 2 | Cysticercosis | No |
| Q8NFI3 | CHEMBL5172 | ENASE | Drugged | Single protein | CHEMBL20 | Acetazolamide | Small molecule | Inhibitor | ChEMBL | Indication | All | 2 | Pulmonary hypertension | Yes |
| Q8NFI3 | CHEMBL5172 | ENASE | Drugged | Single protein | CHEMBL20 | Acetazolamide | Small molecule | Inhibitor | ChEMBL | Indication | All | 1 | Schizophrenia | No |
| Q8NFI3 | CHEMBL5172 | ENASE | Drugged | Single protein | CHEMBL20 | Acetazolamide | Small molecule | Inhibitor | ChEMBL | Indication | All | 2 | Status epilepticus | No |
| Q8NFI3 | CHEMBL5172 | ENASE | Drugged | Single protein | CHEMBL20 | Acetazolamide | Small molecule | Inhibitor | ChEMBL | Indication | All | 3 | Hydrocephalus, normal pressure | No |
| Q8NFI3 | CHEMBL5172 | ENASE | Drugged | Single protein | CHEMBL20 | Acetazolamide | Small molecule | Inhibitor | ChEMBL | Indication | All | 2 | Multiple sclerosis | No |
| Q8NFI3 | CHEMBL5172 | ENASE | Drugged | Single protein | CHEMBL20 | Acetazolamide | Small molecule | Inhibitor | ChEMBL | Indication | All | 4 | Edema | No |
| Q8NFI3 | CHEMBL5172 | ENASE | Drugged | Single protein | CHEMBL20 | Acetazolamide | Small molecule | Inhibitor | ChEMBL | Indication | All | 1 | Diabetes mellitus, type 1 | No |
| Q8NFI3 | CHEMBL5172 | ENASE | Drugged | Single protein | CHEMBL20 | Acetazolamide | Small molecule | Inhibitor | ChEMBL | Indication | All | 2 | Chronic pain | No |
| Q8NFI3 | CHEMBL5172 | ENASE | Drugged | Single protein | CHEMBL20 | Acetazolamide | Small molecule | Inhibitor | ChEMBL | Indication | All | 1 | Cancers | No |
| Q8NFI3 | CHEMBL5172 | ENASE | Drugged | Single protein | CHEMBL20 | Acetazolamide | Small molecule | Inhibitor | ChEMBL | Indication | All | 2 | Kidney diseases | No |
| Q8NFI3 | CHEMBL5172 | ENASE | Drugged | Single protein | CHEMBL20 | Acetazolamide | Small molecule | Inhibitor | ChEMBL | Indication | All | 1 | Andersen syndrome | No |
| Q8NFI3 | CHEMBL5172 | ENASE | Drugged | Single protein | CHEMBL20 | Acetazolamide | Small molecule | Inhibitor | ChEMBL | Indication | All | 4 | Cancers | No |
| Q8NFI3 | CHEMBL5172 | ENASE | Drugged | Single protein | CHEMBL20 | Acetazolamide | Small molecule | Inhibitor | ChEMBL | Indication | All | 4 | Heart failure | Yes |
| Q8NFI3 | CHEMBL5172 | ENASE | Drugged | Single protein | CHEMBL20 | Acetazolamide | Small molecule | Inhibitor | ChEMBL | Indication | All | 4 | Glaucoma, angle-closure | No |
| Q8NFI3 | CHEMBL5172 | ENASE | Drugged | Single protein | CHEMBL20 | Acetazolamide | Small molecule | Inhibitor | ChEMBL | Indication | All | 4 | Seizures | No |
| Q8NFI3 | CHEMBL5172 | ENASE | Drugged | Single protein | CHEMBL1269025 | Edoxaban | Small molecule | Inhibitor | ChEMBL | Indication | All | 4 | Pulmonary embolism | No |
| Q8NFI3 | CHEMBL5172 | ENASE | Drugged | Single protein | CHEMBL20 | Acetazolamide | Small molecule | Inhibitor | ChEMBL | Indication | All | 2 | Sleep apnea, obstructive | No |
| Q8NFI3 | CHEMBL5172 | ENASE | Drugged | Single protein | CHEMBL1503 | Omeprazole | Small molecule | Inhibitor | ChEMBL | Indication | All | 1 | Hepatitis b, chronic | No |
| Q8NFI3 | CHEMBL5172 | ENASE | Drugged | Single protein | CHEMBL1503 | Omeprazole | Small molecule | Inhibitor | ChEMBL | Indication | All | 3 | Arthritis | No |
| Q8NFI3 | CHEMBL5172 | ENASE | Drugged | Single protein | CHEMBL1503 | Omeprazole | Small molecule | Inhibitor | ChEMBL | Indication | All | 1 | Communicable diseases | No |
| Q8NFI3 | CHEMBL5172 | ENASE | Drugged | Single protein | CHEMBL1503 | Omeprazole | Small molecule | Inhibitor | ChEMBL | Indication | All | 3 | Gastrointestinal hemorrhage | No |
| Q8NFI3 | CHEMBL5172 | ENASE | Drugged | Single protein | CHEMBL1503 | Omeprazole | Small molecule | Inhibitor | ChEMBL | Indication | All | 4 | Infections | No |
| Q8NFI3 | CHEMBL5172 | ENASE | Drugged | Single protein | CHEMBL1503 | Omeprazole | Small molecule | Inhibitor | ChEMBL | Indication | All | 1 | Arthritis | No |
| Q8NFI3 | CHEMBL5172 | ENASE | Drugged | Single protein | CHEMBL1503 | Omeprazole | Small molecule | Inhibitor | ChEMBL | Indication | All | 1 | Psoriasis | No |
| Q8NFI3 | CHEMBL5172 | ENASE | Drugged | Single protein | CHEMBL1503 | Omeprazole | Small molecule | Inhibitor | ChEMBL | Indication | All | 1 | Crohn disease | No |
| Q8NFI3 | CHEMBL5172 | ENASE | Drugged | Single protein | CHEMBL1503 | Omeprazole | Small molecule | Inhibitor | ChEMBL | Indication | All | 2 | Cancers | No |
| Q8NFI3 | CHEMBL5172 | ENASE | Drugged | Single protein | CHEMBL1503 | Omeprazole | Small molecule | Inhibitor | ChEMBL | Indication | All | 1 | Hepatitis c | No |
| Q8NFI3 | CHEMBL5172 | ENASE | Drugged | Single protein | CHEMBL1503 | Omeprazole | Small molecule | Inhibitor | ChEMBL | Indication | All | 1 | Dermatitis, atopic | No |
| Q8NFI3 | CHEMBL5172 | ENASE | Drugged | Single protein | CHEMBL1503 | Omeprazole | Small molecule | Inhibitor | ChEMBL | Indication | All | 1 | Diabetes mellitus | No |
| Q8NFI3 | CHEMBL5172 | ENASE | Drugged | Single protein | CHEMBL1503 | Omeprazole | Small molecule | Inhibitor | ChEMBL | Indication | All | 1 | Hiv infection | No |
| Q8NFI3 | CHEMBL5172 | ENASE | Drugged | Single protein | CHEMBL1503 | Omeprazole | Small molecule | Inhibitor | ChEMBL | Indication | All | 3 | Esophagitis, peptic | No |
| Q8NFI3 | CHEMBL5172 | ENASE | Drugged | Single protein | CHEMBL20 | Acetazolamide | Small molecule | Inhibitor | ChEMBL | Indication | All | 2 | Stroke | No |
| Q8NFI3 | CHEMBL5172 | ENASE | Drugged | Single protein | CHEMBL20 | Acetazolamide | Small molecule | Inhibitor | ChEMBL | Indication | All | 4 | Epilepsy | No |
| Q8NFI3 | CHEMBL5172 | ENASE | Drugged | Single protein | CHEMBL20 | Acetazolamide | Small molecule | Inhibitor | ChEMBL | Indication | All | 3 | Cancers | No |
| Q8NFI3 | CHEMBL5172 | ENASE | Drugged | Single protein | CHEMBL20 | Acetazolamide | Small molecule | Inhibitor | ChEMBL | Indication | All | 1 | Hypotension, orthostatic | No |
| Q8NFI3 | CHEMBL5172 | ENASE | Drugged | Single protein | CHEMBL1503 | Omeprazole | Small molecule | Inhibitor | ChEMBL | Indication | All | 3 | Cancers | No |
| Q8NFI3 | CHEMBL5172 | ENASE | Drugged | Single protein | CHEMBL20 | Acetazolamide | Small molecule | Inhibitor | ChEMBL | Indication | All | 4 | Altitude sickness | No |
| P13929 | - | ENOB | Not yet druggable | - | - | - | - | - | - | - | - | - | - | No |
| O75356 | CHEMBL4523151 | ENTP5 | Drugged | Single protein | CHEMBL46469 | Anthralin | Small molecule | - | ChEMBL | Indication | All | 4 | Psoriasis | No |
| P54762 | CHEMBL2363043 | EPHB1 | Drugged | Protein family | CHEMBL24828 | Vandetanib | Small molecule | Inhibitor | ChEMBL | Indication | All | 3 | Cancers | No |
| P54762 | CHEMBL2363043 | EPHB1 | Drugged | Protein family | CHEMBL24828 | Vandetanib | Small molecule | Inhibitor | ChEMBL | Indication | All | 4 | Cancers | No |
| P54762 | CHEMBL2363043 | EPHB1 | Drugged | Protein family | CHEMBL24828 | Vandetanib | Small molecule | Inhibitor | ChEMBL | Indication | All | 2 | Cancers | No |
| P54762 | CHEMBL2363043 | EPHB1 | Drugged | Protein family | CHEMBL24828 | Vandetanib | Small molecule | Inhibitor | ChEMBL | Indication | All | 1 | Cancers | No |
| Q9NZ08 | CHEMBL5939 | ERAP1 | Druggable | Single protein | - | - | - | - | - | - | - | - | - | No |
| Q9NZ08 | CHEMBL3831223 | ERAP1 | Druggable | Protein family | CHEMBL2103847 | Tosedostat | Small molecule | Inhibitor | ChEMBL | Indication | All | 2 | Cancers | No |
| Q9NZ08 | CHEMBL3831223 | ERAP1 | Druggable | Protein family | CHEMBL2103847 | Tosedostat | Small molecule | Inhibitor | ChEMBL | Indication | All | 1 | Cancers | No |
| P00742 | CHEMBL244 | FA10 | Drugged | Single protein | CHEMBL231779 | Apixaban | Small molecule | Inhibitor | ChEMBL | Indication | All | 3 | Atrial flutter | Yes |
| P00742 | CHEMBL244 | FA10 | Drugged | Single protein | CHEMBL231779 | Apixaban | Small molecule | Inhibitor | ChEMBL | Indication | All | 3 | Aortic valve stenosis | Yes |
| P00742 | CHEMBL244 | FA10 | Drugged | Single protein | CHEMBL231779 | Apixaban | Small molecule | Inhibitor | ChEMBL | Indication | All | 3 | Anemia | No |
| P00742 | CHEMBL244 | FA10 | Drugged | Single protein | CHEMBL231779 | Apixaban | Small molecule | Inhibitor | ChEMBL | Indication | All | 2 | Takotsubo cardiomyopathy | Yes |
| P00742 | CHEMBL244 | FA10 | Drugged | Single protein | CHEMBL198362 | Rivaroxaban | Small molecule | Inhibitor | BNF | Side-effect | Common or very common | 4 | Anemia | No |
| P00742 | CHEMBL244 | FA10 | Drugged | Single protein | CHEMBL198362 | Rivaroxaban | Small molecule | Inhibitor | BNF | Indication | All | 4 | Prophylaxis of recurrent deep-vein thrombosis | No |
| P00742 | CHEMBL244 | FA10 | Drugged | Single protein | CHEMBL198362 | Rivaroxaban | Small molecule | Inhibitor | BNF | Indication | All | 4 | Prophylaxis of recurrent pulmonary embolism | No |
| P00742 | CHEMBL244 | FA10 | Drugged | Single protein | CHEMBL198362 | Rivaroxaban | Small molecule | Inhibitor | BNF | Indication | All | 4 | Prophylaxis of stroke and systemic embolism | No |
| P00742 | CHEMBL244 | FA10 | Drugged | Single protein | CHEMBL198362 | Rivaroxaban | Small molecule | Inhibitor | BNF | Indication | All | 4 | Prophylaxis of venous thromboembolism following hip replacement surgery | No |
| P00742 | CHEMBL244 | FA10 | Drugged | Single protein | CHEMBL198362 | Rivaroxaban | Small molecule | Inhibitor | BNF | Indication | All | 4 | Prophylaxis of venous thromboembolism following knee replacement surgery | No |
| P00742 | CHEMBL244 | FA10 | Drugged | Single protein | CHEMBL198362 | Rivaroxaban | Small molecule | Inhibitor | BNF | Indication | All | 4 | Treatment of deep-vein thrombosis | No |
| P00742 | CHEMBL244 | FA10 | Drugged | Single protein | CHEMBL198362 | Rivaroxaban | Small molecule | Inhibitor | BNF | Indication | All | 4 | Treatment of pulmonary embolism | No |
| P00742 | CHEMBL244 | FA10 | Drugged | Single protein | CHEMBL198362 | Rivaroxaban | Small molecule | Inhibitor | BNF | Indication | All | 4 | Prophylaxis of atherothrombotic events following an acute coronary syndrome with elevated cardiac biomarkers | No |
| P00742 | CHEMBL244 | FA10 | Drugged | Single protein | CHEMBL198362 | Rivaroxaban | Small molecule | Inhibitor | BNF | Indication | All | 4 | Prophylaxis of atherothrombotic events | No |
| P00742 | CHEMBL244 | FA10 | Drugged | Single protein | CHEMBL231779 | Apixaban | Small molecule | Inhibitor | BNF | Indication | All | 4 | Treatment of deep-vein thrombosis | No |
| P00742 | CHEMBL244 | FA10 | Drugged | Single protein | CHEMBL231779 | Apixaban | Small molecule | Inhibitor | BNF | Side-effect | Uncommon | 4 | Thrombocytopenia | No |
| P00742 | CHEMBL244 | FA10 | Drugged | Single protein | CHEMBL4297951 | Coagulation factor x human | Unknown | Positive modulator | ChEMBL | Indication | All | 4 | Hemorrhage | No |
| P00742 | CHEMBL244 | FA10 | Drugged | Single protein | CHEMBL231779 | Apixaban | Small molecule | Inhibitor | BNF | Indication | All | 4 | Prophylaxis of recurrent deep-vein thrombosis | No |
| P00742 | CHEMBL244 | FA10 | Drugged | Single protein | CHEMBL231779 | Apixaban | Small molecule | Inhibitor | BNF | Indication | All | 4 | Prophylaxis of recurrent pulmonary embolism | No |
| P00742 | CHEMBL244 | FA10 | Drugged | Single protein | CHEMBL231779 | Apixaban | Small molecule | Inhibitor | BNF | Indication | All | 4 | Prophylaxis of stroke and systemic embolism in non-valvular atrial fibrillation and at least one risk factor | No |
| P00742 | CHEMBL244 | FA10 | Drugged | Single protein | CHEMBL231779 | Apixaban | Small molecule | Inhibitor | BNF | Indication | All | 4 | Prophylaxis of venous thromboembolism following hip replacement surgery | No |
| P00742 | CHEMBL244 | FA10 | Drugged | Single protein | CHEMBL231779 | Apixaban | Small molecule | Inhibitor | BNF | Indication | All | 4 | Prophylaxis of venous thromboembolism following knee replacement surgery | No |
| P00742 | CHEMBL244 | FA10 | Drugged | Single protein | CHEMBL198362 | Rivaroxaban | Small molecule | Inhibitor | BNF | Side-effect | Common or very common | 4 | Asthenia | No |
| P00742 | CHEMBL244 | FA10 | Drugged | Single protein | CHEMBL231779 | Apixaban | Small molecule | Inhibitor | BNF | Indication | All | 4 | Treatment of pulmonary embolism | No |
| P00742 | CHEMBL244 | FA10 | Drugged | Single protein | CHEMBL231779 | Apixaban | Small molecule | Inhibitor | BNF | Side-effect | Common or very common | 4 | Anemia | No |
| P00742 | CHEMBL244 | FA10 | Drugged | Single protein | CHEMBL231779 | Apixaban | Small molecule | Inhibitor | BNF | Side-effect | Common or very common | 4 | Hemorrhage | No |
| P00742 | CHEMBL244 | FA10 | Drugged | Single protein | CHEMBL231779 | Apixaban | Small molecule | Inhibitor | BNF | Side-effect | Common or very common | 4 | Nausea | No |
| P00742 | CHEMBL244 | FA10 | Drugged | Single protein | CHEMBL231779 | Apixaban | Small molecule | Inhibitor | BNF | Side-effect | Common or very common | 4 | Skin reactions | No |
| P00742 | CHEMBL244 | FA10 | Drugged | Single protein | CHEMBL231779 | Apixaban | Small molecule | Inhibitor | BNF | Side-effect | Uncommon | 4 | Cns hemorrhage | No |
| P00742 | CHEMBL244 | FA10 | Drugged | Single protein | CHEMBL231779 | Apixaban | Small molecule | Inhibitor | BNF | Side-effect | Uncommon | 4 | Hypotension | No |
| P00742 | CHEMBL244 | FA10 | Drugged | Single protein | CHEMBL231779 | Apixaban | Small molecule | Inhibitor | BNF | Side-effect | Uncommon | 4 | Post procedural hematoma | No |
| P00742 | CHEMBL244 | FA10 | Drugged | Single protein | CHEMBL231779 | Apixaban | Small molecule | Inhibitor | BNF | Side-effect | Uncommon | 4 | Wound complications | No |
| P00742 | CHEMBL244 | FA10 | Drugged | Single protein | CHEMBL198362 | Rivaroxaban | Small molecule | Inhibitor | BNF | Side-effect | Common or very common | 4 | Constipation | No |
| P00742 | CHEMBL244 | FA10 | Drugged | Single protein | CHEMBL198362 | Rivaroxaban | Small molecule | Inhibitor | BNF | Side-effect | Common or very common | 4 | Pain in extremity | No |
| P00742 | CHEMBL244 | FA10 | Drugged | Single protein | CHEMBL198362 | Rivaroxaban | Small molecule | Inhibitor | BNF | Side-effect | Common or very common | 4 | Dizziness | No |
| P00742 | CHEMBL244 | FA10 | Drugged | Single protein | CHEMBL198362 | Rivaroxaban | Small molecule | Inhibitor | BNF | Side-effect | Uncommon | 4 | Hypersensitivity | No |
| P00742 | CHEMBL244 | FA10 | Drugged | Single protein | CHEMBL198362 | Rivaroxaban | Small molecule | Inhibitor | BNF | Side-effect | Uncommon | 4 | Intracranial hemorrhage | No |

**Table S8. Druggability results of the prioritised proteins**

| Target Uniprot ID* | Target ChEMBL ID | Target protein name | Druggability* | Target type | Drug ChEMBL ID | Drug name | Drug molecule type | Drug mechanism | Drug effect source | Drug effect type | Drug effect frequency | Max phase* | Drug effect | Drug effect class |
|---|---|---|---|---|---|---|---|---|---|---|---|---|---|---|
| P00742 | CHEMBL244 | FA10 | Drugged | Single protein | CHEMBL198362 | Rivaroxaban | Small molecule | Inhibitor | BNF | Side-effect | Uncommon | 4 | Malaise | No |
| P00742 | CHEMBL244 | FA10 | Drugged | Single protein | CHEMBL198362 | Rivaroxaban | Small molecule | Inhibitor | BNF | Side-effect | Uncommon | 4 | Syncope | No |
| P00742 | CHEMBL244 | FA10 | Drugged | Single protein | CHEMBL198362 | Rivaroxaban | Small molecule | Inhibitor | BNF | Side-effect | Uncommon | 4 | Tachycardia | Yes |
| P00742 | CHEMBL244 | FA10 | Drugged | Single protein | CHEMBL198362 | Rivaroxaban | Small molecule | Inhibitor | BNF | Side-effect | Uncommon | 4 | Thrombocytopenia | No |
| P00742 | CHEMBL244 | FA10 | Drugged | Single protein | CHEMBL198362 | Rivaroxaban | Small molecule | Inhibitor | BNF | Side-effect | Uncommon | 4 | Thrombocytosis | No |
| P00742 | CHEMBL2111424 | FA10 | Drugged | Selectivity group | CHEMBL3833393 | Emicizumab | Antibody | Other | ChEMBL | Indication | All | 4 | Hemophilia a | No |
| P00742 | CHEMBL2111424 | FA10 | Drugged | Selectivity group | CHEMBL3833393 | Emicizumab | Antibody | Other | ChEMBL | Indication | All | 4 | Hemorrhage | No |
| P00742 | CHEMBL2111424 | FA10 | Drugged | Selectivity group | CHEMBL3833393 | Emicizumab | Antibody | Other | BNF | Indication | All | 4 | Prophylaxis of hemorrhage in hemophilia a | No |
| P00742 | CHEMBL2111424 | FA10 | Drugged | Selectivity group | CHEMBL3833393 | Emicizumab | Antibody | Other | BNF | Side-effect | Common or very common | 4 | Arthralgia | No |
| P00742 | CHEMBL2111424 | FA10 | Drugged | Selectivity group | CHEMBL3833393 | Emicizumab | Antibody | Other | BNF | Side-effect | Common or very common | 4 | Diarrhoea | No |
| P00742 | CHEMBL2111424 | FA10 | Drugged | Selectivity group | CHEMBL3833393 | Emicizumab | Antibody | Other | BNF | Side-effect | Common or very common | 4 | Fever | No |
| P00742 | CHEMBL2111424 | FA10 | Drugged | Selectivity group | CHEMBL3833393 | Emicizumab | Antibody | Other | BNF | Side-effect | Common or very common | 4 | Headache | No |
| P00742 | CHEMBL2111424 | FA10 | Drugged | Selectivity group | CHEMBL3833393 | Emicizumab | Antibody | Other | BNF | Side-effect | Common or very common | 4 | Myalgia | No |
| P00742 | CHEMBL2111424 | FA10 | Drugged | Selectivity group | CHEMBL3833393 | Emicizumab | Antibody | Other | BNF | Side-effect | Uncommon | 4 | Cavernous sinus thrombosis | No |
| P00742 | CHEMBL2111424 | FA10 | Drugged | Selectivity group | CHEMBL3833393 | Emicizumab | Antibody | Other | BNF | Side-effect | Uncommon | 4 | Embolism and thrombosis | No |
| P00742 | CHEMBL244 | FA10 | Drugged | Single protein | CHEMBL231779 | Apixaban | Small molecule | Inhibitor | ChEMBL | Indication | All | 2 | Hemorrhage | No |
| P00742 | CHEMBL244 | FA10 | Drugged | Single protein | CHEMBL198362 | Rivaroxaban | Small molecule | Inhibitor | BNF | Side-effect | Common or very common | 4 | Diarrhoea | No |
| P00742 | CHEMBL244 | FA10 | Drugged | Single protein | CHEMBL231779 | Apixaban | Small molecule | Inhibitor | ChEMBL | Indication | All | 2 | Renal insufficiency | No |
| P00742 | CHEMBL244 | FA10 | Drugged | Single protein | CHEMBL198362 | Rivaroxaban | Small molecule | Inhibitor | BNF | Side-effect | Uncommon | 4 | Dry mouth | No |
| P00742 | CHEMBL244 | FA10 | Drugged | Single protein | CHEMBL198362 | Rivaroxaban | Small molecule | Inhibitor | BNF | Side-effect | Common or very common | 4 | Fever | No |
| P00742 | CHEMBL244 | FA10 | Drugged | Single protein | CHEMBL198362 | Rivaroxaban | Small molecule | Inhibitor | BNF | Side-effect | Common or very common | 4 | Gastrointestinal discomfort | No |
| P00742 | CHEMBL244 | FA10 | Drugged | Single protein | CHEMBL198362 | Rivaroxaban | Small molecule | Inhibitor | BNF | Side-effect | Common or very common | 4 | Hemorrhage | No |
| P00742 | CHEMBL244 | FA10 | Drugged | Single protein | CHEMBL198362 | Rivaroxaban | Small molecule | Inhibitor | BNF | Side-effect | Common or very common | 4 | Headache | No |
| P00742 | CHEMBL244 | FA10 | Drugged | Single protein | CHEMBL198362 | Rivaroxaban | Small molecule | Inhibitor | BNF | Side-effect | Common or very common | 4 | Hypotension | No |
| P00742 | CHEMBL244 | FA10 | Drugged | Single protein | CHEMBL198362 | Rivaroxaban | Small molecule | Inhibitor | BNF | Side-effect | Common or very common | 4 | Menorrhagia | No |
| P00742 | CHEMBL244 | FA10 | Drugged | Single protein | CHEMBL198362 | Rivaroxaban | Small molecule | Inhibitor | BNF | Side-effect | Common or very common | 4 | Nausea | No |
| P00742 | CHEMBL244 | FA10 | Drugged | Single protein | CHEMBL198362 | Rivaroxaban | Small molecule | Inhibitor | BNF | Side-effect | Common or very common | 4 | Oedema | No |
| P00742 | CHEMBL244 | FA10 | Drugged | Single protein | CHEMBL231779 | Apixaban | Small molecule | Inhibitor | ChEMBL | Indication | All | 1 | Ischemic stroke | Yes |
| P00742 | CHEMBL244 | FA10 | Drugged | Single protein | CHEMBL198362 | Rivaroxaban | Small molecule | Inhibitor | BNF | Side-effect | Common or very common | 4 | Renal impairment | No |
| P00742 | CHEMBL244 | FA10 | Drugged | Single protein | CHEMBL198362 | Rivaroxaban | Small molecule | Inhibitor | BNF | Side-effect | Common or very common | 4 | Skin reactions | No |
| P00742 | CHEMBL244 | FA10 | Drugged | Single protein | CHEMBL198362 | Rivaroxaban | Small molecule | Inhibitor | BNF | Side-effect | Common or very common | 4 | Vomiting | No |
| P00742 | CHEMBL244 | FA10 | Drugged | Single protein | CHEMBL198362 | Rivaroxaban | Small molecule | Inhibitor | BNF | Side-effect | Common or very common | 4 | Wound complications | No |
| P00742 | CHEMBL244 | FA10 | Drugged | Single protein | CHEMBL198362 | Rivaroxaban | Small molecule | Inhibitor | BNF | Side-effect | Rare or very rare | 4 | Severe cutaneous adverse reactions scars | No |
| P00742 | CHEMBL244 | FA10 | Drugged | Single protein | CHEMBL198362 | Rivaroxaban | Small molecule | Inhibitor | BNF | Side-effect | Rare or very rare | 4 | Vascular pseudoaneurysm | No |
| P00742 | CHEMBL244 | FA10 | Drugged | Single protein | CHEMBL198362 | Rivaroxaban | Small molecule | Inhibitor | BNF | Side-effect | Uncommon | 4 | Angiodema | No |
| P00742 | CHEMBL244 | FA10 | Drugged | Single protein | CHEMBL231779 | Apixaban | Small molecule | Inhibitor | ChEMBL | Indication | All | 2 | Foramen ovale, patent | No |
| P00742 | CHEMBL244 | FA10 | Drugged | Single protein | CHEMBL198362 | Rivaroxaban | Small molecule | Inhibitor | BNF | Side-effect | Uncommon | 4 | Hepatic disorder | No |
| P00742 | CHEMBL244 | FA10 | Drugged | Single protein | CHEMBL231779 | Apixaban | Small molecule | Inhibitor | ChEMBL | Indication | All | 1 | Blood coagulation disorder | No |
| P00742 | CHEMBL244 | FA10 | Drugged | Single protein | CHEMBL231779 | Apixaban | Small molecule | Inhibitor | ChEMBL | Indication | All | 4 | Thrombosis | No |
| P00742 | CHEMBL244 | FA10 | Drugged | Single protein | CHEMBL231779 | Apixaban | Small molecule | Inhibitor | ChEMBL | Indication | All | 3 | Ischemia | Yes |
| P00742 | CHEMBL244 | FA10 | Drugged | Single protein | CHEMBL1201554 | Antithrombin alfa | Protein | Inhibitor | ChEMBL | Indication | All | 3 | Disseminated intravascular coagulation | No |
| P00742 | CHEMBL244 | FA10 | Drugged | Single protein | CHEMBL1095032 | Letaxaban | Small molecule | Inhibitor | ChEMBL | Indication | All | 2 | Venous thromboembolism | No |
| P00742 | CHEMBL244 | FA10 | Drugged | Single protein | CHEMBL1095032 | Letaxaban | Small molecule | Inhibitor | ChEMBL | Indication | All | 2 | Acute coronary syndrome | Yes |
| P00742 | CHEMBL244 | FA10 | Drugged | Single protein | CHEMBL1908371 | Idraparinux sodium | Oligosaccharide | Inhibitor | ChEMBL | Indication | All | 3 | Venous thrombosis | No |
| P00742 | CHEMBL244 | FA10 | Drugged | Single protein | CHEMBL1908371 | Idraparinux sodium | Oligosaccharide | Inhibitor | ChEMBL | Indication | All | 3 | Pulmonary embolism | No |
| P00742 | CHEMBL244 | FA10 | Drugged | Single protein | CHEMBL1908371 | Idraparinux sodium | Oligosaccharide | Inhibitor | ChEMBL | Indication | All | 3 | Atrial fibrillation | Yes |
| P00742 | CHEMBL244 | FA10 | Drugged | Single protein | CHEMBL512351 | Betrixaban | Small molecule | Inhibitor | ChEMBL | Indication | All | 4 | Venous thromboembolism | No |
| P00742 | CHEMBL244 | FA10 | Drugged | Single protein | CHEMBL512351 | Betrixaban | Small molecule | Inhibitor | ChEMBL | Indication | All | 4 | Thrombosis | No |
| P00742 | CHEMBL244 | FA10 | Drugged | Single protein | CHEMBL512351 | Betrixaban | Small molecule | Inhibitor | ChEMBL | Indication | All | 2 | Thromboembolism | No |
| P00742 | CHEMBL244 | FA10 | Drugged | Single protein | CHEMBL512351 | Betrixaban | Small molecule | Inhibitor | ChEMBL | Indication | All | 2 | Hemorrhage | No |
| P00742 | CHEMBL244 | FA10 | Drugged | Single protein | CHEMBL512351 | Betrixaban | Small molecule | Inhibitor | ChEMBL | Indication | All | 2 | Atrial flutter | Yes |
| P00742 | CHEMBL244 | FA10 | Drugged | Single protein | CHEMBL512351 | Betrixaban | Small molecule | Inhibitor | ChEMBL | Indication | All | 2 | Atrial fibrillation | Yes |
| P00742 | CHEMBL244 | FA10 | Drugged | Single protein | CHEMBL512351 | Betrixaban | Small molecule | Inhibitor | ChEMBL | Indication | All | 1 | Liver diseases | No |
| P00742 | CHEMBL244 | FA10 | Drugged | Single protein | CHEMBL512351 | Betrixaban | Small molecule | Inhibitor | ChEMBL | Indication | All | 1 | Kidney diseases | No |
| P00742 | CHEMBL244 | FA10 | Drugged | Single protein | CHEMBL2105682 | Edoxaban tosylate | Small molecule | Inhibitor | ChEMBL | Indication | All | 4 | Stroke | No |
| P00742 | CHEMBL244 | FA10 | Drugged | Single protein | CHEMBL2105682 | Edoxaban tosylate | Small molecule | Inhibitor | ChEMBL | Indication | All | 4 | Venous thrombosis | No |
| P00742 | CHEMBL244 | FA10 | Drugged | Single protein | CHEMBL2105682 | Edoxaban tosylate | Small molecule | Inhibitor | ChEMBL | Indication | All | 4 | Pulmonary embolism | No |
| P00742 | CHEMBL244 | FA10 | Drugged | Single protein | CHEMBL1201554 | Antithrombin alfa | Protein | Inhibitor | ChEMBL | Indication | All | 3 | Pre-eclampsia | No |
| P00742 | CHEMBL244 | FA10 | Drugged | Single protein | CHEMBL46618 | Otamixaban | Small molecule | Inhibitor | ChEMBL | Indication | All | 1 | Kidney diseases | No |
| P00742 | CHEMBL244 | FA10 | Drugged | Single protein | CHEMBL46618 | Otamixaban | Small molecule | Inhibitor | ChEMBL | Indication | All | 1 | Liver diseases | No |
| P00742 | CHEMBL244 | FA10 | Drugged | Single protein | CHEMBL46618 | Otamixaban | Small molecule | Inhibitor | ChEMBL | Indication | All | 2 | Coronary disease | Yes |
| P00742 | CHEMBL244 | FA10 | Drugged | Single protein | CHEMBL231779 | Apixaban | Small molecule | Inhibitor | ChEMBL | Indication | All | 1 | Nephrotic syndrome | No |
| P00742 | CHEMBL244 | FA10 | Drugged | Single protein | CHEMBL231779 | Apixaban | Small molecule | Inhibitor | ChEMBL | Indication | All | 1 | Thromboembolism | No |
| P00742 | CHEMBL244 | FA10 | Drugged | Single protein | CHEMBL4297951 | Coagulation factor x human | Unknown | Positive modulator | ChEMBL | Indication | All | 3 | Factor x deficiency | No |
| P00742 | CHEMBL244 | FA10 | Drugged | Single protein | CHEMBL231779 | Apixaban | Small molecule | Inhibitor | ChEMBL | Indication | All | 1 | Liver cirrhosis | No |
| P00742 | CHEMBL244 | FA10 | Drugged | Single protein | CHEMBL4297768 | Idrabiotaparinux sodium | Oligosaccharide | Inhibitor | ChEMBL | Indication | All | 3 | Venous thrombosis | No |
| P00742 | CHEMBL244 | FA10 | Drugged | Single protein | CHEMBL4297768 | Idrabiotaparinux sodium | Oligosaccharide | Inhibitor | ChEMBL | Indication | All | 3 | Thrombosis | No |
| P00742 | CHEMBL244 | FA10 | Drugged | Single protein | CHEMBL4297768 | Idrabiotaparinux sodium | Oligosaccharide | Inhibitor | ChEMBL | Indication | All | 3 | Atrial fibrillation | Yes |
| P00742 | CHEMBL244 | FA10 | Drugged | Single protein | CHEMBL1922235 | Darexaban | Small molecule | Inhibitor | ChEMBL | Indication | All | 3 | Venous thromboembolism | No |
| P00742 | CHEMBL244 | FA10 | Drugged | Single protein | CHEMBL2105682 | Edoxaban tosylate | Small molecule | Inhibitor | ChEMBL | Indication | All | 4 | Embolism | No |
| P00742 | CHEMBL244 | FA10 | Drugged | Single protein | CHEMBL1922235 | Darexaban | Small molecule | Inhibitor | ChEMBL | Indication | All | 2 | Acute coronary syndrome | Yes |
| P00742 | CHEMBL244 | FA10 | Drugged | Single protein | CHEMBL1922235 | Darexaban | Small molecule | Inhibitor | ChEMBL | Indication | All | 2 | Atrial fibrillation | Yes |
| P00742 | CHEMBL244 | FA10 | Drugged | Single protein | CHEMBL3707365 | Bemiparin | Oligosaccharide | Inhibitor | ChEMBL | Indication | All | 3 | Diabetic foot | No |
| P00742 | CHEMBL244 | FA10 | Drugged | Single protein | CHEMBL3707365 | Bemiparin | Oligosaccharide | Inhibitor | ChEMBL | Indication | All | 3 | Thrombosis | No |
| P00742 | CHEMBL244 | FA10 | Drugged | Single protein | CHEMBL3707365 | Bemiparin | Oligosaccharide | Inhibitor | ChEMBL | Indication | All | 3 | Liver cirrhosis | No |
| P00742 | CHEMBL244 | FA10 | Drugged | Single protein | CHEMBL3707365 | Bemiparin | Oligosaccharide | Inhibitor | ChEMBL | Indication | All | 2 | Cancers | No |
| P00742 | CHEMBL244 | FA10 | Drugged | Single protein | CHEMBL3707365 | Bemiparin | Oligosaccharide | Inhibitor | ChEMBL | Indication | All | 2 | Severe acute respiratory syndrome | No |
| P00742 | CHEMBL244 | FA10 | Drugged | Single protein | CHEMBL3707365 | Bemiparin | Oligosaccharide | Inhibitor | ChEMBL | Indication | All | 1 | Renal insufficiency | No |
| P00742 | CHEMBL244 | FA10 | Drugged | Single protein | CHEMBL46618 | Otamixaban | Small molecule | Inhibitor | ChEMBL | Indication | All | 3 | Acute coronary syndrome | Yes |
| P00742 | CHEMBL244 | FA10 | Drugged | Single protein | CHEMBL1922235 | Darexaban | Small molecule | Inhibitor | ChEMBL | Indication | All | 2 | Thromboembolism | No |
| P00742 | CHEMBL244 | FA10 | Drugged | Single protein | CHEMBL231779 | Apixaban | Small molecule | Inhibitor | ChEMBL | Indication | All | 3 | Cancers | No |
| P00742 | CHEMBL244 | FA10 | Drugged | Single protein | CHEMBL2105682 | Edoxaban tosylate | Small molecule | Inhibitor | ChEMBL | Indication | All | 4 | Atrial fibrillation | Yes |
| P00742 | CHEMBL244 | FA10 | Drugged | Single protein | CHEMBL2105682 | Edoxaban tosylate | Small molecule | Inhibitor | ChEMBL | Indication | All | 2 | Thrombosis | No |
| P00742 | CHEMBL244 | FA10 | Drugged | Single protein | CHEMBL198362 | Rivaroxaban | Small molecule | Inhibitor | ChEMBL | Indication | All | 2 | Ischemia | Yes |
| P00742 | CHEMBL244 | FA10 | Drugged | Single protein | CHEMBL198362 | Rivaroxaban | Small molecule | Inhibitor | ChEMBL | Indication | All | 2 | Hemorrhage | No |
| P00742 | CHEMBL244 | FA10 | Drugged | Single protein | CHEMBL198362 | Rivaroxaban | Small molecule | Inhibitor | ChEMBL | Indication | All | 2 | Heart diseases | Yes |
| P00742 | CHEMBL244 | FA10 | Drugged | Single protein | CHEMBL198362 | Rivaroxaban | Small molecule | Inhibitor | ChEMBL | Indication | All | 2 | Anemia | No |

**Table S8. Druggability results of the prioritised proteins**

| Target Uniprot ID* | Target ChEMBL ID | Target protein name | Druggability* | Target type | Drug ChEMBL ID | Drug name | Drug molecule type | Drug mechanism | Drug effect source | Drug effect type | Drug effect frequency | Max phase* | Drug effect | Drug effect class |
|---|---|---|---|---|---|---|---|---|---|---|---|---|---|---|
| P00742 | CHEMBL244 | FA10 | Drugged | Single protein | CHEMBL198362 | Rivaroxaban | Small molecule | Inhibitor | ChEMBL | Indication | All | 2 | Aortic valve disease | Yes |
| P00742 | CHEMBL244 | FA10 | Drugged | Single protein | CHEMBL198362 | Rivaroxaban | Small molecule | Inhibitor | ChEMBL | Indication | All | 1 | Renal insufficiency | No |
| P00742 | CHEMBL244 | FA10 | Drugged | Single protein | CHEMBL198362 | Rivaroxaban | Small molecule | Inhibitor | ChEMBL | Indication | All | 1 | Obesity, morbid | No |
| P00742 | CHEMBL244 | FA10 | Drugged | Single protein | CHEMBL198362 | Rivaroxaban | Small molecule | Inhibitor | ChEMBL | Indication | All | 1 | Blood coagulation disorder | No |
| P00742 | CHEMBL244 | FA10 | Drugged | Single protein | CHEMBL231779 | Apixaban | Small molecule | Inhibitor | ChEMBL | Indication | All | 4 | Stroke | No |
| P00742 | CHEMBL244 | FA10 | Drugged | Single protein | CHEMBL231779 | Apixaban | Small molecule | Inhibitor | ChEMBL | Indication | All | 4 | Venous thrombosis | No |
| P00742 | CHEMBL244 | FA10 | Drugged | Single protein | CHEMBL231779 | Apixaban | Small molecule | Inhibitor | ChEMBL | Indication | All | 4 | Pulmonary embolism | No |
| P00742 | CHEMBL244 | FA10 | Drugged | Single protein | CHEMBL231779 | Apixaban | Small molecule | Inhibitor | ChEMBL | Indication | All | 4 | Embolism | No |
| P00742 | CHEMBL244 | FA10 | Drugged | Single protein | CHEMBL231779 | Apixaban | Small molecule | Inhibitor | ChEMBL | Indication | All | 4 | Atrial fibrillation | Yes |
| P00742 | CHEMBL244 | FA10 | Drugged | Single protein | CHEMBL231779 | Apixaban | Small molecule | Inhibitor | ChEMBL | Indication | All | 3 | Venous thromboembolism | No |
| P00742 | CHEMBL244 | FA10 | Drugged | Single protein | CHEMBL231779 | Apixaban | Small molecule | Inhibitor | ChEMBL | Indication | All | 3 | Acute coronary syndrome | Yes |
| P00742 | CHEMBL244 | FA10 | Drugged | Single protein | CHEMBL231779 | Apixaban | Small molecule | Inhibitor | ChEMBL | Indication | All | 3 | Renal insufficiency | No |
| P00742 | CHEMBL244 | FA10 | Drugged | Single protein | CHEMBL231779 | Apixaban | Small molecule | Inhibitor | ChEMBL | Indication | All | 3 | Severe acute respiratory syndrome | No |
| P00742 | CHEMBL244 | FA10 | Drugged | Single protein | CHEMBL231779 | Apixaban | Small molecule | Inhibitor | ChEMBL | Indication | All | 2 | Heart failure | Yes |
| P00742 | CHEMBL244 | FA10 | Drugged | Single protein | CHEMBL198362 | Rivaroxaban | Small molecule | Inhibitor | ChEMBL | Indication | All | 2 | Thromboembolism | No |
| P00742 | CHEMBL244 | FA10 | Drugged | Single protein | CHEMBL198362 | Rivaroxaban | Small molecule | Inhibitor | ChEMBL | Indication | All | 2 | Acute coronary syndrome | Yes |
| P00742 | CHEMBL244 | FA10 | Drugged | Single protein | CHEMBL198362 | Rivaroxaban | Small molecule | Inhibitor | ChEMBL | Indication | All | 3 | Ischemic stroke | Yes |
| P00742 | CHEMBL244 | FA10 | Drugged | Single protein | CHEMBL198362 | Rivaroxaban | Small molecule | Inhibitor | ChEMBL | Indication | All | 4 | Venous thrombosis | No |
| P00742 | CHEMBL244 | FA10 | Drugged | Single protein | CHEMBL198362 | Rivaroxaban | Small molecule | Inhibitor | ChEMBL | Indication | All | 4 | Thrombosis | No |
| P00742 | CHEMBL244 | FA10 | Drugged | Single protein | CHEMBL198362 | Rivaroxaban | Small molecule | Inhibitor | ChEMBL | Indication | All | 4 | Pulmonary embolism | No |
| P00742 | CHEMBL244 | FA10 | Drugged | Single protein | CHEMBL198362 | Rivaroxaban | Small molecule | Inhibitor | ChEMBL | Indication | All | 4 | Atrial fibrillation | Yes |
| P00742 | CHEMBL244 | FA10 | Drugged | Single protein | CHEMBL198362 | Rivaroxaban | Small molecule | Inhibitor | ChEMBL | Indication | All | 3 | Peripheral arterial disease | No |
| P00742 | CHEMBL244 | FA10 | Drugged | Single protein | CHEMBL198362 | Rivaroxaban | Small molecule | Inhibitor | ChEMBL | Indication | All | 3 | Venous thromboembolism | No |
| P00742 | CHEMBL244 | FA10 | Drugged | Single protein | CHEMBL198362 | Rivaroxaban | Small molecule | Inhibitor | ChEMBL | Indication | All | 3 | Renal insufficiency | No |
| P00742 | CHEMBL244 | FA10 | Drugged | Single protein | CHEMBL198362 | Rivaroxaban | Small molecule | Inhibitor | ChEMBL | Indication | All | 3 | Severe acute respiratory syndrome | No |
| P00742 | CHEMBL244 | FA10 | Drugged | Single protein | CHEMBL2105682 | Edoxaban tosylate | Small molecule | Inhibitor | ChEMBL | Indication | All | 3 | Venous thromboembolism | No |
| P00742 | CHEMBL244 | FA10 | Drugged | Single protein | CHEMBL198362 | Rivaroxaban | Small molecule | Inhibitor | ChEMBL | Indication | All | 3 | Stroke | No |
| P00742 | CHEMBL244 | FA10 | Drugged | Single protein | CHEMBL198362 | Rivaroxaban | Small molecule | Inhibitor | ChEMBL | Indication | All | 3 | Thrombocytopenia | No |
| P00742 | CHEMBL244 | FA10 | Drugged | Single protein | CHEMBL198362 | Rivaroxaban | Small molecule | Inhibitor | ChEMBL | Indication | All | 3 | Rheumatic heart disease | No |
| P00742 | CHEMBL244 | FA10 | Drugged | Single protein | CHEMBL198362 | Rivaroxaban | Small molecule | Inhibitor | ChEMBL | Indication | All | 3 | Myocardial infarction | Yes |
| P00742 | CHEMBL244 | FA10 | Drugged | Single protein | CHEMBL198362 | Rivaroxaban | Small molecule | Inhibitor | ChEMBL | Indication | All | 3 | Liver cirrhosis | No |
| P00742 | CHEMBL244 | FA10 | Drugged | Single protein | CHEMBL198362 | Rivaroxaban | Small molecule | Inhibitor | ChEMBL | Indication | All | 3 | Cancers | No |
| P00742 | CHEMBL244 | FA10 | Drugged | Single protein | CHEMBL198362 | Rivaroxaban | Small molecule | Inhibitor | ChEMBL | Indication | All | 3 | Diabetes mellitus, type 2 | No |
| P00742 | CHEMBL244 | FA10 | Drugged | Single protein | CHEMBL198362 | Rivaroxaban | Small molecule | Inhibitor | ChEMBL | Indication | All | 3 | Coronary disease | Yes |
| P00742 | CHEMBL244 | FA10 | Drugged | Single protein | CHEMBL198362 | Rivaroxaban | Small molecule | Inhibitor | ChEMBL | Indication | All | 3 | Aortic valve stenosis | Yes |
| P00742 | CHEMBL244 | FA10 | Drugged | Single protein | CHEMBL198362 | Rivaroxaban | Small molecule | Inhibitor | ChEMBL | Indication | All | 3 | Antiphospholipid syndrome | No |
| P00742 | CHEMBL244 | FA10 | Drugged | Single protein | CHEMBL198362 | Rivaroxaban | Small molecule | Inhibitor | ChEMBL | Indication | All | 2 | Cancers | No |
| P00742 | CHEMBL2111424 | FA10 | Drugged | Selectivity group | CHEMBL3833393 | Emicizumab | Antibody | Other | BNF | Side-effect | Uncommon | 4 | Thrombotic microangiopathy | No |
| P00742 | CHEMBL2111424 | FA10 | Drugged | Selectivity group | CHEMBL3833393 | Emicizumab | Antibody | Other | BNF | Side-effect | Uncommon | 4 | Skin necrosis | No |
| P16930 | - | FAAA | Not yet druggable | - | - | - | - | - | - | - | - | - | - | No |
| P12318 | CHEMBL5841 | FCG2A | Druggable | Single protein | - | - | - | - | - | - | - | - | - | No |
| P08637 | CHEMBL3856162 | FCG3A | Druggable | Single protein | CHEMBL2109389 | Imgatuzumab | Antibody | Cross-linking agent | ChEMBL | Indication | All | 1 | Cancers | No |
| P08637 | CHEMBL3856162 | FCG3A | Druggable | Single protein | CHEMBL2109389 | Imgatuzumab | Antibody | Cross-linking agent | ChEMBL | Indication | All | 2 | Cancers | No |
| Q6BAA4 | - | FCRLB | Not yet druggable | - | - | - | - | - | - | - | - | - | - | No |
| O95479 | - | G6PE | Not yet druggable | - | - | - | - | - | - | - | - | - | - | No |
| P56159 | CHEMBL3833481 | GFRA1 | Druggable | Single protein | CHEMBL2108380 | Liatermin | Protein | Agonist | ChEMBL | Indication | All | 1 | Parkinson disease | No |
| O94923 | - | GLCE | Not yet druggable | - | - | - | - | - | - | - | - | - | - | No |
| Q9P2T1 | CHEMBL4296017 | GMPR2 | Druggable | Single protein | - | - | - | - | - | - | - | - | - | No |
| Q9UJJ9 | - | GNPTG | Not yet druggable | - | - | - | - | - | - | - | - | - | - | No |
| P10144 | CHEMBL2316 | GRAB | Drugged | Single protein | - | - | - | - | - | - | - | - | - | No |
| P21266 | CHEMBL2242 | GSTM3 | Drugged | Single protein | CHEMBL513 | Carmustine | Small molecule | Inhibitor | ChEMBL | Indication | All | 2 | Polyradiculoneuropathy, chronic inflammatory demyelinating | No |
| P21266 | CHEMBL2242 | GSTM3 | Drugged | Single protein | CHEMBL513 | Carmustine | Small molecule | Inhibitor | ChEMBL | Indication | All | 2 | Opsoclonus-myoclonus syndrome | No |
| P21266 | CHEMBL2242 | GSTM3 | Drugged | Single protein | CHEMBL513 | Carmustine | Small molecule | Inhibitor | ChEMBL | Indication | All | 3 | Cancers | No |
| P21266 | CHEMBL2242 | GSTM3 | Drugged | Single protein | CHEMBL513 | Carmustine | Small molecule | Inhibitor | ChEMBL | Indication | All | 2 | Neuromyelitis optica | No |
| P21266 | CHEMBL2242 | GSTM3 | Drugged | Single protein | CHEMBL513 | Carmustine | Small molecule | Inhibitor | ChEMBL | Indication | All | 2 | Cancers | No |
| P21266 | CHEMBL2242 | GSTM3 | Drugged | Single protein | CHEMBL513 | Carmustine | Small molecule | Inhibitor | ChEMBL | Indication | All | 4 | Cancers | No |
| P21266 | CHEMBL2242 | GSTM3 | Drugged | Single protein | CHEMBL513 | Carmustine | Small molecule | Inhibitor | ChEMBL | Indication | All | 4 | Hodgkin disease | No |
| P21266 | CHEMBL2242 | GSTM3 | Drugged | Single protein | CHEMBL513 | Carmustine | Small molecule | Inhibitor | ChEMBL | Indication | All | 1 | Sezary syndrome | No |
| P21266 | CHEMBL2242 | GSTM3 | Drugged | Single protein | CHEMBL513 | Carmustine | Small molecule | Inhibitor | ChEMBL | Indication | All | 2 | Myasthenia gravis | No |
| P21266 | CHEMBL2242 | GSTM3 | Drugged | Single protein | CHEMBL513 | Carmustine | Small molecule | Inhibitor | ChEMBL | Indication | All | 1 | Mycosis fungoides | No |
| P21266 | CHEMBL2242 | GSTM3 | Drugged | Single protein | CHEMBL513 | Carmustine | Small molecule | Inhibitor | ChEMBL | Indication | All | 2 | Stiff-person syndrome | No |
| P21266 | CHEMBL2242 | GSTM3 | Drugged | Single protein | CHEMBL513 | Carmustine | Small molecule | Inhibitor | ChEMBL | Indication | All | 4 | Ependymoma | No |
| P21266 | CHEMBL2242 | GSTM3 | Drugged | Single protein | CHEMBL513 | Carmustine | Small molecule | Inhibitor | ChEMBL | Indication | All | 2 | Multiple sclerosis, relapsing-remitting | No |
| P21266 | CHEMBL2242 | GSTM3 | Drugged | Single protein | CHEMBL513 | Carmustine | Small molecule | Inhibitor | ChEMBL | Indication | All | 2 | Hematologic diseases | No |
| P21266 | CHEMBL2242 | GSTM3 | Drugged | Single protein | CHEMBL513 | Carmustine | Small molecule | Inhibitor | ChEMBL | Indication | All | 1 | Cancers | No |
| P21266 | CHEMBL2242 | GSTM3 | Drugged | Single protein | CHEMBL513 | Carmustine | Small molecule | Inhibitor | ChEMBL | Indication | All | 2 | Graft vs host disease | No |
| P21266 | CHEMBL2242 | GSTM3 | Drugged | Single protein | CHEMBL513 | Carmustine | Small molecule | Inhibitor | ChEMBL | Indication | All | 4 | Medulloblastoma | No |
| Q4G148 | - | GXLT1 | Not yet druggable | - | - | - | - | - | - | - | - | - | - | No |
| Q96D42 | - | HAVR1 | Not yet druggable | - | - | - | - | - | - | - | - | - | - | No |
| Q8NFM7 | - | I17RD | Not yet druggable | - | - | - | - | - | - | - | - | - | - | No |
| P17936 | CHEMBL3997 | IBP3 | Druggable | Single protein | - | - | - | - | - | - | - | - | - | No |
| Q9BXS1 | - | IDI2 | Not yet druggable | - | - | - | - | - | - | - | - | - | - | No |
| Q9H665 | CHEMBL2029192 | IGFR1 | Druggable | Single protein | - | - | - | - | - | - | - | - | - | No |
| Q13478 | CHEMBL4804253 | IL18R | Druggable | Protein complex | CHEMBL2108034 | Iboctadekin | Protein | Agonist | ChEMBL | Indication | All | 1 | Cancers | No |
| Q13478 | CHEMBL4804253 | IL18R | Druggable | Protein complex | CHEMBL2108034 | Iboctadekin | Protein | Agonist | ChEMBL | Indication | All | 2 | Cancers | No |
| P08887 | CHEMBL2364155 | IL6RA | Drugged | Single protein | CHEMBL1237022 | Tocilizumab | Antibody | Inhibitor | BNF | Side-effect | Rare or very rare | 4 | Stevens-johnson syndrome | No |
| P08887 | CHEMBL2364155 | IL6RA | Drugged | Single protein | CHEMBL1237022 | Tocilizumab | Antibody | Inhibitor | BNF | Side-effect | Not known | 4 | Pancytopenia | No |
| P08887 | CHEMBL2364155 | IL6RA | Drugged | Single protein | CHEMBL1237022 | Tocilizumab | Antibody | Inhibitor | BNF | Side-effect | Common or very common | 4 | Cough | No |
| P08887 | CHEMBL2364155 | IL6RA | Drugged | Single protein | CHEMBL1237022 | Tocilizumab | Antibody | Inhibitor | BNF | Side-effect | Common or very common | 4 | Dizziness | No |
| P08887 | CHEMBL2364155 | IL6RA | Drugged | Single protein | CHEMBL1237022 | Tocilizumab | Antibody | Inhibitor | BNF | Side-effect | Not known | 4 | Interstitial lung disease | No |
| P08887 | CHEMBL2364155 | IL6RA | Drugged | Single protein | CHEMBL1237022 | Tocilizumab | Antibody | Inhibitor | BNF | Side-effect | Common or very common | 4 | Dyslipidemia | No |
| P08887 | CHEMBL2364155 | IL6RA | Drugged | Single protein | CHEMBL1237022 | Tocilizumab | Antibody | Inhibitor | BNF | Side-effect | Common or very common | 4 | Dyspnoea | No |
| P08887 | CHEMBL2364155 | IL6RA | Drugged | Single protein | CHEMBL1237022 | Tocilizumab | Antibody | Inhibitor | BNF | Side-effect | Common or very common | 4 | Gastrointestinal disorder | No |
| P08887 | CHEMBL2364155 | IL6RA | Drugged | Single protein | CHEMBL1237022 | Tocilizumab | Antibody | Inhibitor | BNF | Side-effect | Common or very common | 4 | Skin reactions | No |
| P08887 | CHEMBL2364155 | IL6RA | Drugged | Single protein | CHEMBL1237022 | Tocilizumab | Antibody | Inhibitor | BNF | Side-effect | Common or very common | 4 | Headache | No |
| P08887 | CHEMBL2364155 | IL6RA | Drugged | Single protein | CHEMBL1237022 | Tocilizumab | Antibody | Inhibitor | BNF | Side-effect | Common or very common | 4 | Hypersensitivity | No |
| P08887 | CHEMBL2364155 | IL6RA | Drugged | Single protein | CHEMBL1237022 | Tocilizumab | Antibody | Inhibitor | BNF | Side-effect | Common or very common | 4 | Hypertension | Yes |

**Table S8. Druggability results of the prioritised proteins**

| Target Uniprot ID* | Target ChEMBL ID | Target protein name | Druggability* | Target type | Drug ChEMBL ID | Drug name | Drug molecule type | Drug mechanism | Drug effect source | Drug effect type | Drug effect frequency | Max phase* | Drug effect | Drug effect class |
|---|---|---|---|---|---|---|---|---|---|---|---|---|---|---|
| P08887 | CHEMBL2364155 | IL6RA | Drugged | Single protein | CHEMBL1237022 | Tocilizumab | Antibody | Inhibitor | BNF | Side-effect | Not known | 4 | Infusion related reaction | No |
| P08887 | CHEMBL2364155 | IL6RA | Drugged | Single protein | CHEMBL1237022 | Tocilizumab | Antibody | Inhibitor | BNF | Side-effect | Common or very common | 4 | Increased risk of infection | No |
| P08887 | CHEMBL2364155 | IL6RA | Drugged | Single protein | CHEMBL1237022 | Tocilizumab | Antibody | Inhibitor | BNF | Side-effect | Uncommon | 4 | Hypothyroidism | No |
| P08887 | CHEMBL2364155 | IL6RA | Drugged | Single protein | CHEMBL1237022 | Tocilizumab | Antibody | Inhibitor | BNF | Side-effect | Common or very common | 4 | Leucopenia | No |
| P08887 | CHEMBL2364155 | IL6RA | Drugged | Single protein | CHEMBL1237022 | Tocilizumab | Antibody | Inhibitor | BNF | Side-effect | Not known | 4 | Hepatic disorder | No |
| P08887 | CHEMBL2364155 | IL6RA | Drugged | Single protein | CHEMBL1237022 | Tocilizumab | Antibody | Inhibitor | BNF | Side-effect | Common or very common | 4 | Weight increased | No |
| P08887 | CHEMBL2364155 | IL6RA | Drugged | Single protein | CHEMBL1237022 | Tocilizumab | Antibody | Inhibitor | BNF | Side-effect | Common or very common | 4 | Oral disorder | No |
| P08887 | CHEMBL2364155 | IL6RA | Drugged | Single protein | CHEMBL1237022 | Tocilizumab | Antibody | Inhibitor | BNF | Side-effect | Common or very common | 4 | Peripheral oedema | No |
| P08887 | CHEMBL2364155 | IL6RA | Drugged | Single protein | CHEMBL1237022 | Tocilizumab | Antibody | Inhibitor | BNF | Side-effect | Uncommon | 4 | Nephrolithiasis | No |
| P08887 | CHEMBL2364155 | IL6RA | Drugged | Single protein | CHEMBL81 | Raloxifene | Small molecule | Modulator | ChEMBL | Indication | All | 2 | Menopause | No |
| P08887 | CHEMBL3137266 | IL6RA | Drugged | Protein-protein interaction | CHEMBL3833307 | Satralizumab | Antibody | Antagonist | ChEMBL | Indication | All | 3 | Myasthenia gravis | No |
| P08887 | CHEMBL2364155 | IL6RA | Drugged | Single protein | CHEMBL81 | Raloxifene | Small molecule | Modulator | ChEMBL | Indication | All | 4 | Osteoporosis | No |
| P08887 | CHEMBL2364155 | IL6RA | Drugged | Single protein | CHEMBL46740 | Bazedoxifene | Small molecule | Modulator | ChEMBL | Indication | All | 3 | Endometrial hyperplasia | No |
| P08887 | CHEMBL2364155 | IL6RA | Drugged | Single protein | CHEMBL46740 | Bazedoxifene | Small molecule | Modulator | ChEMBL | Indication | All | 2 | Cancers | No |
| P08887 | CHEMBL2364155 | IL6RA | Drugged | Single protein | CHEMBL46740 | Bazedoxifene | Small molecule | Modulator | ChEMBL | Indication | All | 3 | Menopause | No |
| P08887 | CHEMBL2364155 | IL6RA | Drugged | Single protein | CHEMBL46740 | Bazedoxifene | Small molecule | Modulator | ChEMBL | Indication | All | 2 | Multiple sclerosis, relapsing-remitting | No |
| P08887 | CHEMBL2364155 | IL6RA | Drugged | Single protein | CHEMBL46740 | Bazedoxifene | Small molecule | Modulator | ChEMBL | Indication | All | 3 | Osteoporosis | No |
| P08887 | CHEMBL2364155 | IL6RA | Drugged | Single protein | CHEMBL46740 | Bazedoxifene | Small molecule | Modulator | ChEMBL | Indication | All | 4 | Osteoporosis, postmenopausal | No |
| P08887 | CHEMBL2364155 | IL6RA | Drugged | Single protein | CHEMBL81 | Raloxifene | Small molecule | Modulator | ChEMBL | Indication | All | 2 | Alzheimer disease | No |
| P08887 | CHEMBL2364155 | IL6RA | Drugged | Single protein | CHEMBL81 | Raloxifene | Small molecule | Modulator | ChEMBL | Indication | All | 3 | Polycystic ovary syndrome | No |
| P08887 | CHEMBL2364155 | IL6RA | Drugged | Single protein | CHEMBL81 | Raloxifene | Small molecule | Modulator | ChEMBL | Indication | All | 3 | Bone diseases, metabolic | No |
| P08887 | CHEMBL2364155 | IL6RA | Drugged | Single protein | CHEMBL81 | Raloxifene | Small molecule | Modulator | ChEMBL | Indication | All | 3 | Psychotic disorder | No |
| P08887 | CHEMBL2364155 | IL6RA | Drugged | Single protein | CHEMBL81 | Raloxifene | Small molecule | Modulator | ChEMBL | Indication | All | 2 | Severe acute respiratory syndrome | No |
| P08887 | CHEMBL2364155 | IL6RA | Drugged | Single protein | CHEMBL81 | Raloxifene | Small molecule | Modulator | ChEMBL | Indication | All | 3 | Schizophrenia | No |
| P08887 | CHEMBL2364155 | IL6RA | Drugged | Single protein | CHEMBL81 | Raloxifene | Small molecule | Modulator | ChEMBL | Indication | All | 2 | Cancers | No |
| P08887 | CHEMBL2364155 | IL6RA | Drugged | Single protein | CHEMBL81 | Raloxifene | Small molecule | Modulator | ChEMBL | Indication | All | 4 | Cancers | No |
| P08887 | CHEMBL2364155 | IL6RA | Drugged | Single protein | CHEMBL1237022 | Tocilizumab | Antibody | Inhibitor | BNF | Side-effect | Common or very common | 4 | Conjunctivitis | No |
| P08887 | CHEMBL2364155 | IL6RA | Drugged | Single protein | CHEMBL81 | Raloxifene | Small molecule | Modulator | ChEMBL | Indication | All | 3 | Osteoporosis, postmenopausal | No |
| P08887 | CHEMBL3137266 | IL6RA | Drugged | Protein-protein interaction | CHEMBL3833307 | Satralizumab | Antibody | Antagonist | ChEMBL | Indication | All | 4 | Neuromyelitis optica | No |
| P08887 | CHEMBL3137266 | IL6RA | Drugged | Protein-protein interaction | CHEMBL3833307 | Satralizumab | Antibody | Antagonist | ChEMBL | Indication | All | 4 | Immune system diseases | No |
| P08887 | CHEMBL3137266 | IL6RA | Drugged | Protein-protein interaction | CHEMBL3833307 | Satralizumab | Antibody | Antagonist | ChEMBL | Indication | All | 3 | Encephalitis, japanese | No |
| P08887 | CHEMBL2364155 | IL6RA | Drugged | Single protein | CHEMBL1237022 | Tocilizumab | Antibody | Inhibitor | BNF | Side-effect | Common or very common | 4 | Abdominal pain | No |
| P08887 | CHEMBL2364155 | IL6RA | Drugged | Single protein | CHEMBL1237022 | Tocilizumab | Antibody | Inhibitor | BNF | Side-effect | Common or very common | 4 | Neutropenia | No |
| P08887 | CHEMBL2364155 | IL6RA | Drugged | Single protein | CHEMBL1237022 | Tocilizumab | Antibody | Inhibitor | BNF | Indication | All | 4 | Giant cell arteritis | No |
| P08887 | CHEMBL2364155 | IL6RA | Drugged | Single protein | CHEMBL1237022 | Tocilizumab | Antibody | Inhibitor | ChEMBL | Indication | All | 2 | Neuromyelitis optica | No |
| P08887 | CHEMBL2364155 | IL6RA | Drugged | Single protein | CHEMBL1237022 | Tocilizumab | Antibody | Inhibitor | ChEMBL | Indication | All | 2 | Familial mediterranean fever | No |
| P08887 | CHEMBL2364155 | IL6RA | Drugged | Single protein | CHEMBL1237022 | Tocilizumab | Antibody | Inhibitor | ChEMBL | Indication | All | 2 | Pneumonia | No |
| P08887 | CHEMBL2364155 | IL6RA | Drugged | Single protein | CHEMBL1237022 | Tocilizumab | Antibody | Inhibitor | BNF | Indication | All | 4 | Arthritis | No |
| P08887 | CHEMBL2364155 | IL6RA | Drugged | Single protein | CHEMBL1237022 | Tocilizumab | Antibody | Inhibitor | ChEMBL | Indication | All | 2 | Polychondritis, relapsing | No |
| P08887 | CHEMBL2364155 | IL6RA | Drugged | Single protein | CHEMBL1237022 | Tocilizumab | Antibody | Inhibitor | ChEMBL | Indication | All | 2 | Respiratory distress syndrome | No |
| P08887 | CHEMBL2364155 | IL6RA | Drugged | Single protein | CHEMBL1237022 | Tocilizumab | Antibody | Inhibitor | ChEMBL | Indication | All | 2 | Myasthenia gravis | No |
| P08887 | CHEMBL2364155 | IL6RA | Drugged | Single protein | CHEMBL1237022 | Tocilizumab | Antibody | Inhibitor | ChEMBL | Indication | All | 2 | Uveitis | No |
| P08887 | CHEMBL2364155 | IL6RA | Drugged | Single protein | CHEMBL1237022 | Tocilizumab | Antibody | Inhibitor | ChEMBL | Indication | All | 2 | Schnitzler syndrome | No |
| P08887 | CHEMBL2364155 | IL6RA | Drugged | Single protein | CHEMBL1237022 | Tocilizumab | Antibody | Inhibitor | ChEMBL | Indication | All | 2 | Erdheim-chester disease | No |
| P08887 | CHEMBL2364155 | IL6RA | Drugged | Single protein | CHEMBL1237022 | Tocilizumab | Antibody | Inhibitor | ChEMBL | Indication | All | 2 | Graves ophthalmopathy | No |
| P08887 | CHEMBL2364155 | IL6RA | Drugged | Single protein | CHEMBL1237022 | Tocilizumab | Antibody | Inhibitor | ChEMBL | Indication | All | 2 | Lymphohistiocytosis, hemophagocytic | No |
| P08887 | CHEMBL2364155 | IL6RA | Drugged | Single protein | CHEMBL1237022 | Tocilizumab | Antibody | Inhibitor | ChEMBL | Indication | All | 2 | Castleman disease | No |
| P08887 | CHEMBL2364155 | IL6RA | Drugged | Single protein | CHEMBL1237022 | Tocilizumab | Antibody | Inhibitor | ChEMBL | Indication | All | 3 | Epilepsy | No |
| P08887 | CHEMBL2364155 | IL6RA | Drugged | Single protein | CHEMBL1237022 | Tocilizumab | Antibody | Inhibitor | ChEMBL | Indication | All | 2 | Still's disease, adult-onset | No |
| P08887 | CHEMBL2364155 | IL6RA | Drugged | Single protein | CHEMBL1237022 | Tocilizumab | Antibody | Inhibitor | ChEMBL | Indication | All | 3 | Influenza, human | No |
| P08887 | CHEMBL2364155 | IL6RA | Drugged | Single protein | CHEMBL1237022 | Tocilizumab | Antibody | Inhibitor | ChEMBL | Indication | All | 2 | Lung diseases | No |
| P08887 | CHEMBL2364155 | IL6RA | Drugged | Single protein | CHEMBL1237022 | Tocilizumab | Antibody | Inhibitor | ChEMBL | Indication | All | 2 | Graft vs host disease | No |
| P08887 | CHEMBL2364155 | IL6RA | Drugged | Single protein | CHEMBL1237022 | Tocilizumab | Antibody | Inhibitor | ChEMBL | Indication | All | 2 | Fever | No |
| P08887 | CHEMBL2364155 | IL6RA | Drugged | Single protein | CHEMBL1237022 | Tocilizumab | Antibody | Inhibitor | ChEMBL | Indication | All | 2 | Diabetes mellitus, type 1 | No |
| P08887 | CHEMBL2364155 | IL6RA | Drugged | Single protein | CHEMBL1237022 | Tocilizumab | Antibody | Inhibitor | ChEMBL | Indication | All | 2 | Dermatomyositis | No |
| P08887 | CHEMBL2364155 | IL6RA | Drugged | Single protein | CHEMBL1237022 | Tocilizumab | Antibody | Inhibitor | ChEMBL | Indication | All | 2 | Depressive disorder | No |
| P08887 | CHEMBL2364155 | IL6RA | Drugged | Single protein | CHEMBL1237022 | Tocilizumab | Antibody | Inhibitor | ChEMBL | Indication | All | 2 | Cancers | No |
| P08887 | CHEMBL2364155 | IL6RA | Drugged | Single protein | CHEMBL1237022 | Tocilizumab | Antibody | Inhibitor | ChEMBL | Indication | All | 2 | Behcet syndrome | No |
| P08887 | CHEMBL2364155 | IL6RA | Drugged | Single protein | CHEMBL1237022 | Tocilizumab | Antibody | Inhibitor | ChEMBL | Indication | All | 2 | Pulmonary hypertension | Yes |
| P08887 | CHEMBL2364155 | IL6RA | Drugged | Single protein | CHEMBL1237022 | Tocilizumab | Antibody | Inhibitor | ChEMBL | Indication | All | 2 | Amyotrophic lateral sclerosis | No |
| P08887 | CHEMBL2364155 | IL6RA | Drugged | Single protein | CHEMBL1237022 | Tocilizumab | Antibody | Inhibitor | ChEMBL | Indication | All | 1 | Renal insufficiency | No |
| P08887 | CHEMBL2364155 | IL6RA | Drugged | Single protein | CHEMBL1237022 | Tocilizumab | Antibody | Inhibitor | ChEMBL | Indication | All | 1 | Uveitis | No |
| P08887 | CHEMBL2364155 | IL6RA | Drugged | Single protein | CHEMBL1237022 | Tocilizumab | Antibody | Inhibitor | ChEMBL | Indication | All | 1 | Hiv infection | No |
| P08887 | CHEMBL2364155 | IL6RA | Drugged | Single protein | CHEMBL1237022 | Tocilizumab | Antibody | Inhibitor | ChEMBL | Indication | All | 1 | Schizophrenia | No |
| P08887 | CHEMBL2364155 | IL6RA | Drugged | Single protein | CHEMBL1237022 | Tocilizumab | Antibody | Inhibitor | ChEMBL | Indication | All | 1 | Lupus erythematosus, systemic | No |
| P08887 | CHEMBL2364155 | IL6RA | Drugged | Single protein | CHEMBL1237022 | Tocilizumab | Antibody | Inhibitor | ChEMBL | Indication | All | 1 | Cancers | No |
| P08887 | CHEMBL2364155 | IL6RA | Drugged | Single protein | CHEMBL1237022 | Tocilizumab | Antibody | Inhibitor | ChEMBL | Indication | All | 2 | Non-st elevated myocardial infarction | Yes |
| P08887 | CHEMBL2364155 | IL6RA | Drugged | Single protein | CHEMBL1237022 | Tocilizumab | Antibody | Inhibitor | ChEMBL | Indication | All | 3 | Cancers | No |
| P08887 | CHEMBL2364155 | IL6RA | Drugged | Single protein | CHEMBL1237022 | Tocilizumab | Antibody | Inhibitor | ChEMBL | Indication | All | 2 | Macrophage activation syndrome | No |
| P08887 | CHEMBL2364155 | IL6RA | Drugged | Single protein | CHEMBL1237022 | Tocilizumab | Antibody | Inhibitor | ChEMBL | Indication | All | 3 | Pneumonia | No |
| P08887 | CHEMBL2364155 | IL6RA | Drugged | Single protein | CHEMBL2108730 | Sarilumab | Antibody | Antagonist | ChEMBL | Indication | All | 3 | Pneumonia | No |
| P08887 | CHEMBL2364155 | IL6RA | Drugged | Single protein | CHEMBL2108730 | Sarilumab | Antibody | Antagonist | ChEMBL | Indication | All | 3 | Giant cell arteritis | No |
| P08887 | CHEMBL2364155 | IL6RA | Drugged | Single protein | CHEMBL2108730 | Sarilumab | Antibody | Antagonist | ChEMBL | Indication | All | 3 | Severe acute respiratory syndrome | No |
| P08887 | CHEMBL2364155 | IL6RA | Drugged | Single protein | CHEMBL2108730 | Sarilumab | Antibody | Antagonist | ChEMBL | Indication | All | 4 | Arthritis | No |
| P08887 | CHEMBL2364155 | IL6RA | Drugged | Single protein | CHEMBL2108730 | Sarilumab | Antibody | Antagonist | ChEMBL | Indication | All | 4 | Immune system diseases | No |
| P08887 | CHEMBL2364155 | IL6RA | Drugged | Single protein | CHEMBL3833343 | Vobarilizumab | Antibody | Inhibitor | ChEMBL | Indication | All | 2 | Arthritis | No |
| P08887 | CHEMBL2364155 | IL6RA | Drugged | Single protein | CHEMBL3833343 | Vobarilizumab | Antibody | Inhibitor | ChEMBL | Indication | All | 2 | Lupus erythematosus, systemic | No |
| P08887 | CHEMBL2364155 | IL6RA | Drugged | Single protein | CHEMBL2108730 | Sarilumab | Antibody | Antagonist | ChEMBL | Indication | All | 3 | Influenza, human | No |
| P08887 | CHEMBL2364155 | IL6RA | Drugged | Single protein | CHEMBL4650406 | Levilimab | Antibody | Inhibitor | ChEMBL | Indication | All | 3 | Arthritis | No |
| P08887 | CHEMBL2364155 | IL6RA | Drugged | Single protein | CHEMBL2108730 | Sarilumab | Antibody | Antagonist | BNF | Indication | All | 4 | Arthritis | No |
| P08887 | CHEMBL2364155 | IL6RA | Drugged | Single protein | CHEMBL2108730 | Sarilumab | Antibody | Antagonist | BNF | Side-effect | Common or very common | 4 | Dyslipidemia | No |
| P08887 | CHEMBL2364155 | IL6RA | Drugged | Single protein | CHEMBL2108730 | Sarilumab | Antibody | Antagonist | BNF | Side-effect | Common or very common | 4 | Increased risk of infection | No |
| P08887 | CHEMBL2364155 | IL6RA | Drugged | Single protein | CHEMBL2108730 | Sarilumab | Antibody | Antagonist | BNF | Side-effect | Common or very common | 4 | Neutropenia | No |
| P08887 | CHEMBL2364155 | IL6RA | Drugged | Single protein | CHEMBL2108730 | Sarilumab | Antibody | Antagonist | BNF | Side-effect | Common or very common | 4 | Thrombocytopenia | No |
| P08887 | CHEMBL2364155 | IL6RA | Drugged | Single protein | CHEMBL2108730 | Sarilumab | Antibody | Antagonist | BNF | Side-effect | Not known | 4 | Hypersensitivity | No |
| P08887 | CHEMBL2364155 | IL6RA | Drugged | Single protein | CHEMBL2108730 | Sarilumab | Antibody | Antagonist | BNF | Side-effect | Not known | 4 | Skin reactions | No |
| P08887 | CHEMBL2364155 | IL6RA | Drugged | Single protein | CHEMBL4650406 | Levilimab | Antibody | Inhibitor | ChEMBL | Indication | All | 3 | Severe acute respiratory syndrome | No |

| Table S8. Druggability results of the prioritised proteins | | | | | | | | | | | | | | |
|---|---|---|---|---|---|---|---|---|---|---|---|---|---|---|
| Target Uniprot ID* | Target ChEMBL ID | Target protein name | Druggability* | Target type | Drug ChEMBL ID | Drug name | Drug molecule type | Drug mechanism | Drug effect source | Drug effect type | Drug effect frequency | Max phase* | Drug effect | Drug effect class |
| P08887 | CHEMBL2364155 | IL6RA | Drugged | Single protein | CHEMBL2108730 | Sarilumab | Antibody | Antagonist | ChEMBL | Indication | All | 2 | Mastocytosis | No |
| P08887 | CHEMBL2364155 | IL6RA | Drugged | Single protein | CHEMBL2108730 | Sarilumab | Antibody | Antagonist | ChEMBL | Indication | All | 3 | Polymyalgia rheumatica | No |
| P08887 | CHEMBL2364155 | IL6RA | Drugged | Single protein | CHEMBL2108730 | Sarilumab | Antibody | Antagonist | ChEMBL | Indication | All | 2 | Spondylitis, ankylosing | No |
| P08887 | CHEMBL2364155 | IL6RA | Drugged | Single protein | CHEMBL1237022 | Tocilizumab | Antibody | Inhibitor | ChEMBL | Indication | All | 3 | Polymyalgia rheumatica | No |
| P08887 | CHEMBL2364155 | IL6RA | Drugged | Single protein | CHEMBL1237022 | Tocilizumab | Antibody | Inhibitor | ChEMBL | Indication | All | 3 | Spondylitis, ankylosing | No |
| P08887 | CHEMBL2364155 | IL6RA | Drugged | Single protein | CHEMBL1237022 | Tocilizumab | Antibody | Inhibitor | ChEMBL | Indication | All | 3 | Takayasu arteritis | No |
| P08887 | CHEMBL2364155 | IL6RA | Drugged | Single protein | CHEMBL1237022 | Tocilizumab | Antibody | Inhibitor | ChEMBL | Indication | All | 3 | Severe acute respiratory syndrome | No |
| P08887 | CHEMBL2364155 | IL6RA | Drugged | Single protein | CHEMBL1237022 | Tocilizumab | Antibody | Inhibitor | ChEMBL | Indication | All | 4 | Arthritis | No |
| P08887 | CHEMBL2364155 | IL6RA | Drugged | Single protein | CHEMBL1237022 | Tocilizumab | Antibody | Inhibitor | ChEMBL | Indication | All | 4 | Immune system diseases | No |
| P08887 | CHEMBL2364155 | IL6RA | Drugged | Single protein | CHEMBL1237022 | Tocilizumab | Antibody | Inhibitor | ChEMBL | Indication | All | 4 | Scleroderma | No |
| P08887 | CHEMBL2364155 | IL6RA | Drugged | Single protein | CHEMBL1237022 | Tocilizumab | Antibody | Inhibitor | ChEMBL | Indication | All | 4 | Giant cell arteritis | No |
| P08887 | CHEMBL2364155 | IL6RA | Drugged | Single protein | CHEMBL2108730 | Sarilumab | Antibody | Antagonist | ChEMBL | Indication | All | 1 | Scleroderma | No |
| P08887 | CHEMBL2364155 | IL6RA | Drugged | Single protein | CHEMBL2108730 | Sarilumab | Antibody | Antagonist | ChEMBL | Indication | All | 2 | Arthritis | No |
| P08887 | CHEMBL2364155 | IL6RA | Drugged | Single protein | CHEMBL2108730 | Sarilumab | Antibody | Antagonist | ChEMBL | Indication | All | 2 | Uveitis | No |
| P08887 | CHEMBL2364155 | IL6RA | Drugged | Single protein | CHEMBL1237022 | Tocilizumab | Antibody | Inhibitor | ChEMBL | Indication | All | 2 | Fibrous dysplasia of bone | No |
| P08887 | CHEMBL2364155 | IL6RA | Drugged | Single protein | CHEMBL2108730 | Sarilumab | Antibody | Antagonist | ChEMBL | Indication | All | 2 | Cancers | No |
| P20155 | - | ISK2 | Not yet druggable | - | - | - | - | - | - | - | - | - | - | No |
| Q07866 | - | KLC1 | Not yet druggable | - | - | - | - | - | - | - | - | - | - | No |
| P17252 | CHEMBL2093867 | KPCA | Drugged | Protein family | CHEMBL608533 | Midostaurin | Small molecule | Inhibitor | BNF | Side-effect | Common or very common | 4 | Diarrhoea | No |
| P17252 | CHEMBL2093867 | KPCA | Drugged | Protein family | CHEMBL608533 | Midostaurin | Small molecule | Inhibitor | BNF | Side-effect | Common or very common | 4 | Cystitis | No |
| P17252 | CHEMBL2093867 | KPCA | Drugged | Protein family | CHEMBL608533 | Midostaurin | Small molecule | Inhibitor | BNF | Side-effect | Common or very common | 4 | Cough | No |
| P17252 | CHEMBL2093867 | KPCA | Drugged | Protein family | CHEMBL608533 | Midostaurin | Small molecule | Inhibitor | BNF | Side-effect | Common or very common | 4 | Constipation | No |
| P17252 | CHEMBL2093867 | KPCA | Drugged | Protein family | CHEMBL608533 | Midostaurin | Small molecule | Inhibitor | BNF | Indication | All | 4 | Aggressive systemic mastocytosis | No |
| P17252 | CHEMBL2093867 | KPCA | Drugged | Protein family | CHEMBL608533 | Midostaurin | Small molecule | Inhibitor | BNF | Side-effect | Common or very common | 4 | Chills | No |
| P17252 | CHEMBL2093867 | KPCA | Drugged | Protein family | CHEMBL608533 | Midostaurin | Small molecule | Inhibitor | BNF | Indication | All | 4 | Cancers | No |
| P17252 | CHEMBL2093867 | KPCA | Drugged | Protein family | CHEMBL608533 | Midostaurin | Small molecule | Inhibitor | BNF | Side-effect | Common or very common | 4 | Asthenia | No |
| P17252 | CHEMBL2093867 | KPCA | Drugged | Protein family | CHEMBL608533 | Midostaurin | Small molecule | Inhibitor | BNF | Side-effect | Common or very common | 4 | Bruising | No |
| P17252 | CHEMBL2093867 | KPCA | Drugged | Protein family | CHEMBL608533 | Midostaurin | Small molecule | Inhibitor | BNF | Indication | All | 4 | Leukemia | No |
| P17252 | CHEMBL2093867 | KPCA | Drugged | Protein family | CHEMBL608533 | Midostaurin | Small molecule | Inhibitor | BNF | Side-effect | Common or very common | 4 | Vomiting | No |
| P17252 | CHEMBL2093867 | KPCA | Drugged | Protein family | CHEMBL608533 | Midostaurin | Small molecule | Inhibitor | BNF | Side-effect | Common or very common | 4 | Dyspepsia | No |
| P17252 | CHEMBL2093867 | KPCA | Drugged | Protein family | CHEMBL608533 | Midostaurin | Small molecule | Inhibitor | ChEMBL | Indication | All | 4 | Mastocytosis | No |
| P17252 | CHEMBL2093867 | KPCA | Drugged | Protein family | CHEMBL608533 | Midostaurin | Small molecule | Inhibitor | BNF | Side-effect | Not known | 4 | Congestive heart failure | Yes |
| P17252 | CHEMBL2093867 | KPCA | Drugged | Protein family | CHEMBL608533 | Midostaurin | Small molecule | Inhibitor | BNF | Side-effect | Not known | 4 | Qt interval prolongation | Yes |
| P17252 | CHEMBL2093867 | KPCA | Drugged | Protein family | CHEMBL608533 | Midostaurin | Small molecule | Inhibitor | BNF | Side-effect | Not known | 4 | Cardiac disorder | Yes |
| P17252 | CHEMBL2093867 | KPCA | Drugged | Protein family | CHEMBL608533 | Midostaurin | Small molecule | Inhibitor | BNF | Side-effect | Common or very common | 4 | Weight increased | No |
| P17252 | CHEMBL2093867 | KPCA | Drugged | Protein family | CHEMBL608533 | Midostaurin | Small molecule | Inhibitor | BNF | Side-effect | Common or very common | 4 | Vertigo | No |
| P17252 | CHEMBL2093867 | KPCA | Drugged | Protein family | CHEMBL608533 | Midostaurin | Small molecule | Inhibitor | BNF | Side-effect | Common or very common | 4 | Tremor | No |
| P17252 | CHEMBL2093867 | KPCA | Drugged | Protein family | CHEMBL608533 | Midostaurin | Small molecule | Inhibitor | BNF | Side-effect | Common or very common | 4 | Sepsis | No |
| P17252 | CHEMBL2093867 | KPCA | Drugged | Protein family | CHEMBL608533 | Midostaurin | Small molecule | Inhibitor | BNF | Side-effect | Common or very common | 4 | Respiratory disorder | No |
| P17252 | CHEMBL2093867 | KPCA | Drugged | Protein family | CHEMBL608533 | Midostaurin | Small molecule | Inhibitor | BNF | Side-effect | Common or very common | 4 | Oropharyngeal pain | No |
| P17252 | CHEMBL2093867 | KPCA | Drugged | Protein family | CHEMBL608533 | Midostaurin | Small molecule | Inhibitor | BNF | Side-effect | Common or very common | 4 | Dizziness | No |
| P17252 | CHEMBL2093867 | KPCA | Drugged | Protein family | CHEMBL608533 | Midostaurin | Small molecule | Inhibitor | BNF | Side-effect | Common or very common | 4 | Oedema | No |
| P17252 | CHEMBL2093867 | KPCA | Drugged | Protein family | CHEMBL608533 | Midostaurin | Small molecule | Inhibitor | BNF | Side-effect | Common or very common | 4 | Increased risk of infection | No |
| P17252 | CHEMBL2093867 | KPCA | Drugged | Protein family | CHEMBL608533 | Midostaurin | Small molecule | Inhibitor | BNF | Side-effect | Common or very common | 4 | Hypotension | No |
| P17252 | CHEMBL2093867 | KPCA | Drugged | Protein family | CHEMBL608533 | Midostaurin | Small molecule | Inhibitor | BNF | Side-effect | Common or very common | 4 | Hypersensitivity | No |
| P17252 | CHEMBL2093867 | KPCA | Drugged | Protein family | CHEMBL608533 | Midostaurin | Small molecule | Inhibitor | BNF | Side-effect | Common or very common | 4 | Hyperglycemia | No |
| P17252 | CHEMBL2093867 | KPCA | Drugged | Protein family | CHEMBL608533 | Midostaurin | Small molecule | Inhibitor | BNF | Side-effect | Common or very common | 4 | Headache | No |
| P17252 | CHEMBL2093867 | KPCA | Drugged | Protein family | CHEMBL608533 | Midostaurin | Small molecule | Inhibitor | BNF | Side-effect | Common or very common | 4 | Hemorrhage | No |
| P17252 | CHEMBL2093867 | KPCA | Drugged | Protein family | CHEMBL608533 | Midostaurin | Small molecule | Inhibitor | BNF | Side-effect | Common or very common | 4 | Fever | No |
| P17252 | CHEMBL2093867 | KPCA | Drugged | Protein family | CHEMBL608533 | Midostaurin | Small molecule | Inhibitor | BNF | Side-effect | Common or very common | 4 | Febrile neutropenia | No |
| P17252 | CHEMBL2093867 | KPCA | Drugged | Protein family | CHEMBL608533 | Midostaurin | Small molecule | Inhibitor | BNF | Side-effect | Common or very common | 4 | Fall | No |
| P17252 | CHEMBL2093867 | KPCA | Drugged | Protein family | CHEMBL608533 | Midostaurin | Small molecule | Inhibitor | BNF | Side-effect | Common or very common | 4 | Dyspnoea | No |
| P17252 | CHEMBL2093867 | KPCA | Drugged | Protein family | CHEMBL608533 | Midostaurin | Small molecule | Inhibitor | BNF | Side-effect | Common or very common | 4 | Nausea | No |
| P17252 | CHEMBL2093867 | KPCA | Drugged | Protein family | CHEMBL608533 | Midostaurin | Small molecule | Inhibitor | ChEMBL | Indication | All | 4 | Cancers | No |
| P17252 | CHEMBL2093867 | KPCA | Drugged | Protein family | CHEMBL608533 | Midostaurin | Small molecule | Inhibitor | BNF | Side-effect | Common or very common | 4 | Concentration impaired | No |
| P17252 | CHEMBL2093867 | KPCA | Drugged | Protein family | CHEMBL608533 | Midostaurin | Small molecule | Inhibitor | ChEMBL | Indication | All | 2 | Cancers | No |
| P17252 | CHEMBL2093867 | KPCA | Drugged | Protein family | CHEMBL494089 | Gsk-690693 | Small molecule | Inhibitor | ChEMBL | Indication | All | 1 | Cancers | No |
| P17252 | CHEMBL2093867 | KPCA | Drugged | Protein family | CHEMBL3545332 | Cep-2563 | Small molecule | Inhibitor | ChEMBL | Indication | All | - | - | No |
| P17252 | CHEMBL2093867 | KPCA | Drugged | Protein family | CHEMBL565612 | Sotrastaurin | Small molecule | Inhibitor | ChEMBL | Indication | All | 1 | Cancers | No |
| P17252 | CHEMBL2093867 | KPCA | Drugged | Protein family | CHEMBL565612 | Sotrastaurin | Small molecule | Inhibitor | ChEMBL | Indication | All | 2 | Colitis, ulcerative | No |
| P17252 | CHEMBL2093867 | KPCA | Drugged | Protein family | CHEMBL565612 | Sotrastaurin | Small molecule | Inhibitor | ChEMBL | Indication | All | 2 | Psoriasis | No |
| P17252 | CHEMBL2093867 | KPCA | Drugged | Protein family | CHEMBL608533 | Midostaurin | Small molecule | Inhibitor | ChEMBL | Indication | All | 3 | Cancers | No |
| P17252 | CHEMBL2093867 | KPCA | Drugged | Protein family | CHEMBL574737 | Ucn-01 | Small molecule | Inhibitor | ChEMBL | Indication | All | 1 | Cancers | No |
| P17252 | CHEMBL2093867 | KPCA | Drugged | Protein family | CHEMBL574737 | Ucn-01 | Small molecule | Inhibitor | ChEMBL | Indication | All | 2 | Cancers | No |
| P17252 | CHEMBL2093867 | KPCA | Drugged | Protein family | CHEMBL608533 | Midostaurin | Small molecule | Inhibitor | ChEMBL | Indication | All | 1 | Liver diseases | No |
| P17252 | CHEMBL2093867 | KPCA | Drugged | Protein family | CHEMBL608533 | Midostaurin | Small molecule | Inhibitor | ChEMBL | Indication | All | 1 | Cancers | No |
| P17252 | CHEMBL2093867 | KPCA | Drugged | Protein family | CHEMBL608533 | Midostaurin | Small molecule | Inhibitor | ChEMBL | Indication | All | 2 | Blast crisis | No |
| P17252 | CHEMBL2093867 | KPCA | Drugged | Protein family | CHEMBL565612 | Sotrastaurin | Small molecule | Inhibitor | ChEMBL | Indication | All | 2 | Uveitis | No |
| O75023 | - | LIRB5 | Not yet druggable | - | - | - | - | - | - | - | - | - | - | No |
| O15232 | - | MATN3 | Not yet druggable | - | - | - | - | - | - | - | - | - | - | No |
| P61244 | CHEMBL4106127 | MAX | Druggable | Protein-protein interaction | - | - | - | - | - | - | - | - | - | No |
| P61244 | CHEMBL3301395 | MAX | Druggable | Protein complex | - | - | - | - | - | - | - | - | - | No |
| P61244 | CHEMBL1250363 | MAX | Druggable | Single protein | - | - | - | - | - | - | - | - | - | No |
| Q29983 | - | MICA | Not yet druggable | - | - | - | - | - | - | - | - | - | - | No |
| Q29980 | - | MICB | Not yet druggable | - | - | - | - | - | - | - | - | - | - | No |
| Q9UNW1 | - | MINP1 | Not yet druggable | - | - | - | - | - | - | - | - | - | - | No |
| Q13508 | - | NAR3 | Not yet druggable | - | - | - | - | - | - | - | - | - | - | No |
| O14931 | - | NCTR3 | Not yet druggable | - | - | - | - | - | - | - | - | - | - | No |
| P29120 | CHEMBL3182 | NEC1 | Druggable | Single protein | - | - | - | - | - | - | - | - | - | No |
| P15559 | CHEMBL3623 | NQO1 | Drugged | Single protein | CHEMBL71595 | Hydrogen peroxide | Small molecule | - | ChEMBL | Indication | All | 4 | Gingivitis | No |
| P15559 | CHEMBL3623 | NQO1 | Drugged | Single protein | CHEMBL1812161 | Vatiquinone | Small molecule | Modulator | ChEMBL | Indication | All | 1 | Tourette syndrome | No |
| P15559 | CHEMBL3623 | NQO1 | Drugged | Single protein | CHEMBL1812161 | Vatiquinone | Small molecule | Modulator | ChEMBL | Indication | All | 1 | Muscular diseases | No |
| P15559 | CHEMBL3623 | NQO1 | Drugged | Single protein | CHEMBL71595 | Hydrogen peroxide | Small molecule | - | ChEMBL | Indication | All | 0,5 | Dental caries | No |
| P15559 | CHEMBL3623 | NQO1 | Drugged | Single protein | CHEMBL71595 | Hydrogen peroxide | Small molecule | - | ChEMBL | Indication | All | 4 | Keratosis, seborrheic | No |
| P15559 | CHEMBL3623 | NQO1 | Drugged | Single protein | CHEMBL71595 | Hydrogen peroxide | Small molecule | - | ChEMBL | Indication | All | 2 | Cancers | No |
| P15559 | CHEMBL3623 | NQO1 | Drugged | Single protein | CHEMBL71595 | Hydrogen peroxide | Small molecule | - | ChEMBL | Indication | All | 2 | Acne vulgaris | No |
| P15559 | CHEMBL3623 | NQO1 | Drugged | Single protein | CHEMBL1812161 | Vatiquinone | Small molecule | Modulator | ChEMBL | Indication | All | 2 | Rett syndrome | No |

**Table S8. Druggability results of the prioritised proteins**

| Target Uniprot ID* | Target ChEMBL ID | Target protein name | Druggability* | Target type | Drug ChEMBL ID | Drug name | Drug molecule type | Drug mechanism | Drug effect source | Drug effect type | Drug effect frequency | Max phase* | Drug effect | Drug effect class |
|---|---|---|---|---|---|---|---|---|---|---|---|---|---|---|
| P15559 | CHEMBL3623 | NQO1 | Drugged | Single protein | CHEMBL1812161 | Vatiquinone | Small molecule | Modulator | ChEMBL | Indication | All | 2 | Parkinson disease | No |
| P15559 | CHEMBL3623 | NQO1 | Drugged | Single protein | CHEMBL1812161 | Vatiquinone | Small molecule | Modulator | ChEMBL | Indication | All | 2 | Leigh disease | No |
| P15559 | CHEMBL3623 | NQO1 | Drugged | Single protein | CHEMBL1812161 | Vatiquinone | Small molecule | Modulator | ChEMBL | Indication | All | 3 | Mitochondrial diseases | No |
| P15559 | CHEMBL3623 | NQO1 | Drugged | Single protein | CHEMBL71595 | Hydrogen peroxide | Small molecule | - | ChEMBL | Indication | All | 1 | Wounds and injuries | No |
| P15559 | CHEMBL3623 | NQO1 | Drugged | Single protein | CHEMBL71595 | Hydrogen peroxide | Small molecule | - | ChEMBL | Indication | All | 3 | Tooth diseases | No |
| P15559 | CHEMBL3623 | NQO1 | Drugged | Single protein | CHEMBL71595 | Hydrogen peroxide | Small molecule | - | ChEMBL | Indication | All | 3 | Warts | No |
| P15559 | CHEMBL3623 | NQO1 | Drugged | Single protein | CHEMBL1812161 | Vatiquinone | Small molecule | Modulator | ChEMBL | Indication | All | 2 | Hearing loss | No |
| P15559 | CHEMBL3623 | NQO1 | Drugged | Single protein | CHEMBL71595 | Hydrogen peroxide | Small molecule | - | ChEMBL | Indication | All | 4 | Infections | No |
| P15559 | CHEMBL3623 | NQO1 | Drugged | Single protein | CHEMBL71595 | Hydrogen peroxide | Small molecule | - | ChEMBL | Indication | All | 2 | Severe acute respiratory syndrome | No |
| P15559 | CHEMBL3623 | NQO1 | Drugged | Single protein | CHEMBL71595 | Hydrogen peroxide | Small molecule | - | ChEMBL | Indication | All | 4 | Pharyngitis | No |
| P15559 | CHEMBL3623 | NQO1 | Drugged | Single protein | CHEMBL1466 | Dicumarol | Small molecule | Inhibitor | ChEMBL | Indication | All | 4 | Thrombosis | No |
| P15559 | CHEMBL3623 | NQO1 | Drugged | Single protein | CHEMBL590 | Menadione | Small molecule | - | ChEMBL | Indication | All | 4 | Hemorrhage | No |
| P15559 | CHEMBL3623 | NQO1 | Drugged | Single protein | CHEMBL1812161 | Vatiquinone | Small molecule | Modulator | ChEMBL | Indication | All | 2 | Child development disorder, pervasive | No |
| P15559 | CHEMBL3623 | NQO1 | Drugged | Single protein | CHEMBL1812161 | Vatiquinone | Small molecule | Modulator | ChEMBL | Indication | All | 2 | Friedreich ataxia | No |
| Q9BW91 | CHEMBL4105984 | NUDT9 | Druggable | Single protein | - | - | - | - | - | - | - | - | - | No |
| Q14554 | - | PDIA5 | Not yet druggable | - | - | - | - | - | - | - | - | - | - | No |
| Q13018 | CHEMBL3713395 | PLA2R | Druggable | Single protein | - | - | - | - | - | - | - | - | - | No |
| Q9UIW2 | - | PLXA1 | Not yet druggable | - | - | - | - | - | - | - | - | - | - | No |
| O15031 | - | PLXB2 | Not yet druggable | - | - | - | - | - | - | - | - | - | - | No |
| Q96NY8 | CHEMBL3712928 | PVRL4 | Drugged | Single protein | CHEMBL3301589 | Enfortumab vedotin | Antibody | Binding agent | ChEMBL | Indication | All | 4 | Cancers | No |
| Q96NY8 | CHEMBL3712928 | PVRL4 | Drugged | Single protein | CHEMBL3301589 | Enfortumab vedotin | Antibody | Binding agent | ChEMBL | Indication | All | 2 | Cancers | No |
| Q96NY8 | CHEMBL3712928 | PVRL4 | Drugged | Single protein | CHEMBL3301589 | Enfortumab vedotin | Antibody | Binding agent | ChEMBL | Indication | All | 3 | Cancers | No |
| Q6UW15 | - | REG3G | Not yet druggable | - | - | - | - | - | - | - | - | - | - | No |
| P07949 | CHEMBL2041 | RET | Drugged | Single protein | CHEMBL572881 | Motesanib | Small molecule | Inhibitor | ChEMBL | Indication | All | 3 | Cancers | No |
| P07949 | CHEMBL2041 | RET | Drugged | Single protein | CHEMBL1946170 | Regorafenib | Small molecule | Inhibitor | BNF | Side-effect | Common or very common | 4 | Appetite decreased | No |
| P07949 | CHEMBL2041 | RET | Drugged | Single protein | CHEMBL576982 | Quizartinib | Small molecule | Inhibitor | ChEMBL | Indication | All | 1 | Liver diseases | No |
| P07949 | CHEMBL2041 | RET | Drugged | Single protein | CHEMBL576982 | Quizartinib | Small molecule | Inhibitor | ChEMBL | Indication | All | 3 | Cancers | No |
| P07949 | CHEMBL2041 | RET | Drugged | Single protein | CHEMBL1946170 | Regorafenib | Small molecule | Inhibitor | BNF | Side-effect | Common or very common | 4 | Anemia | No |
| P07949 | CHEMBL2041 | RET | Drugged | Single protein | CHEMBL603469 | Lestaurtinib | Small molecule | Inhibitor | ChEMBL | Indication | All | 1 | Cancers | No |
| P07949 | CHEMBL2041 | RET | Drugged | Single protein | CHEMBL603469 | Lestaurtinib | Small molecule | Inhibitor | ChEMBL | Indication | All | 1 | Primary myelofibrosis | No |
| P07949 | CHEMBL2041 | RET | Drugged | Single protein | CHEMBL603469 | Lestaurtinib | Small molecule | Inhibitor | ChEMBL | Indication | All | 2 | Cancers | No |
| P07949 | CHEMBL2041 | RET | Drugged | Single protein | CHEMBL1946170 | Regorafenib | Small molecule | Inhibitor | BNF | Side-effect | Common or very common | 4 | Alopecia | No |
| P07949 | CHEMBL2041 | RET | Drugged | Single protein | CHEMBL1946170 | Regorafenib | Small molecule | Inhibitor | BNF | Indication | All | 4 | Metastatic colorectal cancer | No |
| P07949 | CHEMBL2041 | RET | Drugged | Single protein | CHEMBL4650296 | Bos-589 | Unknown | Inhibitor | ChEMBL | Indication | All | 2 | Irritable bowel syndrome | No |
| P07949 | CHEMBL2041 | RET | Drugged | Single protein | CHEMBL4582651 | Pralsetinib | Small molecule | Inhibitor | ChEMBL | Indication | All | 4 | Cancers | No |
| P07949 | CHEMBL2041 | RET | Drugged | Single protein | CHEMBL603469 | Lestaurtinib | Small molecule | Inhibitor | ChEMBL | Indication | All | 2 | Polycythemia vera | No |
| P07949 | CHEMBL2041 | RET | Drugged | Single protein | CHEMBL4559134 | Selpercatinib | Small molecule | Inhibitor | ChEMBL | Indication | All | 4 | Cancers | No |
| P07949 | CHEMBL2041 | RET | Drugged | Single protein | CHEMBL4559134 | Selpercatinib | Small molecule | Inhibitor | ChEMBL | Indication | All | 1 | Renal insufficiency | No |
| P07949 | CHEMBL2041 | RET | Drugged | Single protein | CHEMBL603469 | Lestaurtinib | Small molecule | Inhibitor | ChEMBL | Indication | All | 2 | Psoriasis | No |
| P07949 | CHEMBL2041 | RET | Drugged | Single protein | CHEMBL4559134 | Selpercatinib | Small molecule | Inhibitor | ChEMBL | Indication | All | 1 | Liver diseases | No |
| P07949 | CHEMBL2041 | RET | Drugged | Single protein | CHEMBL603469 | Lestaurtinib | Small molecule | Inhibitor | ChEMBL | Indication | All | 2 | Thrombocythemia, essential | No |
| P07949 | CHEMBL2041 | RET | Drugged | Single protein | CHEMBL572881 | Motesanib | Small molecule | Inhibitor | ChEMBL | Indication | All | 1 | Cancers | No |
| P07949 | CHEMBL2041 | RET | Drugged | Single protein | CHEMBL1946170 | Regorafenib | Small molecule | Inhibitor | BNF | Indication | All | 4 | Cancers | No |
| P07949 | CHEMBL2041 | RET | Drugged | Single protein | CHEMBL1567 | Sunitinib malate | Small molecule | Inhibitor | ChEMBL | Indication | All | 2 | Cancers | No |
| P07949 | CHEMBL2041 | RET | Drugged | Single protein | CHEMBL2029988 | Cep-32496 | Small molecule | Inhibitor | ChEMBL | Indication | All | 1 | Cancers | No |
| P07949 | CHEMBL2041 | RET | Drugged | Single protein | CHEMBL1946170 | Regorafenib | Small molecule | Inhibitor | ChEMBL | Indication | All | 1 | Cancers | No |
| P07949 | CHEMBL2041 | RET | Drugged | Single protein | CHEMBL1946170 | Regorafenib | Small molecule | Inhibitor | ChEMBL | Indication | All | 2 | Cancers | No |
| P07949 | CHEMBL2041 | RET | Drugged | Single protein | CHEMBL1946170 | Regorafenib | Small molecule | Inhibitor | ChEMBL | Indication | All | 2 | Macular degeneration | No |
| P07949 | CHEMBL2041 | RET | Drugged | Single protein | CHEMBL1946170 | Regorafenib | Small molecule | Inhibitor | ChEMBL | Indication | All | 2 | Pulmonary disease, chronic obstructive | No |
| P07949 | CHEMBL2041 | RET | Drugged | Single protein | CHEMBL1946170 | Regorafenib | Small molecule | Inhibitor | ChEMBL | Indication | All | 3 | Cancers | No |
| P07949 | CHEMBL2041 | RET | Drugged | Single protein | CHEMBL1946170 | Regorafenib | Small molecule | Inhibitor | ChEMBL | Indication | All | 4 | Cancers | No |
| P07949 | CHEMBL2041 | RET | Drugged | Single protein | CHEMBL1200485 | Sorafenib tosylate | Small molecule | Inhibitor | ChEMBL | Indication | All | 1 | Cancers | No |
| P07949 | CHEMBL2041 | RET | Drugged | Single protein | CHEMBL1200485 | Sorafenib tosylate | Small molecule | Inhibitor | ChEMBL | Indication | All | 2 | Cancers | No |
| P07949 | CHEMBL2041 | RET | Drugged | Single protein | CHEMBL1200485 | Sorafenib tosylate | Small molecule | Inhibitor | ChEMBL | Indication | All | 2 | Retroviridae infection | No |
| P07949 | CHEMBL2041 | RET | Drugged | Single protein | CHEMBL1946170 | Regorafenib | Small molecule | Inhibitor | BNF | Side-effect | Common or very common | 4 | Asthenia | No |
| P07949 | CHEMBL2041 | RET | Drugged | Single protein | CHEMBL3545332 | Cep-2563 | Small molecule | Inhibitor | ChEMBL | Indication | All | - | - | No |
| P07949 | CHEMBL2041 | RET | Drugged | Single protein | CHEMBL1200485 | Sorafenib tosylate | Small molecule | Inhibitor | ChEMBL | Indication | All | 3 | Fibromatosis, aggressive | No |
| P07949 | CHEMBL2041 | RET | Drugged | Single protein | CHEMBL1567 | Sunitinib malate | Small molecule | Inhibitor | ChEMBL | Indication | All | 1 | Von hippel-lindau disease | No |
| P07949 | CHEMBL2041 | RET | Drugged | Single protein | CHEMBL1567 | Sunitinib malate | Small molecule | Inhibitor | ChEMBL | Indication | All | 1 | Cancers | No |
| P07949 | CHEMBL2041 | RET | Drugged | Single protein | CHEMBL1567 | Sunitinib malate | Small molecule | Inhibitor | ChEMBL | Indication | All | 3 | Cancers | No |
| P07949 | CHEMBL2041 | RET | Drugged | Single protein | CHEMBL1567 | Sunitinib malate | Small molecule | Inhibitor | ChEMBL | Indication | All | 4 | Cancers | No |
| P07949 | CHEMBL2041 | RET | Drugged | Single protein | CHEMBL24828 | Vandetanib | Small molecule | Inhibitor | ChEMBL | Indication | All | 1 | Cancers | No |
| P07949 | CHEMBL2041 | RET | Drugged | Single protein | CHEMBL24828 | Vandetanib | Small molecule | Inhibitor | ChEMBL | Indication | All | 2 | Cancers | No |
| P07949 | CHEMBL2041 | RET | Drugged | Single protein | CHEMBL24828 | Vandetanib | Small molecule | Inhibitor | ChEMBL | Indication | All | 3 | Cancers | No |
| P07949 | CHEMBL2041 | RET | Drugged | Single protein | CHEMBL24828 | Vandetanib | Small molecule | Inhibitor | ChEMBL | Indication | All | 4 | Cancers | No |
| P07949 | CHEMBL2041 | RET | Drugged | Single protein | CHEMBL3707320 | Alectinib hydrochloride | Small molecule | Inhibitor | ChEMBL | Indication | All | 4 | Cancers | No |
| P07949 | CHEMBL2041 | RET | Drugged | Single protein | CHEMBL574738 | Ast-487 | Small molecule | Inhibitor | ChEMBL | Indication | All | - | - | No |
| P07949 | CHEMBL2041 | RET | Drugged | Single protein | CHEMBL1200485 | Sorafenib tosylate | Small molecule | Inhibitor | ChEMBL | Indication | All | 4 | Cancers | No |
| P07949 | CHEMBL2041 | RET | Drugged | Single protein | CHEMBL1200485 | Sorafenib tosylate | Small molecule | Inhibitor | ChEMBL | Indication | All | 3 | Cancers | No |
| P07949 | CHEMBL2041 | RET | Drugged | Single protein | CHEMBL1946170 | Regorafenib | Small molecule | Inhibitor | BNF | Side-effect | Common or very common | 4 | Diarrhoea | No |
| P07949 | CHEMBL2041 | RET | Drugged | Single protein | CHEMBL1946170 | Regorafenib | Small molecule | Inhibitor | BNF | Side-effect | Common or very common | 4 | Dysphonia | No |
| P07949 | CHEMBL2041 | RET | Drugged | Single protein | CHEMBL1946170 | Regorafenib | Small molecule | Inhibitor | BNF | Side-effect | Common or very common | 4 | Dry mouth | No |
| P07949 | CHEMBL2041 | RET | Drugged | Single protein | CHEMBL1946170 | Regorafenib | Small molecule | Inhibitor | BNF | Side-effect | Rare or very rare | 4 | Cancers | No |
| P07949 | CHEMBL2041 | RET | Drugged | Single protein | CHEMBL1946170 | Regorafenib | Small molecule | Inhibitor | BNF | Side-effect | Rare or very rare | 4 | Posterior reversible encephalopathy syndrome pres | No |
| P07949 | CHEMBL2041 | RET | Drugged | Single protein | CHEMBL1946170 | Regorafenib | Small molecule | Inhibitor | BNF | Side-effect | Rare or very rare | 4 | Severe cutaneous adverse reactions scars | No |
| P07949 | CHEMBL2041 | RET | Drugged | Single protein | CHEMBL1946170 | Regorafenib | Small molecule | Inhibitor | BNF | Side-effect | Uncommon | 4 | - | No |
| P07949 | CHEMBL2041 | RET | Drugged | Single protein | CHEMBL1946170 | Regorafenib | Small molecule | Inhibitor | BNF | Side-effect | Uncommon | 4 | Gastrointestinal fistula | No |
| P07949 | CHEMBL2041 | RET | Drugged | Single protein | CHEMBL1946170 | Regorafenib | Small molecule | Inhibitor | BNF | Side-effect | Uncommon | 4 | Gastrointestinal perforation | No |
| P07949 | CHEMBL2041 | RET | Drugged | Single protein | CHEMBL1946170 | Regorafenib | Small molecule | Inhibitor | BNF | Side-effect | Uncommon | 4 | Hepatic disorder | No |
| P07949 | CHEMBL2041 | RET | Drugged | Single protein | CHEMBL1946170 | Regorafenib | Small molecule | Inhibitor | BNF | Side-effect | Not known | 4 | Aneurysm | No |
| P07949 | CHEMBL2041 | RET | Drugged | Single protein | CHEMBL1946170 | Regorafenib | Small molecule | Inhibitor | BNF | Side-effect | Uncommon | 4 | Myocardial infarction | Yes |
| P07949 | CHEMBL2041 | RET | Drugged | Single protein | CHEMBL1946170 | Regorafenib | Small molecule | Inhibitor | BNF | Side-effect | Uncommon | 4 | Nail disorder | No |
| P07949 | CHEMBL3430888 | RET | Drugged | Chimeric protein | CHEMBL4559134 | Selpercatinib | Small molecule | Inhibitor | ChEMBL | Indication | All | 1 | Liver diseases | No |
| P07949 | CHEMBL3430888 | RET | Drugged | Chimeric protein | CHEMBL4559134 | Selpercatinib | Small molecule | Inhibitor | ChEMBL | Indication | All | 1 | Renal insufficiency | No |
| P07949 | CHEMBL3430888 | RET | Drugged | Chimeric protein | CHEMBL4559134 | Selpercatinib | Small molecule | Inhibitor | ChEMBL | Indication | All | 4 | Cancers | No |
| P07949 | CHEMBL3430904 | RET | Drugged | Chimeric protein | CHEMBL4582651 | Pralsetinib | Small molecule | Inhibitor | ChEMBL | Indication | All | 4 | Cancers | No |
| P07949 | CHEMBL3430904 | RET | Drugged | Chimeric protein | CHEMBL4559134 | Selpercatinib | Small molecule | Inhibitor | ChEMBL | Indication | All | 4 | Cancers | No |

**Table S8. Druggability results of the prioritised proteins**

| Target Uniprot ID* | Target ChEMBL ID | Target protein name | Druggability* | Target type | Drug ChEMBL ID | Drug name | Drug molecule type | Drug mechanism | Drug effect source | Drug effect type | Drug effect frequency | Max phase* | Drug effect | Drug effect class |
|---|---|---|---|---|---|---|---|---|---|---|---|---|---|---|
| P07949 | CHEMBL3430904 | RET | Drugged | Chimeric protein | CHEMBL4559134 | Selpercatinib | Small molecule | Inhibitor | ChEMBL | Indication | All | 1 | Renal insufficiency | No |
| P07949 | CHEMBL3430904 | RET | Drugged | Chimeric protein | CHEMBL4559134 | Selpercatinib | Small molecule | Inhibitor | ChEMBL | Indication | All | 1 | Liver diseases | No |
| P07949 | CHEMBL2041 | RET | Drugged | Single protein | CHEMBL1946170 | Regorafenib | Small molecule | Inhibitor | BNF | Side-effect | Uncommon | 4 | Myocardial ischemia | Yes |
| P07949 | CHEMBL2041 | RET | Drugged | Single protein | CHEMBL1946170 | Regorafenib | Small molecule | Inhibitor | BNF | Side-effect | Common or very common | 4 | Weight decreased | No |
| P07949 | CHEMBL2041 | RET | Drugged | Single protein | CHEMBL1946170 | Regorafenib | Small molecule | Inhibitor | BNF | Side-effect | Not known | 4 | Artery dissection | No |
| P07949 | CHEMBL2041 | RET | Drugged | Single protein | CHEMBL1946170 | Regorafenib | Small molecule | Inhibitor | BNF | Side-effect | Common or very common | 4 | Tremor | No |
| P07949 | CHEMBL2041 | RET | Drugged | Single protein | CHEMBL1946170 | Regorafenib | Small molecule | Inhibitor | BNF | Side-effect | Common or very common | 4 | Electrolyte imbalance | No |
| P07949 | CHEMBL2041 | RET | Drugged | Single protein | CHEMBL1946170 | Regorafenib | Small molecule | Inhibitor | BNF | Side-effect | Common or very common | 4 | Fever | No |
| P07949 | CHEMBL2041 | RET | Drugged | Single protein | CHEMBL1946170 | Regorafenib | Small molecule | Inhibitor | BNF | Side-effect | Common or very common | 4 | Gastrooesophageal reflux disease | No |
| P07949 | CHEMBL2041 | RET | Drugged | Single protein | CHEMBL1946170 | Regorafenib | Small molecule | Inhibitor | BNF | Side-effect | Common or very common | 4 | Headache | No |
| P07949 | CHEMBL2041 | RET | Drugged | Single protein | CHEMBL1946170 | Regorafenib | Small molecule | Inhibitor | BNF | Side-effect | Common or very common | 4 | Hyperbilirubinemia | No |
| P07949 | CHEMBL2041 | RET | Drugged | Single protein | CHEMBL1946170 | Regorafenib | Small molecule | Inhibitor | BNF | Side-effect | Common or very common | 4 | Hypertension | Yes |
| P07949 | CHEMBL2041 | RET | Drugged | Single protein | CHEMBL1946170 | Regorafenib | Small molecule | Inhibitor | BNF | Side-effect | Common or very common | 4 | Hyperuricemia | No |
| P07949 | CHEMBL2041 | RET | Drugged | Single protein | CHEMBL1946170 | Regorafenib | Small molecule | Inhibitor | BNF | Side-effect | Common or very common | 4 | Hypothyroidism | No |
| P07949 | CHEMBL2041 | RET | Drugged | Single protein | CHEMBL1946170 | Regorafenib | Small molecule | Inhibitor | BNF | Side-effect | Common or very common | 4 | Increased risk of infection | No |
| P07949 | CHEMBL2041 | RET | Drugged | Single protein | CHEMBL1946170 | Regorafenib | Small molecule | Inhibitor | BNF | Side-effect | Common or very common | 4 | Vomiting | No |
| P07949 | CHEMBL2041 | RET | Drugged | Single protein | CHEMBL1946170 | Regorafenib | Small molecule | Inhibitor | BNF | Side-effect | Common or very common | 4 | Mucositis | No |
| P07949 | CHEMBL2041 | RET | Drugged | Single protein | CHEMBL1946170 | Regorafenib | Small molecule | Inhibitor | BNF | Side-effect | Common or very common | 4 | Musculoskeletal stiffness | No |
| P07949 | CHEMBL2041 | RET | Drugged | Single protein | CHEMBL1946170 | Regorafenib | Small molecule | Inhibitor | BNF | Side-effect | Common or very common | 4 | Leucopenia | No |
| P07949 | CHEMBL3430888 | RET | Drugged | Chimeric protein | CHEMBL4582651 | Pralsetinib | Small molecule | Inhibitor | ChEMBL | Indication | All | 4 | Cancers | No |
| P07949 | CHEMBL2041 | RET | Drugged | Single protein | CHEMBL1946170 | Regorafenib | Small molecule | Inhibitor | BNF | Side-effect | Common or very common | 4 | Nausea | No |
| P07949 | CHEMBL2041 | RET | Drugged | Single protein | CHEMBL1946170 | Regorafenib | Small molecule | Inhibitor | BNF | Side-effect | Common or very common | 4 | Thrombocytopenia | No |
| P07949 | CHEMBL2041 | RET | Drugged | Single protein | CHEMBL1946170 | Regorafenib | Small molecule | Inhibitor | BNF | Side-effect | Common or very common | 4 | Pain | No |
| P07949 | CHEMBL2041 | RET | Drugged | Single protein | CHEMBL1946170 | Regorafenib | Small molecule | Inhibitor | BNF | Side-effect | Common or very common | 4 | Proteinuria | No |
| P07949 | CHEMBL2041 | RET | Drugged | Single protein | CHEMBL1946170 | Regorafenib | Small molecule | Inhibitor | BNF | Side-effect | Common or very common | 4 | Taste altered | No |
| P07949 | CHEMBL2041 | RET | Drugged | Single protein | CHEMBL1946170 | Regorafenib | Small molecule | Inhibitor | BNF | Side-effect | Common or very common | 4 | Skin reactions | No |
| P07949 | CHEMBL2041 | RET | Drugged | Single protein | CHEMBL1946170 | Regorafenib | Small molecule | Inhibitor | BNF | Side-effect | Common or very common | 4 | Stomatitis | No |
| Q9UGC7 | - | RF1ML | Not yet druggable | - | - | - | - | - | - | - | - | - | - | No |
| Q96DX8 | - | RTP4 | Not yet druggable | - | - | - | - | - | - | - | - | - | - | No |
| Q96F10 | CHEMBL3509592 | SAT2 | Drugged | Single protein | CHEMBL609 | Trientine | Small molecule | Chelating agent | ChEMBL | Indication | All | 1 | Cancers | No |
| Q96F10 | CHEMBL3509592 | SAT2 | Drugged | Single protein | CHEMBL609 | Trientine | Small molecule | Chelating agent | ChEMBL | Indication | All | 2 | Macular edema | No |
| Q96F10 | CHEMBL3509592 | SAT2 | Drugged | Single protein | CHEMBL609 | Trientine | Small molecule | Chelating agent | ChEMBL | Indication | All | 4 | Liver cirrhosis, biliary | No |
| Q96F10 | CHEMBL3509592 | SAT2 | Drugged | Single protein | CHEMBL609 | Trientine | Small molecule | Chelating agent | ChEMBL | Indication | All | 4 | Arthritis | No |
| Q96F10 | CHEMBL3509592 | SAT2 | Drugged | Single protein | CHEMBL609 | Trientine | Small molecule | Chelating agent | ChEMBL | Indication | All | 4 | Hepatolenticular degeneration | No |
| Q96F10 | CHEMBL3509592 | SAT2 | Drugged | Single protein | CHEMBL609 | Trientine | Small molecule | Chelating agent | ChEMBL | Indication | All | 2 | Cardiomyopathy, hypertrophic | Yes |
| Q9UH65 | - | SWP70 | Not yet druggable | - | - | - | - | - | - | - | - | - | - | No |
| P13385 | CHEMBL3713025 | TDGF1 | Druggable | Single protein | CHEMBL2109575 | Biib-015 | Antibody | Binding agent | ChEMBL | Indication | All | 1 | Cancers | No |
| P42680 | CHEMBL4296642 | TEC | Drugged | Protein family | CHEMBL4085457 | Pf-06651600 | Small molecule | Inhibitor | ChEMBL | Indication | All | 3 | Alopecia | No |
| P42680 | CHEMBL4296642 | TEC | Drugged | Protein family | CHEMBL4085457 | Pf-06651600 | Small molecule | Inhibitor | ChEMBL | Indication | All | 2 | Vitiligo | No |
| P42680 | CHEMBL4296642 | TEC | Drugged | Protein family | CHEMBL4085457 | Pf-06651600 | Small molecule | Inhibitor | ChEMBL | Indication | All | 2 | Crohn disease | No |
| P42680 | CHEMBL4296642 | TEC | Drugged | Protein family | CHEMBL4085457 | Pf-06651600 | Small molecule | Inhibitor | ChEMBL | Indication | All | 2 | Colitis, ulcerative | No |
| P42680 | CHEMBL4296642 | TEC | Drugged | Protein family | CHEMBL4085457 | Pf-06651600 | Small molecule | Inhibitor | ChEMBL | Indication | All | 2 | Arthritis | No |
| P42680 | CHEMBL4296642 | TEC | Drugged | Protein family | CHEMBL4085457 | Pf-06651600 | Small molecule | Inhibitor | ChEMBL | Indication | All | 1 | Liver diseases | No |
| P42680 | CHEMBL4296642 | TEC | Drugged | Protein family | CHEMBL4085457 | Pf-06651600 | Small molecule | Inhibitor | ChEMBL | Indication | All | 1 | Kidney diseases | No |
| P22105 | - | TENX | Not yet druggable | - | - | - | - | - | - | - | - | - | - | No |
| Q92563 | - | TICN2 | Not yet druggable | - | - | - | - | - | - | - | - | - | - | No |
| P49788 | - | TIG1 | Not yet druggable | - | - | - | - | - | - | - | - | - | - | No |
| P35625 | - | TIMP3 | Not yet druggable | - | - | - | - | - | - | - | - | - | - | No |
| O15533 | - | TPSN | Not yet druggable | - | - | - | - | - | - | - | - | - | - | No |
| Q15661 | CHEMBL2617 | TRYB1 | Drugged | Single protein | CHEMBL1485 | Arginine | Small molecule | - | ChEMBL | Indication | All | 2 | Alzheimer disease | No |
| Q15661 | CHEMBL2617 | TRYB1 | Drugged | Single protein | CHEMBL1485 | Arginine | Small molecule | - | ChEMBL | Indication | All | 1 | Liver cirrhosis | No |
| Q15661 | CHEMBL2617 | TRYB1 | Drugged | Single protein | CHEMBL1485 | Arginine | Small molecule | - | ChEMBL | Indication | All | 3 | Periodontitis | No |
| Q15661 | CHEMBL2617 | TRYB1 | Drugged | Single protein | CHEMBL1485 | Arginine | Small molecule | - | ChEMBL | Indication | All | 2 | Arthritis | No |
| Q15661 | CHEMBL2617 | TRYB1 | Drugged | Single protein | CHEMBL1485 | Arginine | Small molecule | - | ChEMBL | Indication | All | 0,5 | Kidney diseases | No |
| Q15661 | CHEMBL2617 | TRYB1 | Drugged | Single protein | CHEMBL1485 | Arginine | Small molecule | - | ChEMBL | Indication | All | 2 | Beta-thalassemia | No |
| Q15661 | CHEMBL2617 | TRYB1 | Drugged | Single protein | CHEMBL1485 | Arginine | Small molecule | - | ChEMBL | Indication | All | 3 | Anemia | No |
| Q15661 | CHEMBL2617 | TRYB1 | Drugged | Single protein | CHEMBL1485 | Arginine | Small molecule | - | ChEMBL | Indication | All | 3 | Cardiovascular diseases | Yes |
| Q15661 | CHEMBL2617 | TRYB1 | Drugged | Single protein | CHEMBL1485 | Arginine | Small molecule | - | ChEMBL | Indication | All | 2 | Peripheral arterial disease | No |
| Q15661 | CHEMBL2617 | TRYB1 | Drugged | Single protein | CHEMBL1485 | Arginine | Small molecule | - | ChEMBL | Indication | All | 3 | Severe acute respiratory syndrome | No |
| Q15661 | CHEMBL2617 | TRYB1 | Drugged | Single protein | CHEMBL1485 | Arginine | Small molecule | - | ChEMBL | Indication | All | 2 | Pain | No |
| Q15661 | CHEMBL2617 | TRYB1 | Drugged | Single protein | CHEMBL1485 | Arginine | Small molecule | - | ChEMBL | Indication | All | 3 | Glucose intolerance | No |
| Q15661 | CHEMBL2617 | TRYB1 | Drugged | Single protein | CHEMBL1485 | Arginine | Small molecule | - | ChEMBL | Indication | All | 2 | Schizophrenia | No |
| Q15661 | CHEMBL2617 | TRYB1 | Drugged | Single protein | CHEMBL1485 | Arginine | Small molecule | - | ChEMBL | Indication | All | 2 | Asthma | No |
| Q15661 | CHEMBL2617 | TRYB1 | Drugged | Single protein | CHEMBL1485 | Arginine | Small molecule | - | ChEMBL | Indication | All | 1 | Hypertension | Yes |
| Q15661 | CHEMBL2617 | TRYB1 | Drugged | Single protein | CHEMBL1485 | Arginine | Small molecule | - | ChEMBL | Indication | All | 3 | Tuberculosis, pulmonary | No |
| Q15661 | CHEMBL2617 | TRYB1 | Drugged | Single protein | CHEMBL1485 | Arginine | Small molecule | - | ChEMBL | Indication | All | 2 | Argininosuccinic aciduria | No |
| Q15661 | CHEMBL2617 | TRYB1 | Drugged | Single protein | CHEMBL1485 | Arginine | Small molecule | - | ChEMBL | Indication | All | 1 | Diabetes mellitus, type 2 | No |
| Q15661 | CHEMBL2617 | TRYB1 | Drugged | Single protein | CHEMBL1485 | Arginine | Small molecule | - | ChEMBL | Indication | All | 2 | Hypertension | Yes |
| Q15661 | CHEMBL2617 | TRYB1 | Drugged | Single protein | CHEMBL1485 | Arginine | Small molecule | - | ChEMBL | Indication | All | 1 | Heart failure | Yes |
| Q15661 | CHEMBL2617 | TRYB1 | Drugged | Single protein | CHEMBL1485 | Arginine | Small molecule | - | ChEMBL | Indication | All | 2 | Melas syndrome | No |
| Q15661 | CHEMBL2617 | TRYB1 | Drugged | Single protein | CHEMBL1485 | Arginine | Small molecule | - | ChEMBL | Indication | All | 2 | Sepsis | No |
| Q15661 | CHEMBL2617 | TRYB1 | Drugged | Single protein | CHEMBL1485 | Arginine | Small molecule | - | ChEMBL | Indication | All | 3 | Cancers | No |
| Q15661 | CHEMBL2617 | TRYB1 | Drugged | Single protein | CHEMBL1485 | Arginine | Small molecule | - | ChEMBL | Indication | All | 2 | Death, sudden, cardiac | No |
| Q15661 | CHEMBL2617 | TRYB1 | Drugged | Single protein | CHEMBL1485 | Arginine | Small molecule | - | ChEMBL | Indication | All | 2 | Malaria | No |
| Q15661 | CHEMBL2617 | TRYB1 | Drugged | Single protein | CHEMBL1485 | Arginine | Small molecule | - | ChEMBL | Indication | All | 3 | Pre-eclampsia | No |
| Q15661 | CHEMBL2617 | TRYB1 | Drugged | Single protein | CHEMBL1485 | Arginine | Small molecule | - | ChEMBL | Indication | All | 2 | Hepatitis, alcoholic | No |
| Q15661 | CHEMBL2617 | TRYB1 | Drugged | Single protein | CHEMBL1485 | Arginine | Small molecule | - | ChEMBL | Indication | All | 2 | Dental caries | No |
| Q15661 | CHEMBL2617 | TRYB1 | Drugged | Single protein | CHEMBL1485 | Arginine | Small molecule | - | ChEMBL | Indication | All | 3 | Hypersensitivity | No |
| Q15661 | CHEMBL2617 | TRYB1 | Drugged | Single protein | CHEMBL1485 | Arginine | Small molecule | - | ChEMBL | Indication | All | 2 | Cancers | No |
| Q15661 | CHEMBL2617 | TRYB1 | Drugged | Single protein | CHEMBL1485 | Arginine | Small molecule | - | ChEMBL | Indication | All | 2 | Hernias, diaphragmatic, congenital | No |
| Q15661 | CHEMBL2617 | TRYB1 | Drugged | Single protein | CHEMBL1485 | Arginine | Small molecule | - | ChEMBL | Indication | All | 1 | Muscular dystrophy, duchenne | No |
| Q15661 | CHEMBL2617 | TRYB1 | Drugged | Single protein | CHEMBL1485 | Arginine | Small molecule | - | ChEMBL | Indication | All | 2 | Galactosemias | No |
| Q15661 | CHEMBL2617 | TRYB1 | Drugged | Single protein | CHEMBL1485 | Arginine | Small molecule | - | ChEMBL | Indication | All | 2 | Cystic fibrosis | No |
| Q15661 | CHEMBL2617 | TRYB1 | Drugged | Single protein | CHEMBL1485 | Arginine | Small molecule | - | ChEMBL | Indication | All | 1 | Cancers | No |
| Q15661 | CHEMBL2617 | TRYB1 | Drugged | Single protein | CHEMBL1485 | Arginine | Small molecule | - | ChEMBL | Indication | All | 1 | Coronary disease | Yes |
| P49746 | - | TSP3 | Not yet druggable | - | - | - | - | - | - | - | - | - | - | No |
| P25311 | - | ZA2G | Not yet druggable | - | - | - | - | - | - | - | - | - | - | No |

**Table S8. Druggability results of the prioritised proteins**

| Target Uniprot ID* | Target ChEMBL ID | Target protein name | Druggability* | Target type | Drug ChEMBL ID | Drug name | Drug molecule type | Drug mechanism | Drug effect source | Drug effect type | Drug effect frequency | Max phase* | Drug effect | Drug effect class |
|---|---|---|---|---|---|---|---|---|---|---|---|---|---|---|
| P00326 | CHEMBL3285 | ADH1B | Druggable | Single protein | - | - | - | - | - | - | - | - | - | No |
| P00326 | CHEMBL2096668 | ADH1B | Druggable | Protein family | CHEMBL1909285 | Nitrefazole | Small molecule | Inhibitor | ChEMBL | Indication | All | - | - | No |
| P00326 | CHEMBL2363044 | ADH1B | Druggable | Protein complex group | CHEMBL1308 | Fomepizole | Small molecule | Inhibitor | ChEMBL | Indication | All | 1 | Macular degeneration | No |

* Columns: Target Uniprot ID - Uniprot ID of the protein (target), Druggability - druggability per protein, where druggable is defined as a protein targeted by a compound that is being tested, and drugged is defined as a protein targeted by an approved compound (see Methods section),
Max phase - The maximum phase the drug has reached for the intended indication (0 = pre-clinical phases, 4 = in use in the clinic), see for more information: https://cfinan.gitlab.io/bio-misc/scripts/drug_lookups.html
Abbreviations: BNF = British National Formulary
