## Supplemental Table S9 for "Integrating metabolomics and proteomics to identify novel drug targets for heart failure and atrial fibrillation"

**Table S9. Frequency of indication and side-effects**

| Phenotype | Indications | Side-effects | Total |
|---|---|---|---|
| Cancers | 108 | 1 | 109 |
| Arthritis | 14 | 0 | 14 |
| Renal insufficiency | 12 | 0 | 12 |
| Severe acute respiratory syndrome | 11 | 0 | 11 |
| Hemorrhage | 8 | 3 | 11 |
| Nausea | 1 | 10 | 11 |
| Anemia | 7 | 4 | 11 |
| Headache | 0 | 10 | 10 |
| Atrial fibrillation | 10 | 0 | 10 |
| Diarrhoea | 0 | 9 | 9 |
| Skin reactions | 0 | 9 | 9 |
| Thrombocytopenia | 3 | 6 | 9 |
| Liver diseases | 9 | 0 | 9 |
| Alzheimer disease | 8 | 0 | 8 |
| Thrombosis | 8 | 0 | 8 |
| Heart failure | 8 | 0 | 8 |
| Myasthenia gravis | 7 | 1 | 8 |
| Dizziness | 0 | 8 | 8 |
| Vomiting | 0 | 8 | 8 |
| Schizophrenia | 7 | 0 | 7 |
| Dementia | 7 | 0 | 7 |
| Venous thromboembolism | 7 | 0 | 7 |
| Venous thrombosis | 6 | 0 | 6 |
| Infections | 6 | 0 | 6 |
| Hiv infection | 6 | 0 | 6 |
| Hypertension | 3 | 3 | 6 |
| Cardiovascular diseases | 6 | 0 | 6 |
| Hypotension | 1 | 5 | 6 |
| Parkinson disease | 5 | 0 | 5 |
| Hepatitis c | 5 | 0 | 5 |
| Dyspepsia | 4 | 1 | 5 |
| Increased risk of infection | 0 | 5 | 5 |
| Diabetes mellitus, type 2 | 5 | 0 | 5 |
| Asthenia | 0 | 5 | 5 |
| Pain | 3 | 2 | 5 |
| Pneumonia | 5 | 0 | 5 |
| Gastroesophageal reflux | 5 | 0 | 5 |
| Kidney diseases | 5 | 0 | 5 |
| Depressive disorder | 5 | 0 | 5 |
| Fever | 1 | 4 | 5 |
| Hepatic disorder | 0 | 5 | 5 |
| Stroke | 5 | 0 | 5 |
| Acute coronary syndrome | 5 | 0 | 5 |
| Hypersensitivity | 1 | 4 | 5 |
| Pulmonary embolism | 5 | 0 | 5 |
| Cystic fibrosis | 4 | 0 | 4 |
| Hallucination | 0 | 4 | 4 |
| Immune system diseases | 4 | 0 | 4 |
| Neuromyelitis optica | 4 | 0 | 4 |
| Appetite decreased | 0 | 4 | 4 |
| Psoriasis | 4 | 0 | 4 |
| Peripheral arterial disease | 4 | 0 | 4 |
| Sepsis | 3 | 1 | 4 |
| Gastrointestinal disorder | 0 | 4 | 4 |
| Esophagitis, peptic | 4 | 0 | 4 |
| Coronary disease | 4 | 0 | 4 |
| Thromboembolism | 4 | 0 | 4 |
| Gastrointestinal hemorrhage | 2 | 2 | 4 |
| Liver cirrhosis | 4 | 0 | 4 |
| Uveitis | 4 | 0 | 4 |
| Peptic ulcer | 4 | 0 | 4 |

| Table S9. Frequency of indication and side-effects | | | |
|---|---|---|---|
| Phenotype | Indications | Side-effects | Total |
| Duodenal ulcer | 4 | 0 | 4 |
| Diabetes mellitus, type 1 | 4 | 0 | 4 |
| Helicobacter infection | 4 | 0 | 4 |
| Tachycardia | 1 | 3 | 4 |
| Epilepsy | 3 | 0 | 3 |
| Influenza, human | 3 | 0 | 3 |
| Angiodema | 1 | 2 | 3 |
| Giant cell arteritis | 3 | 0 | 3 |
| Polycystic ovary syndrome | 3 | 0 | 3 |
| Plasmacytoma | 3 | 0 | 3 |
| Spondylitis, ankylosing | 3 | 0 | 3 |
| Waldenstrom macroglobulinemia | 3 | 0 | 3 |
| Hodgkin disease | 3 | 0 | 3 |
| Zollinger-ellison syndrome | 3 | 0 | 3 |
| Esophagitis | 3 | 0 | 3 |
| Colitis, ulcerative | 3 | 0 | 3 |
| Gastrointestinal discomfort | 0 | 3 | 3 |
| Multiple sclerosis, relapsing-remitting | 3 | 0 | 3 |
| Cough | 1 | 2 | 3 |
| Embolism | 3 | 0 | 3 |
| Severe cutaneous adverse reactions scars | 0 | 3 | 3 |
| Leucopenia | 0 | 3 | 3 |
| Myocardial infarction | 2 | 1 | 3 |
| Dyspnoea | 0 | 3 | 3 |
| Graft vs host disease | 3 | 0 | 3 |
| Stomach ulcer | 3 | 0 | 3 |
| Mycosis fungoides | 3 | 0 | 3 |
| Sezary syndrome | 3 | 0 | 3 |
| Ischemic stroke | 3 | 0 | 3 |
| Blood coagulation disorder | 3 | 0 | 3 |
| Aortic valve stenosis | 3 | 0 | 3 |
| Drowsiness | 0 | 3 | 3 |
| Tremor | 0 | 3 | 3 |
| Malaise | 0 | 3 | 3 |
| Cognitive dysfunction | 3 | 0 | 3 |
| Syncope | 0 | 3 | 3 |
| Constipation | 1 | 2 | 3 |
| Urinary tract infection | 2 | 1 | 3 |
| Hypotension, orthostatic | 3 | 0 | 3 |
| Seizure | 0 | 3 | 3 |
| Hypersalivation | 0 | 3 | 3 |
| Confusion | 0 | 3 | 3 |
| Substance-related disorder | 3 | 0 | 3 |
| Anxiety | 2 | 1 | 3 |
| Communicable diseases | 2 | 0 | 2 |
| Laryngopharyngeal reflux | 2 | 0 | 2 |
| Down syndrome | 2 | 0 | 2 |
| Osteoporosis, postmenopausal | 2 | 0 | 2 |
| Amyotrophic lateral sclerosis | 2 | 0 | 2 |
| Muscle cramps | 0 | 2 | 2 |
| Chronic pain | 2 | 0 | 2 |
| Hyperventilation | 0 | 2 | 2 |
| Neutropenia | 0 | 2 | 2 |
| Cysticercosis | 2 | 0 | 2 |
| Heartburn | 2 | 0 | 2 |
| Depression | 0 | 2 | 2 |
| Barrett esophagus | 2 | 0 | 2 |
| Idiopathic pulmonary fibrosis | 2 | 0 | 2 |
| Wounds and injuries | 2 | 0 | 2 |
| Menopause | 2 | 0 | 2 |
| Renal impairment | 0 | 2 | 2 |

| Table S9. Frequency of indication and side-effects | | | |
|---|---|---|---|
| **Phenotype** | **Indications** | **Side-effects** | **Total** |
| Oral disorder | 0 | 2 | 2 |
| Irritable bowel syndrome | 2 | 0 | 2 |
| Hearing loss | 1 | 1 | 2 |
| Taste altered | 0 | 2 | 2 |
| Thrombocytosis | 0 | 2 | 2 |
| Lupus erythematosus, systemic | 2 | 0 | 2 |
| Vertigo | 0 | 2 | 2 |
| Mastocytosis | 2 | 0 | 2 |
| Polymyalgia rheumatica | 2 | 0 | 2 |
| Scleroderma | 2 | 0 | 2 |
| Mild to moderate dementia in alzheimer's disease | 2 | 0 | 2 |
| Osteoporosis | 2 | 0 | 2 |
| Child development disorder, pervasive | 2 | 0 | 2 |
| Pulmonary disease, chronic obstructive | 2 | 0 | 2 |
| Cocaine-related disorder | 2 | 0 | 2 |
| Wound complications | 0 | 2 | 2 |
| Treatment of pulmonary embolism | 2 | 0 | 2 |
| Treatment of deep-vein thrombosis | 2 | 0 | 2 |
| Prophylaxis of venous thromboembolism following knee replacement surgery | 2 | 0 | 2 |
| Prophylaxis of venous thromboembolism following hip replacement surgery | 2 | 0 | 2 |
| Prophylaxis of recurrent pulmonary embolism | 2 | 0 | 2 |
| Prophylaxis of recurrent deep-vein thrombosis | 2 | 0 | 2 |
| Dry mouth | 0 | 2 | 2 |
| Atrial flutter | 2 | 0 | 2 |
| Oedema | 0 | 2 | 2 |
| Crohn disease | 2 | 0 | 2 |
| Urinary incontinence | 0 | 2 | 2 |
| Ischemia | 2 | 0 | 2 |
| Pre-eclampsia | 2 | 0 | 2 |
| Pulmonary hypertension | 2 | 0 | 2 |
| Macular edema | 2 | 0 | 2 |
| Movement disorder | 0 | 2 | 2 |
| Psychotic disorder | 2 | 0 | 2 |
| Heart diseases | 2 | 0 | 2 |
| Delirium | 2 | 0 | 2 |
| Cardiac conduction disorder | 0 | 2 | 2 |
| Vision disorder | 0 | 2 | 2 |
| Attention deficit disorder with hyperactivity | 2 | 0 | 2 |
| Dyslipidemia | 0 | 2 | 2 |
| Hypothyroidism | 0 | 2 | 2 |
| Weight increased | 0 | 2 | 2 |
| Mitochondrial diseases | 2 | 0 | 2 |
| Brain injuries | 2 | 0 | 2 |
| Dental caries | 2 | 0 | 2 |
| Aggression | 0 | 2 | 2 |
| Muscle weakness | 1 | 1 | 2 |
| Muscular atrophy, spinal | 2 | 0 | 2 |
| Arrhythmias | 0 | 2 | 2 |
| Cytomegalovirus infection | 2 | 0 | 2 |
| Pyelonephritis | 2 | 0 | 2 |
| Non-alcoholic fatty liver disease | 2 | 0 | 2 |
| Weight decreased | 0 | 2 | 2 |
| Purpura, thrombocytopenic, idiopathic | 2 | 0 | 2 |
| Castleman disease | 2 | 0 | 2 |
| Amyloidosis, familial | 2 | 0 | 2 |
| Alopecia | 1 | 1 | 2 |
| Von hippel-lindau disease | 2 | 0 | 2 |
| Agitation | 0 | 2 | 2 |
| Rett syndrome | 2 | 0 | 2 |
| Sleep disorder | 0 | 2 | 2 |
| Hepatitis b, chronic | 2 | 0 | 2 |

**Table S9. Frequency of indication and side-effects**

| Phenotype | Indications | Side-effects | Total |
|---|---|---|---|
| Encephalitis, japanese | 2 | 0 | 2 |
| Immunoglobulin light-chain amyloidosis | 2 | 0 | 2 |
| Hyperhidrosis | 0 | 2 | 2 |
| Virus diseases | 2 | 0 | 2 |
| Macular degeneration | 2 | 0 | 2 |
| Spinal cord injuries | 2 | 0 | 2 |
| Multiple sclerosis | 2 | 0 | 2 |
| Eosinophilia | 0 | 2 | 2 |
| Amyloidosis | 2 | 0 | 2 |
| Fall | 0 | 2 | 2 |
| Diabetes mellitus | 2 | 0 | 2 |
| Ependymoma | 1 | 0 | 1 |
| Stiff-person syndrome | 1 | 0 | 1 |
| Opsoclonus-myoclonus syndrome | 1 | 0 | 1 |
| Polyradiculoneuropathy, chronic inflammatory demyelinating | 1 | 0 | 1 |
| Skin necrosis | 0 | 1 | 1 |
| Thrombotic microangiopathy | 0 | 1 | 1 |
| Acne vulgaris | 1 | 0 | 1 |
| Vitiligo | 1 | 0 | 1 |
| Hematologic diseases | 1 | 0 | 1 |
| Medulloblastoma | 1 | 0 | 1 |
| Beta-thalassemia | 1 | 0 | 1 |
| Cardiomyopathy, hypertrophic | 1 | 0 | 1 |
| Hepatolenticular degeneration | 1 | 0 | 1 |
| Liver cirrhosis, biliary | 1 | 0 | 1 |
| Stevens-johnson syndrome | 0 | 1 | 1 |
| Pancytopenia | 0 | 1 | 1 |
| Stomatitis | 0 | 1 | 1 |
| Interstitial lung disease | 0 | 1 | 1 |
| Infusion related reaction | 0 | 1 | 1 |
| Proteinuria | 0 | 1 | 1 |
| Periodontitis | 1 | 0 | 1 |
| Asthma | 1 | 0 | 1 |
| Antiphospholipid syndrome | 1 | 0 | 1 |
| Melas syndrome | 1 | 0 | 1 |
| Hemophilia a | 1 | 0 | 1 |
| Prophylaxis of hemorrhage in hemophilia a | 1 | 0 | 1 |
| Arthralgia | 0 | 1 | 1 |
| Neuroleptic malignant syndrome | 0 | 1 | 1 |
| Myalgia | 0 | 1 | 1 |
| Cavernous sinus thrombosis | 0 | 1 | 1 |
| Muscular dystrophy, duchenne | 1 | 0 | 1 |
| Hernias, diaphragmatic, congenital | 1 | 0 | 1 |
| Hepatitis, alcoholic | 1 | 0 | 1 |
| Embolism and thrombosis | 0 | 1 | 1 |
| Malaria | 1 | 0 | 1 |
| Death, sudden, cardiac | 1 | 0 | 1 |
| Menorrhagia | 0 | 1 | 1 |
| Vascular pseudoaneurysm | 0 | 1 | 1 |
| Rheumatic heart disease | 1 | 0 | 1 |
| Foramen ovale, patent | 1 | 0 | 1 |
| Argininosuccinic aciduria | 1 | 0 | 1 |
| Disseminated intravascular coagulation | 1 | 0 | 1 |
| Rhabdomyolysis | 0 | 1 | 1 |
| Bradycardia | 0 | 1 | 1 |
| Nephrotic syndrome | 1 | 0 | 1 |
| Tuberculosis, pulmonary | 1 | 0 | 1 |
| Factor x deficiency | 1 | 0 | 1 |
| Mucositis | 0 | 1 | 1 |
| Glucose intolerance | 1 | 0 | 1 |
| Diabetic foot | 1 | 0 | 1 |

| Table S9. Frequency of indication and side-effects | | | |
|---|---|---|---|
| **Phenotype** | **Indications** | **Side-effects** | **Total** |
| Aortic valve disease | 1 | 0 | 1 |
| Obesity, morbid | 1 | 0 | 1 |
| Musculoskeletal stiffness | 0 | 1 | 1 |
| Artery dissection | 0 | 1 | 1 |
| Hyperuricemia | 0 | 1 | 1 |
| Bruising | 0 | 1 | 1 |
| Dermatomyositis | 1 | 0 | 1 |
| Behcet syndrome | 1 | 0 | 1 |
| Friedreich ataxia | 1 | 0 | 1 |
| Pharyngitis | 1 | 0 | 1 |
| Non-st elevated myocardial infarction | 1 | 0 | 1 |
| Macrophage activation syndrome | 1 | 0 | 1 |
| Warts | 1 | 0 | 1 |
| Takayasu arteritis | 1 | 0 | 1 |
| Tooth diseases | 1 | 0 | 1 |
| Fibrous dysplasia of bone | 1 | 0 | 1 |
| Cystitis | 0 | 1 | 1 |
| Aggressive systemic mastocytosis | 1 | 0 | 1 |
| Chills | 0 | 1 | 1 |
| Leukemia | 1 | 0 | 1 |
| Primary myelofibrosis | 1 | 0 | 1 |
| Congestive heart failure | 0 | 1 | 1 |
| Qt interval prolongation | 0 | 1 | 1 |
| Cardiac disorder | 0 | 1 | 1 |
| Respiratory disorder | 0 | 1 | 1 |
| Oropharyngeal pain | 0 | 1 | 1 |
| Hyperglycemia | 0 | 1 | 1 |
| Febrile neutropenia | 0 | 1 | 1 |
| Concentration impaired | 0 | 1 | 1 |
| Blast crisis | 1 | 0 | 1 |
| Gingivitis | 1 | 0 | 1 |
| Tourette syndrome | 1 | 0 | 1 |
| Muscular diseases | 1 | 0 | 1 |
| Leigh disease | 1 | 0 | 1 |
| Intracranial hemorrhage | 0 | 1 | 1 |
| Metastatic colorectal cancer | 1 | 0 | 1 |
| Hyperbilirubinemia | 0 | 1 | 1 |
| Posterior reversible encephalopathy syndrome pres | 0 | 1 | 1 |
| Gastrooesophageal reflux disease | 0 | 1 | 1 |
| Peripheral oedema | 0 | 1 | 1 |
| Nephrolithiasis | 0 | 1 | 1 |
| Electrolyte imbalance | 0 | 1 | 1 |
| Keratosis, seborrheic | 1 | 0 | 1 |
| Myocardial ischemia | 0 | 1 | 1 |
| Nail disorder | 0 | 1 | 1 |
| Aneurysm | 0 | 1 | 1 |
| Endometrial hyperplasia | 1 | 0 | 1 |
| Gastrointestinal perforation | 0 | 1 | 1 |
| Gastrointestinal fistula | 0 | 1 | 1 |
| Bone diseases, metabolic | 1 | 0 | 1 |
| Conjunctivitis | 0 | 1 | 1 |
| Abdominal pain | 0 | 1 | 1 |
| Still's disease, adult-onset | 1 | 0 | 1 |
| Dysphonia | 0 | 1 | 1 |
| Fibromatosis, aggressive | 1 | 0 | 1 |
| Retroviridae infection | 1 | 0 | 1 |
| Thrombocythemia, essential | 1 | 0 | 1 |
| Familial mediterranean fever | 1 | 0 | 1 |
| Polychondritis, relapsing | 1 | 0 | 1 |
| Respiratory distress syndrome | 1 | 0 | 1 |
| Adjunct to atropine in the treatment of poisoning by organophosphorus insecticide or nerve agent | 1 | 0 | 1 |

| Table S9. Frequency of indication and side-effects | | | |
|---|---|---|---|
| Phenotype | Indications | Side-effects | Total |
| Schnitzler syndrome | 1 | 0 | 1 |
| Polycythemia vera | 1 | 0 | 1 |
| Erdheim-chester disease | 1 | 0 | 1 |
| Graves ophthalmopathy | 1 | 0 | 1 |
| Lymphohistiocytosis, hemophagocytic | 1 | 0 | 1 |
| Lung diseases | 1 | 0 | 1 |
| Anxiety disorder | 1 | 0 | 1 |
| Pain in extremity | 0 | 1 | 1 |
| Smoldering multiple myeloma | 1 | 0 | 1 |
| Hyperlipoproteinemia type i | 1 | 0 | 1 |
| Metabolic syndrome | 1 | 0 | 1 |
| Infertility | 1 | 0 | 1 |
| Common cold | 0 | 1 | 1 |
| Uremia | 1 | 0 | 1 |
| Myeloproliferative disorder | 1 | 0 | 1 |
| Fatigue | 0 | 1 | 1 |
| Focal tremor | 0 | 1 | 1 |
| Red-cell aplasia, pure | 1 | 0 | 1 |
| Injury | 0 | 1 | 1 |
| Immunoblastic lymphadenopathy | 1 | 0 | 1 |
| Bronchiolitis obliterans | 1 | 0 | 1 |
| Glycogen storage disease type ii | 1 | 0 | 1 |
| Spinal muscular atrophies of childhood | 1 | 0 | 1 |
| Obesity | 1 | 0 | 1 |
| Encephalopathy | 0 | 1 | 1 |
| Thrombophlebitis | 0 | 1 | 1 |
| Flushing | 0 | 1 | 1 |
| Agranulocytosis | 0 | 1 | 1 |
| Antibiotic associated colitis | 0 | 1 | 1 |
| Chest discomfort | 0 | 1 | 1 |
| Colitis hemorrhagic | 0 | 1 | 1 |
| Anaphylactic reaction | 0 | 1 | 1 |
| Cyanosis | 0 | 1 | 1 |
| Empirical treatment of infection in febrile patients with neutropenia | 1 | 0 | 1 |
| Infection caused by | 1 | 0 | 1 |
| Peritonitis | 1 | 0 | 1 |
| Intraabdominal infection | 1 | 0 | 1 |
| Osteomyelitis | 1 | 0 | 1 |
| Bacterial infection | 1 | 0 | 1 |
| Or other less sensitive organisms | 1 | 0 | 1 |
| Ileus | 1 | 0 | 1 |
| Frontotemporal dementia | 1 | 0 | 1 |
| Lymphoproliferative disorder | 1 | 0 | 1 |
| Emergency loading dose, for atrial fibrillation or flutter | 1 | 0 | 1 |
| Gait abnormal | 0 | 1 | 1 |
| Dehydration | 0 | 1 | 1 |
| Psychosis | 0 | 1 | 1 |
| Mild to moderate dementia in parkinson's disease | 1 | 0 | 1 |
| Inflammation | 1 | 0 | 1 |
| Parkinsonism | 0 | 1 | 1 |
| Hepatitis | 0 | 1 | 1 |
| Nightmare | 0 | 1 | 1 |
| Pancreatitis | 0 | 1 | 1 |
| Gynaecomastia | 0 | 1 | 1 |
| Angina | 0 | 1 | 1 |
| Keratosis, actinic | 1 | 0 | 1 |
| Atrioventricular block | 0 | 1 | 1 |
| Shock | 1 | 0 | 1 |
| Maintenance, for atrial fibrillation or flutter | 1 | 0 | 1 |
| Mental disorder | 1 | 0 | 1 |
| Rapid digitalisation, for atrial fibrillation or flutter | 1 | 0 | 1 |

| Table S9. Frequency of indication and side-effects | | | |
|---|---|---|---|
| **Phenotype** | **Indications** | **Side-effects** | **Total** |
| Cerebral impairment | 0 | 1 | 1 |
| Hyperthyroidism | 1 | 0 | 1 |
| Respiratory insufficiency | 1 | 0 | 1 |
| Sarcopenia | 1 | 0 | 1 |
| Brain diseases | 1 | 0 | 1 |
| Acquired immunodeficiency syndrome | 1 | 0 | 1 |
| Lipid metabolism disorder | 1 | 0 | 1 |
| Gastric ulcer | 0 | 1 | 1 |
| Kidney failure, chronic | 1 | 0 | 1 |
| Muscle cramp | 1 | 0 | 1 |
| Acute kidney injury | 1 | 0 | 1 |
| Poisoning | 1 | 0 | 1 |
| Shock, septic | 1 | 0 | 1 |
| Aerobic and anaerobic gram-positive and gram-negative infection | 1 | 0 | 1 |
| Hospital-acquired septicemia | 1 | 0 | 1 |
| Post procedural hematoma | 0 | 1 | 1 |
| Postoperative nausea and vomiting | 1 | 0 | 1 |
| Landau-kleffner syndrome | 1 | 0 | 1 |
| Ulcer | 1 | 0 | 1 |
| Central serous chorioretinopathy | 1 | 0 | 1 |
| Laryngomalacia | 1 | 0 | 1 |
| Snoring | 1 | 0 | 1 |
| Lupus nephritis | 1 | 0 | 1 |
| Sexual dysfunction, physiological | 1 | 0 | 1 |
| Life-threatening infection | 1 | 0 | 1 |
| Pseudomyxoma peritonei | 1 | 0 | 1 |
| Gout | 1 | 0 | 1 |
| Eosinophilic esophagitis | 1 | 0 | 1 |
| Autistic disorder | 1 | 0 | 1 |
| Migraine disorder | 1 | 0 | 1 |
| Neuralgia | 1 | 0 | 1 |
| Apnea | 1 | 0 | 1 |
| Status epilepticus | 1 | 0 | 1 |
| Hydrocephalus, normal pressure | 1 | 0 | 1 |
| Edema | 1 | 0 | 1 |
| Andersen syndrome | 1 | 0 | 1 |
| Glaucoma, angle-closure | 1 | 0 | 1 |
| Seizures | 1 | 0 | 1 |
| Sleep apnea, obstructive | 1 | 0 | 1 |
| Extrapyramidal symptoms | 0 | 1 | 1 |
| Dermatitis, atopic | 1 | 0 | 1 |
| Altitude sickness | 1 | 0 | 1 |
| Takotsubo cardiomyopathy | 1 | 0 | 1 |
| Prophylaxis of stroke and systemic embolism | 1 | 0 | 1 |
| Prophylaxis of atherothrombotic events following an acute coronary syndrome with elevated cardiac biomark | 1 | 0 | 1 |
| Prophylaxis of atherothrombotic events | 1 | 0 | 1 |
| Prophylaxis of stroke and systemic embolism in non-valvular atrial fibrillation and at least one risk factor | 1 | 0 | 1 |
| Cns hemorrhage | 0 | 1 | 1 |
| Neurobehavioral manifestations | 1 | 0 | 1 |
| Cardiomyopathies | 1 | 0 | 1 |
| Attention deficit and disruptive behavior disorder | 1 | 0 | 1 |
| Gastroparesis | 1 | 0 | 1 |
| Urine discolouration | 0 | 1 | 1 |
| Bone marrow disorder | 0 | 1 | 1 |
| Digestive system diseases | 1 | 0 | 1 |
| Psychiatric disorder | 0 | 1 | 1 |
| Aerobic gram-negative infection | 1 | 0 | 1 |
| Tooth discolouration | 0 | 1 | 1 |
| Tongue discolouration | 0 | 1 | 1 |
| Palpitations | 0 | 1 | 1 |
| Paraesthesia | 0 | 1 | 1 |

| Table S9. Frequency of indication and side-effects | | | |
|---|---|---|---|
| Phenotype | Indications | Side-effects | Total |
| Polyarthralgia | 0 | 1 | 1 |
| Polyuria | 0 | 1 | 1 |
| Spinal pain | 0 | 1 | 1 |
| Tinnitus | 0 | 1 | 1 |
| Endocarditis | 1 | 0 | 1 |
| Abdominal cramps | 0 | 1 | 1 |
| Gastritis | 1 | 0 | 1 |
| Gastrointestinal hypermotility | 0 | 1 | 1 |
| Excessive tearing | 0 | 1 | 1 |
| Lecithin cholesterol acyltransferase deficiency | 1 | 0 | 1 |
| Rash | 0 | 1 | 1 |
| Cerebrovascular disorder | 1 | 0 | 1 |
| Gastrinoma | 1 | 0 | 1 |
| Premature birth | 1 | 0 | 1 |
| Amphetamine-related disorder | 1 | 0 | 1 |
| Squamous intraepithelial lesions | 1 | 0 | 1 |
| Clostridium infection | 1 | 0 | 1 |
| Exocrine pancreatic insufficiency | 1 | 0 | 1 |
| Supranuclear palsy, progressive | 1 | 0 | 1 |
| Erectile dysfunction | 1 | 0 | 1 |
| Granuloma | 1 | 0 | 1 |
| Galactosemias | 1 | 0 | 1 |
