## Supplemental Table S10 for "Integrating metabolomics and proteomics to identify novel drug targets for heart failure and atrial fibrillation"

**Table S10. Reactome pathway enrichment**

| Pathway | Pathway identifier | No. proteins in subset* | % proteins in subset* | No. proteins in data* | % proteins in data* | Enrichment (95% CI) | Test statistic | p-value | Adjusted p-value | Proteins in subset | Enriched |
|---|---|---|---|---|---|---|---|---|---|---|---|
| RET signaling | R-HSA-8853659 | 3 | 0,037 | 3 | 0,003 | 0.034 (0.009; 0.099) | 4,199 | $2.68\times10^{-5}$ | 0,004 | GFRA1, KPCA, RET | Yes |
| Antigen Presentation: Folding, assembly and peptide loading of class I MH | R-HSA-983170 | 2 | 0,024 | 1 | 0,001 | 0.023 (0.005; 0.084) | 4,111 | $3.94\times10^{-5}$ | 0,004 | TPSN, ERAP1 | Yes |
| Synthesis, secretion, and deacylation of Ghrelin | R-HSA-422085 | 2 | 0,024 | 2 | 0,002 | 0.023 (0.004; 0.083) | 3,426 | $6.13\times10^{-4}$ | 0,046 | ACES, NEC1 | Yes |
| Anti-inflammatory response favouring Leishmania parasite infection | R-HSA-9662851 | 3 | 0,037 | 6 | 0,005 | 0.031 (0.006; 0.097) | 3,161 | 0,002 | 0,07 | FCG2A, FCG3A, DPEP1 | No |
| Leishmania parasite growth and survival | R-HSA-9664433 | 3 | 0,037 | 6 | 0,005 | 0.031 (0.006; 0.097) | 3,161 | 0,002 | 0,07 | FCG2A, FCG3A, DPEP1 | No |
| Insulin processing | R-HSA-264876 | 1 | 0,012 | 1 | 0,001 | 0.011 (0.0; 0.065) | 2,42 | 0,016 | 0,102 | NEC1 | No |
| LTC4-CYSLTR mediated IL4 production | R-HSA-9664535 | 1 | 0,012 | 1 | 0,001 | 0.011 (0.0; 0.065) | 2,42 | 0,016 | 0,102 | DPEP1 | No |
| MAPK1 (ERK2) activation | R-HSA-112411 | 1 | 0,012 | 1 | 0,001 | 0.011 (0.0; 0.065) | 2,42 | 0,016 | 0,102 | IL6RA | No |
| Miscellaneous transport and binding events | R-HSA-5223345 | 1 | 0,012 | 1 | 0,001 | 0.011 (0.0; 0.065) | 2,42 | 0,016 | 0,102 | ZA2G | No |
| N-glycan trimming in the ER and Calnexin/Calreticulin cycle | R-HSA-532668 | 1 | 0,012 | 1 | 0,001 | 0.011 (0.0; 0.065) | 2,42 | 0,016 | 0,102 | ENASE | No |
| NOTCH2 intracellular domain regulates transcription | R-HSA-2197563 | 1 | 0,012 | 1 | 0,001 | 0.011 (0.0; 0.065) | 2,42 | 0,016 | 0,102 | GRAB | No |
| Nectin/Necl trans heterodimerization | R-HSA-420597 | 1 | 0,012 | 1 | 0,001 | 0.011 (0.0; 0.065) | 2,42 | 0,016 | 0,102 | PVRL4 | No |
| Regulation of KIT signaling | R-HSA-1433559 | 1 | 0,012 | 1 | 0,001 | 0.011 (0.0; 0.065) | 2,42 | 0,016 | 0,102 | KPCA | No |
| Phosphate bond hydrolysis by NUDT proteins | R-HSA-2393930 | 1 | 0,012 | 1 | 0,001 | 0.011 (0.0; 0.065) | 2,42 | 0,016 | 0,102 | NUDT9 | No |
| G alpha (z) signalling events | R-HSA-418597 | 1 | 0,012 | 1 | 0,001 | 0.011 (0.0; 0.065) | 2,42 | 0,016 | 0,102 | KPCA | No |
| Regulation of insulin secretion | R-HSA-422356 | 1 | 0,012 | 1 | 0,001 | 0.011 (0.0; 0.065) | 2,42 | 0,016 | 0,102 | KPCA | No |
| Regulation of signaling by NODAL | R-HSA-1433617 | 1 | 0,012 | 1 | 0,001 | 0.011 (0.0; 0.065) | 2,42 | 0,016 | 0,102 | TDGF1 | No |
| Transcriptional regulation of granulopoiesis | R-HSA-9616222 | 1 | 0,012 | 1 | 0,001 | 0.011 (0.0; 0.065) | 2,42 | 0,016 | 0,102 | IL6RA | No |
| VLDL assembly | R-HSA-8866423 | 1 | 0,012 | 1 | 0,001 | 0.011 (0.0; 0.065) | 2,42 | 0,016 | 0,102 | APOC1 | No |
| VLDL clearance | R-HSA-8964046 | 1 | 0,012 | 1 | 0,001 | 0.011 (0.0; 0.065) | 2,42 | 0,016 | 0,102 | APOC1 | No |
| Opioid Signalling | R-HSA-111885 | 1 | 0,012 | 1 | 0,001 | 0.011 (0.0; 0.065) | 2,42 | 0,016 | 0,102 | KPCA | No |
| Disinhibition of SNARE formation | R-HSA-114516 | 1 | 0,012 | 1 | 0,001 | 0.011 (0.0; 0.065) | 2,42 | 0,016 | 0,102 | KPCA | No |
| Nuclear Envelope Breakdown | R-HSA-2980766 | 1 | 0,012 | 1 | 0,001 | 0.011 (0.0; 0.065) | 2,42 | 0,016 | 0,102 | KPCA | No |
| Beta-oxidation of pristanoyl-CoA | R-HSA-389887 | 1 | 0,012 | 1 | 0,001 | 0.011 (0.0; 0.065) | 2,42 | 0,016 | 0,102 | CACP | No |
| Depolymerization of the Nuclear Lamina | R-HSA-4419969 | 1 | 0,012 | 1 | 0,001 | 0.011 (0.0; 0.065) | 2,42 | 0,016 | 0,102 | KPCA | No |
| FCGR activation | R-HSA-2029481 | 2 | 0,024 | 3 | 0,003 | 0.022 (0.003; 0.082) | 2,944 | 0,003 | 0,102 | FCG2A, FCG3A | No |
| Role of phospholipids in phagocytosis | R-HSA-2029485 | 2 | 0,024 | 3 | 0,003 | 0.022 (0.003; 0.082) | 2,944 | 0,003 | 0,102 | FCG2A, FCG3A | No |
| Acyl chain remodelling of PC | R-HSA-1482788 | 1 | 0,012 | 1 | 0,001 | 0.011 (0.0; 0.065) | 2,42 | 0,016 | 0,102 | PLA2R | No |
| FCGR3A-mediated IL10 synthesis | R-HSA-9664323 | 2 | 0,024 | 4 | 0,004 | 0.021 (0.002; 0.081) | 2,578 | 0,01 | 0,102 | FCG2A, FCG3A | No |
| Acyl chain remodelling of PG | R-HSA-1482925 | 1 | 0,012 | 1 | 0,001 | 0.011 (0.0; 0.065) | 2,42 | 0,016 | 0,102 | PLA2R | No |
| Basigin interactions | R-HSA-210991 | 1 | 0,012 | 1 | 0,001 | 0.011 (0.0; 0.065) | 2,42 | 0,016 | 0,102 | AT1B2 | No |
| Acyl chain remodelling of PE | R-HSA-1482839 | 1 | 0,012 | 1 | 0,001 | 0.011 (0.0; 0.065) | 2,42 | 0,016 | 0,102 | PLA2R | No |
| Acyl chain remodelling of PS | R-HSA-1482801 | 1 | 0,012 | 1 | 0,001 | 0.011 (0.0; 0.065) | 2,42 | 0,016 | 0,102 | PLA2R | No |
| Acyl chain remodelling of PI | R-HSA-1482922 | 1 | 0,012 | 1 | 0,001 | 0.011 (0.0; 0.065) | 2,42 | 0,016 | 0,102 | PLA2R | No |
| Leishmania infection | R-HSA-9658195 | 4 | 0,049 | 16 | 0,014 | 0.034 (0.004; 0.105) | 2,352 | 0,019 | 0,116 | FCG2A, ENTP5, FCG3A, DPEP1 | No |
| Parasitic Infection Pathways | R-HSA-9824443 | 4 | 0,049 | 16 | 0,014 | 0.034 (0.004; 0.105) | 2,352 | 0,019 | 0,116 | FCG2A, ENTP5, FCG3A, DPEP1 | No |
| Regulation of actin dynamics for phagocytic cup formation | R-HSA-2029482 | 2 | 0,024 | 5 | 0,004 | 0.02 (0.001; 0.08) | 2,285 | 0,022 | 0,134 | FCG2A, FCG3A | No |
| Mitotic Prophase | R-HSA-68875 | 1 | 0,012 | 2 | 0,002 | 0.01 (-0.001; 0.064) | 1,821 | 0,069 | 0,213 | KPCA | No |
| EGFR Transactivation by Gastrin | R-HSA-2179392 | 1 | 0,012 | 2 | 0,002 | 0.01 (-0.001; 0.064) | 1,821 | 0,069 | 0,213 | KPCA | No |
| Digestion of dietary lipid | R-HSA-192456 | 1 | 0,012 | 2 | 0,002 | 0.01 (-0.001; 0.064) | 1,821 | 0,069 | 0,213 | CEL | No |
| Deregulated CDK5 triggers multiple neurodegenerative pathways in Alzhei | R-HSA-8862803 | 1 | 0,012 | 2 | 0,002 | 0.01 (-0.001; 0.064) | 1,821 | 0,069 | 0,213 | CAN2 | No |
| Defective Intrinsic Pathway for Apoptosis | R-HSA-9734009 | 1 | 0,012 | 2 | 0,002 | 0.01 (-0.001; 0.064) | 1,821 | 0,069 | 0,213 | CAN2 | No |
| HSP90 chaperone cycle for steroid hormone receptors (SHR) in the presen | R-HSA-3371497 | 1 | 0,012 | 2 | 0,002 | 0.01 (-0.001; 0.064) | 1,821 | 0,069 | 0,213 | DNJA4 | No |
| Inactivation, recovery and regulation of the phototransduction cascade | R-HSA-2514859 | 1 | 0,012 | 2 | 0,002 | 0.01 (-0.001; 0.064) | 1,821 | 0,069 | 0,213 | KPCA | No |
| Interleukin-18 signaling | R-HSA-9012546 | 1 | 0,012 | 2 | 0,002 | 0.01 (-0.001; 0.064) | 1,821 | 0,069 | 0,213 | IL18R | No |
| CRMPs in Sema3A signaling | R-HSA-399956 | 1 | 0,012 | 2 | 0,002 | 0.01 (-0.001; 0.064) | 1,821 | 0,069 | 0,213 | PLXA1 | No |
| Interleukin-33 signaling | R-HSA-9014843 | 1 | 0,012 | 2 | 0,002 | 0.01 (-0.001; 0.064) | 1,821 | 0,069 | 0,213 | IL18R | No |
| Interleukin-37 signaling | R-HSA-9008059 | 1 | 0,012 | 2 | 0,002 | 0.01 (-0.001; 0.064) | 1,821 | 0,069 | 0,213 | IL18R | No |
| Ion transport by P-type ATPases | R-HSA-936837 | 1 | 0,012 | 2 | 0,002 | 0.01 (-0.001; 0.064) | 1,821 | 0,069 | 0,213 | AT1B2 | No |
| NPAS4 regulates expression of target genes | R-HSA-9768919 | 1 | 0,012 | 2 | 0,002 | 0.01 (-0.001; 0.064) | 1,821 | 0,069 | 0,213 | RET | No |
| SEMA3A-Plexin repulsion signaling by inhibiting Integrin adhesion | R-HSA-399955 | 1 | 0,012 | 2 | 0,002 | 0.01 (-0.001; 0.064) | 1,821 | 0,069 | 0,213 | PLXA1 | No |
| Neurodegenerative Diseases | R-HSA-8863678 | 1 | 0,012 | 2 | 0,002 | 0.01 (-0.001; 0.064) | 1,821 | 0,069 | 0,213 | CAN2 | No |
| Phosphate bond hydrolysis by NTPDase proteins | R-HSA-8850843 | 1 | 0,012 | 2 | 0,002 | 0.01 (-0.001; 0.064) | 1,821 | 0,069 | 0,213 | ENTP5 | No |
| RHO GTPases Activate NADPH Oxidases | R-HSA-5668599 | 1 | 0,012 | 2 | 0,002 | 0.01 (-0.001; 0.064) | 1,821 | 0,069 | 0,213 | KPCA | No |
| RND1 GTPase cycle | R-HSA-9696273 | 1 | 0,012 | 2 | 0,002 | 0.01 (-0.001; 0.064) | 1,821 | 0,069 | 0,213 | PLXA1 | No |
| ROBO receptors bind AKAP5 | R-HSA-9010642 | 1 | 0,012 | 2 | 0,002 | 0.01 (-0.001; 0.064) | 1,821 | 0,069 | 0,213 | KPCA | No |
| Sema3A PAK dependent Axon repulsion | R-HSA-399954 | 1 | 0,012 | 2 | 0,002 | 0.01 (-0.001; 0.064) | 1,821 | 0,069 | 0,213 | PLXA1 | No |
| Signaling by NODAL | R-HSA-1181150 | 1 | 0,012 | 2 | 0,002 | 0.01 (-0.001; 0.064) | 1,821 | 0,069 | 0,213 | TDGF1 | No |

**Table S10. Reactome pathway enrichment**

| Pathway | Pathway identifier | No. proteins in subset* | % proteins in subset* | No. proteins in data* | % proteins in data* | Enrichment (95% CI) | Test statistic | p-value | Adjusted p-value | Proteins in subset | Enriched |
|---|---|---|---|---|---|---|---|---|---|---|---|
| Synthesis of PC | R-HSA-1483191 | 1 | 0,012 | 2 | 0,002 | 0.01 (-0.001; 0.064) | 1,821 | 0,069 | 0,213 | ACES | No |
| WNT5A-dependent internalization of FZD4 | R-HSA-5099900 | 1 | 0,012 | 2 | 0,002 | 0.01 (-0.001; 0.064) | 1,821 | 0,069 | 0,213 | KPCA | No |
| Tyrosine catabolism | R-HSA-8963684 | 1 | 0,012 | 2 | 0,002 | 0.01 (-0.001; 0.064) | 1,821 | 0,069 | 0,213 | FAAA | No |
| COPI-dependent Golgi-to-ER retrograde traffic | R-HSA-6811434 | 1 | 0,012 | 2 | 0,002 | 0.01 (-0.001; 0.064) | 1,821 | 0,069 | 0,213 | KLC1 | No |
| Transcriptional Regulation by NPAS4 | R-HSA-9634815 | 1 | 0,012 | 2 | 0,002 | 0.01 (-0.001; 0.064) | 1,821 | 0,069 | 0,213 | RET | No |
| The phototransduction cascade | R-HSA-2514856 | 1 | 0,012 | 2 | 0,002 | 0.01 (-0.001; 0.064) | 1,821 | 0,069 | 0,213 | KPCA | No |
| NR1H3 & NR1H2 regulate gene expression linked to cholesterol transport a | R-HSA-9029569 | 1 | 0,012 | 2 | 0,002 | 0.01 (-0.001; 0.064) | 1,821 | 0,069 | 0,213 | APOC1 | No |
| Activation, myristolyation of BID and translocation to mitochondria | R-HSA-75108 | 1 | 0,012 | 2 | 0,002 | 0.01 (-0.001; 0.064) | 1,821 | 0,069 | 0,213 | GRAB | No |
| Synthesis of PA | R-HSA-1483166 | 1 | 0,012 | 2 | 0,002 | 0.01 (-0.001; 0.064) | 1,821 | 0,069 | 0,213 | PLA2R | No |
| Fcgamma receptor (FCGR) dependent phagocytosis | R-HSA-2029480 | 2 | 0,024 | 7 | 0,006 | 0.018 (-0.001; 0.078) | 1,836 | 0,066 | 0,213 | FCG2A, FCG3A | No |
| Signaling by SCF-KIT | R-HSA-1433557 | 2 | 0,024 | 6 | 0,005 | 0.019 (0.0; 0.079) | 2,042 | 0,041 | 0,213 | TEC, KPCA | No |
| Plasma lipoprotein assembly | R-HSA-8963898 | 2 | 0,024 | 6 | 0,005 | 0.019 (0.0; 0.079) | 2,042 | 0,041 | 0,213 | APOC1, APOC3 | No |
| Glycerophospholipid biosynthesis | R-HSA-1483206 | 2 | 0,024 | 6 | 0,005 | 0.019 (0.0; 0.079) | 2,042 | 0,041 | 0,213 | ACES, PLA2R | No |
| Ethanol oxidation | R-HSA-71384 | 2 | 0,024 | 6 | 0,005 | 0.019 (0.0; 0.079) | 2,042 | 0,041 | 0,213 | ADH1B, ADH4 | No |
| Potential therapeutics for SARS | R-HSA-9679191 | 2 | 0,024 | 8 | 0,007 | 0.017 (-0.002; 0.078) | 1,656 | 0,098 | 0,298 | IL6RA, AT1B2 | No |
| Extrinsic Pathway of Fibrin Clot Formation | R-HSA-140834 | 1 | 0,012 | 3 | 0,003 | 0.01 (-0.002; 0.063) | 1,442 | 0,149 | 0,343 | FA10 | No |
| Aflatoxin activation and detoxification | R-HSA-5423646 | 1 | 0,012 | 3 | 0,003 | 0.01 (-0.002; 0.063) | 1,442 | 0,149 | 0,343 | DPEP1 | No |
| Defective factor IX causes hemophilia B | R-HSA-9668250 | 1 | 0,012 | 3 | 0,003 | 0.01 (-0.002; 0.063) | 1,442 | 0,149 | 0,343 | FA10 | No |
| Diseases of programmed cell death | R-HSA-9645723 | 1 | 0,012 | 3 | 0,003 | 0.01 (-0.002; 0.063) | 1,442 | 0,149 | 0,343 | CAN2 | No |
| Trafficking of GluR2-containing AMPA receptors | R-HSA-416993 | 1 | 0,012 | 3 | 0,003 | 0.01 (-0.002; 0.063) | 1,442 | 0,149 | 0,343 | KPCA | No |
| Trafficking of AMPA receptors | R-HSA-399719 | 1 | 0,012 | 3 | 0,003 | 0.01 (-0.002; 0.063) | 1,442 | 0,149 | 0,343 | KPCA | No |
| Synthesis of Leukotrienes (LT) and Eoxins (EX) | R-HSA-2142691 | 1 | 0,012 | 3 | 0,003 | 0.01 (-0.002; 0.063) | 1,442 | 0,149 | 0,343 | DPEP1 | No |
| Signaling by ALK in cancer | R-HSA-9700206 | 1 | 0,012 | 3 | 0,003 | 0.01 (-0.002; 0.063) | 1,442 | 0,149 | 0,343 | KLC1 | No |
| Signaling by ALK fusions and activated point mutants | R-HSA-9725370 | 1 | 0,012 | 3 | 0,003 | 0.01 (-0.002; 0.063) | 1,442 | 0,149 | 0,343 | KLC1 | No |
| RA biosynthesis pathway | R-HSA-5365859 | 1 | 0,012 | 3 | 0,003 | 0.01 (-0.002; 0.063) | 1,442 | 0,149 | 0,343 | ADH4 | No |
| FCERI mediated Ca+2 mobilization | R-HSA-2871809 | 1 | 0,012 | 3 | 0,003 | 0.01 (-0.002; 0.063) | 1,442 | 0,149 | 0,343 | TEC | No |
| Glutamate binding, activation of AMPA receptors and synaptic plasticity | R-HSA-399721 | 1 | 0,012 | 3 | 0,003 | 0.01 (-0.002; 0.063) | 1,442 | 0,149 | 0,343 | KPCA | No |
| Golgi-to-ER retrograde transport | R-HSA-8856688 | 1 | 0,012 | 3 | 0,003 | 0.01 (-0.002; 0.063) | 1,442 | 0,149 | 0,343 | KLC1 | No |
| HDL remodeling | R-HSA-8964058 | 1 | 0,012 | 3 | 0,003 | 0.01 (-0.002; 0.063) | 1,442 | 0,149 | 0,343 | APOC3 | No |
| RAF-independent MAPK1/3 activation | R-HSA-112409 | 1 | 0,012 | 3 | 0,003 | 0.01 (-0.002; 0.063) | 1,442 | 0,149 | 0,343 | IL6RA | No |
| Interleukin-6 signaling | R-HSA-1059683 | 1 | 0,012 | 3 | 0,003 | 0.01 (-0.002; 0.063) | 1,442 | 0,149 | 0,343 | IL6RA | No |
| Inositol phosphate metabolism | R-HSA-1483249 | 1 | 0,012 | 3 | 0,003 | 0.01 (-0.002; 0.063) | 1,442 | 0,149 | 0,343 | MINP1 | No |
| NR1H2 and NR1H3-mediated signaling | R-HSA-9024446 | 1 | 0,012 | 3 | 0,003 | 0.01 (-0.002; 0.063) | 1,442 | 0,149 | 0,343 | APOC1 | No |
| Neurotransmitter clearance | R-HSA-112311 | 1 | 0,012 | 3 | 0,003 | 0.01 (-0.002; 0.063) | 1,442 | 0,149 | 0,343 | ACES | No |
| POU5F1 (OCT4), SOX2, NANOG activate genes related to proliferation | R-HSA-2892247 | 1 | 0,012 | 3 | 0,003 | 0.01 (-0.002; 0.063) | 1,442 | 0,149 | 0,343 | TDGF1 | No |
| Purine salvage | R-HSA-74217 | 1 | 0,012 | 3 | 0,003 | 0.01 (-0.002; 0.063) | 1,442 | 0,149 | 0,343 | GMPR2 | No |
| Pyroptosis | R-HSA-5620971 | 1 | 0,012 | 3 | 0,003 | 0.01 (-0.002; 0.063) | 1,442 | 0,149 | 0,343 | GRAB | No |
| MAPK3 (ERK1) activation | R-HSA-110056 | 1 | 0,012 | 3 | 0,003 | 0.01 (-0.002; 0.063) | 1,442 | 0,149 | 0,343 | IL6RA | No |
| Ephrin signaling | R-HSA-3928664 | 1 | 0,012 | 3 | 0,003 | 0.01 (-0.002; 0.063) | 1,442 | 0,149 | 0,343 | EPHB1 | No |
| Signaling by Rho GTPases | R-HSA-194315 | 4 | 0,049 | 27 | 0,024 | 0.025 (-0.007; 0.095) | 1,356 | 0,175 | 0,39 | SWP70, KPCA, KLC1, PLXA1 | No |
| Signaling by Rho GTPases, Miro GTPases and RHOBTB3 | R-HSA-9716542 | 4 | 0,049 | 27 | 0,024 | 0.025 (-0.007; 0.095) | 1,356 | 0,175 | 0,39 | SWP70, KPCA, KLC1, PLXA1 | No |
| Phospholipid metabolism | R-HSA-1483257 | 2 | 0,024 | 10 | 0,009 | 0.015 (-0.004; 0.076) | 1,356 | 0,175 | 0,39 | ACES, PLA2R | No |
| Peroxisomal lipid metabolism | R-HSA-390918 | 1 | 0,012 | 4 | 0,004 | 0.009 (-0.003; 0.062) | 1,169 | 0,242 | 0,458 | CACP | No |
| Defective factor VIII causes hemophilia A | R-HSA-9662001 | 1 | 0,012 | 4 | 0,004 | 0.009 (-0.003; 0.062) | 1,169 | 0,242 | 0,458 | FA10 | No |
| EPHB-mediated forward signaling | R-HSA-3928662 | 1 | 0,012 | 4 | 0,004 | 0.009 (-0.003; 0.062) | 1,169 | 0,242 | 0,458 | EPHB1 | No |
| Gastrin-CREB signalling pathway via PKC and MAPK | R-HSA-881907 | 1 | 0,012 | 4 | 0,004 | 0.009 (-0.003; 0.062) | 1,169 | 0,242 | 0,458 | KPCA | No |
| IRE1alpha activates chaperones | R-HSA-381070 | 1 | 0,012 | 4 | 0,004 | 0.009 (-0.003; 0.062) | 1,169 | 0,242 | 0,458 | PDIA5 | No |
| Integration of energy metabolism | R-HSA-163685 | 1 | 0,012 | 4 | 0,004 | 0.009 (-0.003; 0.062) | 1,169 | 0,242 | 0,458 | KPCA | No |
| Ion channel transport | R-HSA-983712 | 1 | 0,012 | 4 | 0,004 | 0.009 (-0.003; 0.062) | 1,169 | 0,242 | 0,458 | AT1B2 | No |
| Ion homeostasis | R-HSA-5578775 | 1 | 0,012 | 4 | 0,004 | 0.009 (-0.003; 0.062) | 1,169 | 0,242 | 0,458 | AT1B2 | No |
| Cardiac conduction | R-HSA-5576891 | 1 | 0,012 | 4 | 0,004 | 0.009 (-0.003; 0.062) | 1,169 | 0,242 | 0,458 | AT1B2 | No |
| Chylomicron assembly | R-HSA-8963888 | 1 | 0,012 | 4 | 0,004 | 0.009 (-0.003; 0.062) | 1,169 | 0,242 | 0,458 | APOC3 | No |
| Signaling by ERBB2 | R-HSA-1227986 | 1 | 0,012 | 4 | 0,004 | 0.009 (-0.003; 0.062) | 1,169 | 0,242 | 0,458 | KPCA | No |
| Signaling by NOTCH2 | R-HSA-1980145 | 1 | 0,012 | 4 | 0,004 | 0.009 (-0.003; 0.062) | 1,169 | 0,242 | 0,458 | GRAB | No |
| TP53 Regulates Transcription of Death Receptors and Ligands | R-HSA-6803211 | 1 | 0,012 | 4 | 0,004 | 0.009 (-0.003; 0.062) | 1,169 | 0,242 | 0,458 | IBP3 | No |
| Transcriptional regulation of pluripotent stem cells | R-HSA-452723 | 1 | 0,012 | 4 | 0,004 | 0.009 (-0.003; 0.062) | 1,169 | 0,242 | 0,458 | TDGF1 | No |
| VEGFR2 mediated cell proliferation | R-HSA-5218921 | 1 | 0,012 | 4 | 0,004 | 0.009 (-0.003; 0.062) | 1,169 | 0,242 | 0,458 | KPCA | No |
| MHC class II antigen presentation | R-HSA-2132295 | 2 | 0,024 | 11 | 0,01 | 0.015 (-0.005; 0.075) | 1,228 | 0,219 | 0,458 | KLC1, CATF | No |

**Table S10. Reactome pathway enrichment**

| Pathway | Pathway identifier | No. proteins in subset* | % proteins in subset* | No. proteins in data* | % proteins in data* | Enrichment (95% CI) | Test statistic | p-value | Adjusted p-value | Proteins in subset | Enriched |
|---|---|---|---|---|---|---|---|---|---|---|---|
| XBP1(S) activates chaperone genes | R-HSA-381038 | 1 | 0,012 | 4 | 0,004 | 0.009 (-0.003; 0.062) | 1,169 | 0,242 | 0,458 | PDIA5 | No |
| SHC1 events in ERBB2 signaling | R-HSA-1250196 | 1 | 0,012 | 4 | 0,004 | 0.009 (-0.003; 0.062) | 1,169 | 0,242 | 0,458 | KPCA | No |
| Transmission across Chemical Synapses | R-HSA-112315 | 2 | 0,024 | 12 | 0,011 | 0.014 (-0.006; 0.074) | 1,111 | 0,266 | 0,495 | KPCA, ACES | No |
| RHO GTPase Effectors | R-HSA-195258 | 2 | 0,024 | 12 | 0,011 | 0.014 (-0.006; 0.074) | 1,111 | 0,266 | 0,495 | KPCA, KLC1 | No |
| Gamma-carboxylation of protein precursors | R-HSA-159740 | 1 | 0,012 | 5 | 0,004 | 0.008 (-0.004; 0.061) | 0,957 | 0,339 | 0,538 | FA10 | No |
| Surfactant metabolism | R-HSA-5683826 | 1 | 0,012 | 5 | 0,004 | 0.008 (-0.004; 0.061) | 0,957 | 0,339 | 0,538 | CECR1 | No |
| Removal of aminoterminal propeptides from gamma-carboxylated protein | R-HSA-159782 | 1 | 0,012 | 5 | 0,004 | 0.008 (-0.004; 0.061) | 0,957 | 0,339 | 0,538 | FA10 | No |
| Regulation of ornithine decarboxylase (ODC) | R-HSA-350562 | 1 | 0,012 | 5 | 0,004 | 0.008 (-0.004; 0.061) | 0,957 | 0,339 | 0,538 | NQO1 | No |
| Gamma-carboxylation, transport, and amino-terminal cleavage of proteins | R-HSA-159854 | 1 | 0,012 | 5 | 0,004 | 0.008 (-0.004; 0.061) | 0,957 | 0,339 | 0,538 | FA10 | No |
| Gluconeogenesis | R-HSA-70263 | 1 | 0,012 | 5 | 0,004 | 0.008 (-0.004; 0.061) | 0,957 | 0,339 | 0,538 | ENOB | No |
| RAC2 GTPase cycle | R-HSA-9013404 | 1 | 0,012 | 5 | 0,004 | 0.008 (-0.004; 0.061) | 0,957 | 0,339 | 0,538 | SWP70 | No |
| Phenylalanine and tyrosine metabolism | R-HSA-8963691 | 1 | 0,012 | 5 | 0,004 | 0.008 (-0.004; 0.061) | 0,957 | 0,339 | 0,538 | FAAA | No |
| Leishmania phagocytosis | R-HSA-9664417 | 1 | 0,012 | 5 | 0,004 | 0.008 (-0.004; 0.061) | 0,957 | 0,339 | 0,538 | FCG3A | No |
| Metabolism of polyamines | R-HSA-351202 | 1 | 0,012 | 5 | 0,004 | 0.008 (-0.004; 0.061) | 0,957 | 0,339 | 0,538 | NQO1 | No |
| Peptide hormone biosynthesis | R-HSA-209952 | 1 | 0,012 | 5 | 0,004 | 0.008 (-0.004; 0.061) | 0,957 | 0,339 | 0,538 | NEC1 | No |
| NFE2L2 regulating anti-oxidant/detoxification enzymes | R-HSA-9818027 | 1 | 0,012 | 5 | 0,004 | 0.008 (-0.004; 0.061) | 0,957 | 0,339 | 0,538 | NQO1 | No |
| Parasite infection | R-HSA-9664407 | 1 | 0,012 | 5 | 0,004 | 0.008 (-0.004; 0.061) | 0,957 | 0,339 | 0,538 | FCG3A | No |
| Neurotransmitter receptors and postsynaptic signal transmission | R-HSA-112314 | 1 | 0,012 | 5 | 0,004 | 0.008 (-0.004; 0.061) | 0,957 | 0,339 | 0,538 | KPCA | No |
| Transport of gamma-carboxylated protein precursors from the endoplasmi | R-HSA-159763 | 1 | 0,012 | 5 | 0,004 | 0.008 (-0.004; 0.061) | 0,957 | 0,339 | 0,538 | FA10 | No |
| FCGR3A-mediated phagocytosis | R-HSA-9664422 | 1 | 0,012 | 5 | 0,004 | 0.008 (-0.004; 0.061) | 0,957 | 0,339 | 0,538 | FCG3A | No |
| Nucleotide salvage | R-HSA-8956321 | 1 | 0,012 | 5 | 0,004 | 0.008 (-0.004; 0.061) | 0,957 | 0,339 | 0,538 | GMPR2 | No |
| Phase I - Functionalization of compounds | R-HSA-211945 | 2 | 0,024 | 13 | 0,012 | 0.013 (-0.007; 0.073) | 1,004 | 0,316 | 0,538 | ADH4, ADH1B | No |
| Cholesterol biosynthesis | R-HSA-191273 | 1 | 0,012 | 5 | 0,004 | 0.008 (-0.004; 0.061) | 0,957 | 0,339 | 0,538 | IDI2 | No |
| Immunoregulatory interactions between a Lymphoid and a non-Lymphoid | R-HSA-198933 | 5 | 0,061 | 44 | 0,039 | 0.022 (-0.015; 0.096) | 0,954 | 0,34 | 0,538 | NCTR3, MICB, FCG3A, LIRB5, MICA | No |
| Nucleotide catabolism | R-HSA-8956319 | 2 | 0,024 | 13 | 0,012 | 0.013 (-0.007; 0.073) | 1,004 | 0,316 | 0,538 | NUDT9, ENTP5 | No |
| Cell Cycle, Mitotic | R-HSA-69278 | 2 | 0,024 | 14 | 0,013 | 0.012 (-0.008; 0.072) | 0,904 | 0,366 | 0,575 | KPCA, MAX | No |
| Adaptive Immune System | R-HSA-1280218 | 9 | 0,11 | 91 | 0,081 | 0.028 (-0.025; 0.115) | 0,897 | 0,37 | 0,577 | FCG3A, KLC1, MICA, NCTR3, MICB, ERAP1, LIRB5, TPSN, CA | No |
| Chylomicron remodeling | R-HSA-8963901 | 1 | 0,012 | 6 | 0,005 | 0.007 (-0.005; 0.061) | 0,783 | 0,433 | 0,604 | APOC3 | No |
| Cyclin A:Cdk2-associated events at S phase entry | R-HSA-69656 | 1 | 0,012 | 6 | 0,005 | 0.007 (-0.005; 0.061) | 0,783 | 0,433 | 0,604 | MAX | No |
| Cyclin E associated events during G1/S transition | R-HSA-69202 | 1 | 0,012 | 6 | 0,005 | 0.007 (-0.005; 0.061) | 0,783 | 0,433 | 0,604 | MAX | No |
| Interleukin-3, Interleukin-5 and GM-CSF signaling | R-HSA-512988 | 1 | 0,012 | 6 | 0,005 | 0.007 (-0.005; 0.061) | 0,783 | 0,433 | 0,604 | TEC | No |
| Signaling by Retinoic Acid | R-HSA-5362517 | 1 | 0,012 | 6 | 0,005 | 0.007 (-0.005; 0.061) | 0,783 | 0,433 | 0,604 | ADH4 | No |
| Purinergic signaling in leishmaniasis infection | R-HSA-9660826 | 1 | 0,012 | 6 | 0,005 | 0.007 (-0.005; 0.061) | 0,783 | 0,433 | 0,604 | ENTP5 | No |
| Cell-cell junction organization | R-HSA-421270 | 1 | 0,012 | 6 | 0,005 | 0.007 (-0.005; 0.061) | 0,783 | 0,433 | 0,604 | PVRL4 | No |
| RAC3 GTPase cycle | R-HSA-9013423 | 1 | 0,012 | 6 | 0,005 | 0.007 (-0.005; 0.061) | 0,783 | 0,433 | 0,604 | SWP70 | No |
| Syndecan interactions | R-HSA-3000170 | 1 | 0,012 | 6 | 0,005 | 0.007 (-0.005; 0.061) | 0,783 | 0,433 | 0,604 | KPCA | No |
| TP53 Regulates Transcription of Cell Death Genes | R-HSA-5633008 | 1 | 0,012 | 6 | 0,005 | 0.007 (-0.005; 0.061) | 0,783 | 0,433 | 0,604 | IBP3 | No |
| Mitotic G1 phase and G1/S transition | R-HSA-453279 | 1 | 0,012 | 6 | 0,005 | 0.007 (-0.005; 0.061) | 0,783 | 0,433 | 0,604 | MAX | No |
| Cell recruitment (pro-inflammatory response) | R-HSA-9664424 | 1 | 0,012 | 6 | 0,005 | 0.007 (-0.005; 0.061) | 0,783 | 0,433 | 0,604 | ENTP5 | No |
| G1/S Transition | R-HSA-69206 | 1 | 0,012 | 6 | 0,005 | 0.007 (-0.005; 0.061) | 0,783 | 0,433 | 0,604 | MAX | No |
| Transport of small molecules | R-HSA-382551 | 4 | 0,049 | 36 | 0,032 | 0.017 (-0.015; 0.087) | 0,807 | 0,42 | 0,604 | AT1B2, APOC3, APOC1, ZA2G | No |
| Cell Cycle | R-HSA-1640170 | 2 | 0,024 | 15 | 0,013 | 0.011 (-0.009; 0.071) | 0,811 | 0,417 | 0,604 | KPCA, MAX | No |
| Infectious disease | R-HSA-5663205 | 7 | 0,085 | 69 | 0,062 | 0.024 (-0.022; 0.105) | 0,848 | 0,396 | 0,604 | AT1B2, ENTP5, FCG3A, HAVR1, FCG2A, DPEP1, IL6RA | No |
| Adherens junctions interactions | R-HSA-418990 | 1 | 0,012 | 6 | 0,005 | 0.007 (-0.005; 0.061) | 0,783 | 0,433 | 0,604 | PVRL4 | No |
| RHO GTPase cycle | R-HSA-9012999 | 2 | 0,024 | 16 | 0,014 | 0.01 (-0.01; 0.071) | 0,724 | 0,469 | 0,641 | SWP70, PLXA1 | No |
| Plasma lipoprotein assembly, remodeling, and clearance | R-HSA-174824 | 2 | 0,024 | 16 | 0,014 | 0.01 (-0.01; 0.071) | 0,724 | 0,469 | 0,641 | APOC1, APOC3 | No |
| Peptide hormone metabolism | R-HSA-2980736 | 2 | 0,024 | 16 | 0,014 | 0.01 (-0.01; 0.071) | 0,724 | 0,469 | 0,641 | NEC1, ACES | No |
| Biological oxidations | R-HSA-211859 | 4 | 0,049 | 38 | 0,034 | 0.015 (-0.017; 0.085) | 0,703 | 0,482 | 0,655 | ADH1B, DPEP1, ADH4, GSTM3 | No |
| Metabolism of nucleotides | R-HSA-15869 | 3 | 0,037 | 27 | 0,024 | 0.012 (-0.014; 0.078) | 0,696 | 0,487 | 0,658 | GMPR2, NUDT9, ENTP5 | No |
| Purine catabolism | R-HSA-74259 | 1 | 0,012 | 7 | 0,006 | 0.006 (-0.006; 0.06) | 0,637 | 0,524 | 0,676 | NUDT9 | No |
| Plasma lipoprotein clearance | R-HSA-8964043 | 1 | 0,012 | 7 | 0,006 | 0.006 (-0.006; 0.06) | 0,637 | 0,524 | 0,676 | APOC1 | No |
| Peroxisomal protein import | R-HSA-9033241 | 1 | 0,012 | 7 | 0,006 | 0.006 (-0.006; 0.06) | 0,637 | 0,524 | 0,676 | CACP | No |
| Intrinsic Pathway for Apoptosis | R-HSA-109606 | 1 | 0,012 | 7 | 0,006 | 0.006 (-0.006; 0.06) | 0,637 | 0,524 | 0,676 | GRAB | No |
| HS-GAG biosynthesis | R-HSA-2022928 | 1 | 0,012 | 7 | 0,006 | 0.006 (-0.006; 0.06) | 0,637 | 0,524 | 0,676 | GLCE | No |
| S Phase | R-HSA-69242 | 1 | 0,012 | 7 | 0,006 | 0.006 (-0.006; 0.06) | 0,637 | 0,524 | 0,676 | MAX | No |
| Glycolysis | R-HSA-70171 | 1 | 0,012 | 7 | 0,006 | 0.006 (-0.006; 0.06) | 0,637 | 0,524 | 0,676 | ENOB | No |
| Translation | R-HSA-72766 | 1 | 0,012 | 7 | 0,006 | 0.006 (-0.006; 0.06) | 0,637 | 0,524 | 0,676 | RF1ML | No |
| MAPK1/MAPK3 signaling | R-HSA-5684996 | 4 | 0,049 | 41 | 0,037 | 0.012 (-0.02; 0.083) | 0,557 | 0,578 | 0,74 | GFRA1, IL6RA, RET, I17RD | No |

**Table S10. Reactome pathway enrichment**

| Pathway | Pathway identifier | No. proteins in subset* | % proteins in subset* | No. proteins in data* | % proteins in data* | Enrichment (95% CI) | Test statistic | p-value | Adjusted p-value | Proteins in subset | Enriched |
|---|---|---|---|---|---|---|---|---|---|---|---|
| Digestion | R-HSA-8935690 | 1 | 0,012 | 8 | 0,007 | 0.005 (-0.007; 0.059) | 0,51 | 0,61 | 0,76 | CEL | No |
| Digestion and absorption | R-HSA-8963743 | 1 | 0,012 | 8 | 0,007 | 0.005 (-0.007; 0.059) | 0,51 | 0,61 | 0,76 | CEL | No |
| Early SARS-CoV-2 Infection Events | R-HSA-9772572 | 1 | 0,012 | 8 | 0,007 | 0.005 (-0.007; 0.059) | 0,51 | 0,61 | 0,76 | HAVR1 | No |
| RAC1 GTPase cycle | R-HSA-9013149 | 1 | 0,012 | 8 | 0,007 | 0.005 (-0.007; 0.059) | 0,51 | 0,61 | 0,76 | SWP70 | No |
| Interleukin-6 family signaling | R-HSA-6783589 | 1 | 0,012 | 8 | 0,007 | 0.005 (-0.007; 0.059) | 0,51 | 0,61 | 0,76 | IL6RA | No |
| Attachment and Entry | R-HSA-9694614 | 1 | 0,012 | 9 | 0,008 | 0.004 (-0.008; 0.058) | 0,398 | 0,69 | 0,794 | HAVR1 | No |
| Diseases of hemostasis | R-HSA-9671793 | 1 | 0,012 | 9 | 0,008 | 0.004 (-0.008; 0.058) | 0,398 | 0,69 | 0,794 | FA10 | No |
| Beta-catenin independent WNT signaling | R-HSA-3858494 | 1 | 0,012 | 9 | 0,008 | 0.004 (-0.008; 0.058) | 0,398 | 0,69 | 0,794 | KPCA | No |
| Cell junction organization | R-HSA-446728 | 1 | 0,012 | 9 | 0,008 | 0.004 (-0.008; 0.058) | 0,398 | 0,69 | 0,794 | PVRL4 | No |
| Attachment and Entry | R-HSA-9678110 | 1 | 0,012 | 9 | 0,008 | 0.004 (-0.008; 0.058) | 0,398 | 0,69 | 0,794 | HAVR1 | No |
| Defects of contact activation system (CAS) and kallikrein/kinin system (KKS | R-HSA-9651496 | 1 | 0,012 | 9 | 0,008 | 0.004 (-0.008; 0.058) | 0,398 | 0,69 | 0,794 | FA10 | No |
| MAPK family signaling cascades | R-HSA-5683057 | 4 | 0,049 | 44 | 0,039 | 0.009 (-0.023; 0.08) | 0,42 | 0,674 | 0,794 | GFRA1, IL6RA, RET, I17RD | No |
| NCAM1 interactions | R-HSA-419037 | 1 | 0,012 | 9 | 0,008 | 0.004 (-0.008; 0.058) | 0,398 | 0,69 | 0,794 | GFRA1 | No |
| PCP/CE pathway | R-HSA-4086400 | 1 | 0,012 | 9 | 0,008 | 0.004 (-0.008; 0.058) | 0,398 | 0,69 | 0,794 | KPCA | No |
| Regulated Necrosis | R-HSA-5218859 | 1 | 0,012 | 9 | 0,008 | 0.004 (-0.008; 0.058) | 0,398 | 0,69 | 0,794 | GRAB | No |
| Intra-Golgi and retrograde Golgi-to-ER traffic | R-HSA-6811442 | 1 | 0,012 | 9 | 0,008 | 0.004 (-0.008; 0.058) | 0,398 | 0,69 | 0,794 | KLC1 | No |
| Visual phototransduction | R-HSA-2187338 | 2 | 0,024 | 20 | 0,018 | 0.007 (-0.014; 0.067) | 0,423 | 0,672 | 0,794 | KPCA, APOC3 | No |
| Regulation of mRNA stability by proteins that bind AU-rich elements | R-HSA-450531 | 1 | 0,012 | 9 | 0,008 | 0.004 (-0.008; 0.058) | 0,398 | 0,69 | 0,794 | KPCA | No |
| Arachidonic acid metabolism | R-HSA-2142753 | 1 | 0,012 | 9 | 0,008 | 0.004 (-0.008; 0.058) | 0,398 | 0,69 | 0,794 | DPEP1 | No |
| MAP2K and MAPK activation | R-HSA-5674135 | 1 | 0,012 | 9 | 0,008 | 0.004 (-0.008; 0.058) | 0,398 | 0,69 | 0,794 | I17RD | No |
| Class I MHC mediated antigen processing & presentation | R-HSA-983169 | 2 | 0,024 | 21 | 0,019 | 0.006 (-0.015; 0.066) | 0,357 | 0,721 | 0,824 | TPSN, ERAP1 | No |
| Unfolded Protein Response (UPR) | R-HSA-381119 | 1 | 0,012 | 10 | 0,009 | 0.003 (-0.009; 0.057) | 0,298 | 0,766 | 0,84 | PDIA5 | No |
| Other semaphorin interactions | R-HSA-416700 | 1 | 0,012 | 10 | 0,009 | 0.003 (-0.009; 0.057) | 0,298 | 0,766 | 0,84 | PLXA1 | No |
| VEGFA-VEGFR2 Pathway | R-HSA-4420097 | 1 | 0,012 | 10 | 0,009 | 0.003 (-0.009; 0.057) | 0,298 | 0,766 | 0,84 | KPCA | No |
| Plasma lipoprotein remodeling | R-HSA-8963899 | 1 | 0,012 | 10 | 0,009 | 0.003 (-0.009; 0.057) | 0,298 | 0,766 | 0,84 | APOC3 | No |
| Muscle contraction | R-HSA-397014 | 1 | 0,012 | 10 | 0,009 | 0.003 (-0.009; 0.057) | 0,298 | 0,766 | 0,84 | AT1B2 | No |
| Sensory Perception | R-HSA-9709957 | 2 | 0,024 | 22 | 0,02 | 0.005 (-0.016; 0.065) | 0,294 | 0,769 | 0,84 | APOC3, KPCA | No |
| Gamma carboxylation, hypusine formation and arylsulfatase activation | R-HSA-163841 | 1 | 0,012 | 10 | 0,009 | 0.003 (-0.009; 0.057) | 0,298 | 0,766 | 0,84 | FA10 | No |
| Fatty acid metabolism | R-HSA-8978868 | 2 | 0,024 | 22 | 0,02 | 0.005 (-0.016; 0.065) | 0,294 | 0,769 | 0,84 | CACP, DPEP1 | No |
| Glucose metabolism | R-HSA-70326 | 1 | 0,012 | 10 | 0,009 | 0.003 (-0.009; 0.057) | 0,298 | 0,766 | 0,84 | ENOB | No |
| SARS-CoV Infections | R-HSA-9679506 | 3 | 0,037 | 35 | 0,031 | 0.005 (-0.022; 0.071) | 0,263 | 0,792 | 0,862 | AT1B2, IL6RA, HAVR1 | No |
| ER-Phagosome pathway | R-HSA-1236974 | 1 | 0,012 | 11 | 0,01 | 0.002 (-0.01; 0.056) | 0,207 | 0,836 | 0,892 | TPSN | No |
| NCAM signaling for neurite out-growth | R-HSA-375165 | 1 | 0,012 | 11 | 0,01 | 0.002 (-0.01; 0.056) | 0,207 | 0,836 | 0,892 | GFRA1 | No |
| Protein localization | R-HSA-9609507 | 1 | 0,012 | 11 | 0,01 | 0.002 (-0.01; 0.056) | 0,207 | 0,836 | 0,892 | CACP | No |
| TNFs bind their physiological receptors | R-HSA-5669034 | 1 | 0,012 | 11 | 0,01 | 0.002 (-0.01; 0.056) | 0,207 | 0,836 | 0,892 | EDAR | No |
| Glutathione conjugation | R-HSA-156590 | 1 | 0,012 | 12 | 0,011 | 0.001 (-0.011; 0.055) | 0,123 | 0,902 | 0,949 | GSTM3 | No |
| Fc epsilon receptor (FCERI) signaling | R-HSA-2454202 | 1 | 0,012 | 12 | 0,011 | 0.001 (-0.011; 0.055) | 0,123 | 0,902 | 0,949 | TEC | No |
| EPH-ephrin mediated repulsion of cells | R-HSA-3928665 | 1 | 0,012 | 12 | 0,011 | 0.001 (-0.011; 0.055) | 0,123 | 0,902 | 0,949 | EPHB1 | No |
| Transcriptional Regulation by TP53 | R-HSA-3700989 | 1 | 0,012 | 13 | 0,012 | 0.001 (-0.012; 0.054) | 0,046 | 0,963 | 0,981 | IBP3 | No |
| Semaphorin interactions | R-HSA-373755 | 1 | 0,012 | 13 | 0,012 | 0.001 (-0.012; 0.054) | 0,046 | 0,963 | 0,981 | PLXA1 | No |
| KEAP1-NFE2L2 pathway | R-HSA-9755511 | 1 | 0,012 | 13 | 0,012 | 0.001 (-0.012; 0.054) | 0,046 | 0,963 | 0,981 | NQO1 | No |
| Non-integrin membrane-ECM interactions | R-HSA-3000171 | 1 | 0,012 | 13 | 0,012 | 0.001 (-0.012; 0.054) | 0,046 | 0,963 | 0,981 | KPCA | No |
| M Phase | R-HSA-68886 | 1 | 0,012 | 13 | 0,012 | 0.001 (-0.012; 0.054) | 0,046 | 0,963 | 0,981 | KPCA | No |
| EPH-Ephrin signaling | R-HSA-2682334 | 1 | 0,012 | 13 | 0,012 | 0.001 (-0.012; 0.054) | 0,046 | 0,963 | 0,981 | EPHB1 | No |
| Nuclear events mediated by NFE2L2 | R-HSA-9759194 | 1 | 0,012 | 13 | 0,012 | 0.001 (-0.012; 0.054) | 0,046 | 0,963 | 0,981 | NQO1 | No |
| RAF/MAP kinase cascade | R-HSA-5673001 | 3 | 0,037 | 40 | 0,036 | 0.001 (-0.026; 0.067) | 0,038 | 0,97 | 0,983 | GFRA1, RET, I17RD | No |
| Neuronal System | R-HSA-112316 | 2 | 0,024 | 27 | 0,024 | 0.0 (-0.02; 0.061) | 0,014 | 0,989 | 0,989 | KPCA, ACES | No |
| Intracellular signaling by second messengers | R-HSA-9006925 | 2 | 0,024 | 27 | 0,024 | 0.0 (-0.02; 0.061) | 0,014 | 0,989 | 0,989 | KPCA, IL18R | No |
| Signaling by Nuclear Receptors | R-HSA-9006931 | 2 | 0,024 | 27 | 0,024 | 0.0 (-0.02; 0.061) | 0,014 | 0,989 | 0,989 | ADH4, APOC1 | No |

* Columns: No. proteins in subset - number of prioritised proteins belonging to indicated pathway, % proteins in subset - percentage of proteins in subset belonging to indicated pathway, No. of proteins in data - number of proteins in reference belonging to indicated pathway, % proteins in data - percentage of proteins in reference belonging to indicated pathway, see Methods section for more detailed description.

Abbreviations: CI = confidence interval
