## Supplemental Table S11 for "Integrating metabolomics and proteomics to identify novel drug targets for heart failure and atrial fibrillation"

**Table S11. Replicates of the proteins associated with cardiac outcomes**

| Protein name | Point estimate* | p-value* | Outcome | Discovery study* | No. other occurences* | Other point estimates* | Other p-values* | Nominal replicates* | Adjusted replicates* | Studies* |
|---|---|---|---|---|---|---|---|---|---|---|
| G6PE | -0,041 | $7.8\times10^{-7}$ | AF | deCODE | 2 | -0.003; -0.025 | $7.5\times10^{-1}$; $1.1\times10^{-1}$ | 0 | 0 | AGES-Reykjavik; INTERVAL |
| PDIA5 | 0,033 | $1.5\times10^{-6}$ | AF | deCODE | 2 | 0.017; 0.026 | $2.5\times10^{-9}$; $6.6\times10^{-3}$ | 2 | 1 | AGES-Reykjavik; INTERVAL |
| GNPTG | -0,059 | $4.3\times10^{-9}$ | AF | deCODE | 2 | -0.157; -0.001 | $1.2\times10^{-2}$; $9.7\times10^{-1}$ | 1 | 0 | AGES-Reykjavik; INTERVAL |
| GNPTG | 0,075 | $1.4\times10^{-10}$ | HF | deCODE | 2 | 0.115; 0.074 | $6.0\times10^{-5}$; $2.1\times10^{-4}$ | 2 | 2 | AGES-Reykjavik; INTERVAL |
| GNPTG | 0,338 | $4.1\times10^{-6}$ | DCM | deCODE | 2 | 1.279; 0.114 | $1.3\times10^{-3}$; $2.7\times10^{-1}$ | 1 | 0 | AGES-Reykjavik; INTERVAL |
| CECR1 | -0,193 | $8.3\times10^{-6}$ | DCM | deCODE | 2 | -0.214; -0.243 | $7.0\times10^{-9}$; $2.1\times10^{-3}$ | 2 | 1 | AGES-Reykjavik; INTERVAL |
| TIMP3 | 0,03 | $1.3\times10^{-11}$ | AF | deCODE | 1 | 0.013 | $5.2\times10^{-2}$ | 0 | 0 | AGES-Reykjavik |
| GRAB | 0,189 | $1.5\times10^{-5}$ | HF | deCODE | 1 | -0.02 | $1.5\times10^{-1}$ | 0 | 0 | INTERVAL |
| FAAA | -0,106 | $1.0\times10^{-100}$ | AF | deCODE | 2 | -0.069; -0.066 | $1.7\times10^{-3}$; $4.2\times10^{-5}$ | 2 | 1 | AGES-Reykjavik; INTERVAL |
| LIRB5 | 0,021 | $2.3\times10^{-6}$ | AF | deCODE | 2 | 0.009; 0.017 | $2.2\times10^{-1}$; $5.8\times10^{-3}$ | 1 | 0 | AGES-Reykjavik; INTERVAL |
| TICN2 | -0,092 | $1.2\times10^{-9}$ | AF | deCODE | 2 | 0.143; -0.029 | $3.5\times10^{-2}$; $1.7\times10^{-1}$ | 0 | 0 | AGES-Reykjavik; INTERVAL |
| MINP1 | -0,22 | $5.6\times10^{-6}$ | HF | deCODE | 2 | 0.129; -0.05 | $1.2\times10^{-1}$; $1.7\times10^{-1}$ | 0 | 0 | AGES-Reykjavik; INTERVAL |
| MINP1 | 0,826 | $1.4\times10^{-6}$ | NICM | deCODE | 1 | 0.16 | $1.1\times10^{-1}$ | 0 | 0 | AGES-Reykjavik |
| CCL8 | 0,023 | $2.5\times10^{-7}$ | AF | deCODE | 1 | 0.055 | $7.1\times10^{-3}$ | 1 | 0 | AGES-Reykjavik |
| APOC3 | 0,255 | $5.5\times10^{-8}$ | HF | deCODE | 1 | 0.065 | $2.9\times10^{-2}$ | 1 | 0 | AGES-Reykjavik |
| APOC3 | 0,764 | $1.2\times10^{-7}$ | DCM | deCODE | 1 | -0.172 | $5.8\times10^{-1}$ | 0 | 0 | AGES-Reykjavik |
| HAVR1 | 0,052 | $3.7\times10^{-8}$ | AF | deCODE | 4 | 0.017; 0.048; 0.043; -0.007 | $7.5\times10^{-2}$; $1.3\times10^{-5}$; $4.1\times10^{-2}$; $4.2\times10^{-1}$ | 2 | 1 | SCALLOP; AGES-Reykjavik; INTERVAL; Gilly |
| HAVR1 | 0,059 | $7.6\times10^{-8}$ | HF | deCODE | 4 | 0.02; 0.049; -0.03; 0.025 | $1.7\times10^{-1}$; $1.5\times10^{-4}$; $2.2\times10^{-1}$; $1.6\times10^{-2}$ | 2 | 1 | SCALLOP; AGES-Reykjavik; INTERVAL; Gilly |
| PLXA1 | 0,086 | $8.9\times10^{-10}$ | AF | deCODE | 1 | -0.098 | $9.2\times10^{-2}$ | 0 | 0 | AGES-Reykjavik |
| PLXA1 | -0,108 | $1.0\times10^{-100}$ | HF | deCODE | 1 | -0.079 | $2.9\times10^{-2}$ | 1 | 0 | AGES-Reykjavik |
| PLXA1 | -0,283 | $1.6\times10^{-11}$ | NICM | deCODE | 1 | -0.273 | $1.0\times10^{-1}$ | 0 | 0 | AGES-Reykjavik |
| IL18R | -0,047 | $3.8\times10^{-11}$ | AF | deCODE | 2 | -0.034; -0.024 | $1.5\times10^{-5}$; $1.8\times10^{-9}$ | 2 | 2 | AGES-Reykjavik; INTERVAL |
| TIG1 | 0,026 | $1.0\times10^{-100}$ | HF | deCODE | 2 | 0.017; 0.004 | $5.1\times10^{-4}$; $7.3\times10^{-1}$ | 1 | 0 | AGES-Reykjavik; INTERVAL |
| TIG1 | 0,19 | $8.0\times10^{-14}$ | DCM | deCODE | 2 | 0.165; 0.176 | $1.0\times10^{-100}$; $3.7\times10^{-6}$ | 2 | 2 | AGES-Reykjavik; INTERVAL |
| FA10 | 0,102 | $8.6\times10^{-7}$ | AF | deCODE | 2 | 0.175; 0.078 | $6.3\times10^{-3}$; $3.1\times10^{-12}$ | 2 | 1 | AGES-Reykjavik; INTERVAL |
| IGFR1 | 0,053 | $6.5\times10^{-8}$ | AF | deCODE | 2 | 0.062; -0.007 | $4.8\times10^{-8}$; $7.8\times10^{-1}$ | 1 | 1 | AGES-Reykjavik; INTERVAL |
| IGFR1 | -0,169 | $1.8\times10^{-5}$ | DCM | deCODE | 2 | -0.079; -0.064 | $2.8\times10^{-1}$; $2.5\times10^{-1}$ | 0 | 0 | AGES-Reykjavik; INTERVAL |
| ISK2 | 0,097 | $7.8\times10^{-9}$ | AF | deCODE | 2 | 0.107; 0.027 | $1.1\times10^{-4}$; $4.9\times10^{-1}$ | 1 | 1 | AGES-Reykjavik; INTERVAL |
| AT1B2 | 0,288 | $5.4\times10^{-7}$ | NICM | deCODE | 2 | 0.439; 0.188 | $1.0\times10^{-100}$; $1.7\times10^{-6}$ | 2 | 2 | AGES-Reykjavik; INTERVAL |
| AT1B2 | 0,315 | $3.1\times10^{-9}$ | DCM | deCODE | 2 | 0.164; 0.254 | $1.5\times10^{-2}$; $1.1\times10^{-5}$ | 2 | 1 | AGES-Reykjavik; INTERVAL |
| C1QRF | 0,053 | $3.3\times10^{-9}$ | AF | deCODE | 1 | 0.038 | $1.2\times10^{-5}$ | 1 | 1 | AGES-Reykjavik |
| SWP70 | -0,045 | $4.0\times10^{-9}$ | HF | deCODE | 2 | -0.03; 0.015 | $1.6\times10^{-3}$; $3.7\times10^{-1}$ | 1 | 0 | AGES-Reykjavik; INTERVAL |
| GSTM3 | 0,035 | $3.1\times10^{-7}$ | AF | deCODE | 1 | -0.03 | $5.1\times10^{-1}$ | 0 | 0 | AGES-Reykjavik |
| EDAR | -0,088 | $4.1\times10^{-6}$ | HF | deCODE | 2 | 0.005; 0.001 | $7.8\times10^{-1}$; $9.4\times10^{-1}$ | 0 | 0 | AGES-Reykjavik; INTERVAL |
| GLCE | -0,059 | $1.0\times10^{-100}$ | AF | deCODE | 2 | -0.026; -0.047 | $8.5\times10^{-8}$; $1.9\times10^{-3}$ | 2 | 1 | AGES-Reykjavik; INTERVAL |
| SAT2 | 0,14 | $1.2\times10^{-5}$ | AF | deCODE | 1 | 0.214 | $5.0\times10^{-2}$ | 1 | 0 | INTERVAL |
| PVRL4 | -1,136 | $1.3\times10^{-6}$ | NICM | deCODE | 1 | -0.477 | $1.3\times10^{-4}$ | 1 | 1 | AGES-Reykjavik |
| FCG2A | -0,015 | $1.1\times10^{-8}$ | AF | deCODE | 2 | 0.002; -0.02 | $7.0\times10^{-1}$; $5.1\times10^{-6}$ | 1 | 1 | AGES-Reykjavik; INTERVAL |
| FCG2A | -0,015 | $3.5\times10^{-7}$ | HF | deCODE | 2 | -0.013; -0.024 | $1.8\times10^{-4}$; $3.6\times10^{-13}$ | 2 | 2 | AGES-Reykjavik; INTERVAL |
| REG3G | -0,029 | $3.2\times10^{-6}$ | AF | deCODE | 1 | -0.025 | $4.8\times10^{-5}$ | 1 | 1 | AGES-Reykjavik |
| I17RD | -0,271 | $1.0\times10^{-5}$ | DCM | deCODE | 2 | -0.255; -0.183 | $1.1\times10^{-8}$; $4.3\times10^{-2}$ | 2 | 1 | AGES-Reykjavik; INTERVAL |
| GXLT1 | 0,518 | $6.0\times10^{-6}$ | DCM | deCODE | 2 | 0.307; 0.243 | $1.8\times10^{-6}$; $2.2\times10^{-2}$ | 2 | 1 | AGES-Reykjavik; INTERVAL |
| GFRA1 | -0,091 | $3.4\times10^{-9}$ | AF | deCODE | 2 | -0.034; -0.051 | $1.6\times10^{-2}$; $8.5\times10^{-4}$ | 2 | 0 | AGES-Reykjavik; INTERVAL |
| GFRA1 | -0,38 | $2.3\times10^{-6}$ | NICM | deCODE | 2 | -0.343; -0.047 | $3.9\times10^{-9}$; $5.2\times10^{-1}$ | 1 | 1 | AGES-Reykjavik; INTERVAL |
| PLA2R | 0,011 | $8.6\times10^{-6}$ | HF | deCODE | 2 | 0.006; 0.005 | $1.2\times10^{-1}$; $6.3\times10^{-2}$ | 0 | 0 | AGES-Reykjavik; INTERVAL |
| PLA2R | -0,081 | $1.0\times10^{-100}$ | DCM | deCODE | 2 | -0.172; -0.165 | $1.4\times10^{-3}$; $6.5\times10^{-5}$ | 2 | 1 | AGES-Reykjavik; INTERVAL |
| NAR3 | 0,072 | $1.6\times10^{-5}$ | AF | deCODE | 2 | -0.139; 0.018 | $1.1\times10^{-4}$; $1.0\times10^{-1}$ | 0 | 0 | AGES-Reykjavik; INTERVAL |
| IL6RA | -0,036 | $9.1\times10^{-15}$ | AF | deCODE | 5 | -0.024; -0.071; -0.039; -0.612; -0.03 | $2.8\times10^{-5}$; $1.0\times10^{-100}$; $1.0\times10^{-100}$; $1.1\times10^{-5}$; $2.5\times10^{-14}$ | 5 | 5 | SCALLOP; AGES-Reykjavik; Gilly; Yang; INTERVAL |
| IL6RA | -0,023 | $6.0\times10^{-7}$ | HF | deCODE | 5 | -0.014; -0.009; -0.031; -0.18; -0.016 | $5.1\times10^{-3}$; $6.7\times10^{-2}$; $1.0\times10^{-100}$; $6.0\times10^{-3}$; $7.1\times10^{-3}$ | 4 | 1 | SCALLOP; AGES-Reykjavik; Gilly; Yang; INTERVAL |
| CAN2 | 0,189 | $2.7\times10^{-8}$ | HF | deCODE | 1 | 0.126 | $3.2\times10^{-2}$ | 1 | 0 | AGES-Reykjavik |
| ERAP1 | -0,028 | $3.5\times10^{-11}$ | HF | deCODE | 3 | -0.016; -0.013; -0.017 | $4.3\times10^{-5}$; $1.8\times10^{-7}$; $3.7\times10^{-1}$ | 2 | 2 | AGES-Reykjavik; INTERVAL; Yang |
| RET | -0,058 | $1.0\times10^{-100}$ | AF | deCODE | 2 | -0.043; -0.083 | $1.5\times10^{-2}$; $5.3\times10^{-11}$ | 2 | 1 | AGES-Reykjavik; INTERVAL |
| RET | -0,028 | $2.2\times10^{-10}$ | HF | deCODE | 2 | -0.02; -0.034 | $2.7\times10^{-3}$; $2.2\times10^{-3}$ | 2 | 0 | AGES-Reykjavik; INTERVAL |
| RET | -0,102 | $6.7\times10^{-6}$ | NICM | deCODE | 2 | -0.162; -0.164 | $3.5\times10^{-8}$; $4.1\times10^{-3}$ | 2 | 1 | AGES-Reykjavik; INTERVAL |

**Table S11. Replicates of the proteins associated with cardiac outcomes**

| Protein name | Point estimate* | p-value* | Outcome | Discovery study* | No. other occurences* | Other point estimates* | Other p-values* | Nominal replicates* | Adjusted replicates* | Studies* |
|---|---|---|---|---|---|---|---|---|---|---|
| CC126 | 0,063 | $2.2\times10^{-9}$ | HF | deCODE | 2 | 0.039; 0.106 | $6.1\times10^{-4}$; $3.2\times10^{-8}$ | 2 | 1 | AGES-Reykjavik; INTERVAL |
| CC126 | 0,29 | $8.8\times10^{-14}$ | NICM | deCODE | 2 | 0.049; 0.139 | $3.5\times10^{-1}$; $1.5\times10^{-1}$ | 0 | 0 | AGES-Reykjavik; INTERVAL |
| NUDT9 | -1,298 | $9.6\times10^{-6}$ | DCM | deCODE | 2 | -0.057; 0.392 | $5.4\times10^{-1}$; $9.8\times10^{-2}$ | 0 | 0 | AGES-Reykjavik; INTERVAL |
| ACYP2 | -0,176 | $1.1\times10^{-10}$ | AF | deCODE | 1 | 0.029 | $6.6\times10^{-2}$ | 0 | 0 | AGES-Reykjavik |
| CEL | -0,051 | $6.1\times10^{-7}$ | AF | deCODE | 2 | 0.017; 0.015 | $4.2\times10^{-1}$; $2.5\times10^{-1}$ | 0 | 0 | AGES-Reykjavik; INTERVAL |
| CL12A | -0,014 | $2.0\times10^{-6}$ | HF | deCODE | 2 | -0.023; -0.021 | $1.8\times10^{-4}$; $1.0\times10^{-100}$ | 2 | 2 | AGES-Reykjavik; INTERVAL |
| CL12A | -0,099 | $1.2\times10^{-8}$ | NICM | deCODE | 2 | 0.015; -0.029 | $5.4\times10^{-1}$; $1.2\times10^{-2}$ | 1 | 0 | AGES-Reykjavik; INTERVAL |
| CATF | -0,084 | $8.7\times10^{-10}$ | HF | deCODE | 1 | -0.087 | $1.9\times10^{-4}$ | 1 | 1 | INTERVAL |
| NEC1 | -0,043 | $1.0\times10^{-6}$ | AF | deCODE | 2 | -0.03; -0.021 | $3.3\times10^{-9}$; $3.5\times10^{-4}$ | 2 | 2 | AGES-Reykjavik; INTERVAL |
| NEC1 | -0,027 | $1.1\times10^{-5}$ | HF | deCODE | 2 | -0.02; -0.015 | $1.6\times10^{-3}$; $3.2\times10^{-2}$ | 2 | 0 | AGES-Reykjavik; INTERVAL |
| NQO1 | -0,028 | $1.1\times10^{-9}$ | HF | deCODE | 2 | -0.075; -0.039 | $1.3\times10^{-2}$; $1.1\times10^{-6}$ | 2 | 1 | AGES-Reykjavik; INTERVAL |
| ENTP5 | -0,117 | $5.8\times10^{-9}$ | AF | deCODE | 3 | -0.075; -0.02; -0.031 | $3.2\times10^{-12}$; $2.5\times10^{-1}$; $5.8\times10^{-4}$ | 2 | 1 | AGES-Reykjavik; INTERVAL; Gilly |
| PLXB2 | 0,049 | $1.0\times10^{-100}$ | AF | deCODE | 2 | 0.03; 0.027 | $1.3\times10^{-5}$; $1.3\times10^{-7}$ | 2 | 2 | AGES-Reykjavik; INTERVAL |
| PLXB2 | 0,032 | $1.6\times10^{-7}$ | HF | deCODE | 2 | 0.029; 0.017 | $1.9\times10^{-4}$; $1.9\times10^{-3}$ | 2 | 1 | AGES-Reykjavik; INTERVAL |
| ADH4 | -0,36 | $2.0\times10^{-7}$ | HF | deCODE | 1 | -0.074 | $1.3\times10^{-1}$ | 0 | 0 | AGES-Reykjavik |
| TDGF1 | 0,029 | $6.3\times10^{-9}$ | AF | deCODE | 3 | 0.01; 0.008; 0.007 | $1.2\times10^{-2}$; $9.8\times10^{-2}$; $7.3\times10^{-1}$ | 1 | 0 | AGES-Reykjavik; INTERVAL; Yang |
| TDGF1 | -0,126 | $3.7\times10^{-6}$ | DCM | deCODE | 3 | -0.035; -0.049; 0.245 | $1.6\times10^{-1}$; $2.0\times10^{-1}$; $3.1\times10^{-2}$ | 0 | 0 | AGES-Reykjavik; INTERVAL; Yang |

* Columns: Point estimate - odds ratio of indicated association between protein and outcome in the discovery study, p-value - p-value of indicated association in the discovery study, Discovery study - study with largest sample size,
No. other occurences - number of other studies in which replication was attempted, Other point estimates - odds ratio(s) of indicated association for each study in which replication was attempted,
Other p-values - p-value of indicated association for each study in which replication was attempted, Nominal replicates - number of times association was replicated with a p-value < 0.05,
Adjusted replicates - number of times association was replicated with an adjusted p-value, Studies - studies in which replication was attempted, see Methods section for more details.
Abbreviations: AF = atrial fibrillation, DCM = dilated cardiomyopathy, HF = heart failure, NICM = non-ischemic cardiomyopathy
