## Supplemental Table S12 for "Integrating metabolomics and proteomics to identify novel drug targets for heart failure and atrial fibrillation"

**Table S12. Replicates of the proteins associated with metabolites**

| Protein name | Point estimate* | p-value* | Outcome | Discovery study* | No. other occurences* | Other point estimates* | Other p-values* | Nominal replicates* | Adjusted replicates* | Studies* |
|---|---|---|---|---|---|---|---|---|---|---|
| G6PE | 0,061 | $1.9\times10^{-9}$ | Taurine | deCODE | 2 | 0.036; 0.031 | $5.7\times10^{-8}$; $6.5\times10^{-6}$ | 2 | 2 | AGES-Reykjavik; INTERVAL |
| G6PE | 0,048 | $3.3\times10^{-11}$ | Propionylcarnitine | deCODE | 2 | 0.029; 0.035 | $2.6\times10^{-6}$; $8.8\times10^{-6}$ | 2 | 2 | AGES-Reykjavik; INTERVAL |
| G6PE | 0,042 | $2.0\times10^{-8}$ | Hexadecanoylcarnitine | deCODE | 2 | 0.044; 0.028 | $2.7\times10^{-11}$; $9.0\times10^{-5}$ | 2 | 2 | AGES-Reykjavik; INTERVAL |
| G6PE | 0,058 | $1.0\times10^{-100}$ | Octadecadienoylcarnitine | deCODE | 2 | 0.052; 0.035 | $1.0\times10^{-100}$; $1.4\times10^{-7}$ | 2 | 2 | AGES-Reykjavik; INTERVAL |
| G6PE | 0,091 | $2.2\times10^{-16}$ | PC aa C42:6 | deCODE | 2 | 0.057; 0.083 | $2.5\times10^{-8}$; $2.0\times10^{-12}$ | 2 | 2 | AGES-Reykjavik; INTERVAL |
| G6PE | 0,065 | $5.6\times10^{-11}$ | SM C16:0 | deCODE | 2 | 0.048; 0.024 | $8.8\times10^{-13}$; $8.1\times10^{-4}$ | 2 | 1 | AGES-Reykjavik; INTERVAL |
| PDIA5 | 0,044 | $5.2\times10^{-10}$ | Aspartate | deCODE | 2 | 0.003; 0.017 | $6.1\times10^{-1}$; $1.0\times10^{-1}$ | 0 | 0 | AGES-Reykjavik; INTERVAL |
| PDIA5 | -0,063 | $2.7\times10^{-7}$ | Acetylornithine | deCODE | 2 | -0.012; 0.049 | $1.0\times10^{-4}$; $3.3\times10^{-2}$ | 1 | 1 | AGES-Reykjavik; INTERVAL |
| PDIA5 | -0,064 | $2.3\times10^{-11}$ | PC ae C38:3 | deCODE | 2 | -0.036; -0.074 | $8.6\times10^{-12}$; $1.7\times10^{-6}$ | 2 | 2 | AGES-Reykjavik; INTERVAL |
| GNPTG | -0,137 | $2.9\times10^{-11}$ | PC aa C42:6 | deCODE | 2 | -0.557; -0.21 | $9.3\times10^{-6}$; $3.3\times10^{-10}$ | 2 | 2 | AGES-Reykjavik; INTERVAL |
| GNPTG | -0,079 | $8.8\times10^{-10}$ | SM C16:0 | deCODE | 2 | -0.128; -0.091 | $4.4\times10^{-4}$; $1.1\times10^{-5}$ | 2 | 1 | AGES-Reykjavik; INTERVAL |
| CECR1 | 0,045 | $9.5\times10^{-9}$ | Butyrylcarnitine | deCODE | 2 | 0.031; 0.05 | $4.8\times10^{-9}$; $3.0\times10^{-3}$ | 2 | 1 | AGES-Reykjavik; INTERVAL |
| CECR1 | 0,042 | $3.1\times10^{-7}$ | Octadecadienoylcarnitine | deCODE | 2 | 0.018; 0.028 | $4.9\times10^{-3}$; $2.0\times10^{-1}$ | 1 | 0 | AGES-Reykjavik; INTERVAL |
| CECR1 | 0,075 | $6.0\times10^{-10}$ | LPC a C26:1 | deCODE | 2 | 0.075; 0.111 | $3.9\times10^{-3}$; $1.0\times10^{-100}$ | 2 | 1 | AGES-Reykjavik; INTERVAL |
| CECR1 | 0,067 | $1.1\times10^{-8}$ | PC aa C36:4 | deCODE | 2 | 0.05; 0.079 | $2.3\times10^{-8}$; $1.3\times10^{-11}$ | 2 | 2 | AGES-Reykjavik; INTERVAL |
| CECR1 | 0,062 | $5.0\times10^{-8}$ | PC aa C38:5 | deCODE | 2 | 0.036; 0.052 | $1.2\times10^{-4}$; $7.3\times10^{-6}$ | 2 | 2 | AGES-Reykjavik; INTERVAL |
| CECR1 | 0,053 | $8.8\times10^{-7}$ | PC aa C42:1 | deCODE | 2 | -0.006; -0.062 | $7.5\times10^{-1}$; $5.1\times10^{-2}$ | 0 | 0 | AGES-Reykjavik; INTERVAL |
| CECR1 | 0,08 | $2.0\times10^{-12}$ | PC ae C36:5 | deCODE | 2 | 0.053; 0.03 | $2.2\times10^{-3}$; $3.4\times10^{-1}$ | 1 | 0 | AGES-Reykjavik; INTERVAL |
| CECR1 | 0,071 | $8.0\times10^{-10}$ | PC ae C42:3 | deCODE | 2 | 0.017; 0.051 | $7.7\times10^{-2}$; $1.0\times10^{-5}$ | 1 | 1 | AGES-Reykjavik; INTERVAL |
| TIMP3 | -0,058 | $1.3\times10^{-7}$ | PC aa C42:6 | deCODE | 1 | -0.045 | $2.8\times10^{-5}$ | 1 | 1 | AGES-Reykjavik |
| GRAB | 0,233 | $4.6\times10^{-7}$ | Hexadecanoylcarnitine | deCODE | 1 | -0.017 | $2.6\times10^{-3}$ | 0 | 0 | INTERVAL |
| GRAB | 0,68 | $1.0\times10^{-7}$ | PC ae C36:5 | deCODE | 1 | -0.012 | $4.6\times10^{-1}$ | 0 | 0 | INTERVAL |
| GRAB | 0,415 | $4.2\times10^{-7}$ | PC ae C38:3 | deCODE | 1 | -0.039 | $3.1\times10^{-6}$ | 0 | 0 | INTERVAL |
| GRAB | 0,706 | $3.4\times10^{-8}$ | PC ae C38:4 | deCODE | 1 | -0.03 | $5.3\times10^{-4}$ | 0 | 0 | INTERVAL |
| FAAA | 0,08 | $1.3\times10^{-7}$ | Taurine | deCODE | 2 | 0.066; 0.041 | $1.3\times10^{-8}$; $5.4\times10^{-2}$ | 1 | 1 | AGES-Reykjavik; INTERVAL |
| FAAA | 0,11 | $1.5\times10^{-7}$ | PC ae C38:3 | deCODE | 2 | -0.008; 0.021 | $6.3\times10^{-1}$; $4.4\times10^{-1}$ | 0 | 0 | AGES-Reykjavik; INTERVAL |
| LIRB5 | 0,023 | $4.9\times10^{-8}$ | Asparagine | deCODE | 2 | 0.016; 0.034 | $4.0\times10^{-3}$; $1.3\times10^{-7}$ | 2 | 1 | AGES-Reykjavik; INTERVAL |
| LIRB5 | 0,015 | $6.4\times10^{-10}$ | Tyrosine | deCODE | 2 | 0.019; 0.019 | $6.3\times10^{-4}$; $4.0\times10^{-8}$ | 2 | 1 | AGES-Reykjavik; INTERVAL |
| LIRB5 | 0,042 | $3.8\times10^{-14}$ | PC ae C44:6 | deCODE | 2 | 0.012; 0.052 | $3.3\times10^{-1}$; $5.9\times10^{-7}$ | 1 | 1 | AGES-Reykjavik; INTERVAL |
| TICN2 | -0,117 | $1.6\times10^{-7}$ | Tetradecanoylcarnitine | deCODE | 2 | -0.034; -0.062 | $1.3\times10^{-1}$; $5.4\times10^{-3}$ | 1 | 0 | AGES-Reykjavik; INTERVAL |
| TICN2 | -0,121 | $2.8\times10^{-8}$ | Hexadecanoylcarnitine | deCODE | 2 | -0.027; -0.08 | $1.9\times10^{-1}$; $2.1\times10^{-4}$ | 1 | 0 | AGES-Reykjavik; INTERVAL |
| TICN2 | -0,137 | $1.8\times10^{-8}$ | Octadecanoylcarnitine | deCODE | 2 | 0.135; -0.077 | $1.0\times10^{-1}$; $3.9\times10^{-4}$ | 1 | 0 | AGES-Reykjavik; INTERVAL |
| TICN2 | -0,217 | $1.2\times10^{-11}$ | PC ae C40:5 | deCODE | 2 | -0.001; -0.166 | $9.9\times10^{-1}$; $4.3\times10^{-7}$ | 1 | 1 | AGES-Reykjavik; INTERVAL |
| TICN2 | -0,203 | $1.4\times10^{-9}$ | PC ae C42:3 | deCODE | 2 | -0.19; 0.122 | $4.5\times10^{-10}$; $3.5\times10^{-1}$ | 1 | 1 | AGES-Reykjavik; INTERVAL |
| MINP1 | 0,276 | $8.6\times10^{-11}$ | Butyrylcarnitine | deCODE | 2 | 0.145; -0.164 | $1.8\times10^{-14}$; $2.2\times10^{-2}$ | 1 | 1 | AGES-Reykjavik; INTERVAL |
| MINP1 | -0,435 | $4.0\times10^{-9}$ | PC aa C38:6 | deCODE | 2 | -0.159; -0.091 | $2.5\times10^{-4}$; $2.6\times10^{-2}$ | 2 | 0 | AGES-Reykjavik; INTERVAL |
| CCL8 | 0,027 | $6.3\times10^{-8}$ | Tryptophan | deCODE | 1 | 0.075 | $1.3\times10^{-2}$ | 1 | 0 | AGES-Reykjavik |
| CCL8 | 0,041 | $7.2\times10^{-7}$ | LPC a C28:1 | deCODE | 1 | 0.004 | $8.8\times10^{-1}$ | 0 | 0 | AGES-Reykjavik |
| CCL8 | 0,074 | $5.2\times10^{-8}$ | PC aa C38:5 | deCODE | 1 | 0.01 | $5.3\times10^{-1}$ | 0 | 0 | AGES-Reykjavik |
| CCL8 | 0,068 | $1.9\times10^{-7}$ | PC aa C38:6 | deCODE | 1 | 0.146 | $1.2\times10^{-3}$ | 1 | 0 | AGES-Reykjavik |
| APOC3 | 0,19 | $4.7\times10^{-9}$ | Acetylcarnitine | deCODE | 1 | 0.124 | $1.9\times10^{-4}$ | 1 | 0 | AGES-Reykjavik |
| APOC3 | 0,189 | $2.8\times10^{-7}$ | Hexadecanoylcarnitine | deCODE | 1 | 0.166 | $9.4\times10^{-6}$ | 1 | 1 | AGES-Reykjavik |
| APOC3 | 0,157 | $5.3\times10^{-7}$ | Octadecanoylcarnitine | deCODE | 1 | 0.19 | $1.8\times10^{-8}$ | 1 | 1 | AGES-Reykjavik |
| APOC3 | 0,304 | $5.8\times10^{-9}$ | LPC a C26:1 | deCODE | 1 | 0.429 | $3.2\times10^{-14}$ | 1 | 1 | AGES-Reykjavik |
| APOC3 | -0,236 | $6.4\times10^{-8}$ | LPC a C20:3 | deCODE | 1 | -0.281 | $1.2\times10^{-8}$ | 1 | 1 | AGES-Reykjavik |
| APOC3 | 0,39 | $1.0\times10^{-100}$ | PC aa C38:4 | deCODE | 1 | 0.522 | $1.0\times10^{-100}$ | 1 | 1 | AGES-Reykjavik |
| APOC3 | 0,255 | $3.5\times10^{-7}$ | PC aa C38:5 | deCODE | 1 | 0.44 | $1.0\times10^{-100}$ | 1 | 1 | AGES-Reykjavik |
| APOC3 | 0,438 | $2.2\times10^{-16}$ | PC aa C38:6 | deCODE | 1 | 0.593 | $1.0\times10^{-100}$ | 1 | 1 | AGES-Reykjavik |
| APOC3 | 0,629 | $2.0\times10^{-7}$ | PC ae C40:5 | deCODE | 1 | 0.458 | $8.1\times10^{-12}$ | 1 | 1 | AGES-Reykjavik |
| HAVR1 | 0,039 | $7.6\times10^{-7}$ | Serine | deCODE | 4 | 0.014; -0.004; 0.075; 0.027 | $2.3\times10^{-1}$; $7.7\times10^{-1}$; $5.1\times10^{-3}$; $2.0\times10^{-2}$ | 2 | 0 | SCALLOP; AGES-Reykjavik; INTERVAL; Gilly |
| HAVR1 | 0,041 | $6.8\times10^{-7}$ | Tryptophan | deCODE | 4 | 0.029; -0.001; 0.038; 0.037 | $1.8\times10^{-2}$; $9.4\times10^{-1}$; $3.7\times10^{-3}$; $4.7\times10^{-4}$ | 3 | 0 | SCALLOP; AGES-Reykjavik; INTERVAL; Gilly |
| PLXA1 | -0,093 | $7.1\times10^{-9}$ | Asparagine | deCODE | 1 | -0.146 | $3.2\times10^{-4}$ | 1 | 0 | AGES-Reykjavik |
| PLXA1 | 0,127 | $2.9\times10^{-7}$ | Acetylornithine | deCODE | 1 | 0.033 | $3.3\times10^{-1}$ | 0 | 0 | AGES-Reykjavik |
| PLXA1 | -0,086 | $8.2\times10^{-8}$ | SM C16:0 | deCODE | 1 | -0.145 | $1.7\times10^{-5}$ | 1 | 1 | AGES-Reykjavik |
| IL18R | -0,032 | $8.4\times10^{-9}$ | Tryptophan | deCODE | 2 | 0.051; -0.004 | $1.8\times10^{-2}$; $3.6\times10^{-1}$ | 0 | 0 | AGES-Reykjavik; INTERVAL |
| IL18R | -0,025 | $2.3\times10^{-8}$ | Taurine | deCODE | 2 | -0.033; -0.016 | $8.6\times10^{-5}$; $1.1\times10^{-5}$ | 2 | 2 | AGES-Reykjavik; INTERVAL |

**Table S12. Replicates of the proteins associated with metabolites**

| Protein name | Point estimate* | p-value* | Outcome | Discovery study* | No. other occurences* | Other point estimates* | Other p-values* | Nominal replicates* | Adjusted replicates* | Studies* |
|---|---|---|---|---|---|---|---|---|---|---|
| IL18R | -0,039 | $1.3\times10^{-9}$ | Propionylcarnitine | deCODE | 2 | -0.04; -0.019 | $3.5\times10^{-6}$; $7.9\times10^{-2}$ | 1 | 1 | AGES-Reykjavik; INTERVAL |
| IL18R | -0,031 | $6.0\times10^{-8}$ | Butyrylcarnitine | deCODE | 2 | -0.053; -0.025 | $2.4\times10^{-11}$; $1.7\times10^{-3}$ | 2 | 1 | AGES-Reykjavik; INTERVAL |
| IL18R | -0,05 | $2.1\times10^{-9}$ | LPC a C26:1 | deCODE | 2 | -0.058; -0.06 | $6.4\times10^{-4}$; $8.8\times10^{-9}$ | 2 | 1 | AGES-Reykjavik; INTERVAL |
| TIG1 | 0,029 | $7.4\times10^{-12}$ | PC ae C38:4 | deCODE | 2 | -0.003; -0.001 | $4.7\times10^{-1}$; $9.5\times10^{-1}$ | 0 | 0 | AGES-Reykjavik; INTERVAL |
| TIG1 | 0,037 | $1.0\times10^{-100}$ | PC ae C38:5 | deCODE | 2 | 0.009; 0.013 | $2.6\times10^{-1}$; $1.3\times10^{-1}$ | 0 | 0 | AGES-Reykjavik; INTERVAL |
| TIG1 | 0,031 | $4.6\times10^{-14}$ | SM C16:0 | deCODE | 2 | 0.006; 0.015 | $1.4\times10^{-1}$; $1.0\times10^{-4}$ | 1 | 1 | AGES-Reykjavik; INTERVAL |
| FA10 | 0,25 | $4.4\times10^{-11}$ | PC aa C38:5 | deCODE | 2 | 0.108; 0.071 | $1.3\times10^{-2}$; $2.0\times10^{-5}$ | 2 | 1 | AGES-Reykjavik; INTERVAL |
| FA10 | 0,217 | $1.0\times10^{-8}$ | PC aa C40:4 | deCODE | 2 | 0.115; 0.043 | $3.1\times10^{-3}$; $3.3\times10^{-2}$ | 2 | 0 | AGES-Reykjavik; INTERVAL |
| FA10 | 0,216 | $2.2\times10^{-7}$ | PC aa C42:6 | deCODE | 2 | 0.079; 0.001 | $7.7\times10^{-2}$; $9.7\times10^{-1}$ | 0 | 0 | AGES-Reykjavik; INTERVAL |
| FA10 | 0,204 | $4.2\times10^{-7}$ | PC ae C38:4 | deCODE | 2 | 0.11; 0.106 | $2.1\times10^{-2}$; $2.7\times10^{-9}$ | 2 | 1 | AGES-Reykjavik; INTERVAL |
| FA10 | 0,187 | $8.6\times10^{-7}$ | PC ae C42:2 | deCODE | 2 | 0.106; 0.061 | $1.5\times10^{-2}$; $2.7\times10^{-4}$ | 2 | 0 | AGES-Reykjavik; INTERVAL |
| IGFR1 | -0,043 | $5.8\times10^{-7}$ | Acetylornithine | deCODE | 2 | -0.053; 0.047 | $4.5\times10^{-2}$; $1.7\times10^{-1}$ | 1 | 0 | AGES-Reykjavik; INTERVAL |
| IGFR1 | -0,061 | $6.5\times10^{-8}$ | PC aa C36:4 | deCODE | 2 | 0.027; -0.039 | $5.6\times10^{-1}$; $7.6\times10^{-2}$ | 0 | 0 | AGES-Reykjavik; INTERVAL |
| ISK2 | 0,115 | $3.1\times10^{-7}$ | PC ae C40:5 | deCODE | 2 | 0.064; 0.239 | $1.9\times10^{-3}$; $4.7\times10^{-3}$ | 2 | 0 | AGES-Reykjavik; INTERVAL |
| ISK2 | 0,135 | $6.5\times10^{-10}$ | PC ae C42:2 | deCODE | 2 | 0.078; 0.087 | $2.5\times10^{-4}$; $2.3\times10^{-5}$ | 2 | 1 | AGES-Reykjavik; INTERVAL |
| AT1B2 | 0,065 | $1.6\times10^{-7}$ | Tryptophan | deCODE | 2 | 0.016; 0.054 | $2.9\times10^{-1}$; $3.9\times10^{-5}$ | 1 | 1 | AGES-Reykjavik; INTERVAL |
| AT1B2 | 0,036 | $1.3\times10^{-8}$ | Tyrosine | deCODE | 2 | 0.049; 0.052 | $6.0\times10^{-6}$; $2.0\times10^{-13}$ | 2 | 2 | AGES-Reykjavik; INTERVAL |
| AT1B2 | -0,109 | $3.3\times10^{-7}$ | Taurine | deCODE | 2 | -0.018; -0.18 | $2.7\times10^{-1}$; $5.4\times10^{-5}$ | 1 | 1 | AGES-Reykjavik; INTERVAL |
| AT1B2 | -0,068 | $9.3\times10^{-8}$ | Hexadecanoylcarnitine | deCODE | 2 | -0.055; -0.069 | $3.8\times10^{-3}$; $2.7\times10^{-5}$ | 2 | 1 | AGES-Reykjavik; INTERVAL |
| AT1B2 | -0,076 | $7.1\times10^{-7}$ | Octadecadienoylcarnitine | deCODE | 2 | -0.029; 0.173 | $5.4\times10^{-2}$; $1.5\times10^{-3}$ | 0 | 0 | AGES-Reykjavik; INTERVAL |
| C1QRF | -0,064 | $6.5\times10^{-10}$ | Serine | deCODE | 1 | -0.018 | $5.0\times10^{-2}$ | 1 | 0 | AGES-Reykjavik |
| C1QRF | -0,07 | $2.2\times10^{-10}$ | Tetradecanoylcarnitine | deCODE | 1 | -0.04 | $5.6\times10^{-5}$ | 1 | 1 | AGES-Reykjavik |
| C1QRF | -0,077 | $1.2\times10^{-8}$ | Octadecadienoylcarnitine | deCODE | 1 | -0.088 | $5.2\times10^{-3}$ | 1 | 0 | AGES-Reykjavik |
| C1QRF | 0,073 | $1.1\times10^{-7}$ | PC aa C36:4 | deCODE | 1 | 0.107 | $2.2\times10^{-15}$ | 1 | 1 | AGES-Reykjavik |
| C1QRF | 0,079 | $7.4\times10^{-8}$ | PC aa C38:4 | deCODE | 1 | 0.203 | $7.8\times10^{-5}$ | 1 | 1 | AGES-Reykjavik |
| C1QRF | 0,073 | $2.0\times10^{-7}$ | PC aa C38:6 | deCODE | 1 | 0.061 | $3.8\times10^{-6}$ | 1 | 1 | AGES-Reykjavik |
| C1QRF | 0,067 | $6.3\times10^{-9}$ | SM C16:0 | deCODE | 1 | 0.105 | $1.4\times10^{-4}$ | 1 | 0 | AGES-Reykjavik |
| SWP70 | -0,042 | $4.1\times10^{-7}$ | Propionylcarnitine | deCODE | 2 | -0.027; -0.031 | $6.2\times10^{-3}$; $8.3\times10^{-2}$ | 1 | 0 | AGES-Reykjavik; INTERVAL |
| SWP70 | -0,059 | $2.2\times10^{-7}$ | PC aa C38:5 | deCODE | 2 | -0.172; -0.048 | $3.8\times10^{-4}$; $6.4\times10^{-2}$ | 1 | 0 | AGES-Reykjavik; INTERVAL |
| SWP70 | -0,069 | $1.5\times10^{-9}$ | PC aa C38:6 | deCODE | 2 | -0.081; -0.024 | $1.7\times10^{-8}$; $3.5\times10^{-1}$ | 1 | 1 | AGES-Reykjavik; INTERVAL |
| SWP70 | -0,103 | $4.2\times10^{-7}$ | PC ae C42:2 | deCODE | 2 | -0.064; -0.033 | $6.2\times10^{-5}$; $2.1\times10^{-1}$ | 1 | 1 | AGES-Reykjavik; INTERVAL |
| GSTM3 | 0,028 | $2.5\times10^{-14}$ | Tyrosine | deCODE | 1 | 0.059 | $4.3\times10^{-5}$ | 1 | 1 | AGES-Reykjavik |
| GSTM3 | -0,031 | $3.1\times10^{-8}$ | Acetylornithine | deCODE | 1 | -0.051 | $2.3\times10^{-2}$ | 1 | 0 | AGES-Reykjavik |
| GSTM3 | -0,047 | $2.6\times10^{-14}$ | Acetylcarnitine | deCODE | 1 | -0.14 | $1.0\times10^{-10}$ | 1 | 1 | AGES-Reykjavik |
| GSTM3 | -0,07 | $1.6\times10^{-13}$ | Propionylcarnitine | deCODE | 1 | -0.096 | $2.7\times10^{-5}$ | 1 | 1 | AGES-Reykjavik |
| GSTM3 | 0,03 | $3.9\times10^{-7}$ | SM C16:0 | deCODE | 1 | 0.087 | $5.8\times10^{-5}$ | 1 | 1 | AGES-Reykjavik |
| EDAR | -0,153 | $4.3\times10^{-8}$ | LPC a C26:1 | deCODE | 2 | -0.093; -0.058 | $8.2\times10^{-4}$; $2.6\times10^{-2}$ | 2 | 0 | AGES-Reykjavik; INTERVAL |
| EDAR | -0,171 | $5.8\times10^{-12}$ | LPC a C20:3 | deCODE | 2 | -0.074; -0.104 | $1.3\times10^{-2}$; $9.3\times10^{-5}$ | 2 | 1 | AGES-Reykjavik; INTERVAL |
| EDAR | -0,132 | $4.0\times10^{-8}$ | LPC a C28:1 | deCODE | 2 | -0.054; -0.06 | $9.8\times10^{-2}$; $2.5\times10^{-2}$ | 1 | 0 | AGES-Reykjavik; INTERVAL |
| EDAR | -0,175 | $1.0\times10^{-9}$ | PC aa C36:4 | deCODE | 2 | -0.088; -0.099 | $9.7\times10^{-4}$; $2.2\times10^{-3}$ | 2 | 0 | AGES-Reykjavik; INTERVAL |
| EDAR | -0,212 | $1.3\times10^{-7}$ | PC aa C38:5 | deCODE | 2 | -0.094; -0.12 | $4.2\times10^{-4}$; $6.4\times10^{-6}$ | 2 | 1 | AGES-Reykjavik; INTERVAL |
| EDAR | -0,175 | $3.3\times10^{-12}$ | PC ae C38:4 | deCODE | 2 | -0.099; -0.127 | $1.0\times10^{-3}$; $1.4\times10^{-5}$ | 2 | 1 | AGES-Reykjavik; INTERVAL |
| EDAR | -0,246 | $2.0\times10^{-9}$ | PC ae C38:5 | deCODE | 2 | 0.015; -0.107 | $7.9\times10^{-1}$; $1.2\times10^{-3}$ | 1 | 0 | AGES-Reykjavik; INTERVAL |
| EDAR | -0,223 | $3.2\times10^{-8}$ | PC ae C40:5 | deCODE | 2 | -0.096; -0.104 | $6.6\times10^{-4}$; $2.0\times10^{-3}$ | 2 | 0 | AGES-Reykjavik; INTERVAL |
| EDAR | -0,129 | $6.9\times10^{-8}$ | PC ae C42:3 | deCODE | 2 | -0.057; -0.101 | $6.2\times10^{-2}$; $3.2\times10^{-3}$ | 1 | 0 | AGES-Reykjavik; INTERVAL |
| EDAR | -0,121 | $4.4\times10^{-7}$ | SM C16:1 | deCODE | 2 | -0.098; -0.093 | $2.3\times10^{-4}$; $1.2\times10^{-3}$ | 2 | 0 | AGES-Reykjavik; INTERVAL |
| GLCE | -0,093 | $7.3\times10^{-10}$ | PC aa C36:4 | deCODE | 2 | -0.039; -0.045 | $1.0\times10^{-4}$; $6.8\times10^{-6}$ | 2 | 2 | AGES-Reykjavik; INTERVAL |
| GLCE | 0,069 | $5.2\times10^{-9}$ | PC aa C42:6 | deCODE | 2 | -0.036; -0.018 | $1.3\times10^{-1}$; $4.5\times10^{-1}$ | 0 | 0 | AGES-Reykjavik; INTERVAL |
| GLCE | -0,049 | $4.2\times10^{-7}$ | SM C16:0 | deCODE | 2 | -0.021; 0.028 | $8.0\times10^{-5}$; $5.5\times10^{-2}$ | 1 | 1 | AGES-Reykjavik; INTERVAL |
| PVRL4 | 0,427 | $7.7\times10^{-11}$ | Taurine | deCODE | 1 | 0.942 | $1.6\times10^{-3}$ | 1 | 0 | AGES-Reykjavik |
| FCG2A | -0,032 | $1.5\times10^{-10}$ | Octadecanoylcarnitine | deCODE | 2 | -0.001; -0.008 | $8.5\times10^{-1}$; $1.8\times10^{-2}$ | 1 | 0 | AGES-Reykjavik; INTERVAL |
| FCG2A | -0,025 | $4.5\times10^{-7}$ | Octadecadienoylcarnitine | deCODE | 2 | -0.009; -0.014 | $5.4\times10^{-3}$; $4.2\times10^{-6}$ | 2 | 1 | AGES-Reykjavik; INTERVAL |
| FCG2A | -0,025 | $8.3\times10^{-9}$ | PC aa C40:4 | deCODE | 2 | -0.005; -0.0 | $3.1\times10^{-1}$; $9.6\times10^{-1}$ | 0 | 0 | AGES-Reykjavik; INTERVAL |
| REG3G | -0,057 | $6.2\times10^{-7}$ | PC aa C36:4 | deCODE | 1 | -0.037 | $6.0\times10^{-5}$ | 1 | 1 | AGES-Reykjavik |
| REG3G | -0,06 | $1.8\times10^{-7}$ | PC aa C38:4 | deCODE | 1 | 0.019 | $4.0\times10^{-1}$ | 0 | 0 | AGES-Reykjavik |
| REG3G | -0,071 | $1.1\times10^{-9}$ | PC aa C38:5 | deCODE | 1 | -0.035 | $1.1\times10^{-1}$ | 0 | 0 | AGES-Reykjavik |

**Table S12. Replicates of the proteins associated with metabolites**

| Protein name | Point estimate* | p-value* | Outcome | Discovery study* | No. other occurences* | Other point estimates* | Other p-values* | Nominal replicates* | Adjusted replicates* | Studies* |
|---|---|---|---|---|---|---|---|---|---|---|
| REG3G | -0,076 | $1.6\times10^{-10}$ | PC aa C38:6 | deCODE | 1 | -0.047 | $9.9\times10^{-7}$ | 1 | 1 | AGES-Reykjavik |
| REG3G | -0,05 | $1.3\times10^{-9}$ | PC ae C38:3 | deCODE | 1 | -0.06 | $8.5\times10^{-13}$ | 1 | 1 | AGES-Reykjavik |
| REG3G | -0,055 | $2.9\times10^{-11}$ | PC ae C38:5 | deCODE | 1 | -0.041 | $5.3\times10^{-2}$ | 0 | 0 | AGES-Reykjavik |
| I17RD | 0,081 | $6.1\times10^{-7}$ | Aspartate | deCODE | 2 | 0.021; 0.062 | $3.2\times10^{-2}$; $5.3\times10^{-4}$ | 2 | 0 | AGES-Reykjavik; INTERVAL |
| I17RD | 0,075 | $8.5\times10^{-8}$ | Propionylcarnitine | deCODE | 2 | 0.066; 0.023 | $1.6\times10^{-2}$; $2.0\times10^{-1}$ | 1 | 0 | AGES-Reykjavik; INTERVAL |
| I17RD | -0,053 | $7.2\times10^{-9}$ | Octadecanoylcarnitine | deCODE | 2 | -0.104; -0.035 | $1.0\times10^{-100}$; $5.9\times10^{-4}$ | 2 | 1 | AGES-Reykjavik; INTERVAL |
| I17RD | -0,068 | $1.5\times10^{-10}$ | LPC a C20:4 | deCODE | 2 | -0.013; -0.058 | $7.6\times10^{-1}$; $3.5\times10^{-2}$ | 1 | 0 | AGES-Reykjavik; INTERVAL |
| I17RD | -0,081 | $6.0\times10^{-13}$ | LPC a C20:3 | deCODE | 2 | -0.063; -0.05 | $2.0\times10^{-1}$; $7.1\times10^{-4}$ | 1 | 0 | AGES-Reykjavik; INTERVAL |
| GXLT1 | 0,122 | $4.0\times10^{-15}$ | Octadecadienoylcarnitine | deCODE | 2 | 0.052; 0.04 | $9.1\times10^{-5}$; $3.5\times10^{-2}$ | 2 | 1 | AGES-Reykjavik; INTERVAL |
| GFRA1 | -0,104 | $1.9\times10^{-7}$ | Hexadecanoylcarnitine | deCODE | 2 | -0.063; 0.003 | $1.3\times10^{-5}$; $8.5\times10^{-1}$ | 1 | 1 | AGES-Reykjavik; INTERVAL |
| GFRA1 | -0,107 | $1.1\times10^{-7}$ | Octadecanoylcarnitine | deCODE | 2 | -0.07; -0.041 | $3.5\times10^{-5}$; $2.8\times10^{-2}$ | 2 | 1 | AGES-Reykjavik; INTERVAL |
| GFRA1 | -0,157 | $1.2\times10^{-7}$ | PC ae C38:4 | deCODE | 2 | -0.038; 0.004 | $5.9\times10^{-2}$; $9.1\times10^{-1}$ | 0 | 0 | AGES-Reykjavik; INTERVAL |
| PLA2R | 0,012 | $2.5\times10^{-9}$ | Tryptophan | deCODE | 2 | 0.009; 0.002 | $4.6\times10^{-2}$; $5.8\times10^{-1}$ | 1 | 0 | AGES-Reykjavik; INTERVAL |
| PLA2R | 0,022 | $1.0\times10^{-100}$ | Tyrosine | deCODE | 2 | 0.048; 0.023 | $8.9\times10^{-16}$; $1.0\times10^{-100}$ | 2 | 2 | AGES-Reykjavik; INTERVAL |
| PLA2R | -0,011 | $1.6\times10^{-7}$ | Propionylcarnitine | deCODE | 2 | -0.038; -0.005 | $6.2\times10^{-4}$; $4.8\times10^{-2}$ | 2 | 0 | AGES-Reykjavik; INTERVAL |
| PLA2R | -0,017 | $1.4\times10^{-11}$ | Tetradecanoylcarnitine | deCODE | 2 | -0.037; -0.02 | $1.5\times10^{-3}$; $1.3\times10^{-14}$ | 2 | 1 | AGES-Reykjavik; INTERVAL |
| PLA2R | -0,016 | $4.9\times10^{-12}$ | Octadecanoylcarnitine | deCODE | 2 | -0.023; -0.018 | $7.7\times10^{-2}$; $5.0\times10^{-10}$ | 1 | 1 | AGES-Reykjavik; INTERVAL |
| PLA2R | -0,053 | $2.8\times10^{-8}$ | LPC a C28:1 | deCODE | 2 | -0.024; 0.002 | $8.7\times10^{-4}$; $6.1\times10^{-1}$ | 1 | 0 | AGES-Reykjavik; INTERVAL |
| NAR3 | 0,063 | $2.3\times10^{-10}$ | Aspartate | deCODE | 2 | 0.013; 0.007 | $5.4\times10^{-1}$; $5.8\times10^{-1}$ | 0 | 0 | AGES-Reykjavik; INTERVAL |
| NAR3 | 0,081 | $9.5\times10^{-9}$ | LPC a C28:1 | deCODE | 2 | 0.018; 0.078 | $5.1\times10^{-1}$; $1.6\times10^{-4}$ | 1 | 0 | AGES-Reykjavik; INTERVAL |
| NAR3 | -0,091 | $9.2\times10^{-9}$ | PC aa C40:4 | deCODE | 2 | -0.048; -0.036 | $4.1\times10^{-1}$; $7.1\times10^{-2}$ | 0 | 0 | AGES-Reykjavik; INTERVAL |
| NAR3 | 0,136 | $4.2\times10^{-9}$ | PC aa C42:1 | deCODE | 2 | 0.075; 0.061 | $9.4\times10^{-3}$; $5.3\times10^{-3}$ | 2 | 0 | AGES-Reykjavik; INTERVAL |
| NAR3 | -0,078 | $2.2\times10^{-7}$ | PC ae C40:5 | deCODE | 2 | -0.099; -0.102 | $2.7\times10^{-4}$; $3.5\times10^{-7}$ | 2 | 1 | AGES-Reykjavik; INTERVAL |
| IL6RA | -0,03 | $2.2\times10^{-11}$ | SM C16:0 | deCODE | 5 | -0.032; -0.022; -0.014; 0.004; -0.014 | $5.1\times10^{-6}$; $4.7\times10^{-10}$; $5.8\times10^{-4}$; $9.6\times10^{-1}$; $2.6\times10^{-3}$ | 4 | 2 | SCALLOP; AGES-Reykjavik; Gilly; Yang; INTERVAL |
| CAN2 | 0,109 | $7.8\times10^{-10}$ | Tyrosine | deCODE | 1 | 0.071 | $6.4\times10^{-2}$ | 0 | 0 | AGES-Reykjavik |
| CAN2 | 0,194 | $6.5\times10^{-10}$ | Propionylcarnitine | deCODE | 2 | 0.091; 0.071 | $1.5\times10^{-1}$; $8.2\times10^{-2}$ | 0 | 0 | AGES-Reykjavik; INTERVAL |
| CAN2 | 0,234 | $5.0\times10^{-7}$ | PC ae C38:5 | deCODE | 1 | 0.109 | $4.1\times10^{-1}$ | 0 | 0 | AGES-Reykjavik |
| ERAP1 | -0,029 | $8.8\times10^{-10}$ | Propionylcarnitine | deCODE | 3 | -0.014; -0.012; -0.063 | $3.3\times10^{-3}$; $1.0\times10^{-5}$; $2.1\times10^{-3}$ | 3 | 1 | AGES-Reykjavik; INTERVAL; Yang |
| ERAP1 | -0,026 | $1.6\times10^{-7}$ | Octadecadienoylcarnitine | deCODE | 3 | -0.004; -0.007; 0.148 | $3.3\times10^{-2}$; $8.0\times10^{-3}$; $4.2\times10^{-4}$ | 2 | 0 | AGES-Reykjavik; INTERVAL; Yang |
| ERAP1 | -0,031 | $7.2\times10^{-7}$ | PC aa C38:5 | deCODE | 3 | -0.049; -0.04; -0.075 | $5.8\times10^{-12}$; $1.0\times10^{-100}$; $2.1\times10^{-1}$ | 2 | 2 | AGES-Reykjavik; INTERVAL; Yang |
| ERAP1 | -0,035 | $4.9\times10^{-8}$ | PC aa C38:6 | deCODE | 3 | -0.051; -0.05; -0.059 | $6.3\times10^{-12}$; $1.0\times10^{-100}$; $3.3\times10^{-1}$ | 2 | 2 | AGES-Reykjavik; INTERVAL; Yang |
| RET | -0,037 | $7.5\times10^{-10}$ | Asparagine | deCODE | 2 | 0.053; 0.02 | $2.7\times10^{-3}$; $2.0\times10^{-1}$ | 0 | 0 | AGES-Reykjavik; INTERVAL |
| RET | 0,034 | $1.5\times10^{-12}$ | Aspartate | deCODE | 2 | 0.005; -0.011 | $8.0\times10^{-1}$; $4.2\times10^{-1}$ | 0 | 0 | AGES-Reykjavik; INTERVAL |
| RET | -0,04 | $4.2\times10^{-9}$ | Taurine | deCODE | 2 | -0.078; -0.021 | $4.7\times10^{-5}$; $1.0\times10^{-1}$ | 1 | 1 | AGES-Reykjavik; INTERVAL |
| RET | -0,03 | $7.0\times10^{-10}$ | Acetylcarnitine | deCODE | 2 | -0.004; -0.039 | $5.7\times10^{-1}$; $6.2\times10^{-3}$ | 1 | 0 | AGES-Reykjavik; INTERVAL |
| RET | -0,041 | $1.0\times10^{-100}$ | Propionylcarnitine | deCODE | 2 | 0.038; -0.032 | $3.6\times10^{-2}$; $2.3\times10^{-2}$ | 1 | 0 | AGES-Reykjavik; INTERVAL |
| RET | -0,093 | $2.9\times10^{-7}$ | PC aa C36:4 | deCODE | 2 | -0.045; -0.065 | $4.6\times10^{-5}$; $6.0\times10^{-5}$ | 2 | 2 | AGES-Reykjavik; INTERVAL |
| RET | -0,076 | $1.0\times10^{-100}$ | PC aa C40:4 | deCODE | 2 | -0.069; -0.112 | $9.2\times10^{-9}$; $5.4\times10^{-12}$ | 2 | 2 | AGES-Reykjavik; INTERVAL |
| RET | -0,105 | $4.6\times10^{-7}$ | PC aa C42:1 | deCODE | 2 | -0.028; -0.03 | $1.3\times10^{-2}$; $7.0\times10^{-2}$ | 1 | 0 | AGES-Reykjavik; INTERVAL |
| RET | -0,07 | $1.0\times10^{-100}$ | PC ae C38:3 | deCODE | 2 | -0.034; -0.078 | $4.5\times10^{-3}$; $1.5\times10^{-6}$ | 2 | 1 | AGES-Reykjavik; INTERVAL |
| RET | -0,044 | $3.7\times10^{-7}$ | PC ae C38:5 | deCODE | 2 | -0.019; -0.124 | $7.7\times10^{-2}$; $4.5\times10^{-2}$ | 1 | 0 | AGES-Reykjavik; INTERVAL |
| CC126 | 0,04 | $2.6\times10^{-8}$ | Tyrosine | deCODE | 2 | 0.042; -0.014 | $6.3\times10^{-9}$; $4.0\times10^{-1}$ | 1 | 1 | AGES-Reykjavik; INTERVAL |
| CC126 | -0,103 | $3.0\times10^{-13}$ | Acetylornithine | deCODE | 2 | -0.111; -0.047 | $5.4\times10^{-10}$; $1.8\times10^{-3}$ | 2 | 1 | AGES-Reykjavik; INTERVAL |
| CC126 | 0,061 | $1.2\times10^{-10}$ | Octadecanoylcarnitine | deCODE | 2 | 0.007; -0.008 | $6.2\times10^{-1}$; $6.5\times10^{-1}$ | 0 | 0 | AGES-Reykjavik; INTERVAL |
| CC126 | 0,115 | $1.1\times10^{-7}$ | PC aa C42:6 | deCODE | 2 | 0.102; 0.029 | $1.7\times10^{-9}$; $2.8\times10^{-1}$ | 1 | 1 | AGES-Reykjavik; INTERVAL |
| NUDT9 | -0,186 | $1.7\times10^{-14}$ | Acetylcarnitine | deCODE | 2 | -0.073; -0.073 | $5.1\times10^{-5}$; $6.1\times10^{-5}$ | 2 | 2 | AGES-Reykjavik; INTERVAL |
| NUDT9 | -0,14 | $4.9\times10^{-7}$ | Hexadecanoylcarnitine | deCODE | 2 | -0.087; -0.052 | $1.5\times10^{-5}$; $3.5\times10^{-3}$ | 2 | 1 | AGES-Reykjavik; INTERVAL |
| NUDT9 | -0,261 | $6.2\times10^{-7}$ | PC aa C38:5 | deCODE | 2 | -0.109; -0.089 | $3.0\times10^{-4}$; $2.2\times10^{-2}$ | 2 | 0 | AGES-Reykjavik; INTERVAL |
| ACYP2 | -0,104 | $6.0\times10^{-12}$ | Aspartate | deCODE | 2 | 0.05; -0.024 | $4.8\times10^{-3}$; $3.6\times10^{-1}$ | 0 | 0 | AGES-Reykjavik; INTERVAL |
| ACYP2 | 0,074 | $2.2\times10^{-7}$ | Serine | deCODE | 2 | -0.049; -0.073 | $1.5\times10^{-1}$; $8.2\times10^{-3}$ | 0 | 0 | AGES-Reykjavik; INTERVAL |
| ACYP2 | 0,087 | $5.0\times10^{-9}$ | Tryptophan | deCODE | 2 | -0.055; -0.014 | $2.4\times10^{-3}$; $5.8\times10^{-1}$ | 0 | 0 | AGES-Reykjavik; INTERVAL |
| ACYP2 | 0,065 | $8.0\times10^{-7}$ | Propionylcarnitine | deCODE | 2 | 0.008; -0.019 | $6.2\times10^{-1}$; $5.8\times10^{-1}$ | 0 | 0 | AGES-Reykjavik; INTERVAL |
| ACYP2 | 0,107 | $1.2\times10^{-9}$ | Hexadecanoylcarnitine | deCODE | 2 | -0.044; -0.048 | $6.2\times10^{-3}$; $7.1\times10^{-2}$ | 0 | 0 | AGES-Reykjavik; INTERVAL |
| ACYP2 | 0,262 | $1.0\times10^{-100}$ | Octadecanoylcarnitine | deCODE | 2 | -0.162; -0.093 | $1.5\times10^{-5}$; $8.8\times10^{-3}$ | 0 | 0 | AGES-Reykjavik; INTERVAL |
| ACYP2 | 0,098 | $5.5\times10^{-7}$ | LPC a C28:1 | deCODE | 2 | -0.003; -0.016 | $9.1\times10^{-1}$; $7.0\times10^{-1}$ | 0 | 0 | AGES-Reykjavik; INTERVAL |
| CEL | 0,074 | $8.7\times10^{-10}$ | Acetylornithine | deCODE | 2 | 0.021; 0.014 | $4.2\times10^{-1}$; $3.9\times10^{-1}$ | 0 | 0 | AGES-Reykjavik; INTERVAL |

**Table S12. Replicates of the proteins associated with metabolites**

| Protein name | Point estimate* | p-value* | Outcome | Discovery study* | No. other occurences* | Other point estimates* | Other p-values* | Nominal replicates* | Adjusted replicates* | Studies* |
|---|---|---|---|---|---|---|---|---|---|---|
| CL12A | -0,023 | $8.9\times10^{-8}$ | Asparagine | deCODE | 2 | -0.027; -0.027 | $2.1\times10^{-5}$; $3.6\times10^{-4}$ | 2 | 1 | AGES-Reykjavik; INTERVAL |
| CL12A | -0,019 | $3.7\times10^{-10}$ | Tryptophan | deCODE | 2 | -0.027; -0.022 | $1.9\times10^{-5}$; $5.5\times10^{-13}$ | 2 | 2 | AGES-Reykjavik; INTERVAL |
| CL12A | 0,032 | $8.9\times10^{-8}$ | PC aa C38:5 | deCODE | 2 | 0.009; 0.006 | $4.1\times10^{-1}$; $2.6\times10^{-1}$ | 0 | 0 | AGES-Reykjavik; INTERVAL |
| CL12A | 0,032 | $3.0\times10^{-7}$ | PC ae C42:2 | deCODE | 2 | -0.008; -0.003 | $4.9\times10^{-1}$; $7.3\times10^{-1}$ | 0 | 0 | AGES-Reykjavik; INTERVAL |
| CATF | -0,087 | $6.2\times10^{-8}$ | Serine | deCODE | 1 | -0.075 | $4.1\times10^{-4}$ | 1 | 0 | INTERVAL |
| CATF | -0,139 | $2.6\times10^{-12}$ | Tyrosine | deCODE | 1 | -0.033 | $1.4\times10^{-2}$ | 1 | 0 | INTERVAL |
| CATF | -0,087 | $6.0\times10^{-10}$ | Octadecadienoylcarnitine | deCODE | 1 | -0.049 | $2.1\times10^{-2}$ | 1 | 0 | INTERVAL |
| CATF | 0,134 | $9.1\times10^{-8}$ | PC aa C36:4 | deCODE | 1 | 0.075 | $2.2\times10^{-2}$ | 1 | 0 | INTERVAL |
| CATF | 0,14 | $2.8\times10^{-8}$ | PC aa C38:5 | deCODE | 1 | 0.025 | $4.7\times10^{-1}$ | 0 | 0 | INTERVAL |
| CATF | 0,204 | $4.4\times10^{-16}$ | PC aa C38:6 | deCODE | 1 | 0.062 | $5.9\times10^{-2}$ | 0 | 0 | INTERVAL |
| CATF | 0,211 | $3.9\times10^{-11}$ | PC ae C36:5 | deCODE | 1 | 0.083 | $3.8\times10^{-2}$ | 1 | 0 | INTERVAL |
| CATF | 0,125 | $7.0\times10^{-7}$ | PC ae C38:5 | deCODE | 1 | 0.031 | $3.5\times10^{-1}$ | 0 | 0 | INTERVAL |
| NEC1 | 0,039 | $1.4\times10^{-7}$ | Taurine | deCODE | 2 | 0.033; -0.004 | $2.5\times10^{-2}$; $6.1\times10^{-1}$ | 1 | 0 | AGES-Reykjavik; INTERVAL |
| NEC1 | -0,067 | $2.8\times10^{-10}$ | PC aa C36:4 | deCODE | 2 | -0.002; -0.013 | $9.2\times10^{-1}$; $2.6\times10^{-1}$ | 0 | 0 | AGES-Reykjavik; INTERVAL |
| NEC1 | -0,068 | $9.1\times10^{-11}$ | PC ae C36:5 | deCODE | 2 | -0.013; -0.024 | $5.9\times10^{-1}$; $1.8\times10^{-2}$ | 1 | 0 | AGES-Reykjavik; INTERVAL |
| NEC1 | -0,06 | $6.4\times10^{-9}$ | PC ae C38:4 | deCODE | 2 | -0.025; -0.006 | $3.3\times10^{-1}$; $6.1\times10^{-1}$ | 0 | 0 | AGES-Reykjavik; INTERVAL |
| NEC1 | -0,053 | $1.5\times10^{-7}$ | SM C16:1 | deCODE | 2 | -0.011; -0.035 | $6.4\times10^{-1}$; $3.0\times10^{-3}$ | 1 | 0 | AGES-Reykjavik; INTERVAL |
| NQO1 | 0,022 | $9.2\times10^{-9}$ | Taurine | deCODE | 2 | 0.057; 0.002 | $2.2\times10^{-2}$; $9.0\times10^{-1}$ | 1 | 0 | AGES-Reykjavik; INTERVAL |
| NQO1 | -0,033 | $4.4\times10^{-16}$ | Butyrylcarnitine | deCODE | 2 | -0.029; -0.035 | $2.4\times10^{-1}$; $6.1\times10^{-2}$ | 0 | 0 | AGES-Reykjavik; INTERVAL |
| NQO1 | 0,025 | $1.1\times10^{-9}$ | Octadecadienoylcarnitine | deCODE | 2 | 0.088; 0.012 | $1.4\times10^{-3}$; $6.3\times10^{-3}$ | 2 | 0 | AGES-Reykjavik; INTERVAL |
| NQO1 | -0,028 | $1.3\times10^{-7}$ | SM C16:0 | deCODE | 2 | 0.007; -0.036 | $7.8\times10^{-1}$; $2.6\times10^{-11}$ | 1 | 1 | AGES-Reykjavik; INTERVAL |
| ENTP5 | -0,125 | $2.1\times10^{-10}$ | Tetradecanoylcarnitine | deCODE | 3 | -0.085; -0.08; -0.032 | $3.8\times10^{-3}$; $1.3\times10^{-3}$; $5.9\times10^{-3}$ | 3 | 0 | AGES-Reykjavik; INTERVAL; Gilly |
| ENTP5 | -0,119 | $1.8\times10^{-8}$ | Hexadecanoylcarnitine | deCODE | 3 | -0.093; -0.087; -0.031 | $2.6\times10^{-10}$; $2.6\times10^{-4}$; $2.2\times10^{-2}$ | 3 | 1 | AGES-Reykjavik; INTERVAL; Gilly |
| ENTP5 | -0,114 | $2.4\times10^{-8}$ | LPC a C20:3 | deCODE | 3 | -0.101; -0.11; -0.044 | $4.4\times10^{-2}$; $1.3\times10^{-10}$; $2.9\times10^{-2}$ | 3 | 1 | AGES-Reykjavik; INTERVAL; Gilly |
| PLXB2 | 0,032 | $2.5\times10^{-8}$ | Aspartate | deCODE | 2 | 0.014; 0.025 | $9.4\times10^{-2}$; $5.6\times10^{-8}$ | 1 | 1 | AGES-Reykjavik; INTERVAL |
| PLXB2 | 0,033 | $8.9\times10^{-8}$ | Serine | deCODE | 2 | 0.022; -0.001 | $4.1\times10^{-3}$; $9.2\times10^{-1}$ | 1 | 0 | AGES-Reykjavik; INTERVAL |
| PLXB2 | 0,115 | $1.0\times10^{-100}$ | PC aa C42:1 | deCODE | 2 | 0.048; 0.034 | $4.4\times10^{-2}$; $4.0\times10^{-4}$ | 2 | 0 | AGES-Reykjavik; INTERVAL |
| PLXB2 | 0,072 | $1.5\times10^{-14}$ | PC aa C42:6 | deCODE | 2 | 0.062; 0.009 | $1.2\times10^{-5}$; $6.4\times10^{-1}$ | 1 | 1 | AGES-Reykjavik; INTERVAL |
| PLXB2 | 0,086 | $8.3\times10^{-8}$ | PC ae C42:2 | deCODE | 2 | 0.048; 0.018 | $1.7\times10^{-5}$; $4.3\times10^{-1}$ | 1 | 1 | AGES-Reykjavik; INTERVAL |
| PLXB2 | -0,051 | $3.6\times10^{-7}$ | PC ae C44:6 | deCODE | 2 | 0.019; 0.061 | $4.4\times10^{-1}$; $1.9\times10^{-2}$ | 0 | 0 | AGES-Reykjavik; INTERVAL |
| ADH4 | -0,256 | $5.3\times10^{-9}$ | Tetradecanoylcarnitine | deCODE | 1 | -0.152 | $3.6\times10^{-2}$ | 1 | 0 | AGES-Reykjavik |
| ADH4 | -0,757 | $1.2\times10^{-8}$ | PC aa C36:4 | deCODE | 1 | -0.595 | $2.9\times10^{-2}$ | 1 | 0 | AGES-Reykjavik |
| ADH4 | -0,251 | $2.5\times10^{-9}$ | PC aa C40:4 | deCODE | 1 | -0.126 | $1.8\times10^{-1}$ | 0 | 0 | AGES-Reykjavik |
| ADH4 | -0,3 | $1.3\times10^{-14}$ | PC aa C42:1 | deCODE | 1 | -0.076 | $7.7\times10^{-1}$ | 0 | 0 | AGES-Reykjavik |
| ADH4 | -0,654 | $3.1\times10^{-8}$ | PC ae C38:3 | deCODE | 1 | -0.152 | $4.6\times10^{-2}$ | 1 | 0 | AGES-Reykjavik |
| ADH4 | -0,607 | $2.8\times10^{-7}$ | PC ae C38:5 | deCODE | 1 | -0.073 | $2.9\times10^{-1}$ | 0 | 0 | AGES-Reykjavik |
| ADH4 | -0,728 | $6.1\times10^{-8}$ | PC ae C40:5 | deCODE | 1 | -0.626 | $1.3\times10^{-2}$ | 1 | 0 | AGES-Reykjavik |
| TDGF1 | 0,03 | $7.6\times10^{-10}$ | Serine | deCODE | 3 | 0.014; 0.022; 0.059 | $4.1\times10^{-5}$; $4.0\times10^{-9}$; $1.8\times10^{-2}$ | 3 | 2 | AGES-Reykjavik; INTERVAL; Yang |
| TDGF1 | -0,038 | $5.7\times10^{-9}$ | PC aa C42:6 | deCODE | 3 | -0.019; -0.032; -0.045 | $1.0\times10^{-4}$; $1.6\times10^{-9}$; $3.4\times10^{-1}$ | 2 | 2 | AGES-Reykjavik; INTERVAL; Yang |
| TDGF1 | 0,033 | $1.2\times10^{-11}$ | SM C16:0 | deCODE | 3 | 0.005; 0.008; -0.024 | $3.7\times10^{-1}$; $8.1\times10^{-2}$; $4.0\times10^{-1}$ | 0 | 0 | AGES-Reykjavik; INTERVAL; Yang |

* Columns: Point estimate - mean difference of indicated association between protein and metabolite in the discovery study, p-value - p-value of indicated association in the discovery study, Discovery study - study with largest sample size,
No. other occurences - number of other studies in which replication was attempted, Other point estimates - mean difference of indicated association for each study in which replication was attempted,
Other p-values - p-value of indicated association for each study in which replication was attempted, Nominal replicates - number of times association was replicated with a p-value < 0.05,
Adjusted replicates - number of times association was replicated with an adjusted p-value, Studies - studies in which replication was attempted, see Methods section for more details.
Abbreviations: a = acyl residue, aa = diacyl residue, ae = acyl-alkyl residue, LPC = lysophosphatidylcholine, PC = phosphatidylcholine, SM = sphingomyelin
