## Appendix for "Integrating metabolomics and proteomics to identify novel drug targets for heart failure and atrial fibrillation"

Figure S5……………………………………………………………………………………….10

Figure S6……………………………………………………………………………………….11

Figure S7……………………………………………………………………………………..…12

### Supplementary Note

#### Methods

*Data sources*

Genetic associations with the metabolites were sourced from Lotta et al. and the metabolites are grouped in the following classes: amino acids (AAs), biogenic amines, acylcarnitines (ACs), phosphatidylcholines (PCs), lysophosphatidylcholines (LPCs), sphingomyelins (SMs), and hexoses.

*mRNA expression and enrichment*

Cardiac mRNA expression was obtained from the human protein atlas (HPA), sourcing the consensus expression obtained by normalised transcripts per million (nTPM) values from three independent transcriptomics datasets: GTEx, Fantom5, and HPA's own data. Overexpressed genes were identified by comparing cardiac expression with average expression in other tissues, testing against a standard normal quantile of 1.96. Enrichment analysis, evaluating the frequency of association across metabolite classes was conducted using Fisher's exact.

*Networks*

We identified metabolites clusters based on a shared protein profile (defined as 20% or more protein in common), and visualised the triangulated associations using annotated networks. Reactome pathway enrichment of the proteins belonging to the metabolite networks compared to the full set of 1,567 proteins was tested using a Wald test.

#### Results

*Replicating protein associations with metabolites*

Out of the 82 prioritised proteins, 49 were available in more than one GWAS, allowing for replication of the associations with plasma metabolites. Applying a nominal replication p-value of 0.05 we were able to replicate the metabolite association of 45 proteins (91.8%; **Appendix Table S12**). Applying a more stringent p-value cut-off resulted in 38 replicated proteins.

*Networks*

Cluster analysis was performed to identify plasma metabolites sharing common protein effects, identifying four groups (**Figure 1C**, **Appendix Figure S3-5**). This highlighted the following biological pathways: “TP53 regulates transcription of cell death genes” and “metabolism of RNA” (**Appendix Figure S6**).

### Figure legends

**Figure S1. Volcano plots displaying proteins associated with the metabolites.**

NB. Labelled proteins are drugged, which is defined as proteins targeted by a compound (see Methods); the p-value was truncated to a -log10 of 16 for visualisation purposes only. Abbreviations: a = acyl residue, aa = diacyl residue, ae = acyl-alkyl residue, LPC = lysophosphatidylcholine, MD = mean difference, PC = phosphatidylcholine, SM = sphingomyelin.

**Figure S2. Volcano plots displaying proteins associated with the cardiac outcomes.**

NB. Labelled proteins are drugged, which is defined as proteins targeted by a compound (see Methods); the p-value was truncated to a -log10 of 16 for visualisation purposes only. Abbreviations: AF = atrial fibrillation, DCM = dilated cardiomyopathy, HF = heart failure, NICM = non-ischemic cardiomyopathy, OR = odds ratio.

**Figure S3. Heatmap displaying the percentage of overlapping associated proteins between metabolite pairs.**

NB. Orange boxes represent clusters further studied in detail.

Abbreviations: a = acyl residue, aa = diacyl residue, ae = acyl-alkyl residue, LPC = lysophosphatidylcholine, PC = phosphatidylcholine, SM = sphingomyelin.

**Figure S4. Annotated network of prioritised metabolites, proteins, and outcomes for which the metabolites have at least 20% associated common proteins belonging to the metabolite classes LPC and PC.**

NB. Prioritised proteins are represented by circles, metabolites by diamonds, outcomes by triangles. Circle colours represent protein druggability, where drugged is defined as targeted by an approved compound and druggable as targeted by a compound, see Methods. Increasing effect is displayed by a red arrow, decreasing effect by a blue arrow.

Abbreviations: a = acyl residue, aa = diacyl residue, AF = atrial fibrillation, HF = heart failure, LPC = lysophosphatidylcholine, PC = phosphatidylcholine.

**Figure S5. Annotated network of prioritised metabolites, proteins, and outcomes for which the metabolites have at least 20% associated common proteins belonging to the metabolite class PC.**

NB. Prioritised proteins are represented by circles, metabolites by diamonds, outcomes by triangles. Circle colours represent protein druggability, where drugged is defined as targeted by an approved compound and druggable as targeted by a compound, see Methods. Increasing effect is displayed by a red arrow, decreasing effect by a blue arrow.
Abbreviations: ae = acyl-alkyl residue, AF = atrial fibrillation, HF = heart failure, PC = phosphatidylcholine.

**Figure S6. Annotated network of prioritised metabolites, proteins, and outcomes for which the metabolites have at least 20% associated common proteins belonging to the metabolite classes LPC and PC.**

NB. Prioritised proteins are represented by circles, metabolites by diamonds, outcomes by triangles. Circle colours represent protein druggability, where drugged is defined as targeted by an approved compound and druggable as targeted by a compound, see Methods. Increasing effect is displayed by a red arrow, decreasing effect by a blue arrow.
Abbreviations: a = acyl residue, aa = diacyl residue, ae = acyl-alkyl residue, AF = atrial fibrillation, HF = heart failure, LPC = lysophosphatidylcholine, PC = phosphatidylcholine.

**Figure S7. Enriched Reactome pathways and their corresponding -log 10(p-value) per metabolite cluster.**

CAS = contact activation system (CAS); KKS = kallikrein/kinin system

Abbreviations: LPC = lysophosphatidylcholine, PC = phosphatidylcholine.

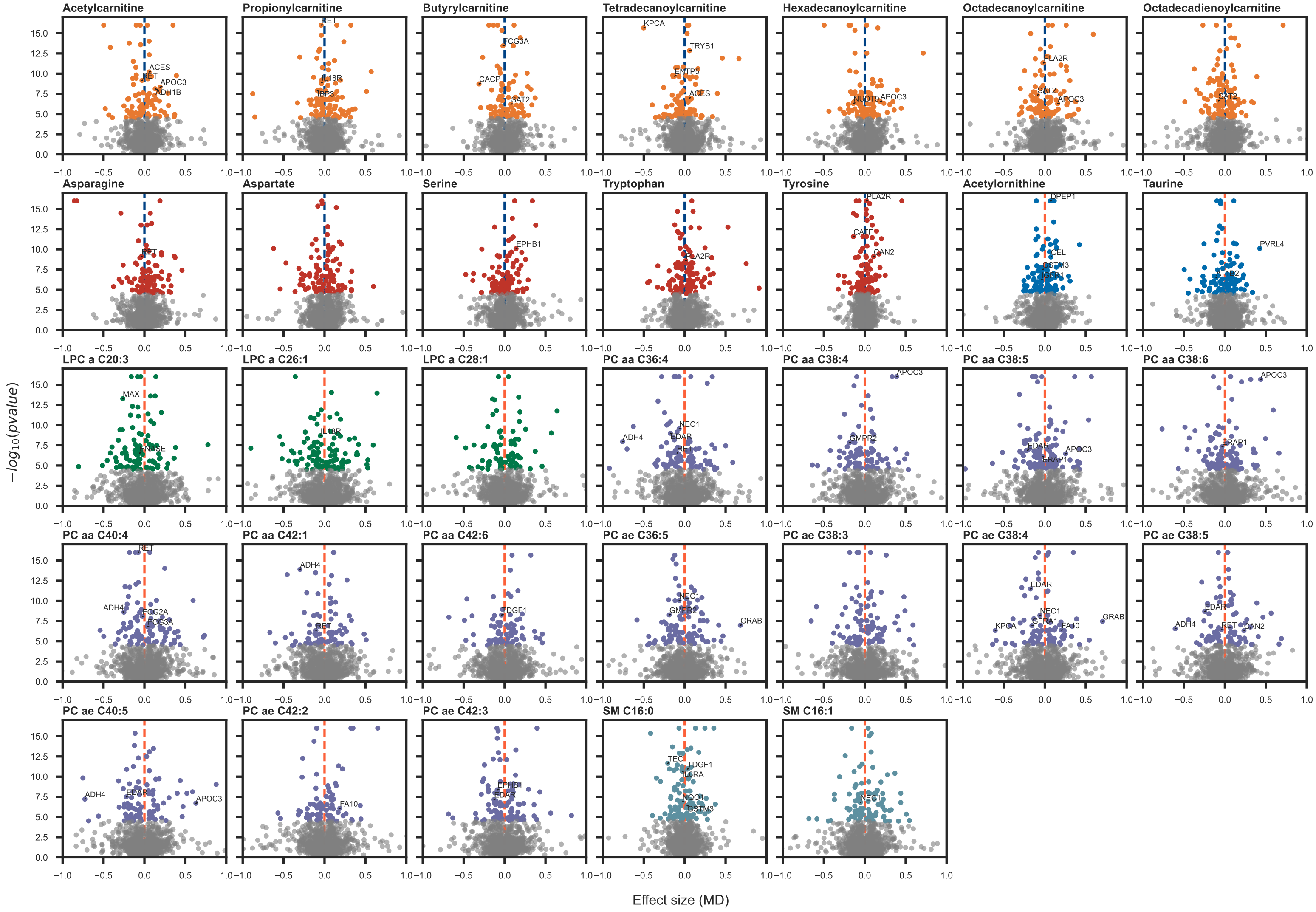

Acetylcarnitine
Propionylcarnitine
Butyrylcarnitine
Tetradecanoylcarnitine
Hexadecanoylcarnitine
Octadecanoylcarnitine
Octadecadienoylcarnitine
Asparagine
Aspartate
Serine
Tryptophan
Tyrosine
Acetylornithine
Taurine
LPC a C20:3
LPC a C26:1
LPC a C28:1
PC aa C36:4
PC aa C38:4
PC aa C38:5
PC aa C38:6
PC aa C40:4
PC aa C42:1
PC aa C42:6
PC ae C36:5
PC ae C38:3
PC ae C38:4
PC ae C38:5
PC ae C40:5
PC ae C42:2
PC ae C42:3
SM C16:0
SM C16:1
$-log_{10}(pvalue)$
Effect size (MD)

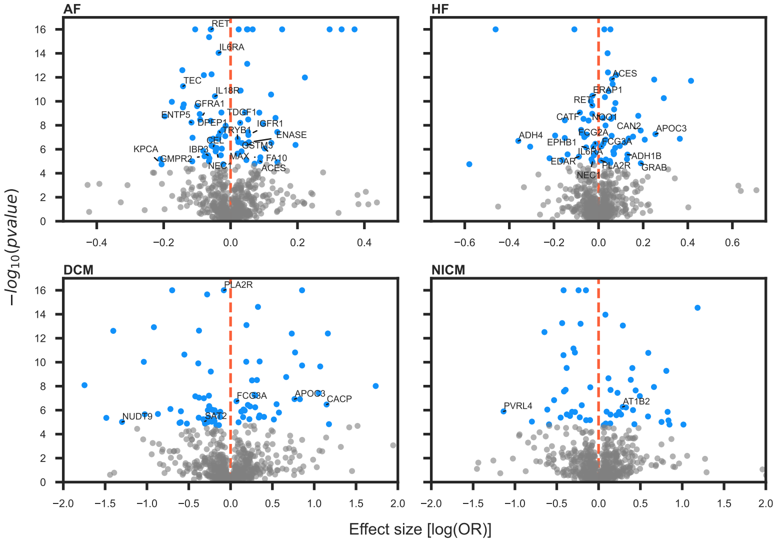

AF
HF
DCM
NICM
$-log_{10}(pvalue)$
Effect size [log(OR)]
RET
IL6RA
TEC
IL18R
GFRA1
ENTP5
TDGF1
DPEP1
IGFR1
TRYB1
ENASE
CEL
GSTM3
KPCA
IBP3
MAX
FA10
GMPR2
NEC1
ACES
ERAP1
CATF
NQO1
CAN2
APOC3
ADH4
FCG2A
FCG3A
EPHB1
IL6RA
ADH1B
EDAR
PLA2R
GRAB
PLA2R
FCG3A
APOC3
CACP
NUDT9
SAT2
PVRL4
AT1B2

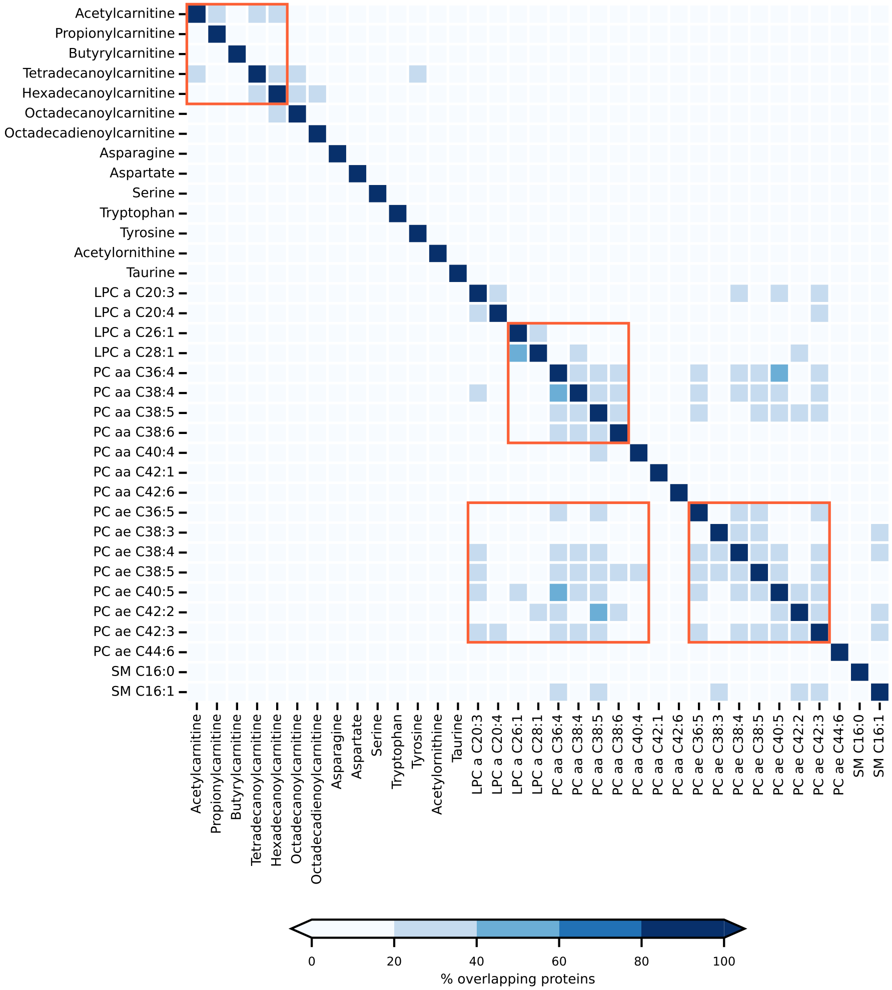

Acetylcarnitine
Propionylcarnitine
Butyrylcarnitine
Tetradecanoylcarnitine
Hexadecanoylcarnitine
Octadecanoylcarnitine
Octadecadienoylcarnitine
Asparagine
Aspartate
Serine
Tryptophan
Tyrosine
Acetylornithine
Taurine
LPC a C20:3
LPC a C20:4
LPC a C26:1
LPC a C28:1
PC aa C36:4
PC aa C38:4
PC aa C38:5
PC aa C38:6
PC aa C40:4
PC aa C42:1
PC aa C42:6
PC ae C36:5
PC ae C38:3
PC ae C38:4
PC ae C38:5
PC ae C40:5
PC ae C42:2
PC ae C42:3
PC ae C44:6
SM C16:0
SM C16:1
0
20
40
60
80
100
% overlapping proteins

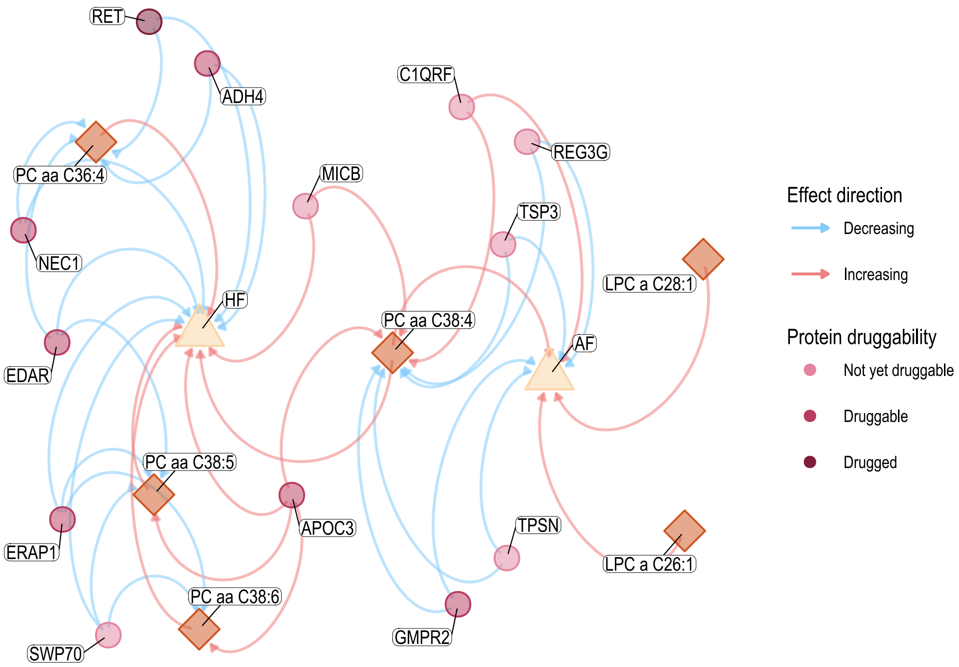
RET
ADH4
C1QRF
REG3G
MICB
TSP3
PC aa C36:4
NEC1
HF
LPC a C28:1
PC aa C38:4
AF
EDAR
PC aa C38:5
ERAP1
APOC3
TPSN
LPC a C26:1
PC aa C38:6
SWP70
GMPR2
Effect direction
Decreasing
Increasing
Protein druggability
Not yet druggable
Druggable
Drugged

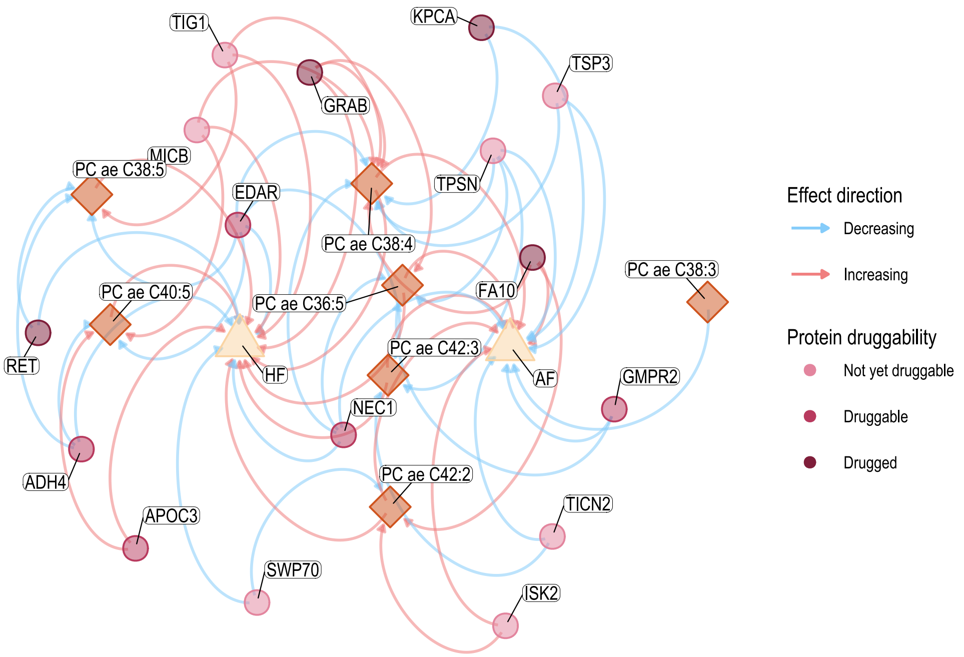

TIG1
KPCA
TSP3
GRAB
MICB
PC ae C38:5
EDAR
TPSN
PC ae C38:4
FA10
PC ae C38:3
PC ae C40:5
PC ae C36:5
RET
HF
PC ae C42:3
AF
GMPR2
NEC1
ADH4
PC ae C42:2
APOC3
TICN2
SWP70
ISK2
Effect direction
Decreasing
Increasing
Protein druggability
Not yet druggable
Druggable
Drugged

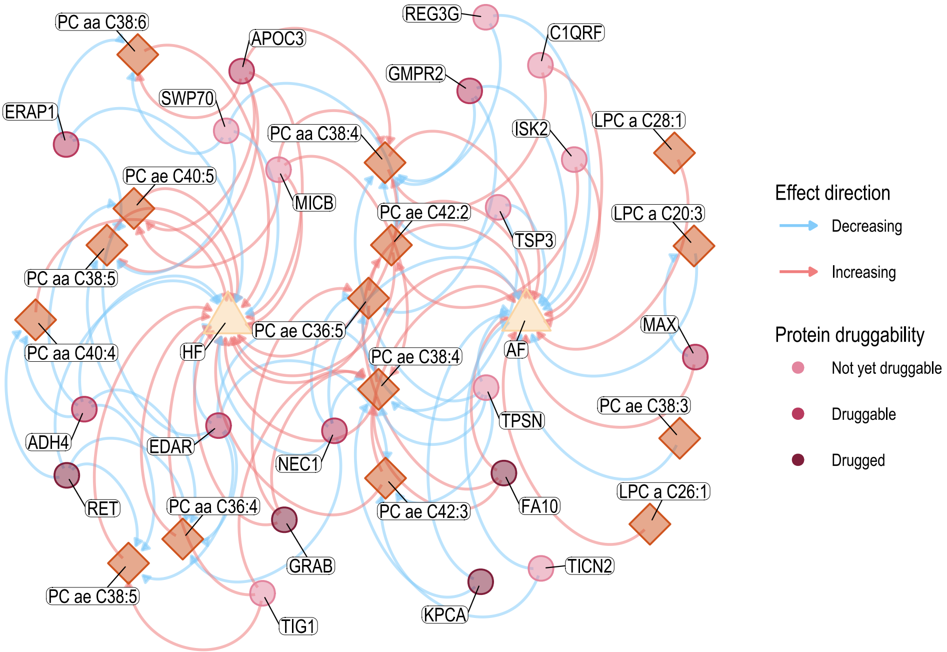
PC aa C38:6
APOC3
REG3G
C1QRF
GMPR2
SWP70
ERAP1
PC aa C38:4
ISK2
LPC a C28:1
MICB
PC ae C40:5
PC ae C42:2
TSP3
LPC a C20:3
PC aa C38:5
PC ae C36:5
HF
AF
MAX
PC aa C40:4
PC ae C38:4
TPSN
PC ae C38:3
ADH4
EDAR
NEC1
FA10
PC aa C36:4
PC ae C42:3
LPC a C26:1
RET
GRAB
TICN2
PC ae C38:5
KPCA
TIG1
Effect direction
Decreasing
Increasing
Protein druggability
Not yet druggable
Druggable
Drugged

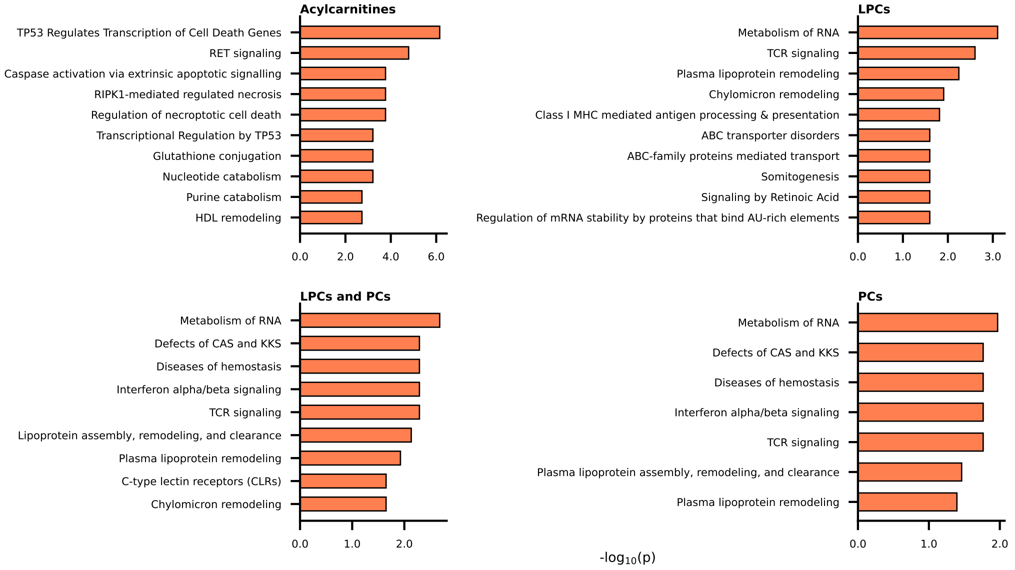
Acylcarnitines
TP53 Regulates Transcription of Cell Death Genes
RET signaling
Caspase activation via extrinsic apoptotic signalling
RIPK1-mediated regulated necrosis
Regulation of necroptotic cell death
Transcriptional Regulation by TP53
Glutathione conjugation
Nucleotide catabolism
Purine catabolism
HDL remodeling
0.0
2.0
4.0
6.0
LPCs
Metabolism of RNA
TCR signaling
Plasma lipoprotein remodeling
Chylomicron remodeling
Class I MHC mediated antigen processing & presentation
ABC transporter disorders
ABC-family proteins mediated transport
Somitogenesis
Signaling by Retinoic Acid
Regulation of mRNA stability by proteins that bind AU-rich elements
0.0
1.0
2.0
3.0
LPCs and PCs
Metabolism of RNA
Defects of CAS and KKS
Diseases of hemostasis
Interferon alpha/beta signaling
TCR signaling
Lipoprotein assembly, remodeling, and clearance
Plasma lipoprotein remodeling
C-type lectin receptors (CLRs)
Chylomicron remodeling
0.0
1.0
2.0
PCs
Metabolism of RNA
Defects of CAS and KKS
Diseases of hemostasis
Interferon alpha/beta signaling
TCR signaling
Plasma lipoprotein assembly, remodeling, and clearance
Plasma lipoprotein remodeling
0.0
1.0
2.0
$-\log_{10}(p)$
